# Development of a dynamic counterfactual risk stratification strategy for newly diagnosed acute myeloid leukemia patients treated with venetoclax and azacitidine

**DOI:** 10.1101/2024.11.25.24317750

**Authors:** Nazmul Islam, Justin L. Dale, Jamie S. Reuben, Karan Sapiah, James W. Coates, Frank R. Markson, Jingjing Zhang, Lezhou Wu, Maura Gasparetto, Brett M. Stevens, Sarah E. Staggs, William M. Showers, Monica R. Ransom, Jairav Desai, Ujjwal V. Kulkarni, Krysta L. Engel, Craig T. Jordan, Michael Boyiadzis, Clayton A. Smith

**Affiliations:** RefinedScience, Aurora, Colorado; Department of Pathology, University of Colorado Anschutz Campus, Aurora, Colorado; Department of Medicine, University of Colorado Anschutz Campus, Aurora, Colorado; OncoVerity, Aurora, Colorado

**Keywords:** Acute Myeloid Leukemia, Venetoclax, Azacitidine, Risk Stratification, Machine Learning

## Abstract

**Objective:** The objective of this study was to develop a flexible risk stratification strategy for Acute Myeloid Leukemia (AML) that is specific for venetoclax plus azacitidine (ven/aza), addresses real-world data (RWD) issues, and is also adaptable to different use cases.

**Materials and Methods:** A series of tunable risk models (RMs) were generated from a dynamic counterfactual machine learning (ML) strategy. These utilized a range of features from diagnostic AML samples and were tested using objective metrics on a single institution cohort of 316 newly diagnosed patients treated with ven/aza.RM performance was tested using various model assumptions, data elements, and endpoints, and with applications to an external AML real world cohort (RWC).

**Results:** Favorable, Intermediate, and Adverse risk groups were identified in a series of ML-based RMs using different assumptions, for genetic only or genetic-plus-phenotypic data elements and with overall survival and event free survival as endpoints. Most RMs demonstrated equitable patient distribution (∼20%-40% in each risk group), significant separation between risk strata (Log-rank based P-values <0.001), and predictability computed by time-dependent survival AUC values of 0.60-0.70. Similar performance was observed when the proposed RM strategy was adapted and compared to the ELN22 using the external RWC.

**Discussion and Conclusion:** The proposed ML strategy addresses a variety of RWD considerations and is readily tunable through coding and parameter updates for different contexts and use case needs. This strategy represents a novel approach to developing more effective RMs for AML and possibly other diseases.

**CONTEXT SUMMARY:** 

**Key Objective:** Can an effective machine learning based risk stratification strategy be developed for Acute Myeloid Leukemia (AML) that address real world challenges and varying use cases?

**Knowledge Generated:** A RM strategy for AML patients treated with venetoclax/azacitidine was developed based on a dynamic counterfactual machine learning (ML) model. This strategy efficiently generated specific RMs that demonstrated robust performance with varying assumptions, data elements, and endpoints, and was adaptable to an external real world cohort dataset. These findings demonstrate that ML-based RMs are feasible and may have potential advantages over existing AML RM strategies in adapting to context dependent settings.

## INTRODUCTION

Acute myeloid leukemia (AML) is an aggressive hematopoietic malignancy diagnosed in ∼20,000 individuals annually in the United States^1^. For young and fit patients, initial treatment typically involves intensive chemotherapy (IC), followed by consolidation with allogeneic hematopoietic cell transplant (allo-HCT) or high-dose chemotherapy^2,3^. For older and less-fit patients, venetoclax (ven) combined with a hypomethylating agent (HMA) such as azacitidine (aza) is now a standard of care^4–6^. For IC therapies, various prognostic models have been developed based on patient- and AML-specific features to stratify patients into subgroups with varying outcomes^7–15^. The European Leukemia Network (ELN) developed a risk categorization approach for AML in 2017 (ELN17), with an update in 2022 (ELN22), which has been widely used^16,17^. ELN categories are divided into Favorable, Intermediate, and Adverse risk groups associated with overall survival (OS) based on AML cytogenetic and next-generation sequencing features^16,17^. However, the ELN categorization approaches were recently found to perform sub-optimally with ven/HMA-treated patients, likely because the ELN criteria are primarily based on treatment outcomes following IC^18^. Based on this, a variety of rapidly evolving new AML risk models (RMs) have been developed with differing risk features to better stratify patients treated with lower intensity agents including ven/HMA^19–24^. In addition to risk features in different RMs varying with treatment, they may also vary according to a variety of other contextual considerations including feature prevalence, missingness and sparsity of feature data, varying endpoints, collinearity among features, testing technologies generating the features, and the impact of allo-HCT and other important confounding effects on outcomes interpretation^21–25^. Furthermore, risk features are contextually dependent on the group being studied, which may vary substantially between academic, clinical trial, community, and other settings; additionally, genetic traits can also pose different risks based on a patient’s genetic background^19,26^. Lastly, RMs’ features and performance vary based on specific use case requirements. For example, up-front patient allocation in clinical trials may prioritize features that can be rapidly resulted, are easy to interpretate, and minimize missingness, while post hoc retrospective analyses can incorporate more complex features that take longer to result.

To address these issues, here we describe the development and initial testing of a flexible AML risk stratification strategy for newly diagnosed AML patients. A machine-learning (ML) based strategy was selected for its efficient adaptability to address various real-world data (RWD) considerations and its readiness to adapt to different datasets and use cases. This strategy was developed and initially tested for performance using multiple objective metrics on a single academic center cohort specifically treated with ven/aza. It was then further evaluated for adaptability and generalizability to other use cases using an external real-world cohort (RWC).

## METHODS

### Internal analytical patient population and outcomes definitions

The internal analytical cohort dataset included 316 adult patients from the University of Colorado (CU) with newly diagnosed AML treated with ven/aza between January 2015 and March 2024. Details of the patient cohort and subset management are shown in Figure 1. Patient demographics are summarized in Supplemental Table 1A. Key AML diagnostic pathology features for developing the RMs are summarized in Supplemental Tables 1B-F, including next generation sequencing (NGS), cytogenetics (CYT), fluorescence in-situ hybridization (FISH), polymerase chain reaction (PCR), flow cytometric (FC) data, and “AML mutations,” which included composite mutations from various tests. Further details are provided in Supplemental Methods S1-S2 and Supplemental Table 2. Responses (OS, event-free survival (EFS), best response (BR)) are defined in Figure 1^17,27^. This retrospective study was approved by an institutional review board (IRB) and used a limited dataset with a waiver of consent.

**Figure 1.**
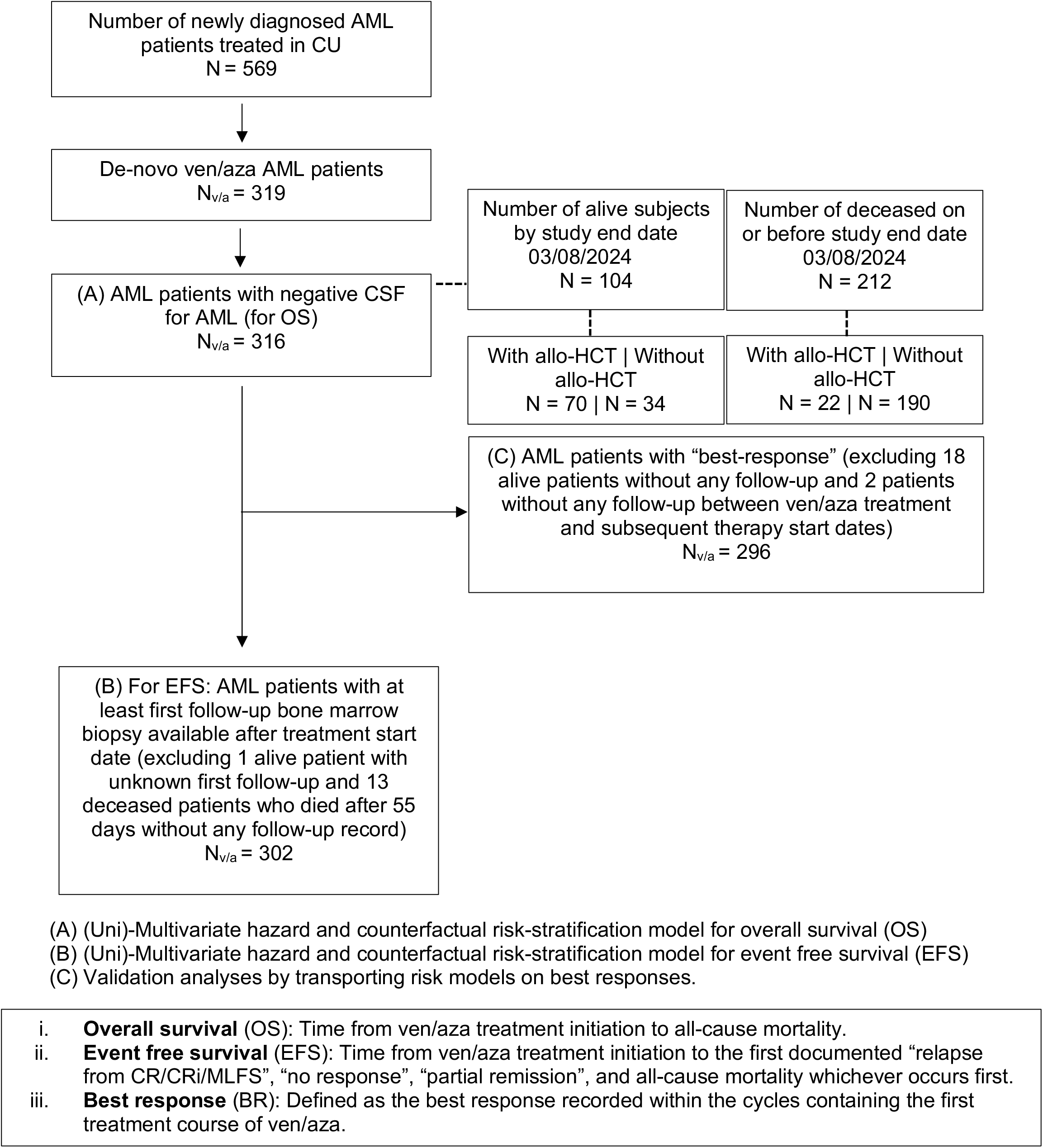
Summary of the de-novo AML ven/aza treated patient cohort. The flow diagram summarizes the patient populations in the CU dataset and how they were utilized in generating the risk stratification models for OS/BR and EFS.

### Exploratory analyses

Standardized mean differences^28^ and Kaplan-Meier (KM) analyses for baseline covariates were performed assuming right censoring with respect to OS and EFS based on a Full Analytical Set (FAS), a dataset censoring allo-HCT recipients, and a dataset excluding allo-HCT recipients. The equality of survival curves was evaluated using various testing approaches^29–35^ and noise variables were filtered to select a parsimonious feature list; further details are provided in Supplemental Methods S3. The resulting list of confounding variables (Supplemental Tables 1B-F) was used for both multivariable OS and EFS models.

### Risk model development

Table 1 summarizes the overall RM specifications, and Figure 2 shows the RM development strategy. Key components include the following.

**Figure 2.**
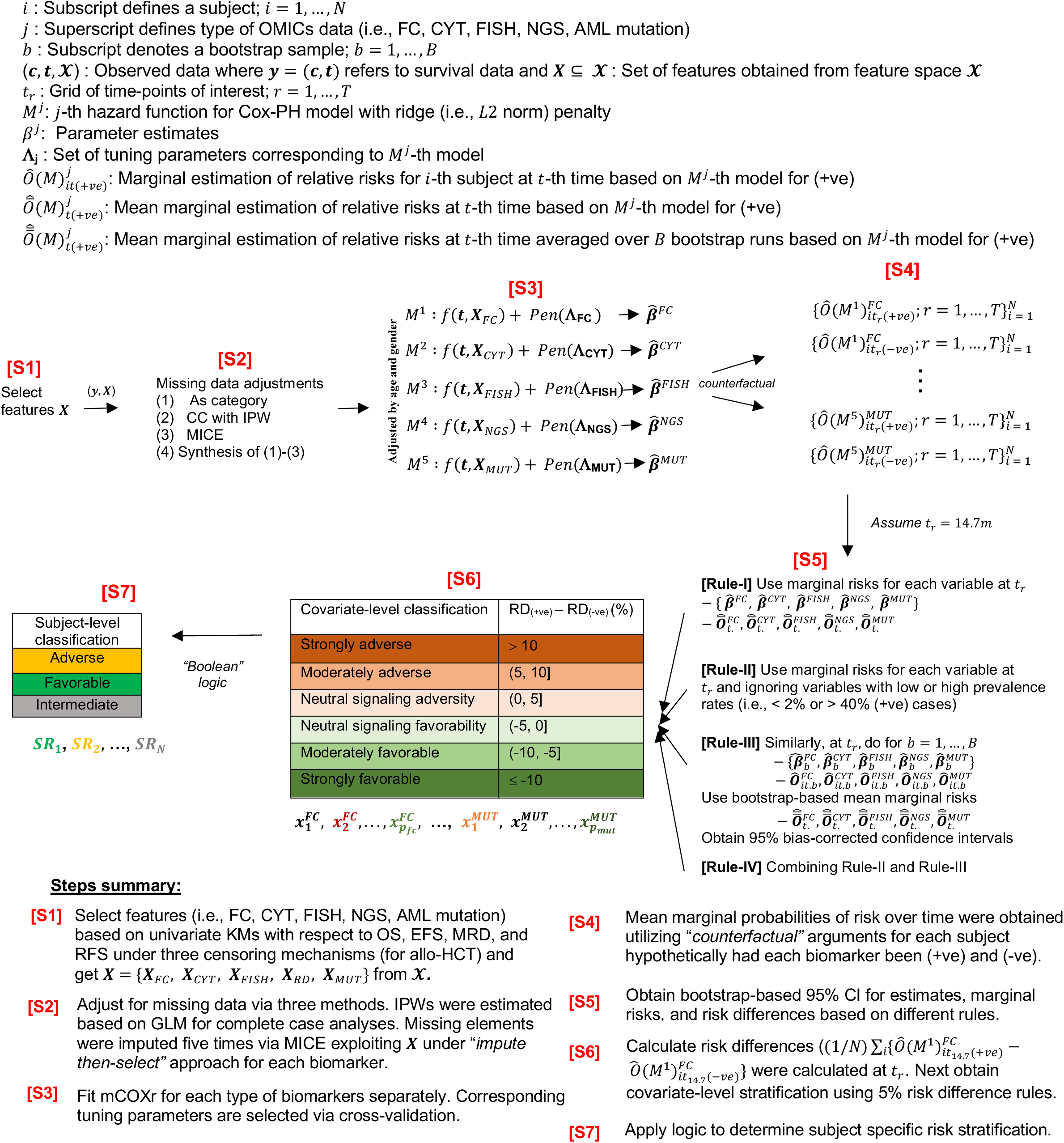
Graphical representation of the machine-learning methodology used to develop risk models with respect to overall survival and event free survival. Details of each step 1-7 are described in the Methods section.

**Table 1.**
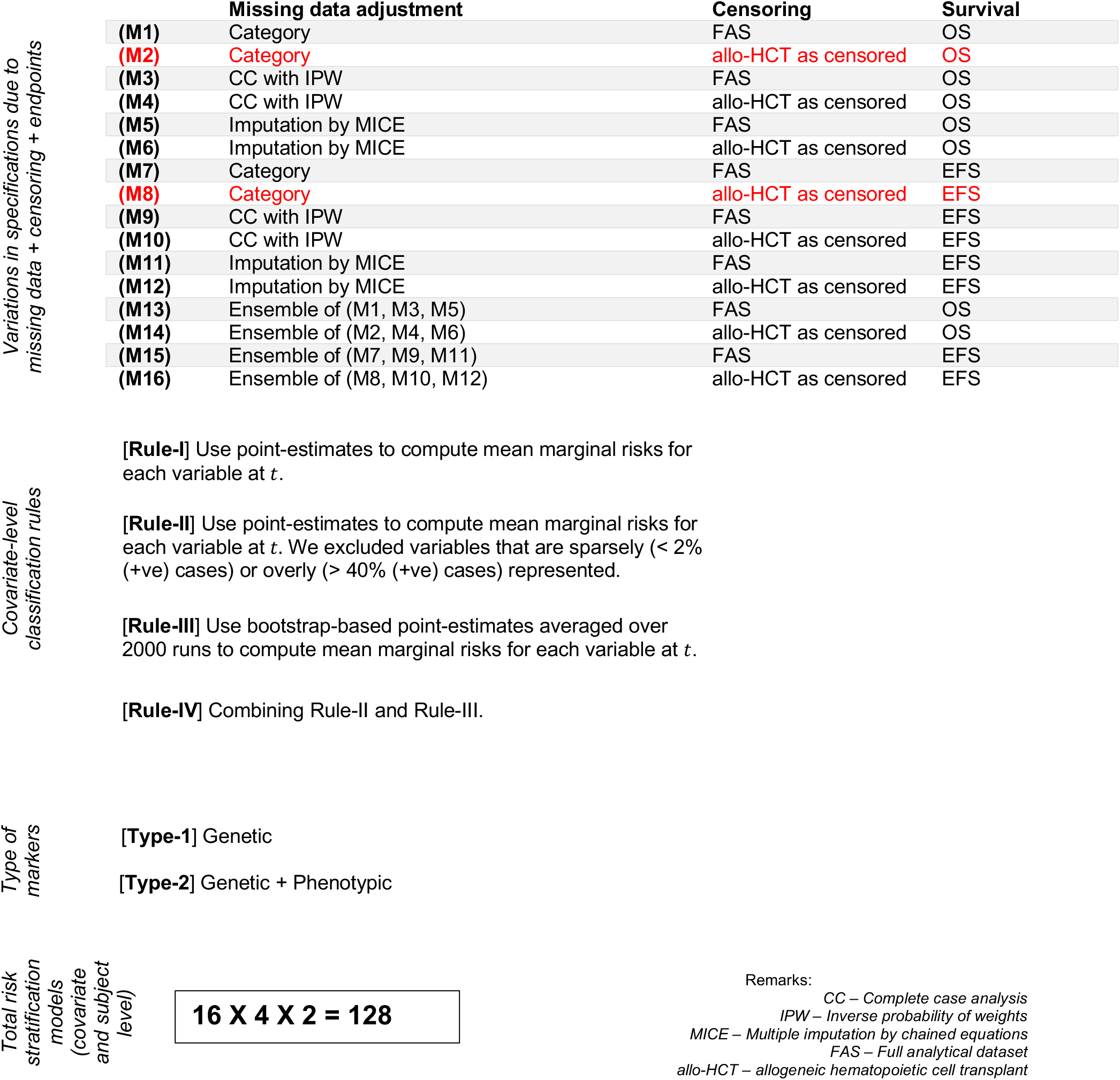
Model specifications for risk stratification models with respect to censoring assumptions, methodologies to deal with missing data, end points, variable selection rules, and feature sets. Primary models were denoted by (M1) – (M16) and reported results were based on (M2) and (M8) (highlighted in red).

### Survival regressions

ML models (i.e., multivariable penalized (ridge) Cox proportional hazard (mCOXr)^36,37^ regressions) were fitted adjusting for age and gender for OS and EFS. Separate models were fitted adjusting for confounding variables selected by null hypothesis significance testing (NHST) (Step 1, Figure 2) to identify influential variables within each group (Step 3, Figure 2). Missing values were treated as separate categories (Step 2, Figure 2). Tuning parameters were selected via cross-validation (CV). Corresponding 95% confidence intervals (CIs) of adjusted hazard ratios (aHRs) were constructed using the fractional random weight bootstrap (FRWB)^38^ approach of 2,000 runs (Supplemental Table 3-4).

### Counterfactual effects estimation

Biomarker-specific mean predicted marginal risks (MRs) over time were calculated using “*recycled*” predictions with counterfactual arguments for the corresponding mCOXr models (Step 4, Figure 2)^39,40^; see Supplemental Method S4 for details. Bootstrap-based MR profiles were constructed by averaging the FRWB estimates, and the corresponding percentile-based 95% CIs were reported. The Kolmogorov-Smirnov test assessed the differences in counterfactual risk distributions^41^. Multivariable penalized (ridge) logistic regressions^37^ were used to model BR confirming the consistency of the mCOXr estimates.

### Testing variations in risk models

#### Treatment variation

Allo-HCT is a potentially curative therapy that can profoundly affect ven/aza treated patient outcomes^42^. To address this confounding factor, RMs were developed by censoring patients with allo-HCT.

#### Varying endpoints

RMs for OS and EFS were developed separately.

#### Dynamic behavior

Counterfactual risk differences (RDs) were computed dynamically over 36 months (Steps 4-5, Figure 2, and Supplemental Figures 1A-1D). Landmarks of 14.7 and 9.8 months were chosen to find optimal feature-specific RD for OS and EFS based on the median time-to-event in the VIALE-A study, which established ven/aza as a new standard of care^6^.

#### Prevalence

Variables with low and high prevalence for (+ve) cases may cause unstable parameter estimates and imbalances in subject-level stratification. To address this, rules with varying degrees of stringency were incorporated. Rule-I (Step 5, Figure 2) is flexible and does not exclude features with respect to prevalence. Rule-II excluded any variables containing less than 2% and more than 40% (+ve) cases in risk stratification even if they exhibited more than 5% RD. Rule-III used FRWB based MR profiles to calculate RD, and Rule-IV combined Rule-II and Rule-III (see Supplemental Table 5A). *Covariate-level classification:* Two levels of classification, covariate and subject, were performed. The magnitude and direction of marginal RDs between (+ve) and (-ve) at a time point of interest were used to classify each feature into either “Adverse” or “Favorable”. Positive RD values of 0-5%, 5-10%, and >10% classified a feature as *slightly*, *moderately*, or *strongly* “Adverse,” respectively. Similarly, negative RD values determined *slightly*, *moderately*, or *strongly* “Favorable” features (Step 6, Figure 2). A cut-off value of 5% was assumed to be the clinically relevant minimum RD. Supplemental Tables 5B-5E depict covariate-level classifications under different combinations of parameters.

#### Subject-level classification

Patient-specific risk stratification (“Adverse”, “Intermediate”, and “Favorable”) was determined by combining covariate-level classifications using a ‘Boolean’ operator (AND/OR). This assigns a patient to the “Favorable” risk group without any “Adverse” features and vice versa. A patient was assigned to the “Intermediate” risk group without any “Favorable” and “Adverse” features or with at least one “Favorable” and one “Adverse” feature (Step 7, Figure 2).

#### Model specification

RMs using genetic and genetic-plus-phenotypic features were developed to reflect different scenarios and their implications on model performance (Table 1).

#### Missingness

Primary analyses treated missing values as a separate category for simplicity, avoiding imputation or exclusion. Reported results were based on this approach unless otherwise specified. Additional analyses were performed to check the robustness of different missing data handling techniques at the covariate-level classification step including: (A) Biases stemming from complete cases (CC) analyses were adjusted by incorporating inverse probability weights (IPW) into mCOXr, where subject-specific weights signaling the propensity of having any missing data were estimated from multivariable logistic regression models^43^. (B) Multiple imputations (five) were generated for each missing data element by exploiting multivariate data sequentially (multivariate imputation by chained equations (MICE))^44,45^. The corresponding aHRs were averaged over five samples and 2,000 FRWB runs, and percentile-based 95% CIs were reported. Subsequently, the marginal RDs were calculated as before and averaged over five samples. (C) Covariate-level classifications were performed based on different missing methods, with final stratification obtained by synthesizing with respect to *majority-votes*.

### Evaluation of numerical performance

Multiple evaluation criteria were used to provide a holistic view of numerical performance in terms of *equitability* (extent of risk group size variation), *separability* (extent of outcome variation by risk strata), *conformity* (extent of risk group overlap between methods), and *predictability* (degree of prognostic relationship with outcomes). Internal validation of *predictability* of the RMs and ELN22 for OS and EFS was conducted using 15-fold CV; the corresponding steps are illustrated in Figure 3. Further details of the computation and evaluation criteria are provided in the Supplemental Methods S5-S6.

**Figure 3.**
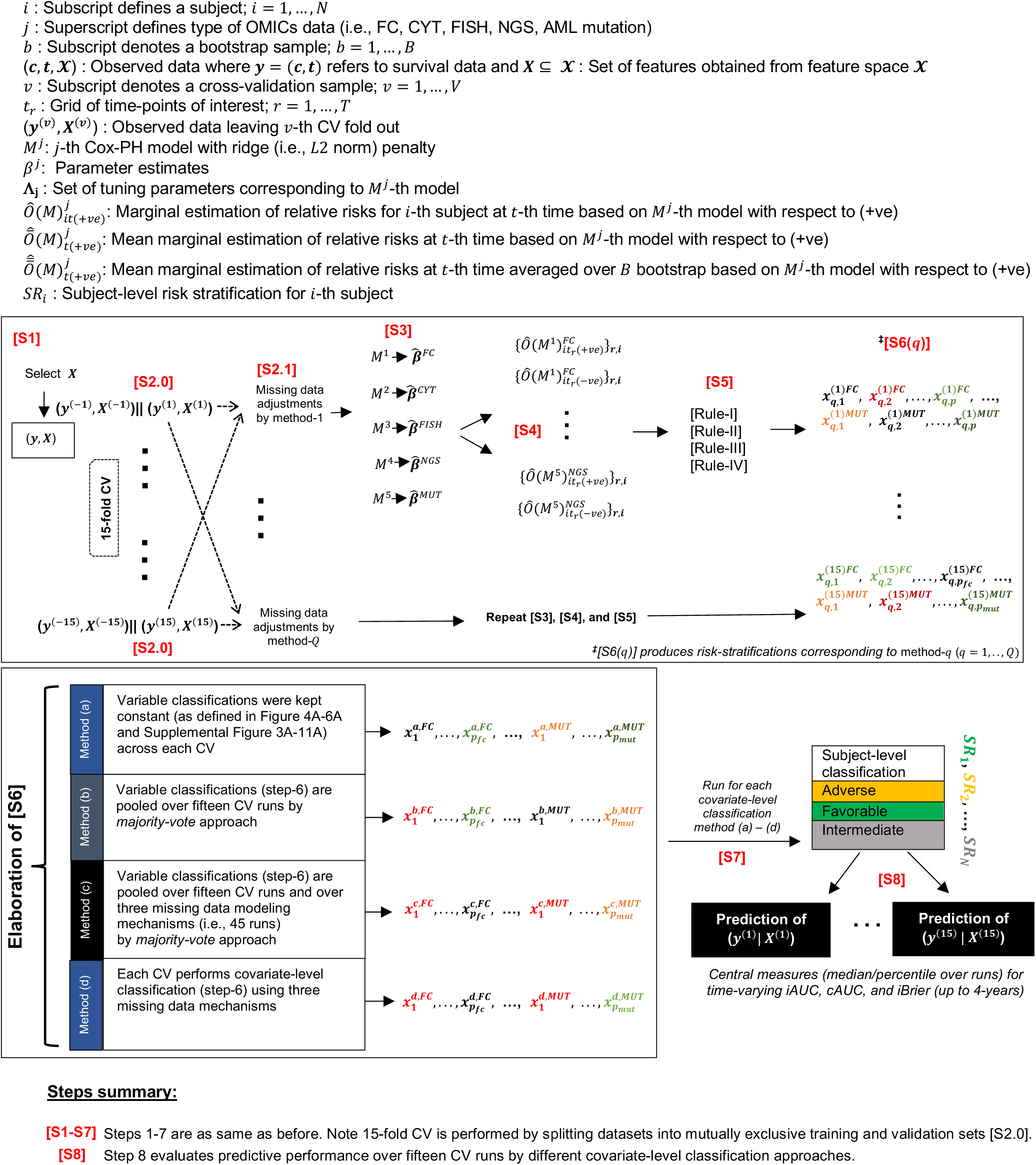
Internal validation of predictive performance for proposed risk stratification variables. Architecture of internal validation steps assessing predictive performance for proposed risk stratification variables.

### Application of risk models on an external analytical dataset

A set of specific RMs for OS was tested on an independent and heterogeneous RWC comprised of 971 AML patients treated with ven/aza at 87 unique sites of care. The RWC was derived from the nationwide Flatiron Health electronic health record-derived, de-identified, longitudinal database consisting of patient-level structured and unstructured de-identified data curated via technology-enabled abstraction^46,47^. Missing values in the RWC were generated by *imputation-by-mode* approach; further evaluation with respect to MICE was performed for sensitivity. Additional details are discussed in Supplementary Methods S7-S10.

All statistical tests, except for the comparison of MR curves, predicted curves, and survival AUC values, were two-sided with 5% significance level. As the analyses were exploratory, no multiplicity adjustments were made. All analyses were performed using R, version 4.2.3.

## RESULTS

The overall strategy for the ML-based AML RMs developed and tested here is summarized in Figure 2. By applying this strategy, we first assessed how the varying prevalence rules detailed in Table 1 influenced model performance. Rule I was identified as the least stringent, whereas Rule IV was determined to be the most stringent (Supplemental Figure 5A). The initial RMs were populated with genetic features only (termed RM_G_) and derived from the CU analytical cohort. Performance was assessed with respect to *equitability, separability, conformity,* and *predictability.* The RM_G_ that used covariate-level classification Rule-II (termed RM_G,II_) demonstrated superior performance in both *equitable* distribution of patients and OS *separability* compared to Rules I, III, and IV (Figures 4A-4F, Supplemental Figure 5 and Supplemental Table 6B). RM_G,II_ demonstrated similar performance characteristics in the allo-HCT excluded, FAS, and allo-HCT recipients censored cohorts for OS as well as BR (Figures 4B-4D). In the cohort including only allo-HCT recipients, the RM did not distinguish Favorable and Intermediate risk patients, however it did distinguish the Adverse risk population from the other two categories (Figure 4D). Covariate-level classifications of the RM_G_ variants using Rule-I (RM_G,I_), Rule-III (RM_G,III_), and Rule-IV (RM_G,IV_) are presented in Supplemental Tables 5B-5E. In general, these varying models demonstrated competitive performance in terms of *separability* and *conformity*. However, they showed reduced *equitability* in assigning patients to different risk categories, which further led to lower *predictability* in risk assessment (Supplemental Figures 2-4, Supplemental Tables 6A, 6C, and 6D). An additional evaluation of the performance of the ML-based RM strategy was conducted by comparison to ELN22. The latter demonstrated a relatively skewed distribution of patients assigned to the Adverse risk category and differences in separation of OS curves for both the allo-HCT excluded cohort and the FAS (Supplemental Figure 5A-C). Inter-group conformity analyses using Fleiss kappa demonstrated the highest level of agreement in patient assignment for the Favorable risk category and predictive evaluations (time-varying AUCs and iBrier) favored the RM_G,II_ model over ELN22 (Supplemental Figure 5E-F, Supplemental Table 6B) which is consistent with other recent reports demonstrating the limitations of ELN22 for non-IC treated patients^18,19,21–24^.

**Figure 4.**
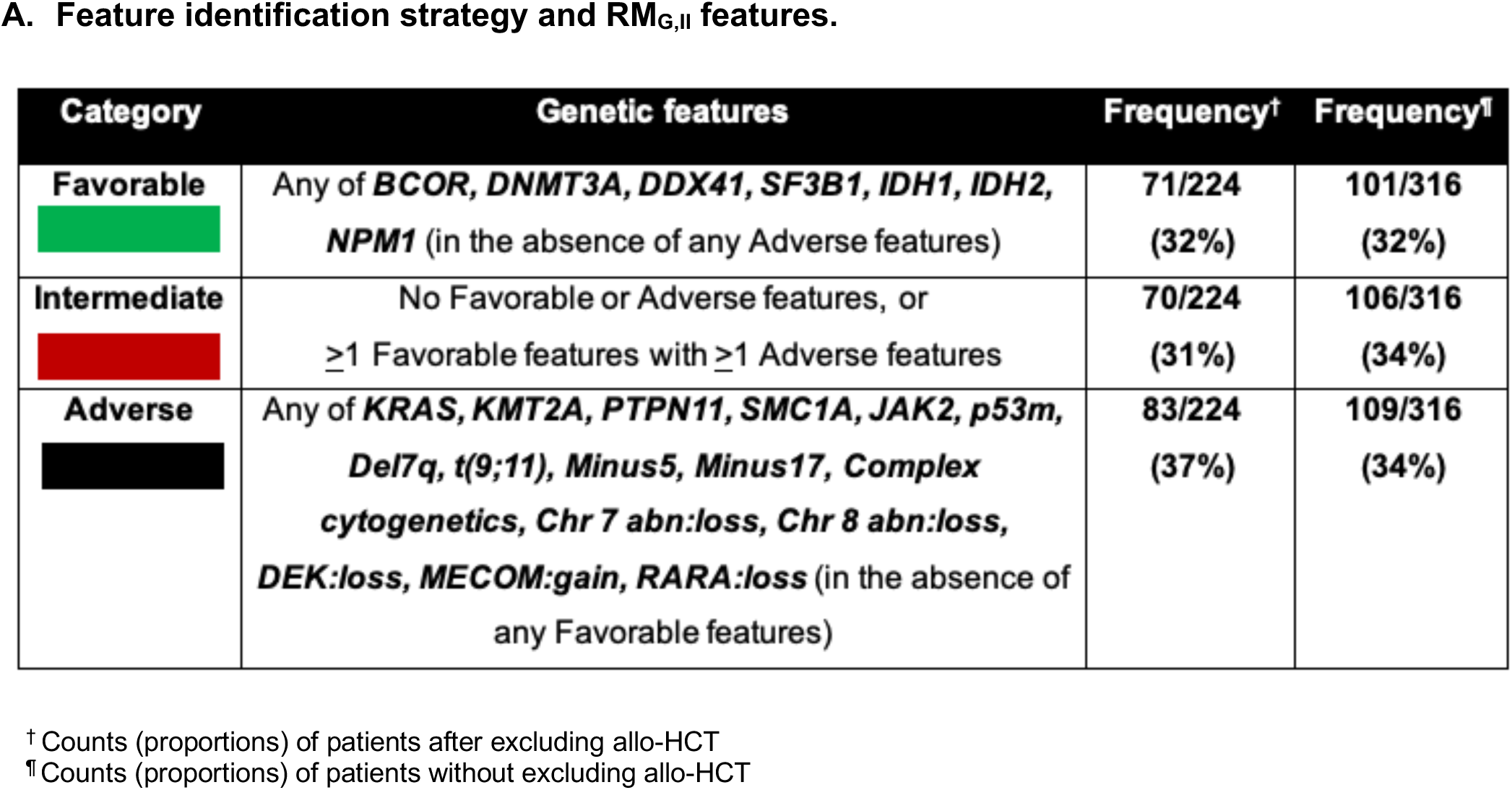
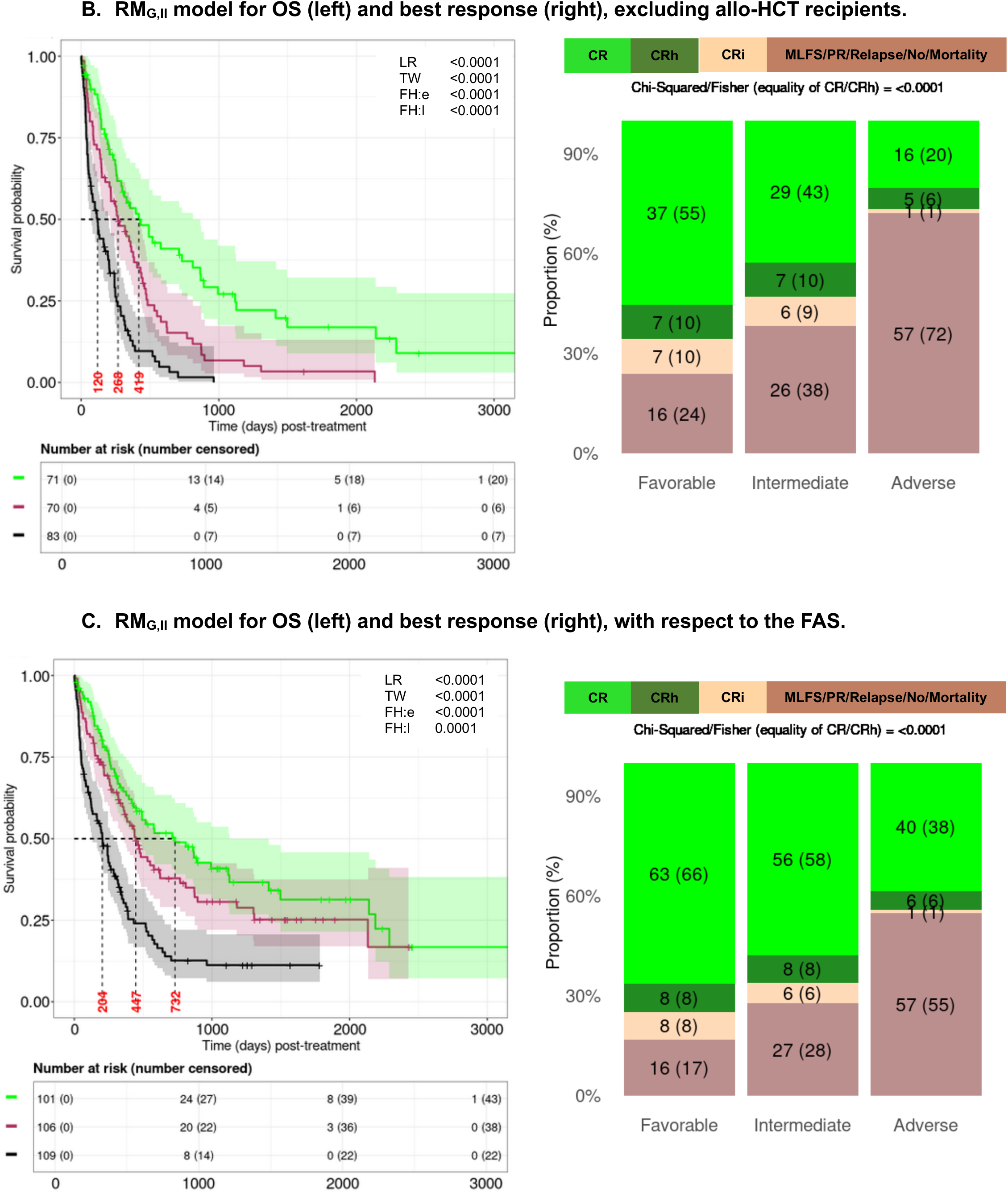
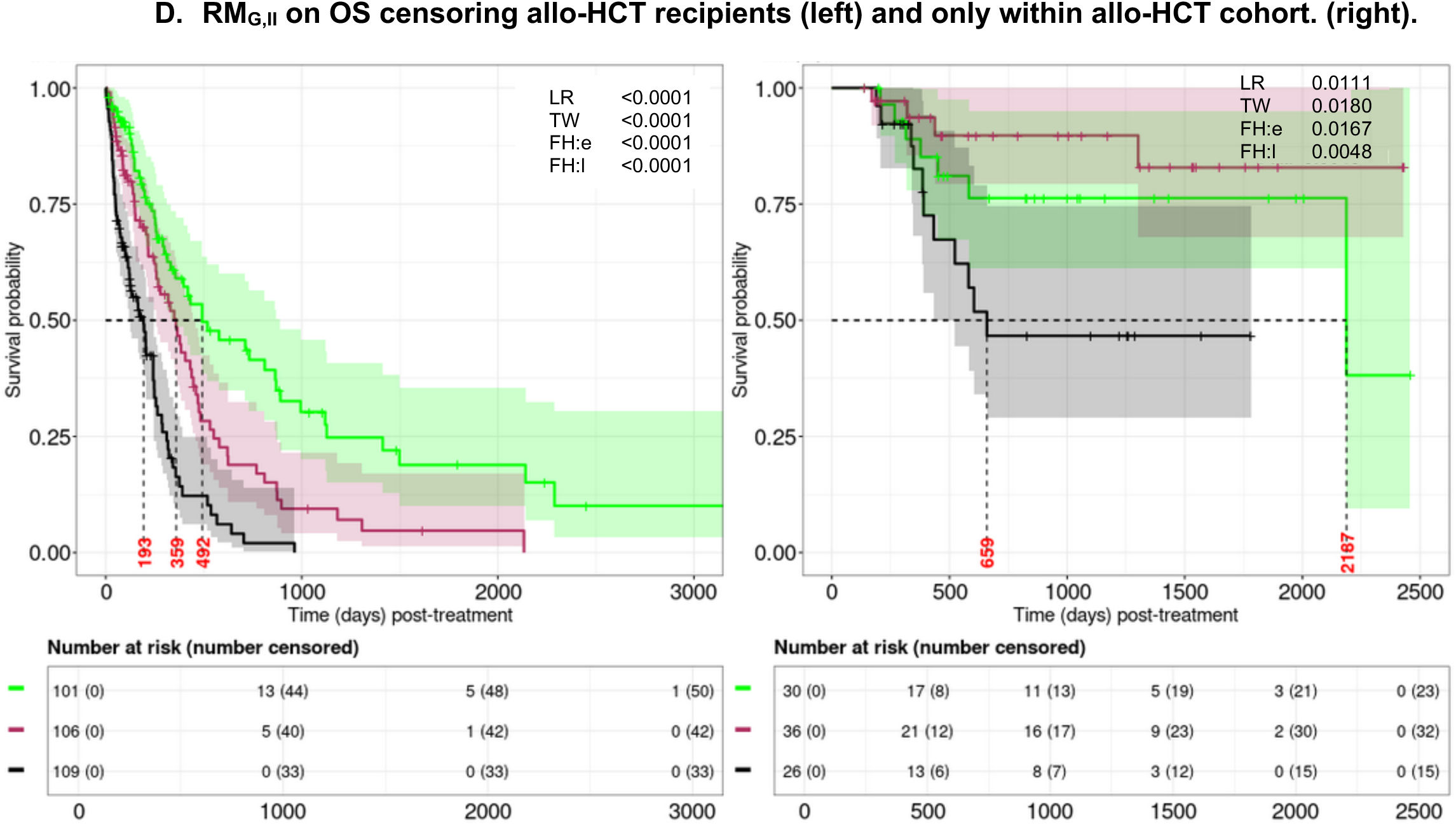
Overall survival (OS) following treatment based on the genetic features-based risk model. The genetic features-based covariate-level classification Rule-II risk model (RM_G,II_) is depicted. A) Definitions of the feature identification strategy and genetic features in RM_G,II_. The category column includes color legends for reference; B) OS (left panel) and best response (BR) (right panel) based on RM_G,II_ applied to the cohort excluding allo-HCT recipients; C) OS (left panel) and BR (right panel) based on RM_G,II_ applied to the full analytical set (FAS); and D) RM_G,II_ based on OS treating allo-HCT recipients as censored (left panel) and within the cohort comprised solely of allo-HCT recipients.

To test the adaptability of this RM strategy to a dataset with more expanded data elements, we explored the impact of including both genetic and phenotypic (flow cytometry) data on RM performance (termed RM_GP_). The impacts of applying Rule-I, II, III, and IV and the various Models in Table 1 to RM_GP_ were tested as described above. The specific features in the Adverse, Intermediate, and Favorable groups defined by the ML-based strategy varied from the RM_G_ models (Supplemental Figures 6-9, Supplemental Tables 6A-6D). In general, the overall performance of the RM_GP_ models with these different features paralleled those of the RM_G_ models, but in some cases with less *equitable* assignment of patients to the Favorable risk category depending on models, rules, and assumptions.

To test whether a new RM could be developed using the ML-based strategy for an endpoint other than OS, we developed and explored the RMs using the EFS endpoint^48^. Again, the specific risk features for Adverse, Intermediate, and Favorable risk populations varied from the OS endpoint-based models (Figure 5A) further confirming the possibility that RMs would be context dependent. Covariate-level classification Rule-II specific RM for EFS using genetic features only (i.e., RM_G,II-EFS_) demonstrated the best performance (Figure 5B-5C) and superior *predictability* (Supplementary Table 7B). In general, other EFS RMs based on the various Rules as described above demonstrated performance roughly similar to that of OS (Supplemental Tables 7A-7D, Supplemental Figures 10-17). Supplemental Figure 18 presents heatmaps that illustrate the variations in risk stratification across the different rules, endpoints, assumptions, and models, underscoring the necessity for context-specific RMs.

**Figure 5.**
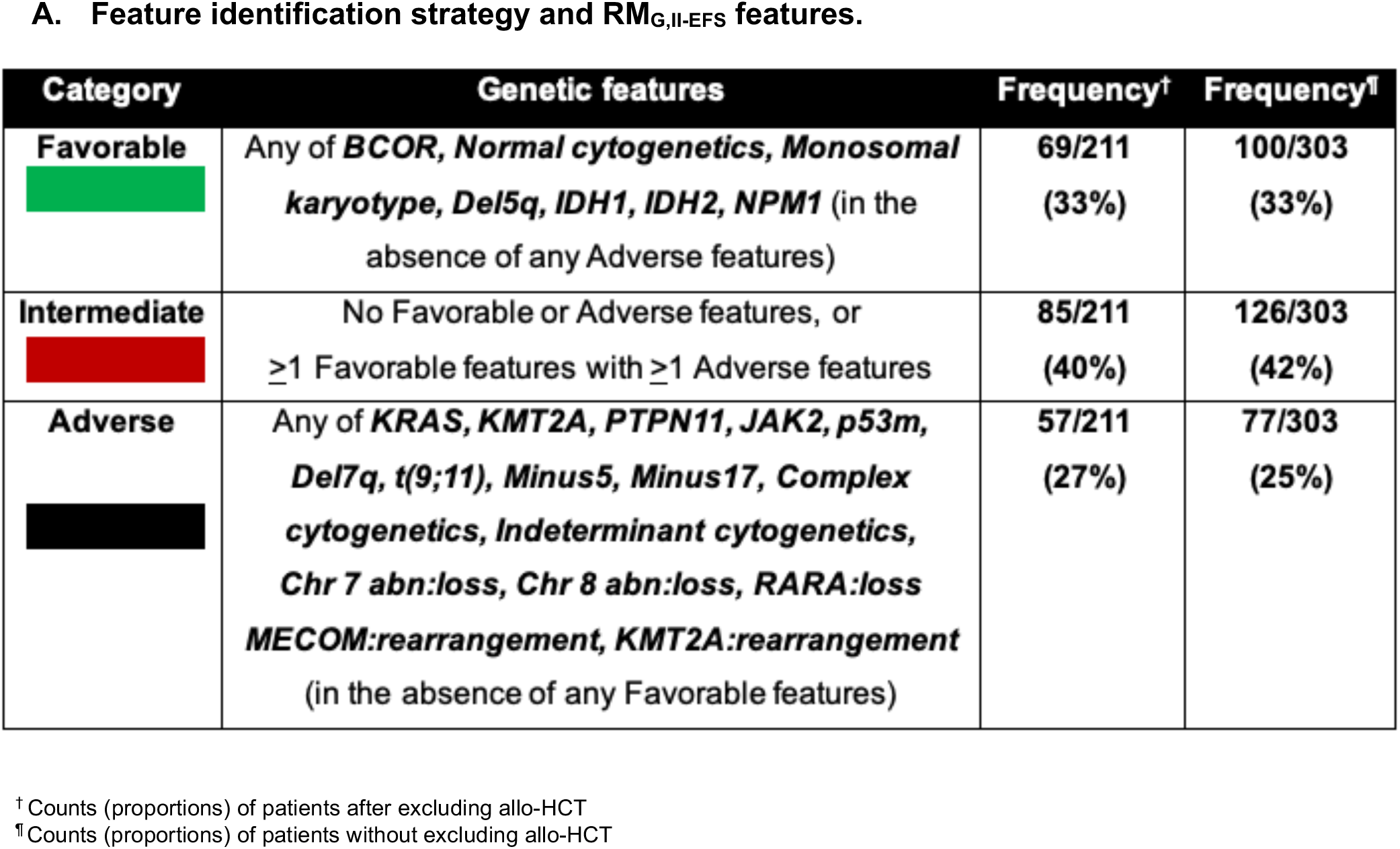
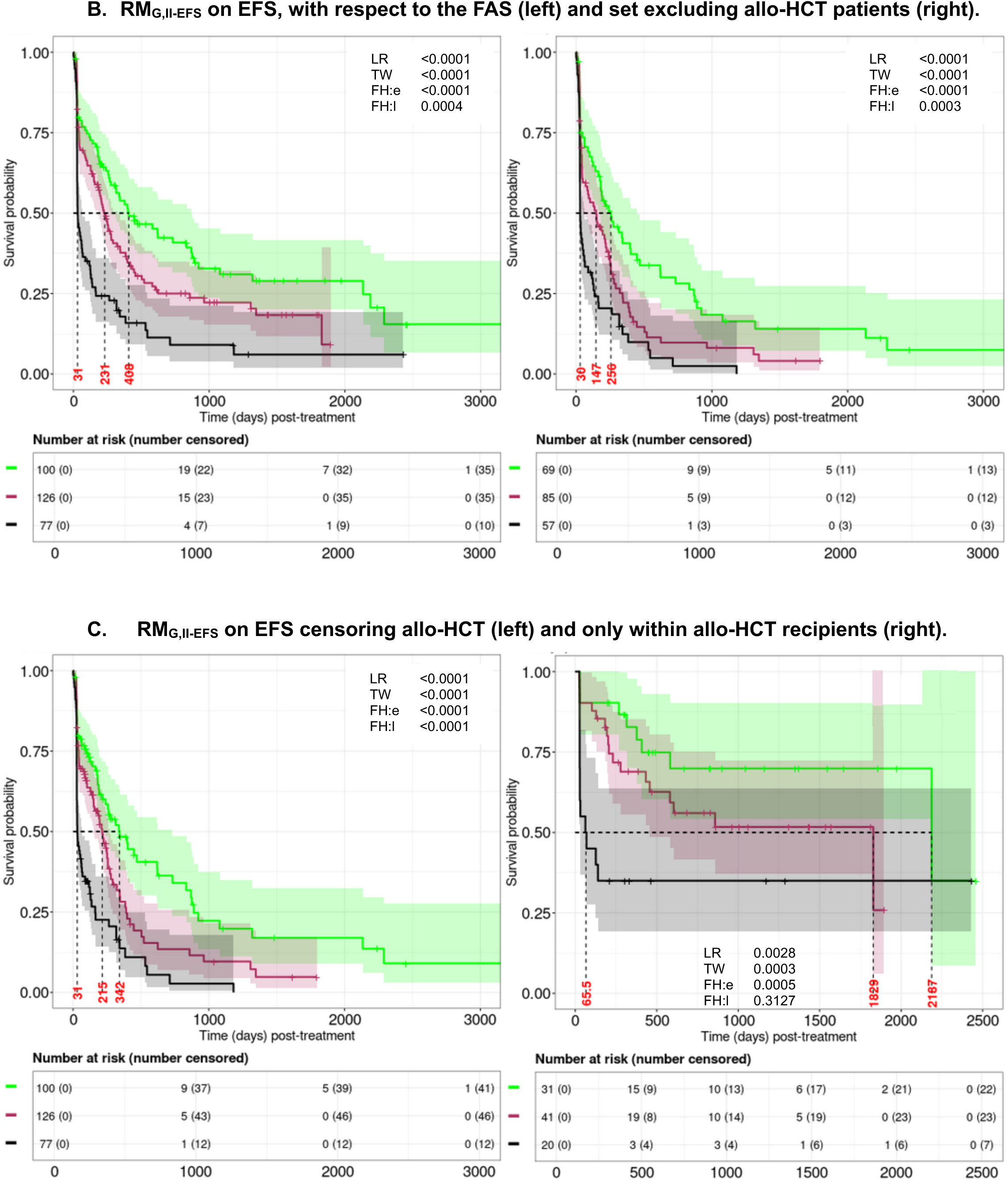
Event free survival (EFS) based on the genetic model using covariate-level classification. The covariate-level classification Rule-II Risk Model based on genetic features and specific to EFS (RM_G,II-EFS_) is depicted. A) Definitions of features associated with the RM_G,II-EFS_. The category column includes color legends for reference; B) EFS based on the RM_G,II-EFS_ applied to the full analytical set (FAS) (left panel) and set excluding (right panel) allo-HCT recipients; and C) EFS based on the RM_G,II-EFS_ treating allo-HCT recipients as censored (left panel) and only within the cohort comprised of allo-HCT recipients.

Lastly, we assessed the adaptability of the RM strategy to a different use case than retrospective analysis of an academic center cohort. The strategy was adapted to retrospective risk stratification of patients in a different external dataset (the “RWC”) derived from patients treated primarily in community centers. This RWC had differences in populations and data availability as described in the main and Supplementary Methods. These RWC specific RMs, termed RWC-RMs were tested across the different Rules for performance characteristics as above. A subset of the original RM_G,II_ risk features for Adverse, Intermediate, and Favorable risk populations was identified based on available features in the RWC (Figure 6A). Supplementary Figures 20-21 illustrate the distribution of patients with observed outcomes across risk groups for the RWC-RM_G,II_ and ELN22. The RWC-RM_G,II_ demonstrated somewhat less *equitable* distribution than the parental RM_G,II_ using the academic center cohort but with strong *separation* of OS in both the RWC FAS and RWC allo-HCT excluded recipients (Figure 6A-6B). The RWC-RM_G,II_ showed lower empirical survival probabilities for the Adverse risk group than ELN22 while achieving significant conformity with ELN22 in assigning patients to this group (Figure 6C-6D). For OS risk *prediction*, it demonstrated a higher cAUC and iAUC than an ELN22-based approach, with median cAUC values of 0.62 (95% CI: 0.60, 0.75) versus 0.52 (95% CI: 0.50, 0.59); see Figure 6E-6F. The performance for RM incorporating Rules I, III, and IV for the RWC as well as a comparison to ELN22 roughly paralleled those for the academic cohort albeit again with different specific risk features identified for each RWC-RM (Supplemental Figures 22-25). This further demonstrates the context-specific adaptability and transportability of the proposed strategy.

**Figure 6.**
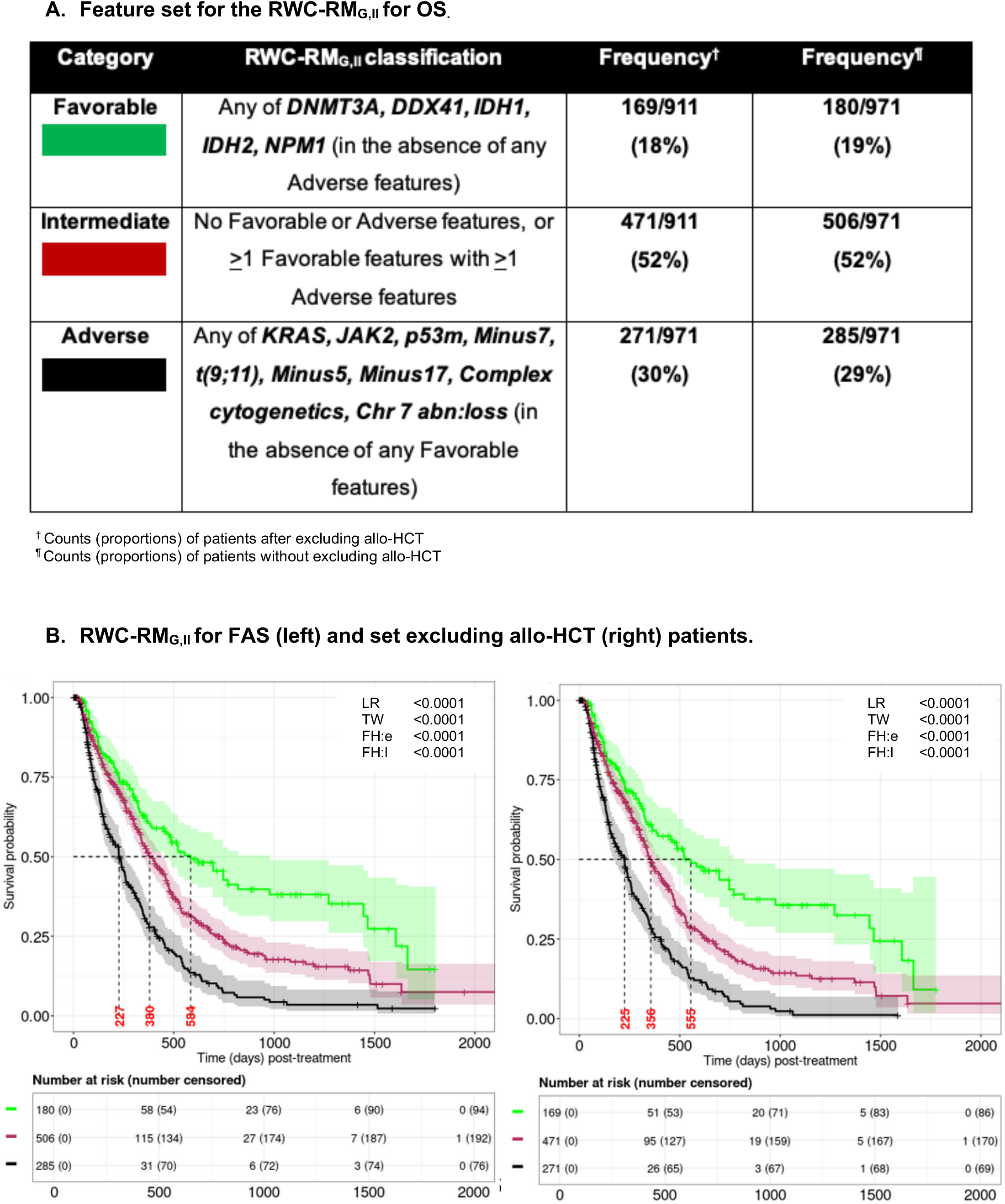
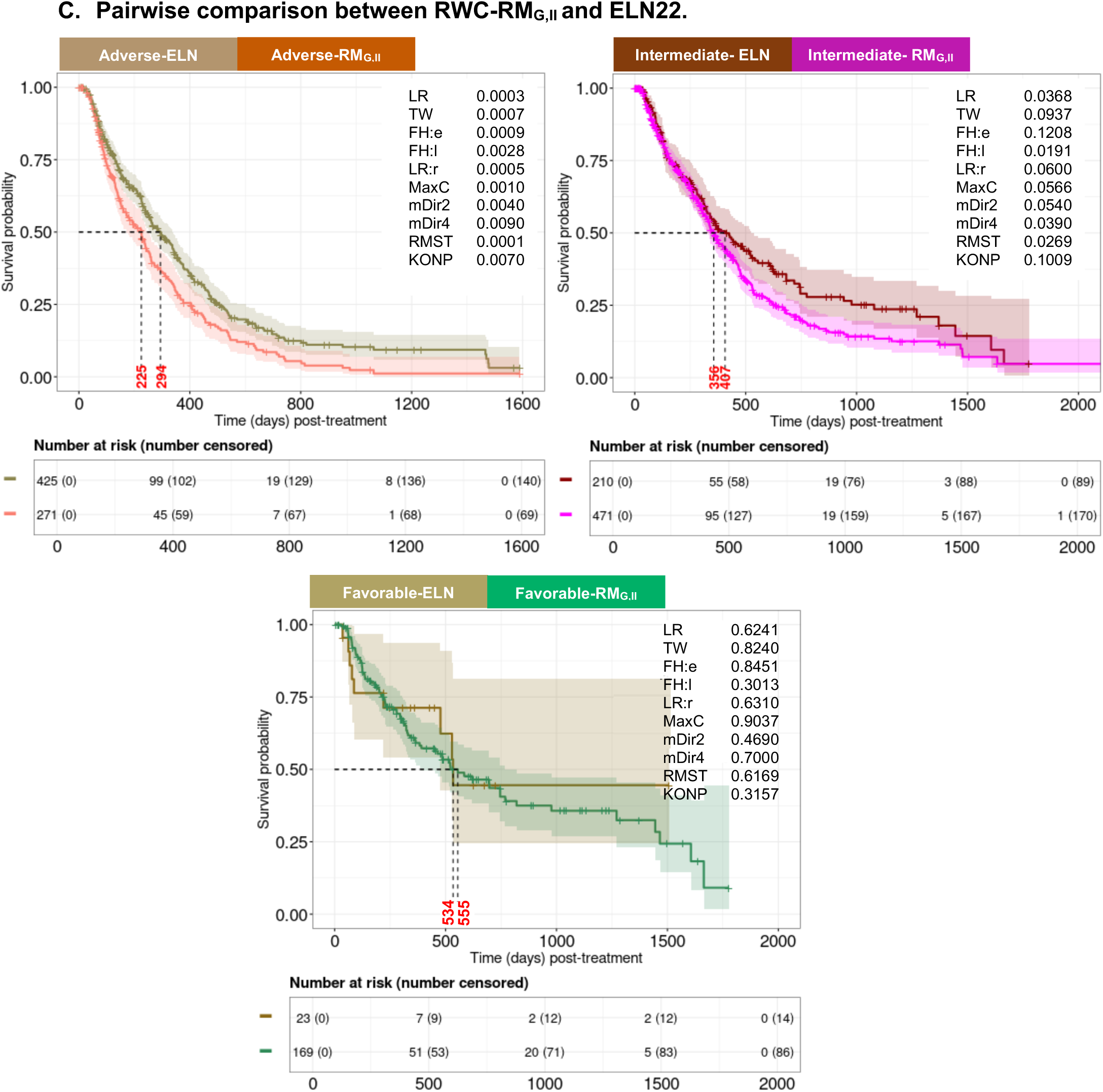
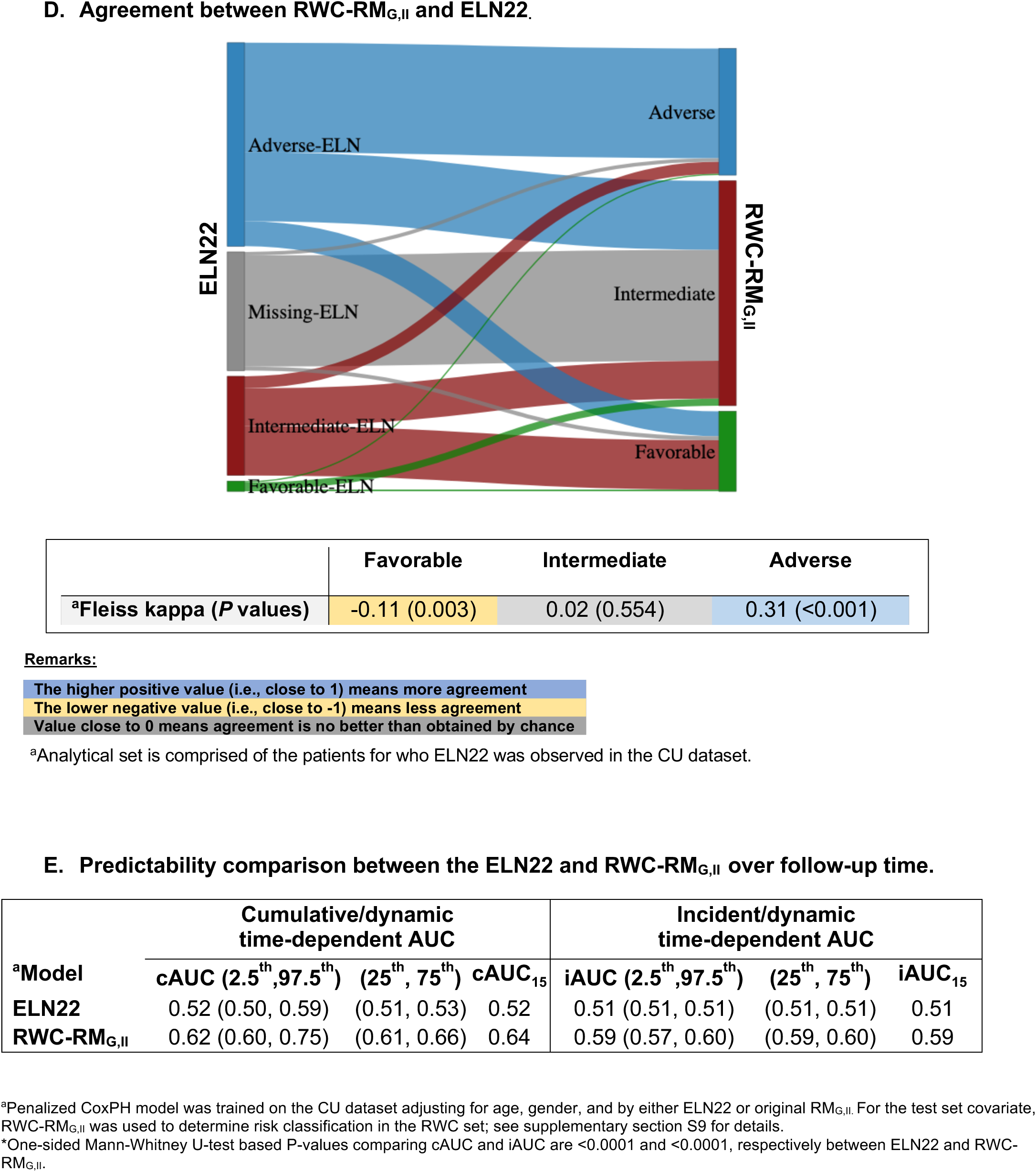
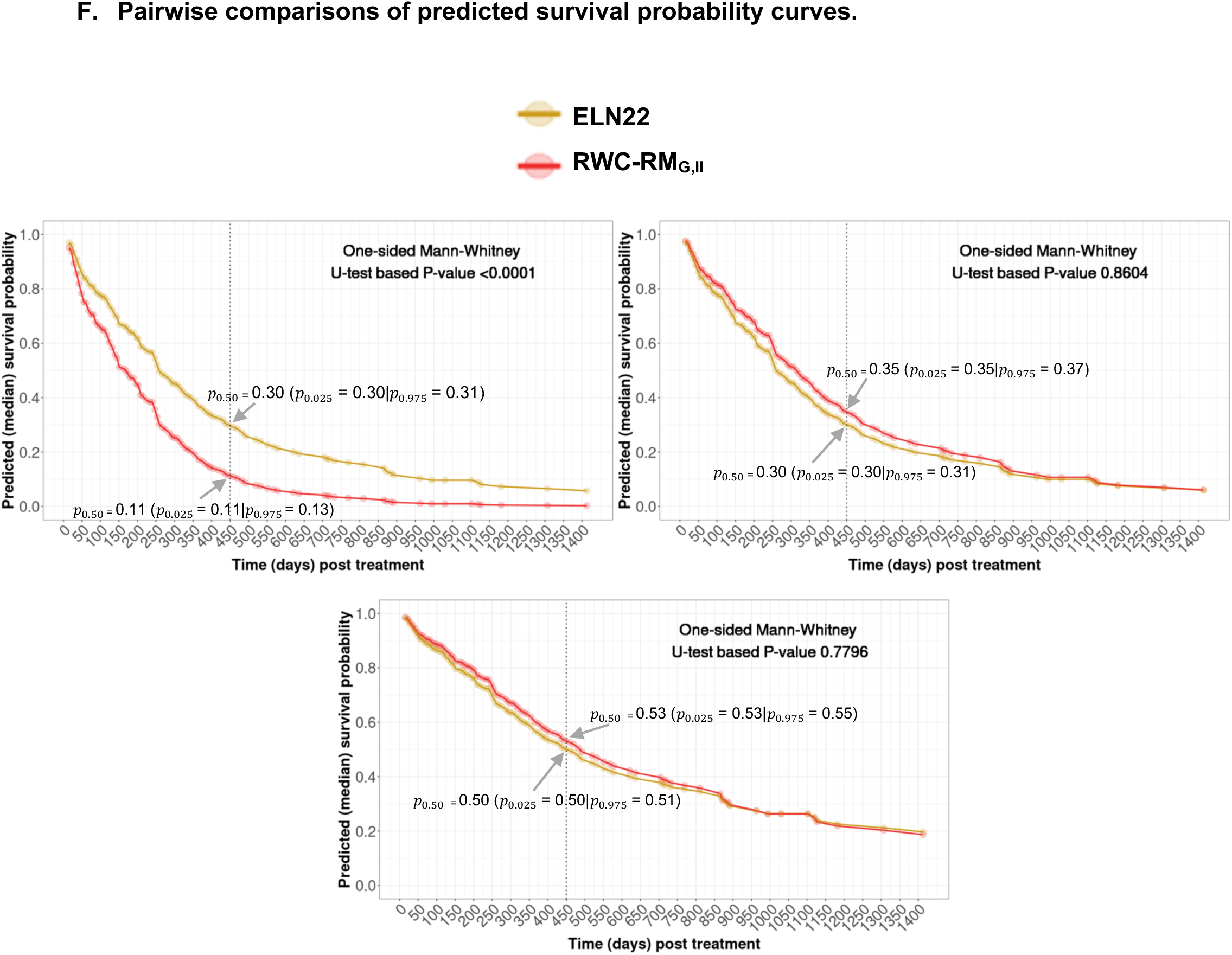
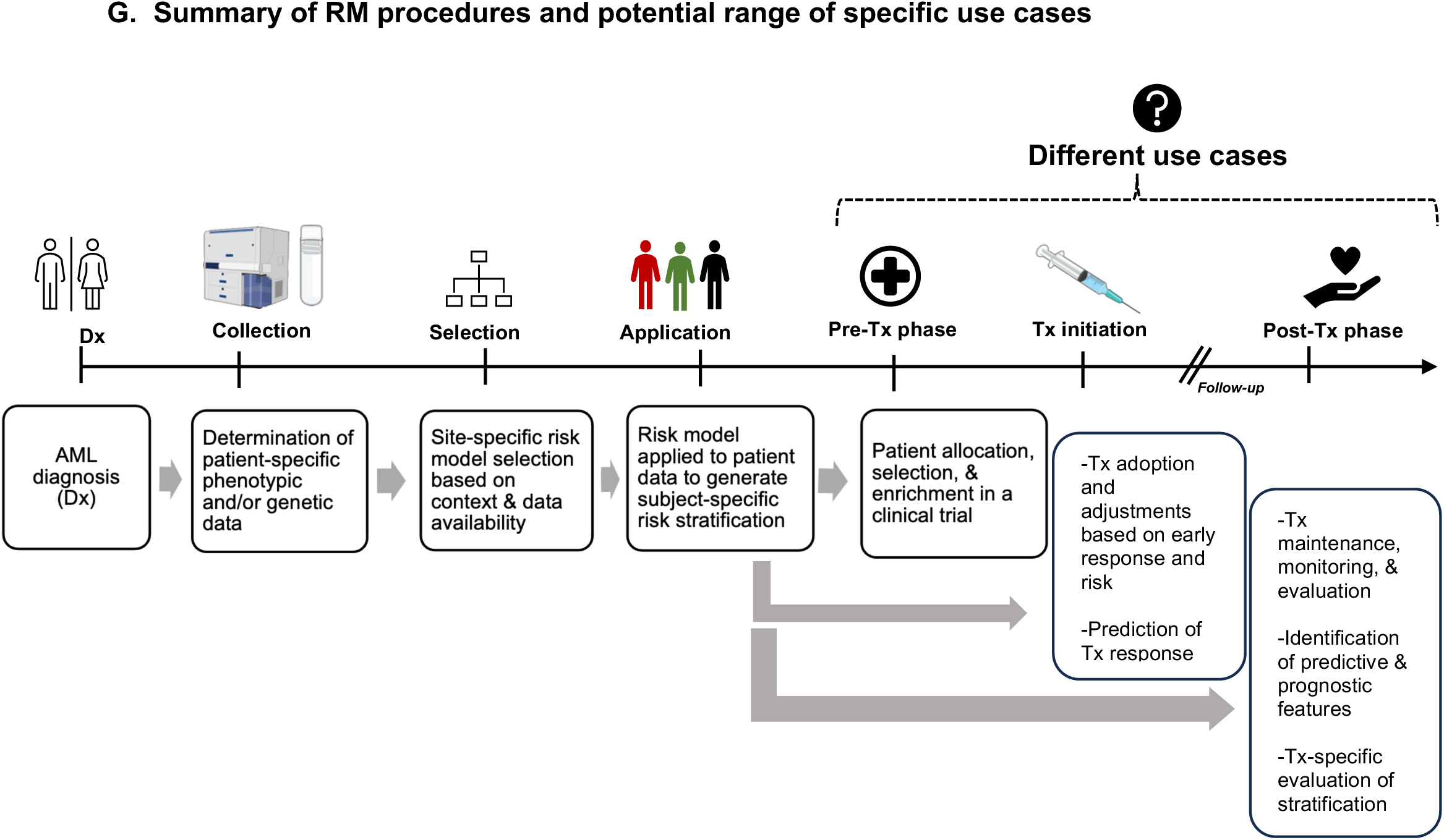
External validation for the RM_G,II_ on the real-world cohort (RWC) with respect to overall survival (OS) A) Definitions of features for the RWC-RM_G,II_. The category column includes color legends for reference. Biomarkers such as *BCOR, SF3B1, SMC1A, KMT2A, PTPN11, Chr 8 abn:loss, DEK:loss, MECOM:gain,* and *RARA:loss* were unavailable in the RWC. *Chromosome 7 deletion* (i.e., *Minus 7*) was used as a proxy of *Chromosome 7q deletion* (i.e., *Del7Q*); B) OS based on the RWC-RM_G,II_ with respect to the full analytical set (FAS) (left) and the set excluding allo-HCT (right) patients; C) Pairwise comparison between the ELN 2012 and RWC-RM_G,II_ for the Adverse, Intermediate, and Favorable risk groups in the cohort excluding allo-HCT patients; D) Comparison of inter-risk group conformity between the RWC-RM_G,II_ and ELN22 based on the FAS; E) Predictability of OS based on the RWC-RM_G,II_ and ELN22 over follow-up time up to four years. Composite molecular definitions used in classifying risk groups in the RWC are defined in Supplemental Methods section; F) Pairwise comparisons of predicted survival probability curves between the ELN 2012 and RWC-RM_G,II_ for the Adverse, Intermediate, and Favorable risk groups; G) A flowchart illustrating the practical application of the ML strategy to different potential use cases, note the availability and requirements of risk stratification features may vary greatly across the different use cases leading to the need for context dependent specific RMs.

## DISCUSSION

We developed a novel ML-based risk stratification strategy for AML patients that was designed to address a variety of RWD challenges and be flexible and adaptable to varying use cases. Having a common strategy is important for RM development to provide a consistent, objective framework for developing RMs across different populations, data elements, and applications. Unique features of the ML strategy include: (1) it was based on probabilistic RDs, (2) dealt with feature interactions using a logic-based approach, (3) addressed dynamic effects of features, (4) supported development of different specific RMs for different endpoints, (5) could account for important confounding effects (e.g., allo-HCT), and 6) included various features adjusting for collinearity. We deployed multiple objective metrics to assess internal performance and cross model comparisons to define the performance of RMs derived from this strategy. These metrics provide insights into model-specific effectiveness, while demonstrating the impact of modeling assumptions and rules on risk classifications. Testing this strategy showed: 1) proof of principle for using counterfactual dynamic estimation to develop RMs in AML, 2) confirmation that risk features depend on context, 3) adaptability of RMs to different modeling assumptions and rules, 4) adaptability to varying data types, endpoints, and use cases, and 5) ability to tune and generate specific RMs through coding, parameter, and feature modifications to accommodate changes in evolving diagnostics, assumptions, and rules. This capability is particularly important in AML, where advances in clinical care, treatment, disease biology, and testing modalities have yielded rapidly changing datasets. At a population level, unlike ELN22, the ML-based strategy effectively generated RMs tailored to specific use cases that demonstrated robust performance across the range of metrics and comparators used in both single academic center and RWC for retrospective analysis. Figure 6G summarizes how this strategy may be deployed to generate RMs for a variety of other potential use cases occurring before, during, and after treatment, including rapid pre-therapy patient allocation and more deliberate post-therapy retrospective analysis for discovery work (see S11 for additional details). To support testing and development, the code for applying this strategy to various datasets and applications has been made publicly accessible; refer to the Code Sharing section. Please note that this code is intended for research and exploratory purposes only; extensive testing and validation on specific RMs is essential before applying them in practice.

There are several limitations to this study. The simplification of genetic, FC, FISH, and composite AML mutation features into binary variables for numerical convenience and interpretability may lead to information loss and overlook critical biological aspects of AML. Similarly, treating all composite mutation categories for RUNX and other features (Supplemental Table 1E) as equally biologically relevant may lead to an oversimplification. Reintroducing the complexity of the constituents of these features may enhance RM stratification ability, offering additional clinical insights; however, future studies with larger datasets are needed for validation of such insights. Although RMs integrating genetic-and-phenotypic features were developed, lack of FC standardization across centers indicates that the models may be most applicable in single-center or small consortium settings with consistent FC datasets. There is also a risk of underreporting samples with dimly expressed FC features (e.g., CD45). Some RMs showed suboptimal stratification for allo-HCT recipients, which may be improved by developing RMs with features specific to this cohort. Using the same dataset for variable selection, risk classifications, and internal validation may cause overfitting. Additionally, all models, particularly those with higher stringency, risk omitting important features due to underestimated effects, small samples, and prespecified cut-offs for prevalence rates and RDs. In contrast, less stringent RMs may incorporate noisy features with spurious associations. To optimize RM’s performance, it is essential to not only address the above limitations but also to employ a flexible non-parametric approach, such as tree-or deep-learning-based survival models, coupled with a robust feature selection scheme. Lastly, this model was solely based on ven/aza treated patients so its further transportability to other treatments, including different lower intensity therapies or ven/aza based triplets, will need to be independently tested. Additional testing and validation with external, diverse datasets from multiple institutions, through prospective studies, and via comparison with published findings and emerging RMs are critical next steps to further evolve this strategy and the RMs generated from it^21,23,25^.

## CONCLUSION

In summary, we developed a novel ML-based risk stratification strategy for AML patients treated with ven/aza. We generated and tested a range of specific RMs that exhibited robust performance across various data elements and endpoints with respect to important metrics, demonstrating adaptability to a distinct external RWC use case. This indicates the proposed strategy’s effectiveness and the RMs’ ability to generalize across different contexts. This ML-based strategy may also be applicable to other AML use cases and diseases beyond AML.

## Supporting information

Supplementary Sections

## ACKNOWLEDGMENTS

The authors gratefully thank Austin E. Gillen from the Department of Medicine, University of Colorado Anschutz Campus, Aurora, Colorado; in addition, Bin Yao and Adam George from OncoVerity, Aurora, Colorado; and Grant Weller from RefinedScience, Aurora, Colorado for their feedback and reviews during this project.

## AUTHOR CONTRIBUTIONS

CAS and NI designed the study and drafted the manuscript. JLD, JSR, KS, JWC, FRM, and UVK processed and pulled the structured analytical datasets. CAS, NI, JSR, JLD, JZ, KS, JWC, and LW assessed the validity and quality of data. NI performed numerical analyses. CAS, NI, CTJ, JSR, JLD, JZ, MB, SES, MG, BMS, WMS, MRR, and KLE interpreted the results of the analyses. All authors reviewed, provided constructive comments, and agreed to its publication.

## SUPPLEMENTARY MATERIAL

Supplementary material is available along with the submission detailing additional methods and numerical results pertinent to the study.

## DATA AVAILABILITY

The raw, individual patient data are protected and not available due to data privacy laws. The processed data are available at reasonable request to the corresponding author.

## CODE SHARING

We simulated a synthetic survival dataset consisting of 200 subjects, distinct from the original analytical set, to illustrate the proposed strategy for developing risk models. This simulated dataset was used to implement the methodology, and the implementation details are provided in an HTML file included as a supplementary document. The corresponding R codes are incorporated within the HTML file and are also available in a public GitHub repository https://github.com/RefinedScience/Risk_Stratification.

## ABBREVIATIONS

AML: Acute myeloid leukemia
IC: Intensive chemotherapy
allo-HCT: Allogeneic hematopoietic cell transplant
ELN: European Leukemia Network
CR: Complete response
BR: Best response
CRi: CR with incomplete hematologic recovery
CRh: CR with partial hematologic recovery
MLFS: Morphologic leukemia free state
PR: Partial remission
CYT: Cytogenetic
FC: Flow cytometric
FISH: Fluorescence in situ hybridization
NGS: Next generation sequencing
PCR: Polymerase chain reaction
ven/aza: Venetoclax plus azacytidine
FAS: Full analytical set
EFS: Event-free survival
OS: Overall survival
IRB: Institutional review board
UC: University of Colorado
KM: Kaplan-Meier
CI: Confidence interval
NHST: Null hypothesis significance testing
LR: Log-rank
TW: Tarone-Ware
FH: Fleming-Harrington
mdir: Weighted multiple direction
MC: Max-Combo
KONP: K-sample omnibus non-proportional hazard
RMST: Restricted mean survival times
ML: Machine learning
mCOXr: Multivariable Cox proportional hazard models with ridge (L2-norm) penalty
PH: Proportional hazard
FRWB: Fractional random weight bootstrap
aHR: Adjusted hazard ratio
CV: Cross-validation
KS: Kolmogorov-Smirnov
AUC: Area under the curve
ROC: Receiver operative characteristics
cAUC: Cumulative case dynamic control ROC
iAUC: Incident case dynamic control ROC
IBS: Integrated Brier scores
CC: Complete cases
IPW: Inverse probability weights
MICE: Multivariate imputation by chained equations
MR: Marginal risk
RM: Risk model
RD: Risk difference
RM_GP_: RM based on genetic-plus-phenotypic features with respect to OS
RM_G_: RM based on genetic features with respect to OS
RM_GP-EFS_: RM based on genetic-plus-phenotypic features with respect to EFS
RM_G-EFS_: RM based on genetic features with respect to EFS
RWD: Real world data
I: Rule-I
II: Rule-II
III: Rule-III
IV: Rule-IV

