## Supplementary Sections for "Development of a dynamic counterfactual risk stratification strategy for newly diagnosed acute myeloid leukemia patients treated with venetoclax and azacitidine"

### SUPPLEMENTARY MATERIALS

This section provides additional pertinent results related to the study. There was one Supplemental Methods section with twelve subsections (S1-S12), seven Supplemental Tables, and twenty-five Supplemental Figures. All sections, tables, and figures are referenced in the main body of the paper.

### SUPPLEMENTAL METHODS

#### S1. Description of analytical dataset

A locked dataset was generated from CU's periodically updated Electronic Health Record (EHR) system. All pertinent subject-level data were extracted, cleaned, and validated at a specific point in time, after which the dataset was "locked," meaning no further changes or updates were permitted. This method ensures the integrity, reproducibility, and reliability of analyses, as all results are derived from an unchanging dataset throughout the research process.

#### S2. Definitions of features

Patients diagnosed with acute promyelocytic leukemia (APL), bi-phenotypic leukemia, blast crisis chronic myeloid leukemia, or central nervous system involvement with acute myeloid leukemia (AML) at diagnosis were excluded from the study. The cohort of AML-diagnosed patients included in the analysis received treatment with venetoclax and azacitidine (ven/aza) either as part of standard care or within a clinical trial. In cases of composite mutations, results from multiple tests were consolidated; thus, patients with multiple positive test results were counted only once. For an instance, the feature “p53m” composite mutation is a combination of p53 abnormalities by NGS and losses or gains of 17p by FISH. Complex cytogenetics were defined as $\geq$3 karyotype abnormalities. FISH data were simplified by combining all chromosomal specific probes into "Chr Abn". FISH data for DEK and MECOM were not routinely available before mid-2019 (0 DEK results, 3/7 positive MECOM results) and thus were not used before that date. Data, including NGS and FC results, were simplified into binary values as described previously in Islam et al[27]. Additional feature nomenclature is provided in the respective figure legends and Supplemental Table 2.

**S3. Exploratory analyses and feature selection**

Univariate Kaplan-Meier (KM) analyses were performed for screening features. The corresponding 95% confidence intervals (CIs) and median survival times were reported. Zero-variation and low-count variables (i.e., ≤ 4 counts on any categorical label) were not included in the analyses. Univariate association filtering by null hypothesis significance testing (NHST) excluded noise variables. Two tailed-tests testing the quality of survival curves were evaluated using Log-rank (LR), Tarone-Ware (TW), Fleming-Harrington (FH) emphasizing early and late periods, weighted multiple direction (mdir) LR, Max-Combo (MC), K-sample omnibus non-proportional hazard (KONP), and restricted mean survival times (RMST) tests[25-31]. The corresponding P-values were reported. Different testing approaches have addressed complexities of violating proportionality assumptions in survival curves and differences over time. Variables with a P-value < 0.30 based on at least one univariate test in any KM analyses (overall survival (OS) and event-free survival (EFS)) were considered candidates for multivariate models that to be used for parameter and counterfactual profile estimation.

**S4. Counterfactual estimation strategy**

The mean predicted marginal risks (MR) (expressed as probabilities) of experiencing mortality (OS) or adverse events (EFS) in relation to wild-type (-ve) or mutated (+ve) conditions are determined through "*recycled*" predictions for each test type. Such estimation process is more robust as a fixed patient cohort was employed, adjusting for the mix of risk factors and severity. This approach utilizes the full analytical set, thereby reducing potential bias from sparse features or skewed distributions of patient characteristics. At each time point, the MRs of a biomarker being positive or negative were calculated by averaging the predicted MR probabilities for all patients, assuming they were (hypothetically) (+ve) or (-ve) for genetic features, or if they expressed a marker by FC. This method is referred to as marginal standardization or S-estimation in the literature, where the objective is to calculate the marginal effect of each genetic and phenotypic biomarker (i.e., the instantaneous effect on the predicted probability of response over time due to a change from a biomarker being (-ve) to (+ve) while keeping other variables constant) for each subject. Next a sample average of individual marginal effects is computed over a grid of pre-specified time points, resulting in overall marginal effects profile.

Let $\{\hat{O}(M^1)^{j}_{it_r(+ve)}; r = 1, \dots, T\}^{N}_{i=1}$ be the marginal estimation of relative risks for the $i$-th subject at the $t_r$-th time based on the $M^j$-th model for being (+ve) where $j = 1, \dots 5$ with the nominal values 1 to 5 referring to FC, CYT, FISH, NGS, and AML mutation test type, respectively. Here $N$ corresponds to the total number of subjects. The model-specific survival function is expressed as $S(t|\boldsymbol{X}_{FC}) = \exp\left(-\int_0^t M^{FC}(\boldsymbol{r}, \boldsymbol{X}_{FC})dr\right)$. The corresponding estimated cumulative distribution function for each subject with respect to the $l$-th FC variable being (+ve) over time is $\hat{F}_{i(+ve)}\left(t|\boldsymbol{X}_{FC_l,FC_{l'}}\right) = 1 - \hat{S}\left(t|\boldsymbol{X}_{i,FC_{l(+ve)}}, \boldsymbol{X}_{i,FC_{l' \neq l}}\right)$; here $l'$ refers to the set of all FC variables except the $l$-th one. We denoted this by $\hat{O}(M)^{FC_l}_{it(+ve)}$ in Figure 2. The mean marginal risk is defined by $\hat{\hat{O}}(M)^{FC_l}_{t(+ve)} = \sum_{i=1}^{N} \hat{O}(M)^{FC_l}_{it(+ve)} \; / \; N$ where all other FC variables except the $l$ −th remain fixed at their observed values. For bootstrap based counterfactual estimation (i.e., Rule-III and Rule-IV), we computed $\hat{F}_{ib(+ve)}\left(t|\boldsymbol{X}_{FC_l,FC_{l'}}\right)$ for $B$ times where $b = 1, \dots, B$ and then we averaged over $B$ estimates resulting in a bootstrap-based marginal risk profiled denoted by $\hat{\hat{\hat{O}}}(M)^{FC_l}_{t(+ve)}$. Using the similar intuition as above, we estimated dynamic marginal risk over time for the $l$-th FC feature being (-ve) and the process is repeated for other test types (i.e., NGS, CYT, FISH, Mutation).

#### S5. Details of evaluation criteria

*Equitability:* Summary statistics (counts and percentages) for each group were provided to assess balance.

*Separability:* KM analyses of OS and EFS and hypothesis testing were performed for the equality Log-rank and its weighted counterparts. Best responses (BRs) were evaluated for the proportion of CR/CRh across risk groups. The corresponding P-values testing the equality of proportions by Chi-Squared or Fisher-exact tests were provided.

*Conformity:* Pairwise comparison in survival curves between the ELN22 and RMs was conducted for each risk strata. Level of agreements between methods were assessed via Fleiss kappa and corresponding P-values testing random agreement were provided[42,43].

*Predictability:* Figure 3 summarizes the steps used to run internal validation for predictability of RMs. Step 1 in Figure 3 remains unchanged across the CV runs relative to Figure 2, while each CV run in Steps 2-5 of Figure 3 generated an independent list of risk stratification variables relative to the same steps in Figure 2. Four validation strategies (Step 6, Figure 3) were used to combine these results for covariate-level classification (Step 7, Figure 3): (a) using the original list of variables as defined prior to CV runs, (b) combining risk factors from each CV run based on the *majority-votes* approach across fifteen runs, (c) combining risk factors from each CV run and three missing data mechanisms (45 runs total), and (d) classifying risk factors separately for each CV run and a missing data method. Two levels of uncertainty were addressed: in the selection of risk stratification and in the time-to-event prediction for a specific RM. While strategy (a) focused primarily on predictive validation keeping risk stratification constant across CV runs, the other strategies (b-d) incorporated both selection and predictive validations at varying extents with (d) being the most stringent. Predictive performance was evaluated at unique follow-up times, at least five days apart, up to four years using the KM and model-based approaches treating allo-HCT patients as censored. In the KM approach, test CV patients were categorized as "Adverse", "Intermediate", or "Favorable" with survival probabilities generated using KM estimates from the training set. Similarly, the mCOXr model (adjusting for age, gender, race, and either ELN22 or a proposed RM) based classifications were generated. Note that the baseline features (i.e., age, gender, and race) were added to control for potential confounding effects and ensure risk variable's (RM or ELN22) predictions are not skewed by demographic factors. Results were evaluated on the test CV folds and compared using time-dependent area under the curve (AUC) of receiver operative characteristics (ROC) based on cumulative case dynamic control (cAUC), time-dependent AUC under ROC curve based on incident case dynamic control definition (iAUC), and integrated Brier scores (iBrier). Reported results were median (over CV folds) of medians (over time). P-values based on one-sided Mann-Whitney U-test comparing the distributions of iAUC and cAUC values over time between the proposed and ELN22 were reported; median (over CV folds) of P-values were reported.

**S6. Rationale of multiplicity of models**

A series of RMs were proposed emphasizing various aspects of real-world and clinical trial data, objectives, assumptions, and requirements. The "best" performing model is typically in part subjective, depending on the context and dataset where it is applied. The proposed approach differs from  one-size-fits-all approach, which assumes the model works well for all scenarios regardless of underlying differences in datasets. In contrast, the proposed strategy requires investigators to consider specific features and elements in the respective analytical dataset and choose model parameters accordingly. For example, for a site like CU, where the proportion of allo-HCT is high, adjustments for allo-HCT needs to be made. However, this may be redundant for sites where the proportion of allo-HCT is low or negligible. One also needs to choose how rigorously to account for missingness in the dataset which primarily depends on the magnitude and pattern of missingness. Furthermore, a user may prefer phenotypic-and-genetic models over genetic models alone. The proposed approach enables potential clinical implementation of a risk stratification methodology adapted to site-specific preferences and underlying patient populations.

**S7. External validation cohort for risk models**

The Flatiron Health dataset is a licensed, de-identified dataset comprising patient information on individuals diagnosed with acute myeloid leukemia (AML) using ICD-9 and ICD-10 codes. These individuals had a minimum of two documented clinical visits between January 2014 and December 2023 and received venetoclax/azacitidine as frontline therapy. During the study period, the de-identified data were sourced from approximately 280 cancer clinics across the United States, encompassing around 800 sites of care, primarily within community oncology settings. The data were de-identified and subject to obligations to prevent re-identification and protect confidentiality. For ease of notation, the real-world cohort from Flatiron Health is referred to as RWC throughout this paper.

**S8. Standardization and normalization for external validation**

OS was defined as same as before which is the time from ven/aza treatment initiation to all-cause mortality where alive patients were censored at their last known alive dates. Due to the absence of follow-up responses in the RWC, EFS based RMs (i.e., $RM_{G,I\text{-}EFS}$, $RM_{G,II\text{-}EFS}$, $RM_{G,III\text{-}EFS}$, $RM_{G,IV\text{-}EFS}$, $RM_{GP,I\text{-}EFS}$, $RM_{GP,II\text{-}EFS}$, $RM_{GP,III\text{-}EFS}$, and $RM_{GP,IV\text{-}EFS}$) were not validated externally.

We simplified the corresponding feature sets to calculate genetics and genetics-plus-phenotypic specific RMs by using only the features that were available in the RWC. For the ELN22 calculations using the CU dataset, NGS testing along with VAF score were used to categorize composite features (e.g., TP53 mutation) as positive or negative. In contrast, VAF information was unavailable in the RWC set and thus was not used in categorization. Additionally, bzip in-frame mutation for CEBPA for ELN “Favorable” risk was not assessed in the RWC due to the lack of information. In the RWC, monosomal karyotype for ELN “Adverse” risk group was not evaluated due to insufficient data. In addition, the term "Chromosome x deletion" refers to either partial or full chromosomal deletions in the RWC, whereas the CU karyotype data differentiates between partial and full deletions or other abnormalities. In calculating the ELN22 and other CU-based risk models using the RWC, Chromosome 7 deletion was used as a proxy measure for Chromosome 7q deletion due to its unavailability. Note that only 21 karyotype abnormalities were provided in the RWC, in contrast the CU dataset was comprised of complete karyotype information. In the RWC, patient-specific genetic testing data for CSF3R, DDX41, DNMT3A, KIT, IDH1, IDH2, JAK2, KRAS, MPL, NPM1, TP53 were provided on a per marker basis using next-generation sequencing (NGS), polymerase chain reaction (PCR), or unknown (other/unrecorded) test types. Similarly, Inv3, T9;11, Minus5, Minus7, Minus17 were detected using cytogenetics/karyotyping (CYT), FISH, or unknown (i.e., other/unrecorded abbreviated as UNK) test types. Phenotypic biomarkers like CD19, CD34, and CD36 were assessed by flow-cytometry (FC), Immunohistochemistry (IHC), or unknown test types. As we applied the RMs on the RWC, we used the composite definitions for the aforementioned biomarkers utilizing NGS, PCR, FISH, FC, IHC, and UNK test types to ensure completeness.

**S9. Numerical assessment for external validation**

To account for missingness, we applied both the *imputation-by-mode* and MICE approach. The model-based MICE imputation uses logistic and polytomous ridge regressions and imputes values multiple times to account for uncertainty. Since the proportion of missingness is large in the RWC for many biomarkers, the use of RWC alone in the MICE imputation models is not sufficient. To resolve this sparsity, we used a pooled analytical dataset to impute missing biomarker values in the RWC cohort by aggregating the CU and RWC together resulting in a larger sample size; this pooled approach is more reliable as it borrows information leveraging the underlying shared patterns in the distribution of patient characteristics across two sources. Some features have high proportion of missingness (approximately >40%) and were only available in the RWC (e.g., FC observed by IHC test type). These RWC features were not imputed by model-based approach due to the risk of unreliable missing value estimation.

However, instead of excluding them from the feature list in classifying subject-level classification, these features were estimated as *imputation-by-mode* approach as a practical and convenient solution.

KM analyses, exploratory analyses with summary statistics, and null hypothesis significance testing (NHST) based on the RWC were performed to assess *equitability*, *conformity*, and *separability* as before; reported are the results based on the *imputation-by-mode* approach unless otherwise specified. Analyses with respect to 10 imputed analytical sets were performed similarly and exhibited similar phenomenon across all these analytical sets. To assess *predictability*, the penalized CoxPH models were trained on the CU dataset, adjusting for age, gender, and risk groups (Adverse, Intermediate, Favorable) defined by either ELN22 or parental RM. For evaluation in the RWC test set with the proposed model, risk level was first assigned using the RWC-RMs and then the corresponding trained models were applied to evaluate survival AUCs over time; see Supplemental Figure 24. While this method preserves semantic consistency in the RWC, it may introduce differences in the composition and distribution of risk classification owing to definitional adjustments needed for missing features. Indeed, this approach reflects the practical realities of model application, shows how the models can be adapted in different real-world situations where information is inevitably missing, and assesses the transportability of the corresponding parental RMs. For sensitivity, we retrained the penalized CoxPH models adjusting for age, gender, and RWC-RM (constructed using only features available in both the CU and RWC datasets) to quantify any differences introduced by feature reduction and definitional modification; the pertinent predictive results are provided in Supplementary Figure 25. Note that the variable defining race of patients was missing in the RWC and thus was not added as a confounding feature in the mCOXr predictive model.

Reported are the 2.5th, 25th, 50th, 75th, and 97.5th percentile values of AUCs with respect to cumulative case/dynamic control (cAUC) receiver operator curves over time. Evaluations are enumerated at discrete unique follow-up times of the CU dataset that are at-least 5-days apart; $cAUC_{15}$ refers to cAUC value corresponding to 450 days (i.e., ~14.7 months).

**S10. Numerical challenges in missing data imputation**

The numerical performance of model-based imputation is influenced by the extent and severity of missing data, the completeness of the dataset, and the pattern of missingness in the RWC which was assumed to be missing at random (MAR). If the imputed model's results overestimate missing biomarkers, such as tp53m and NPM1, it can significantly impact risk classification at the subject level. This may lead to a skewed distribution across the Adverse, Intermediate, and Favorable risk groups, with the majority of AML patients being categorized as Intermediate. This occurs because these patients possess at least one Adverse feature and one Favorable feature.

**S11. Practical application**

Figure 6F illustrates the use of a RM in a clinical environment; while we illustrate this with respect to a version of the $RM_{G,II}$. Indeed, this illustration is RM agnostic and thus can be applied for any RM in general. When diagnosing a patient with AML, it is crucial to identify, gather, and document diagnostic biomarker information in a machine-readable structured format for analysis. Following this, a site selects context-specific parameters based on its preferences and goals. A site can either adopt one of the RMs mentioned in the paper or modify codes to develop a customized, site-specific RM. For instance, if a site aims to maximize OS differences at 2 years instead of 14.7 months, it should choose biomarkers that exhibit risk differences (RDs) of -5% or 5% at 2 years (or approximately 730 days) and then construct RMs accordingly. Additionally, a site may opt to set a stricter (or more relaxed) RD threshold, such as -10% or

10%, to select genetic and/or phenotypic features in classifying risk groups. Note that a tighter RD threshold may result in a sparser and more limited list of features. Furthermore, depending on the objectives, availability, and resources, a site might prefer genetic models over phenotypic-and-genetic ones. Similarly, based on the extent and pattern of missing data and the proportion of allo-HCT, a site should either choose one of the reported models or develop its own by adjusting the relevant context-specific parameters according to the proposed strategy.

**S12. Computational details and codes**

A list of R packages used for computation and implementation is stipulated in the HTML file detailing the implementation of the proposed methodology. In addition,15-fold cross-validation to evaluate predictive performances, fractional random weight bootstrap (FRWB) to estimate uncertainty, and counterfactual estimation to generate risk differences over time were generated using custom R codes. The corresponding R functions are provided in the HTML illustration file and also in a public GitHub repository https://github.com/RefinedScience/Risk_Stratification.
We generated a synthetic survival dataset for 200 subjects (which is completely unrelated to the original analytical set) and illustrated the proposed strategy to develop risk models using this simulated dataset for the purpose of reproducibility.

| | |
|---|---|
| *Testing the equality of survival curves to generate P-values* | *TSHRC, survRM2, survRM2perm, mdir.logrank, KONPsurv, RBT4TCSC, nph, Mvtnorm* |
| *Evaluating numerical performances to compute survival predictability measures* | survivalROC, risksetROC, SurvMetrics, irr |
| *Fitting penalized mCOXr and mLRr models* | survMisc, glmnet, survival |
| *Optimizing computation to distribute work loads* | doParallel, foreach |
| *Generating survival curves and plots* | ggplot2, survminer |

### SUPPLEMENTAL TABLE LEGENDS

**Supplemental Table 1. Summary statistics and data distribution for candidate prognostic risk features.** A) Demographics; B) Next Generation Sequencing (NGS); C) Cytogenetics; D) Fluorescence in situ hybridization (FISH); E) "AML composite mutations" where genetic features defined by multiple test types including NGS, FISH and PCR were bundled together in a single feature definition; and F) Flow cytometry (FC). In the FISH data, "PML" = PML/RARA and "RARA" = RARA break-apart assays.

**Supplemental Table 2. Definition of genetic and phenotypic features.**

**Supplemental Table 3. Penalized regressions based adjusted hazard ratios (aHRs) of prognostic factors for time-to-mortality (Overall survival: 0 = alive/censored and 1 = mortality) and adjusted odds ratios (aORs) for CR/CR$_h$ as best response (binary: 0 = CR/CR$_h$ and 1 = Non CR/CR$_h$).** Bootstrap based 95% confidence intervals and summary statistics (counts and median time-to-event) for potential model features are reported based on A) Next Generation Sequencing (NGS); B) Cytogenetics; C) Fluorescence in situ hybridization (FISH); D) AML specific composite mutations; and E) Flow cytometry (FC). Reported are the results corresponding to the model (M2) in Table 1 for aHRs which are computed based on three analytical datasets - "*allo-HCT*" refers to the full analytical set treating allo-HCT patients as censored, "*ex-allo-HCT*" refers to the set excluding allo-HCT patients, and "*All*" refers to the full analytical set (FAS). aORs were calculated based on the set for which "best response" was available.

**Supplemental Table 4. Penalized regressions based adjusted hazard ratios of prognostic factors for time-to-event (event free survival: 0 = alive without event/censored and 1 =**

**event).** Bootstrap based 95% confidence intervals and summary statistics (counts and median time-to-event) for potential model features are based on A) Next Generation Sequencing (NGS); B) Cytogenetics; C) Fluorescence in situ hybridization (FISH); D) AML specific composite mutations; and E) Flow cytometry (FC). Definition of an event is defined in Figure 1. Reported are the results corresponding to the model (M8) in Table 1 for aHRs which are computed based on three analytical datasets - "*allo-HCT*" refers to the full analytical set treating allo-HCT patients as censored, "e*x-allo-HCT*" refers to the set excluding allo-HCT patients, and "*All*" refers to the full analytical set.

**Supplemental Table 5. Covariate-level risk classification based on different variable selection rules.** A) Relationship between different covariate selection rules; B) Rule-I: Variables selected based on point estimates; C) Rule-II: Variables selected based on point estimates excluding variables with low (< 2%) and high (>40%) prevalence for (+ve) cases; D) Rule-III: Variables selected based on bootstrap-based point estimates; E) Rule-IV: Variables selected based on bootstrap-based point estimates and excluding variables with low (< 2%) and high (>40%) prevalence for (+ve) cases.

**Supplemental Table 6. Predictive validation of risk stratification based on counterfactual point estimates for overall survival (OS).** A) Rule-I: Variables selected based on point estimates; B) Rule-II: Variables selected based on point estimates excluding variables with low (< 2%) and high (>40%) prevalence for (+ve) cases; C) Rule-III: Variables selected based on bootstrap-based point estimates; D) Rule-IV: Variables selected based on bootstrap-based point estimates and excluding variables with low (< 2%) and high (>40%) prevalence for (+ve) cases. Median of cAUCs (M-cAUC) and median of iAUCs (M-iAUC) over follow-up time were reported.

**Supplemental Table 7. Predictive validation of risk stratification based on counterfactual point estimates for event free survival (EFS).** A) Rule-I: Variables selected based on point estimates; B) Rule-II: Variables selected based on point estimates excluding variables with low (< 2%) and high (>40%) prevalence for (+ve) cases; C) Rule-III: Variables selected based on bootstrap-based point estimates; D) Rule-IV: Variables selected based on bootstrap-based point estimates and excluding variables with low (< 2%) and high (>40%) prevalence for (+ve) cases. Median of cAUCs (M-cAUC) and median of iAUCs (M-iAUC) over follow-up time were reported.

**Supplemental Table 1A. Summary statistics for patient demographics.**

| | Total | Alive | Deceased | SMD |
|---|---|---|---|---|
| **N =** | 316 | 104 | 212 | |
| **Demographics** | | | | |
| **Age, Median (IQR) [Range], y** | 71.0 (11.0) [69.0] | 67.0 (12.0) [66.0] | 73.0 (11.0) [67.0] | 0.52 |
| **Age, n (%)** | | | | 0.48 |
| **[18, 50)** | 26 (8.2) | 14 (13.5) | 12 (5.7) | |
| **(50, 60]** | 23 (7.3) | 10 (9.6) | 13 (6.1) | |
| **(60, 70]** | 103 (32.6) | 39 (37.5) | 64 (30.2) | |
| **(70, 80]** | 123 (38.9) | 35 (33.7) | 88 (41.5) | |
| **>80** | 41 (13.0) | 6 (5.8) | 35 (16.5) | |
| **Female, n (%)** | 145 (45.9) | 43 (41.3) | 102 (48.1) | 0.14 |
| ***Race, n (%)** | | | | 0.22 |
| **Black** | 11 (3.5) | 4 (3.8) | 7 (3.3) | |
| **Other** | 2 (0.6) | 2 (1.9) | 0 (0.0) | |
| **White** | 264 (83.5) | 85 (81.7) | 102 (48.1) | |
| ***Ethnicity, n (%)** | | | | 0.02 |
| **Non-Hispanic** | 281 (88.9) | 92 (88.5) | 189 (89.2) | |
| **Hispanic** | 23 (7.3) | 8 (7.7) | 15 (7.1) | |
| **Secondary AML, n (%)** | 111 (35.1) | 21 (20.2) | 90 (42.5) | 0.49 |
| **Prior MDS, n (%)** | 16 (5.1) | 4 (3.8) | 12 (5.7) | 0.09 |
| ***ECOG** | | | | 0.27 |
| **0** | 48 (15.2) | 20 (19.2) | 28 (13.2) | |
| **1** | 112 (35.4) | 36 (34.6) | 76 (35.8) | |
| **2+** | 42 (13.3) | 10 (9.6) | 32 (15.1) | |
| ***ELN risk group, n (%)** | | | | 0.27 |
| **Favorable** | 54 (17.1) | 21 (20.2) | 33 (15.6) | |
| **Intermediate** | 50 (15.8) | 22 (21.2) | 28 (13.2) | |
| **Adverse** | 208 (65.8) | 60 (57.7) | 148 (69.8) | |

*Standardized mean differences (SMD) were calculated after excluding missing cases. SMD > 0.10 refers to systematic differences between alive and deceased patients in the sample.

**Supplemental Table 1B. Summary statistics for next generation sequencing (NGS) features.**

| | | Total | Alive | Deceased | *SMD |
|---|---|---|---|---|---|
| **N =** | | 316 | 104 | 212 | |
| **NGS, n (%)** | | | | | |
| **ASXL1** | (-ve) | 231 (73.1) | 78 (75.0) | 153 (72.2) | 0.10 |
| | (+ve) | 63 (19.9) | 18 (17.3) | 45 (21.2) | |
| **BCOR** | (-ve) | 274 (86.7) | 86 (82.7) | 188 (88.7) | 0.20 |
| | (+ve) | 20 (6.3) | 10 (9.6) | 10 (4.7) | |
| **BCORL1** | (-ve) | 277 (87.7) | 90 (86.5) | 187 (88.2) | 0.05 |
| | (+ve) | 11 (3.5) | 3 (2.9) | 8 (3.8) | |
| **CSF3R** | (-ve) | 286 (90.5) | 92 (88.5) | 194 (91.5) | 0.05 |
| | (+ve) | 5 (1.6) | 2 (1.9) | 3 (1.4) | |
| **CEBPA** | (-ve) | 271 (85.8) | 88 (84.6) | 183 (86.3) | 0.07 |
| | (+ve) | 21 (6.6) | 8 (7.7) | 13 (6.1) | |
| **DNMT3A** | (-ve) | 230 (72.8) | 69 (66.3) | 161 (75.9) | 0.21 |
| | (+ve) | 63 (19.9) | 26 (25.0) | 37 (17.5) | |
| **DDX41** | (-ve) | 140 (44.3) | 56 (53.8) | 84 (39.6) | 0.46 |
| | (+ve) | 6 (1.9) | 6 (5.8) | 0 (0.0) | |
| **ETV6** | (-ve) | 281 (88.9) | 92 (88.5) | 189 (89.2) | 0.02 |
| | (+ve) | 13 (4.1) | 4 (3.8) | 9 (4.2) | |
| **EZH2** | (-ve) | 279 (88.3) | 92 (88.5) | 187 (88.2) | 0.09 |
| | (+ve) | 16 (5.1) | 4 (3.8) | 12 (5.7) | |
| **GATA2** | (-ve) | 276 (87.3) | 90 (86.5) | 186 (87.7) | 0.02 |
| | (+ve) | 13 (4.1) | 4 (3.8) | 9 (4.2) | |
| **IDH1** | (-ve) | 274 (86.7) | 84 (80.8) | 190 (89.6) | 0.27 |
| | (+ve) | 24 (7.6) | 13 (12.5) | 11 (5.2) | |
| **IDH2** | (-ve) | 253 (80.1) | 78 (75.0) | 175 (82.5) | 0.17 |
| | (+ve) | 46 (14.6) | 19 (18.3) | 27 (12.7) | |
| **JAK2** | (-ve) | 284 (89.9) | 95 (91.3) | 189 (89.2) | 0.16 |
| | (+ve) | 12 (3.8) | 2 (1.9) | 10 (4.7) | |
| **KIT** | (-ve) | 289 (91.5) | 94 (90.4) | 195 (92.0) | 0.04 |
| | (+ve) | 5 (1.6) | 2 (1.9) | 3 (1.4) | |
| **KRAS** | (-ve) | 282 (89.2) | 94 (90.4) | 188 (88.7) | 0.16 |
| | (+ve) | 12 (3.8) | 2 (1.9) | 10 (4.7) | |
| **MPL** | (-ve) | 283 (89.6) | 93 (89.4) | 190 (89.6) | 0.08 |
| | (+ve) | 5 (1.6) | 1 (1.0) | 4 (1.9) | |
| **NF1** | (-ve) | 136 (43.0) | 58 (55.8) | 78 (36.8) | 0.07 |
| | (+ve) | 11 (3.5) | 4 (3.8) | 7 (3.3) | |
| **NPM1** | (-ve) | 235 (74.0) | 74 (71.2) | 161 (75.9) | 0.09 |
| | (+ve) | 60 (19.0) | 22 (21.2) | 38 (17.9) | |
| **NRAS** | (-ve) | 265 (83.9) | 86 (82.7) | 179 (84.4) | 0.03 |
| | (+ve) | 30 (9.5) | 9 (8.7) | 21 (9.9) | |

**Supplemental Table 1B, cont'd**

| | | Total | Alive | Deceased | *SMD |
|---|---|---|---|---|---|
| **N =** | | 316 | 104 | 212 | |
| **NGS, n (%)** | | | | | |
| **PHF6** | (-ve) | 278 (88.0) | 90 (86.5) | 188 (88.7) | 0.01 |
| | (+ve) | 12 (3.8) | 4 (3.8) | 8 (3.8) | |
| **PTPN11** | (-ve) | 275 (87.0) | 93 (89.4) | 182 (85.8) | 0.22 |
| | (+ve) | 19 (6.0) | 3 (2.9) | 16 (7.5) | |
| **RAD21** | (-ve) | 281 (88.9) | 91 (87.5) | 190 (89.6) | 0.04 |
| | (+ve) | 8 (2.5) | 3 (2.9) | 5 (2.4) | |
| **RUNX1** | (-ve) | 236 (74.7) | 73 (70.2) | 163 (76.9) | 0.14 |
| | (+ve) | 59 (18.7) | 23 (22.1) | 36 (17.0) | |
| **SETBP1** | (-ve) | 285 (90.2) | 92 (88.5) | 193 (91.0) | 0.09 |
| | (+ve) | 9 (2.8) | 4 (3.8) | 5 (2.4) | |
| **SF3B1** | (-ve) | 268 (84.8) | 85 (81.7) | 183 (86.3) | 0.11 |
| | (+ve) | 19 (6.0) | 8 (7.7) | 11 (5.2) | |
| **SH2B3** | (-ve) | 140 (44.3) | 59 (56.7) | 81 (38.2) | 0.09 |
| | (+ve) | 6 (1.9) | 3 (2.9) | 3 (1.4) | |
| **SMC1A** | (-ve) | 153 (48.4) | 36 (34.6) | 117 (55.2) | 0.32 |
| | (+ve) | 6 (1.9) | 0 (0.0) | 6 (2.8) | |
| **SRSF2** | (-ve) | 240 (75.9) | 78 (75.0) | 162 (76.4) | 0.01 |
| | (+ve) | 50 (15.8) | 16 (15.4) | 34 (16.0) | |
| **STAG2** | (-ve) | 272 (86.1) | 90 (86.5) | 182 (85.8) | 0.05 |
| | (+ve) | 21 (6.6) | 6 (5.8) | 15 (7.1) | |
| **TET2** | (-ve) | 234 (74.1) | 84 (80.8) | 150 (70.8) | 0.32 |
| | (+ve) | 61 (19.3) | 12 (11.5) | 49 (23.1) | |
| **TP53** | (-ve) | 230 (72.8) | 87 (83.7) | 143 (67.5) | 0.49 |
| | (+ve) | 64 (20.3) | 9 (8.7) | 55 (25.9) | |
| **U2AF1** | (-ve) | 263 (83.2) | 85 (81.7) | 178 (84.0) | 0.03 |
| | (+ve) | 26 (8.2) | 9 (8.7) | 17 (8.0) | |
| **WT1** | (-ve) | 287 (90.8) | 93 (89.4) | 194 (91.5) | 0.07 |
| | (+ve) | 7 (2.2) | 3 (2.9) | 4 (1.9) | |
| **ZRSR2** | (-ve) | 283 (89.6) | 90 (86.5) | 193 (91.0) | 0.20 |
| | (+ve) | 6 (1.9) | 4 (3.8) | 2 (0.9) | |

*Standardized mean differences (SMD) were calculated after excluding missing cases. SMD > 0.10 refers to systematic differences between alive and deceased patients in the sample.

**Supplemental Table 1C. Summary statistics for cytogenetic features.**

| | | Total | Alive | Deceased | *SMD |
|---|---|---|---|---|---|
| **N =** | | 316 | 104 | 212 | |
| **Cytogenetics, n (%)** | | | | | |
| **Complex** | (-ve) | 209 (66.1) | 78 (75.0) | 131(61.8) | 0.24 |
| | (+ve) | 93 (29.4) | 24 (23.1) | 69 (32.5) | |
| **Del5q** | (-ve) | 269 (85.1) | 93 (89.4) | 176 (83.0) | 0.10 |
| | (+ve) | 33 (10.4) | 9 (8.7) | 24 (11.3) | |
| **Del7q** | (-ve) | 290 (91.8) | 99 (89.4) | 191 (90.1) | 0.08 |
| | (+ve) | 12 (3.8) | 3 (2.9) | 9 (4.2) | |
| **Inv(3)** | (-ve) | 297 (94.0) | 101 (97.1) | 196 (92.5) | 0.08 |
| | (+ve) | 5 (1.6) | 1 (1.0) | 4 (1.9) | |
| **Minus17** | (-ve) | 278 (88.0) | 99 (95.2) | 179 (84.4) | 0.31 |
| | (+ve) | 24 (7.6) | 3 (2.9) | 21 (9.9) | |
| **Minus5** | (-ve) | 293 (92.7) | 101 (97.1) | 192 (90.6) | 0.20 |
| | (+ve) | 9 (2.8) | 1 (1.0) | 8 (3.8) | |
| **Minus7** | (-ve) | 273 (86.4) | 94 (90.4) | 179 (84.4) | 0.09 |
| | (+ve) | 29 (9.2) | 8 (7.7) | 21 (9.9) | |
| **Monosomal** | (-ve) | 245 (77.5) | 90 (86.5) | 155 (73.1) | 0.29 |
| | (+ve) | 57 (18.0) | 12 (11.5) | 45 (21.2) | |
| **Normal** | (-ve) | 192 (60.8) | 60 (57.7) | 132 (62.3) | 0.15 |
| | (+ve) | 110 (34.8) | 42 (40.4) | 68 (32.1) | |
| **T(9;11)** | (-ve) | 295 (93.4) | 99 (95.2) | 196 (92.5) | 0.06 |
| | (+ve) | 7 (2.2) | 3 (2.9) | 4 (1.9) | |
| **Risk categories** | | | | | |
| **Good** | (-ve) | 311 (98.4) | 102 (98.1) | 209 (98.6) | 0.04 |
| | (+ve) | 5 (1.6) | 2 (1.9) | 3 (1.4) | |
| **Intermediate** | (-ve) | 186 (58.9) | 56 (53.8) | 130 (61.3) | 0.15 |
| | (+ve) | 130 (41.1) | 48 (46.2) | 82 (38.7) | |
| **Indeterminant** | (-ve) | 237 (75.0) | 76 (83.1) | 161 (75.9) | 0.07 |
| | (+ve) | 79 (25.0) | 28 (26.9) | 51 (24.1) | |
| **Poor** | (-ve) | 215 (68.0) | 76 (83.1) | 139 (65.6) | 0.16 |
| | (+ve) | 101 (32.0) | 28 (26.9) | 73 (34.4) | |

of curv[illegible]

*Standardized mean differences (SMD) were calculated after excluding missing cases. SMD > 0.10 refers to systematic differences between alive and deceased patients in the sample.

**Supplemental Table 1D. Summary statistics for fluorescence in situ hybridization (FISH) features.**

| | | Total | Alive | Deceased | *SMD |
|---|---|---|---|---|---|
| **N =** | | 316 | 104 | 212 | |
| **FISH, n (%)** | | | | | |
| **Chr 5 abn, gain** | (-ve) | 249 (78.8) | 86 (82.7) | 163 (76.9) | 0.04 |
| | (+ve) | 10 (3.2) | 3 (2.9) | 7 (3.3) | |
| **Chr 5 abn, loss** | (-ve) | 200 (63.3) | 77 (74.0) | 123 (58.0) | 0.36 |
| | (+ve) | 59 (18.7) | 12 (11.5) | 47 (22.2) | |
| **Chr 5 abn, normal** | (-ve) | 65 (20.6) | 13 (12.5) | 52 (24.5) | 0.39 |
| | (+ve) | 194 (61.4) | 76 (73.1) | 118 (55.7) | |
| **Chr 7 abn, gain** | (-ve) | 265 (83.9) | 91 (87.5) | 174 (82.1) | 0.04 |
| | (+ve) | 10 (3.2) | 3 (2.9) | 7 (3.3) | |
| **Chr 7 abn, loss** | (-ve) | 216 (68.4) | 82 (78.8) | 134 (63.2) | 0.34 |
| | (+ve) | 59 (18.7) | 12 (11.5) | 47 (22.2) | |
| **Chr 7 abn, normal** | (-ve) | 69 (21.8) | 14 (13.5) | 55 (25.9) | 0.38 |
| | (+ve) | 206 (65.2) | 80 (76.9) | 126 (59.4) | |
| **Chr 8 abn, gain** | (-ve) | 131 (41.5) | 38 (36.5) | 93 (43.9) | 0.39 |
| | (+ve) | 45 (14.2) | 6 (5.8) | 39 (18.4) | |
| **Chr 8 abn, loss** | (-ve) | 162 (51.3) | 39 (37.5) | 123 (58.0) | 0.16 |
| | (+ve) | 14 (4.4) | 5 (4.8) | 9 (4.2) | |
| **Chr 8 abn, normal** | (-ve) | 57 (18.0) | 11 (10.6) | 46 (21.7) | 0.22 |
| | (+ve) | 119 (37.7) | 33 (31.7) | 86 (40.6) | |
| **CBFB, rearrangement** | (-ve) | 243 (76.7) | 84 (80.8) | 159 (75.0) | 0.22 |
| | (+ve) | 4 (1.3) | 0 (0.0) | 4 (1.9) | |
| **CBFB, gain** | (-ve) | 238 (75.3) | 82 (78.8) | 156 (73.6) | 0.11 |
| | (+ve) | 9 (2.8) | 2 (1.9) | 7 (3.3) | |
| **CBFB, loss** | (-ve) | 237 (75.0) | 81 (77.9) | 156 (73.6) | 0.04 |
| | (+ve) | 10 (3.2) | 3 (2.9) | 7 (3.3) | |
| **CBFB, normal** | (-ve) | 22 (7.0) | 4 (3.8) | 18 (8.5) | 0.23 |
| | (+ve) | 225 (71.2) | 80 (76.9) | 145 (68.4) | |
| **DEK, rearrangement** | (-ve) | 105 (33.2) | 49 (47.1) | 56 (26.4) | 0.19 |
| | (+ve) | 1 (0.3) | 0 (0.0) | 1 (0.5) | |
| **DEK, gain** | (-ve) | 98 (31.0) | 45 (43.3) | 53 (25.0) | 0.04 |
| | (+ve) | 8 (2.5) | 4 (3.8) | 4 (1.9) | |
| **DEK, loss** | (-ve) | 100 (31.6) | 46 (44.2) | 54 (25.5) | 0.04 |
| | (+ve) | 6 (1.9) | 3 (2.9) | 3 (1.4) | |
| **DEK, normal** | (-ve) | 15 (4.7) | 7 (6.7) | 8 (3.8) | 0.01 |
| | (+ve) | 91 (28.8) | 42 (40.4) | 49 (23.1) | |
| **MECOM, rearrangement** | (-ve) | 94 (29.7) | 46 (44.2) | 48 (22.6) | 0.37 |
| | (+ve) | 7 (2.2) | 1 (1.0) | 6 (2.8) | |
| **MECOM, gain** | (-ve) | 90 (28.5) | 43 (41.3) | 47 (22.2) | 0.14 |
| | (+ve) | 11 (3.5) | 4 (3.8) | 7 (3.3) | |
| **MECOM, loss** | (-ve) | 98 (62.5) | 46 (44.2) | 52 (24.5) | 0.09 |
| | (+ve) | 3 (0.9) | 1 (1.0) | 2 (0.9) | |
| **MECOM, normal** | (-ve) | 21 (6.6) | 5 (4.8) | 16 (7.5) | 0.49 |
| | (+ve) | 80 (25.3) | 42 (40.4) | 38 (17.9) | |
| **KMT2A, rearrangement** | (-ve) | 246 (77.8) | 82 (78.8) | 164 (77.4) | 0.02 |
| | (+ve) | 17 (5.8) | 6 (5.8) | 11 (5.2) | |
| **KMT2A, gain** | (-ve) | 226 (71.5) | 79 (76.0) | 147 (69.3) | 0.17 |
| | (+ve) | 37 (11.7) | 9 (8.7) | 28 (13.2) | |

**Supplemental Table 1D, cont'd**

| | | Total | Alive | Deceased | *SMD |
|---|---|---|---|---|---|
| **N =** | | 316 | 104 | 212 | |
| **FISH, n (%)** | | | | | |
| **KMT2A, loss** | (-ve) | 260 (82.3) | 88 (84.6) | 172 (81.1) | 0.19 |
| | (+ve) | 3 (0.9) | 0 (0.0) | 3 (1.4) | |
| **KMT2A, normal** | (-ve) | 57 (18.0) | 15 (14.4) | 42 (19.8) | 0.17 |
| | (+ve) | 206 (65.2) | 73 (70.2) | 133 (62.7) | |
| **PML, rearrangement** | (-ve) | 229 (72.5) | 81 (77.9) | 148 (69.8) | <0.001 |
| | (+ve) | 0 (0.0) | 0 (0.0) | 0 (0.0) | |
| **PML, gain** | (-ve) | 222 (70.3) | 78 (75.0) | 144 (67.9) | 0.06 |
| | (+ve) | 7 (2.2) | 3 (2.9) | 4 (1.9) | |
| **PML, loss** | (-ve) | 221 (69.5) | 80 (76.9) | 141 (66.5) | 0.21 |
| | (+ve) | 8 (2.5) | 1 (1.0) | 7 (3.3) | |
| **PML, normal** | (-ve) | 16 (5.1) | 4 (3.8) | 12 (5.7) | 0.13 |
| | (+ve) | 213 (67.4) | 77 (74.0) | 136 (64.2) | |
| **RARA, rearrangement** | (-ve) | 71 (22.5) | 23 (22.1) | 48 (46.2) | <0.001 |
| | (+ve) | 0 (0.0) | 0 (0.0) | 0 (0.0) | |
| **RARA, gain** | (-ve) | 70 (22.2) | 23 (22.1) | 47 (22.2) | 0.21 |
| | (+ve) | 1 (0.3) | 0 (0.0) | 1 (0.5) | |
| **RARA, loss** | (-ve) | 64 (20.3) | 22 (21.2) | 42 (19.8) | 0.29 |
| | (+ve) | 7 (2.2) | 1 (1.0) | 6 (2.8) | |
| **RARA, normal** | (-ve) | 8 (2.5) | 1 (1.0) | 7 (3.3) | 0.36 |
| | (+ve) | 63 (19.9) | 22 (21.2) | 41 (19.3) | |
| **RUNX, rearrangement** | (-ve) | 241 (76.3) | 81 (77.9) | 160 (75.5) | 0.09 |
| | (+ve) | 4 (1.3) | 2 (1.9) | 2 (0.9) | |
| **RUNX, gain** | (-ve) | 191 (60.4) | 70 (67.3) | 121 (57.1) | 0.24 |
| | (+ve) | 54 (17.1) | 13 (12.5) | 41 (19.3) | |
| **RUNX, loss** | (-ve) | 241 (76.3) | 81 (77.9) | 160 (75.5) | 0.16 |
| | (+ve) | 4 (1.3) | 2 (1.9) | 2 (0.9) | |
| **RUNX, normal** | (-ve) | 61 (19.3) | 15 (14.4) | 46 (21.7) | 0.25 |
| | (+ve) | 184 (58.2) | 68 (65.4) | 116 (54.7) | |
| **TP53, rearrangement** | (-ve) | 38 (12.0) | 20 (19.2) | 18 (8.5) | <0.001 |
| | (+ve) | 0 (0.0) | 0 (0.0) | 0 (0.0) | |
| **TP53, gain** | (-ve) | 36 (11.4) | 19 (18.3) | 17 (8.0) | 0.03 |
| | (+ve) | 2 (0.6) | 1 (1.0) | 1 (0.5) | |
| **TP53, loss** | (-ve) | 25 (7.6) | 12 (11.5) | 13 (6.1) | 0.26 |
| | (+ve) | 13 (4.4) | 8 (7.7) | 5 (2.4) | |
| **TP53, normal** | (-ve) | 16 (5.1) | 9 (8.7) | 7 (3.3) | 0.12 |
| | (+ve) | 22 (7.0) | 11 (10.6) | 11 (5.2) | |

*Standardized mean differences (SMD) were calculated after excluding missing cases. SMD > 0.10 refers to systematic differences between alive and deceased patients in the sample.

**Supplemental Table 1E. Summary statistics for AML composite mutation features.**

| | | Total | Alive | Deceased | *SMD |
|---|---|---|---|---|---|
| **N =** | | 316 | 104 | 212 | |
| **mutation, test types** | | n (%) | n (%) | n (%) | |
| **CBFB, FISH/PCR** | (-ve) | 226 (71.5) | 80 (76.9) | 146 (68.9) | 0.23 |
| | (+ve) | 22 (7.0) | 4 (3.8) | 18 (8.5) | |
| **CEBPA, PCR/NGS** | (-ve) | 272 (86.1) | 89 (85.6) | 183 (86.3) | 0.06 |
| | (+ve) | 21 (6.6) | 8 (7.7) | 13 (6.1) | |
| **FLT3-ITD, PCR/NGS** | (-ve) | 267 (84.5) | 85 (81.7) | 182 (85.8) | 0.05 |
| | (+ve) | 34 (10.8) | 12 (11.5) | 22 (10.4) | |
| **IDH1, PCR/NGS** | (-ve) | 275 (87.0) | 84 (80.8) | 191 (90.1) | 0.28 |
| | (+ve) | 24 (7.6) | 13 (12.5) | 11 (5.2) | |
| **IDH2, PCR/NGS** | (-ve) | 254 (80.4) | 78 (75.0) | 176 (83.0) | 0.17 |
| | (+ve) | 46 (14.6) | 19 (18.3) | 27 (12.7) | |
| **JAK2, NGS/PCR** | (-ve) | 284 (89.9) | 95 (91.3) | 189 (89.2) | 0.16 |
| | (+ve) | 12 (3.8) | 2 (1.9) | 10 (4.7) | |
| **NPM1, PCR/NGS** | (-ve) | 236 (74.7) | 74 (71.2) | 162 (76.4) | 0.10 |
| | (+ve) | 60 (19.0) | 22 (21.2) | 38 (17.9) | |
| **PML, FISH/PCR/NGS** | (-ve) | 244 (77.2) | 83 (79.8) | 161 (75.9) | 0.10 |
| | (+ve) | 16 (5.1) | 4 (3.8) | 12 (5.7) | |
| **RUNX, FISH/PCR/NGS** | (-ve) | 193 (61.1) | 65 (62.5) | 128 (60.4) | 0.10 |
| | (+ve) | 109 (34.5) | 32 (30.8) | 77 (36.3) | |
| **TP53, FISH/NGS** | (-ve) | 222 (70.3) | 80 (76.9) | 142 (67.0) | 0.30 |
| | (+ve) | 74 (23.4) | 16 (15.4) | 58 (27.4) | |

*Standardized mean differences (SMD) were calculated after excluding missing cases. SMD > 0.10 refers to systematic differences between alive and deceased patients in the sample.

**Supplemental Table 1F. Summary statistics for flow cytometry (FC) features.**

| | | Total | Alive | Deceased | *SMD |
|---|---|---|---|---|---|
| **N =** | | 316 | 104 | 212 | |
| **Flow cytometry, n (%)** | | | | | |
| **CD5** | (-ve) | 66 (20.9) | 29 (27.9) | 37 (17.5) | 0.39 |
| | (+ve) | 7 (2.2) | 1 (1.0) | 6 (2.8) | |
| **CD7** | (-ve) | 201 (63.6) | 73 (70.2) | 128 (60.4) | 0.21 |
| | (+ve) | 84 (26.6) | 22 (21.2) | 62 (29.2) | |
| **CD10** | (-ve) | 205 (64.9) | 82 (78.8) | 123 (58.0) | 0.07 |
| | (+ve) | 12 (3.8) | 4 (3.8) | 8 (3.8) | |
| **CD13** | (-ve) | 34 (10.8) | 14 (13.5) | 20 (9.4) | 0.13 |
| | (+ve) | 265 (83.9) | 85 (81.7) | 180 (84.9) | |
| **CD14** | (-ve) | 277 (87.7) | 94 (90.4) | 183 (86.3) | 0.09 |
| | (+ve) | 16 (5.1) | 4 (3.8) | 12 (5.7) | |
| **CD15** | (-ve) | 181 (57.3) | 65 (62.5) | 116 (54.7) | 0.13 |
| | (+ve) | 32 (10.1) | 9 (8.7) | 23 (10.8) | |
| **CD16** | (-ve) | 248 (78.5) | 94 (90.4) | 154 (72.6) | 0.11 |
| | (+ve) | 5 (1.6) | 1 (1.0) | 4 (1.9) | |
| **CD19** | (-ve) | 280 (88.6) | 95 (91.3) | 185 (87.3) | 0.07 |
| | (+ve) | 8 (2.5) | 2 (1.9) | 6 (2.8) | |
| **CD22** | (-ve) | 226 (71.5) | 77 (74.0) | 149 (70.3) | 0.28 |
| | (+ve) | 6 (1.9) | 0 (0.0) | 6 (2.8) | |
| **CD33** | (-ve) | 38 (12.0) | 12 (11.5) | 26 (12.3) | 0.03 |
| | (+ve) | 261 (82.6) | 87 (83.7) | 174 (82.1) | |
| **CD34** | (-ve) | 63 (19.9) | 22 (21.2) | 41 (19.3) | 0.04 |
| | (+ve) | 243 (76.9) | 79 (76.0) | 164 (77.4) | |
| **CD36** | (-ve) | 124 (39.2) | 50 (48.1) | 74 (34.9) | 0.30 |
| | (+ve) | 37 (11.7) | 9 (8.7) | 28 (13.2) | |
| **CD38** | (-ve) | 25 (7.9) | 10 (9.6) | 15 (7.1) | 0.09 |
| | (+ve) | 266 (84.2) | 87 (83.7) | 179 (84.4) | |
| **CD45** | (-ve) | 69 (21.8) | 32 (30.8) | 37 (17.5) | 0.32 |
| | (+ve) | 193 (61.1) | 58 (55.8) | 135 (63.7) | |
| **CD56** | (-ve) | 202 (63.9) | 71 (68.3) | 131 (61.8) | 0.11 |
| | (+ve) | 77 (24.4) | 23 (22.1) | 54 (25.5) | |
| **CD64** | (-ve) | 189 (59.8) | 70 (67.3) | 119 (56.1) | 0.23 |
| | (+ve) | 94 (29.7) | 25 (24.0) | 69 (32.5) | |
| **CD71** | (-ve) | 8 (2.5) | 4 (3.8) | 4 (1.9) | 0.89 |
| | (+ve) | 7 (2.2) | 1 (1.0) | 6 (2.8) | |
| **CD117** | (-ve) | 19 (6.0) | 7 (6.7) | 12 (5.7) | 0.04 |
| | (+ve) | 286 (90.5) | 94 (90.4) | 192 (90.6) | |
| **CD123** | (-ve) | 33 (9.3) | 16 (15.4) | 17 (8.0) | 0.18 |
| | (+ve) | 133 (44.1) | 50 (48.1) | 83 (39.2) | |
| **CD11B** | (-ve) | 203 (64.2) | 82 (78.8) | 121 (57.1) | 0.52 |
| | (+ve) | 84 (26.6) | 14 (13.5) | 70 (33.0) | |
| **MPO** | (-ve) | 104 (32.9) | 34 (32.7) | 70 (33.0) | <0.01 |
| | (+ve) | 160 (50.6) | 52 (50.0) | 108 (50.9) | |
| **TDT** | (-ve) | 186 (58.9) | 61 (58.7) | 125 (59.0) | 0.16 |
| | (+ve) | 25 (7.9) | 11 (10.6) | 14 (6.6) | |
| **HLA-DR** | (-ve) | 48 (15.2) | 19 (18.3) | 29 (13.7) | 0.13 |
| | (+ve) | 253 (80.1) | 80 (76.9) | 173 (81.6) | |

*Standardized mean differences (SMD) were calculated after excluding missing cases. SMD > 0.10 refers to systematic differences between alive and deceased patients in the sample.

**Supplemental Table 2. Definition of genetic and phenotypic features.**

| | |
|---|---|
| *Next generation sequencing* | *Dichotomized as either positive for an abnormality (regardless of degree) or negative* |
| *Flow cytometry* | Dichotomized as either positive for an abnormality (regardless of degree) or negative |
| *FISH* | Characterized as five non-mutually exclusive groups as 'normal', 'loss', 'gain', 'rearrangement' or any 'other' abnormality. Each of the FISH categories for each FISH marker is represented as a binary (positive/negative) variable in the dataset where positive indicates the presence of that category. |
| *Cytogenetics* | Taken from the full karyotype notation and marked as positive if the abnormality was present and negative if not present |
| *Composite mutation* | Marked as positive if the mutation is present by any testing method. |

Remark: For all markers, if the test was not run, it would be marked as 'not performed' or ‘missing’.

**Supplemental Table 3. Penalized regressions based adjusted hazard ratios (aHRs) of prognostic factors for overall survival (OS) and adjusted odds ratios (aORs) for CR/CR$_h$ best response.**

### A. Next generation sequencing (NGS).

| Label | Counts | $t_{0.5}$: All | $t_{0.5}$: allo-HCT | Reference | Counts | $t_{0.5}$: All | $t_{0.5}$: allo-HCT | aHR: allo-HCT (95%) | aHR: ex-allo-HCT (95%) | aHR: All (95%) | aOR: CR/CRh (95%) |
|---|---|---|---|---|---|---|---|---|---|---|---|
| Age :: >75 | 80/96 | 322 | 299 | ≤75 | 132/220 | 431 | 286 | 1.05 (1.00,1.39) | 0.95 (0.80,1.00) | 1.35 (1.16,1.85) | 1.02 (0.90,1.67) |
| NGS-KRAS :: (+ve) | 10/12 | 82 | 82 | (-ve) | 188/282 | 386 | 311 | 1.44 (1.08,4.06) | 1.31 (1.00,2.78) | 1.76 (1.16,4.23) | 1.15 (0.87,3.24) |
| NGS-MPL :: (+ve) | 4/5 | 87 | 87 | (-ve) | 190/283 | 391 | 311 | 1.37 (1.06,2.64) | 1.13 (1.00,1.68) | 1.72 (1.17,3.74) | NA |
| NGS-PTPN11 :: (+ve) | 16/19 | 151 | 149 | (-ve) | 182/275 | 406 | 317 | 1.30 (1.11,2.16) | 1.16 (1.01,1.63) | 1.52 (1.23,2.37) | 1.10 (0.93,2.87) |
| NGS-SMC1A :: (+ve) | 6/6 | 176 | 176 | (-ve) | 117/153 | 460 | 359 | 1.47 (1.21,2.52) | 1.19 (1.02,1.96) | 1.78 (1.43,3.25) | 0.88 (0.23,1.66) |
| NGS-KMT2A :: (+ve) | 4/6 | 219.5 | 171 | (-ve) | 176/267 | 384 | 289 | 1.31 (1.08,3.58) | NA | 1.47 (1.03,4.02) | NA |
| NGS-NF1 :: (+ve) | 7/11 | 247 | 247 | (-ve) | 78/136 | 377 | 242 | 0.98 (0.71,1.06) | 1.02 (0.95,1.20) | 1.18 (1.00,1.98) | 1.06 (0.71,4.45) |
| NGS-CSF3R :: (+ve) | 3/5 | 259 | 259 | (-ve) | 194/286 | 378 | 299 | 1.20 (1.02,2.20) | NA | 1.08 (0.81,2.21) | NA |
| NGS-RAD21 :: (+ve) | 5/8 | 259 | 257.5 | (-ve) | 190/281 | 384 | 299 | 0.91 (0.51,1.01) | 0.95 (0.62,1.03) | 0.90 (0.52,1.04) | 0.92 (0.19,1.14) |
| NGS-STAG2 :: (+ve) | 15/21 | 267 | 242 | (-ve) | 182/272 | 384 | 311 | 1.03 (0.93,1.36) | 1.03 (0.97,1.36) | 1.08 (0.95,1.53) | 1.24 (1.00,7.42) |
| NGS-U2AF1 :: (+ve) | 17/26 | 268 | 242 | (-ve) | 178/263 | 386 | 311 | 1.07 (1.00,1.46) | 1.03 (0.98,1.33) | 1.12 (1.00,1.53) | 1.20 (1.00,5.29) |
| NGS-NRAS :: (+ve) | 21/30 | 339.5 | 311 | (-ve) | 179/265 | 378 | 292 | 1.00 (0.81,1.11) | 0.99 (0.78,1.06) | 0.94 (0.48,1.03) | 1.05 (0.79,1.89) |
| NGS-TET2 :: (+ve) | 49/61 | 342 | 264 | (-ve) | 150/234 | 386 | 299 | 0.98 (0.79,1.04) | 0.96 (0.80,1.00) | 1.12 (1.00,1.42) | 1.02 (0.85,1.84) |
| NGS-GATA2 :: (+ve) | 9/13 | 386 | 342 | (-ve) | 186/276 | 378 | 289 | 0.99 (0.70,1.10) | 0.98 (0.76,1.06) | 0.95 (0.62,1.08) | 1.09 (0.77,2.74) |
| NGS-DNMT3A :: (+ve) | 37/63 | 397 | 319 | (-ve) | 161/230 | 375 | 286 | 0.84 (0.52,0.95) | 0.90 (0.59,1.00) | 0.81 (0.52,0.95) | 0.94 (0.47,1.03) |
| NGS-ASXL1 :: (+ve) | 45/63 | 419 | 289 | (-ve) | 153/231 | 378 | 314 | 0.99 (0.84,1.05) | 0.98 (0.81,1.03) | 0.99 (0.79,1.08) | 1.09 (0.89,1.76) |
| NGS-WT1 :: (+ve) | 4/7 | 431 | 431 | (-ve) | 194/287 | 377 | 292 | 0.96 (0.67,1.05) | 0.96 (0.67,1.03) | 0.90 (0.50,1.04) | 1.05 (0.65,4.78) |
| NGS-SRSF2 :: (+ve) | 34/50 | 432 | 342 | (-ve) | 162/240 | 375 | 289 | 0.95 (0.56,1.00) | 0.97 (0.74,1.01) | 0.89 (0.58,1.00) | 1.03 (0.84,1.74) |
| NGS-BCORL1 :: (+ve) | 8/11 | 521 | 519 | (-ve) | 187/277 | 377 | 292 | 0.89 (0.57,1.00) | 0.96 (0.66,1.04) | 0.92 (0.59,1.02) | 1.04 (0.82,3.39) |
| NGS-SH2B3 :: (+ve) | 3/6 | 537 | 537 | (-ve) | 81/140 | 375 | 242 | 0.88 (0.55,1.00) | NA | 0.81 (0.42,1.00) | 1.00 (0.23,1.79) |
| NGS-EZH2 :: (+ve) | 12/16 | 551 | 470 | (-ve) | 187/279 | 375 | 286 | 0.90 (0.57,1.00) | 0.93 (0.73,1.00) | 0.92 (0.61,1.02) | 1.11 (0.90,3.13) |
| NGS-PHF6 :: (+ve) | 8/12 | 578.5 | 359 | (-ve) | 188/278 | 377 | 289 | 0.98 (0.74,1.11) | 1.02 (0.94,1.40) | 0.95 (0.67,1.14) | 0.96 (0.36,1.24) |
| NGS-SETBP1 :: (+ve) | 5/9 | 579 | 311 | (-ve) | 193/285 | 378 | 292 | 1.05 (0.94,1.82) | 1.06 (0.96,2.04) | 0.95 (0.55,1.17) | 1.14 (0.79,4.52) |
| NGS-KIT :: (+ve) | 3/5 | 659 | 193 | (-ve) | 195/289 | 378 | 299 | 0.81 (0.25,1.00) | NA | 0.88 (0.38,1.10) | 1.02 (0.69,6.87) |
| NGS-BCOR :: (+ve) | 10/20 | 865 | 769 | (-ve) | 188/274 | 359 | 286 | 0.77 (0.42,0.92) | 0.81 (0.52,0.99) | 0.72 (0.38,0.87) | 0.95 (0.29,1.15) |
| NGS-SF3B1 :: (+ve) | 11/19 | 1127 | 519 | (-ve) | 183/268 | 355 | 286 | 0.86 (0.55,0.98) | 0.90 (0.68,1.00) | 0.74 (0.39,0.94) | 0.87 (0.34,1.02) |
| NGS-DDX41 :: (+ve) | 0/6 | NA | NA | (-ve) | 84/140 | 347 | 215 | 0.66 (0.23,0.85) | NA | 0.47 (0.18,0.61) | 0.79 (0.09,1.00) |
| NGS-ZRSR2 :: (+ve) | 2/6 | NA | NA | (-ve) | 193/283 | 378 | 292 | 1.00 (0.77,1.31) | NA | 0.72 (0.38,1.00) | 1.06 (0.65,4.71) |

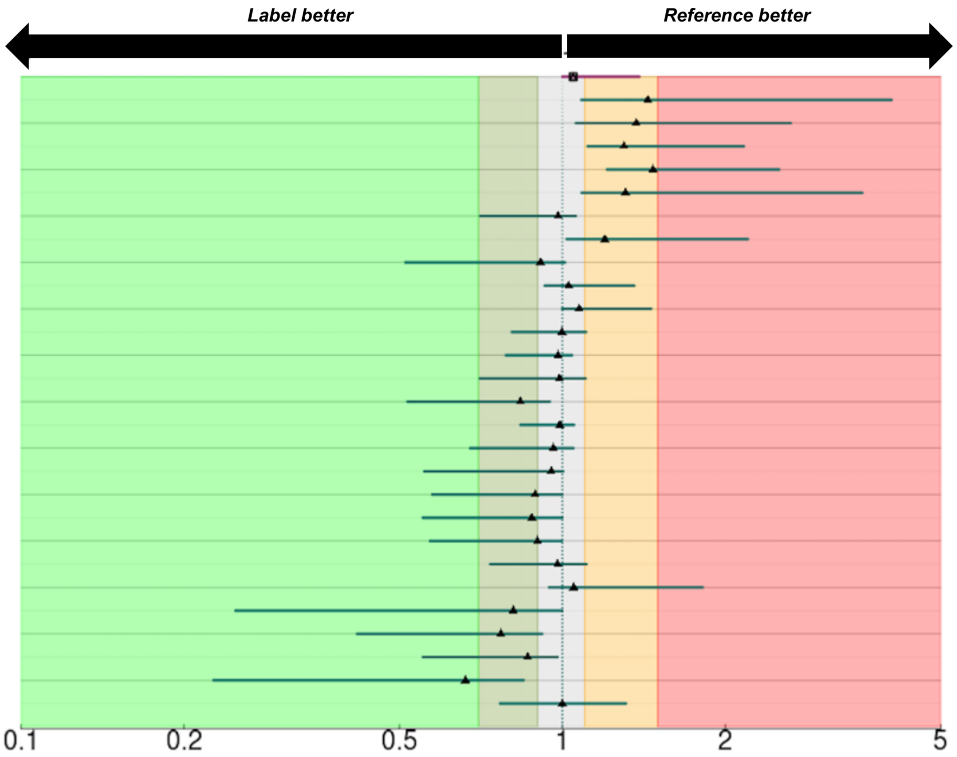

### Supplemental Table 3, cont'd

#### B. Cytogenetics (CYT).

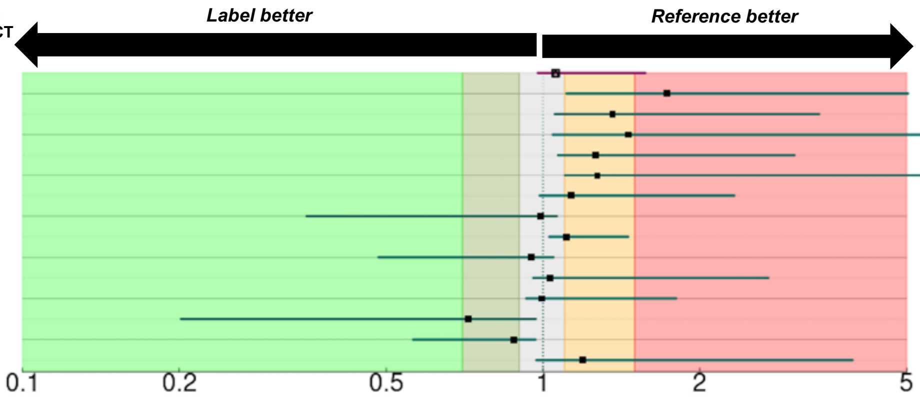


| Label | Counts | $t_{0.5}$: All | $t_{0.5}$: allo-HCT | Reference | Counts | $t_{0.5}$: All | $t_{0.5}$: allo-HCT | aHR: allo-HCT (95%) | aHR: ex-allo-HCT (95%) | aHR: All (95%) | aOR: CR/CRh (95%) |
|---|---|---|---|---|---|---|---|---|---|---|---|
| Age :: >75 | 80/96 | 322 | 299 | ≤75 | 132/220 | 431 | 286 | 1.06 (0.98,1.57) | 0.91 (0.72,1.02) | 1.28 (1.09,2.12) | 1.01 (0.88,1.53) |
| CYT-Del7q :: (+ve) | 9/12 | 66 | 66 | (-ve) | 191/290 | 391 | 311 | 1.73 (1.11,5.03) | 1.92 (1.17,4.55) | 1.33 (1.00,3.20) | 1.08 (0.99,3.68) |
| CYT-Minus5 :: (+ve) | 8/9 | 114 | 114 | (-ve) | 192/293 | 391 | 311 | 1.36 (1.05,3.39) | 1.84 (1.22,5.54) | 1.38 (1.09,2.70) | 1.04 (0.67,3.29) |
| CYT-Inv3 :: (+ve) | 4/5 | 248 | 248 | (-ve) | 196/297 | 390 | 311 | 1.46 (1.04,6.54) | NA | 1.22 (0.87,2.87) | 1.00 (0.70,3.42) |
| CYT-Minus17 :: (+ve) | 21/24 | 256 | 163 | (-ve) | 179/278 | 431 | 314 | 1.26 (1.07,3.04) | 1.21 (1.05,2.09) | 1.29 (1.09,2.46) | 1.00 (0.78,2.55) |
| CYT-Complex :: (+ve) | 69/93 | 279 | 206 | (-ve) | 131/209 | 483 | 355 | 1.27 (1.10,6.24) | 1.41 (1.19,5.77) | 1.17 (1.06,4.59) | 1.01 (0.95,1.59) |
| CYT-Minus7 :: (+ve) | 21/29 | 279 | 248 | (-ve) | 179/273 | 406 | 322 | 1.13 (0.99,2.33) | 1.39 (1.10,3.50) | 1.04 (0.85,1.37) | 1.01 (0.78,2.13) |
| CYT-Monosomal :: (+ve) | 45/57 | 317 | 279 | (-ve) | 155/245 | 432 | 314 | 0.99 (0.35,1.06) | 0.89 (0.26,1.04) | 1.08 (0.95,1.27) | 1.00 (0.85,1.70) |
| CYT risk -Poor :: (+ve) | 73/101 | 317 | 242 | (-ve) | 139/215 | 438 | 331 | 1.11 (1.03,1.46) | 1.13 (1.04,1.68) | 1.05 (0.97,1.15) | 1.01 (0.91,1.39) |
| CYT-Del5q :: (+ve) | 24/33 | 347 | 286 | (-ve) | 176/269 | 390 | 311 | 0.95 (0.48,1.05) | 0.89 (0.51,1.03) | 0.94 (0.56,1.05) | 0.96 (0.53,1.06) |
| CYT risk -Indeterminant :: (+ve) | 51/79 | 419 | 292 | (-ve) | 161/237 | 365 | 286 | 1.03 (0.96,2.71) | 1.07 (0.99,3.11) | 1.00 (0.91,1.21) | 1.02 (0.94,1.49) |
| CYT risk -Intermediate :: (+ve) | 82/130 | 470 | 355 | (-ve) | 130/186 | 322 | 268 | 0.99 (0.93,1.80) | 1.06 (0.98,4.11) | 0.98 (0.90,1.12) | 0.98 (0.83,1.25) |
| CYT risk -Good :: (+ve) | 3/5 | 475 | 475 | (-ve) | 209/311 | 375 | 289 | 0.72 (0.20,0.97) | 0.64 (0.10,0.91) | 0.87 (0.38,1.09) | NA |
| CYT-Normal :: (+ve) | 68/110 | 519 | 397 | (-ve) | 132/192 | 337 | 259 | 0.88 (0.56,0.97) | 0.84 (0.33,0.95) | 0.94 (0.81,1.00) | 0.96 (0.74,1.00) |
| CYT-T911 :: (+ve) | 4/7 | 659 | 242 | (-ve) | 196/295 | 384 | 311 | 1.19 (0.97,3.93) | NA | 0.83 (0.27,1.12) | 1.01 (0.58,3.37) |

#### C. Fluorescence in situ hybridization (FISH).

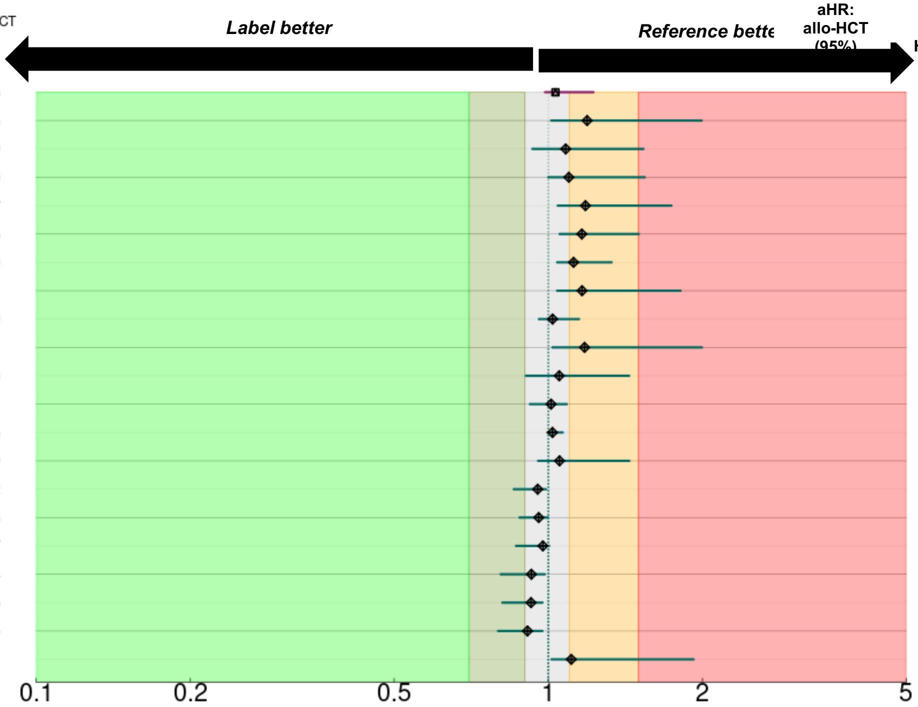


| Label | Counts | $t_{0.5}$: All | $t_{0.5}$: allo-HCT | Reference | Counts | $t_{0.5}$: All t:All | $t_{0.5}$: allo-HCT t:HSCT | aHR: allo-HCT (95%) | aHR: ex-allo-HCT (95%) | aHR: All (95%) | aOR: CR/CRh (95%) R:( |
|---|---|---|---|---|---|---|---|---|---|---|---|
| Age :: >75 | 80/96 | 322 | 299 | ≤75 | 132/220 | 431 | 286 | 1.03 (0.99,1.22) | 0.94 (0.81,1.01) | 1.19 (1.08,1.67) | 1.00 ( |
| FISH-DEK [Loss] :: (+ve) | 3/6 | 44 | 44 | (-ve) | 54/100 | 377 | 256 | 1.19 (1.01,1.99) | NA | 1.16 (0.99,2.22) | 1.00 ( |
| FISH-CHR 7 ABN [Gain] :: (+ve) | 7/10 | 124 | 124 | (-ve) | 174/265 | 386 | 299 | 1.08 (0.93,1.53) | 1.07 (0.87,1.43) | 1.18 (1.03,1.92) | 1.00 ( |
| FISH-MECOM [Rearrangement] :: (+ve) | 6/7 | 167.5 | 147 | (-ve) | 48/94 | 377 | 259 | 1.10 (1.00,1.54) | 1.11 (1.00,1.47) | 1.32 (1.13,2.13) | 1.00 ( |
| FISH-CHR 8 ABN [Loss] :: (+ve) | 9/14 | 196 | 196 | (-ve) | 123/162 | 397 | 317 | 1.18 (1.04,1.74) | 1.27 (1.11,2.34) | 1.03 (0.86,1.35) | 1.00 ( |
| FISH-CHR 7 ABN [Loss] :: (+ve) | 47/59 | 206 | 165 | (-ve) | 134/216 | 475 | 355 | 1.16 (1.05,1.50) | 1.23 (1.11,1.53) | 1.15 (1.04,1.41) | 1.00 ( |
| FISH-CHR 5 ABN [Loss] :: (+ve) | 47/59 | 247 | 242 | (-ve) | 123/200 | 438 | 319 | 1.12 (1.04,1.33) | 1.08 (1.01,1.23) | 1.15 (1.06,1.41) | 1.00 ( |
| FISH-MECOM [Gain] :: (+ve) | 7/11 | 247 | 247 | (-ve) | 47/90 | 377 | 311 | 1.16 (1.04,1.81) | 1.45 (1.25,2.37) | 1.07 (0.93,1.41) | 1.00 ( |
| FISH-CHR 8 ABN [Gain] :: (+ve) | 39/45 | 279 | 264 | (-ve) | 93/131 | 406 | 319 | 1.02 (0.96,1.15) | 0.99 (0.87,1.07) | 1.11 (1.03,1.35) | 1.00 ( |
| FISH-RARA [Loss] :: (+ve) | 6/7 | 289 | 114 | (-ve) | 42/64 | 447 | 391 | 1.18 (1.02,2.00) | 1.30 (1.10,2.37) | 1.20 (1.05,1.85) | 1.00 ( |
| FISH-CHR 5 ABN [Gain] :: (+ve) | 7/10 | 342.5 | 342.5 | (-ve) | 163/249 | 377 | 289 | 1.05 (0.90,1.44) | 1.07 (0.83,1.48) | 0.95 (0.65,1.13) | 1.00 ( |
| FISH-MLL [Gain] :: (+ve) | 28/37 | 349 | 242 | (-ve) | 147/226 | 419 | 311 | 1.01 (0.92,1.09) | 1.00 (0.80,1.07) | 1.02 (0.88,1.11) | 1.00 ( |
| FISH-DEK [Normal] :: (+ve) | 49/91 | 377 | 259 | (-ve) | 8/15 | 320 | 215 | 1.02 (1.00,1.07) | 1.01 (0.93,1.05) | 1.00 (0.97,1.08) | 1.00 ( |
| FISH-DEK [Gain] :: (+ve) | 4/8 | 391 | 215 | (-ve) | 53/98 | 375 | 259 | 1.05 (0.96,1.44) | NA | 0.92 (0.60,1.09) | 1.00 ( |
| FISH-MLL [Normal] :: (+ve) | 133/206 | 419 | 314 | (-ve) | 42/57 | 349 | 242 | 0.95 (0.86,0.99) | 0.94 (0.82,0.98) | 0.97 (0.91,1.01) | 1.00 ( |
| FISH-CHR 8 ABN [Normal] :: (+ve) | 86/119 | 432 | 322 | (-ve) | 46/57 | 279 | 256 | 0.96 (0.88,1.00) | 0.94 (0.84,0.99) | 0.99 (0.93,1.04) | 1.00 ( |
| FISH-MECOM [Normal] :: (+ve) | 38/80 | 447 | 311 | (-ve) | 16/21 | 247 | 147 | 0.98 (0.86,1.00) | 0.97 (0.80,1.01) | 0.95 (0.83,0.99) | 1.00 ( |
| FISH-RARA [Normal] :: (+ve) | 41/63 | 447 | 391 | (-ve) | 7/8 | 114 | 114 | 0.93 (0.81,0.98) | 0.89 (0.75,0.95) | 0.97 (0.87,1.01) | 1.00 ( |
| FISH-CHR 5 ABN [Normal] :: (+ve) | 118/194 | 451 | 342 | (-ve) | 52/65 | 242 | 215 | 0.93 (0.81,0.97) | 0.95 (0.88,1.00) | 0.90 (0.79,0.95) | 1.00 ( |
| FISH-CHR 7 ABN [Normal] :: (+ve) | 126/206 | 483 | 355 | (-ve) | 55/69 | 204 | 165 | 0.91 (0.80,0.97) | 0.88 (0.76,0.95) | 0.88 (0.73,0.95) | 1.00 ( |
| FISH-MLL [Rearrangement] :: (+ve) | 11/17 | 532 | 242 | (-ve) | 164/246 | 384 | 311 | 1.11 (1.01,1.92) | 1.40 (1.16,3.12) | 1.01 (0.89,1.34) | 1.00 ( |

### Supplemental Table 3, cont'd

#### D. AML specific composite mutations.

| Label | Counts | $t_{0.5}$: All | $t_{0.5}$: allo-HCT | Reference | Counts | $t_{0.5}$: All | $t_{0.5}$: allo-HCT | aHR: allo-HCT (95%) | aHR: ex-allo-HCT (95%) | aHR: All (95%) | aOR: CR/CRh (95%) |
|---|---|---|---|---|---|---|---|---|---|---|---|
| Age :: >75 | 80/96 | 322 | 299 | ≤75 | 132/220 | 431 | 286 | 0.97 (0.74,1.06) | 0.84 (0.55,0.95) | 1.36 (1.19,1.93) | 1.03 (0.81,1.60) |
| FISH/NGS-TP53 :: (+ve) | 58/74 | 242 | 203 | (-ve) | 142/222 | 451 | 342 | 1.50 (1.25,2.45) | 1.47 (1.23,2.44) | 1.38 (1.17,2.53) | 1.25 (1.00,2.44) |
| PCR/NGS-JAK2 :: (+ve) | 10/12 | 258 | 151 | (-ve) | 189/284 | 384 | 299 | 1.30 (1.04,2.39) | 1.23 (1.01,2.12) | 1.27 (0.99,2.42) | 1.89 (1.00,6.61) |
| FISH/PCR-CBFB :: (+ve) | 18/22 | 264 | 264 | (-ve) | 146/226 | 386 | 311 | 1.08 (0.89,1.38) | 1.06 (0.93,1.38) | 1.25 (1.02,1.80) | 0.77 (0.23,1.11) |
| PCR/NGS-FLT3 :: (+ve) | 42/64 | 315 | 234 | (-ve) | 162/237 | 391 | 314 | 1.08 (0.98,1.55) | 1.08 (0.99,1.67) | 1.06 (0.93,1.32) | 1.13 (0.91,2.68) |
| PCR/NGS-FLT3ITD :: (+ve) | 22/34 | 315 | 253 | (-ve) | 182/267 | 378 | 311 | 1.01 (0.85,1.33) | 1.06 (0.93,1.61) | 1.06 (0.89,1.46) | 1.08 (0.78,2.50) |
| FISH/PCR/NGS-RUNX :: (+ve) | 77/109 | 359 | 256 | (-ve) | 128/193 | 377 | 317 | 1.12 (1.02,1.41) | 1.08 (1.00,1.36) | 1.09 (0.99,1.36) | 1.13 (0.95,2.01) |
| PCR/NGS-NPM1 :: (+ve) | 38/60 | 378 | 378 | (-ve) | 162/236 | 359 | 259 | 0.79 (0.50,0.90) | 0.75 (0.41,0.88) | 0.98 (0.84,1.24) | 0.71 (0.27,1.00) |
| FISH/PCR/NGS-PML :: (+ve) | 12/16 | 384 | 357 | (-ve) | 161/244 | 386 | 299 | 1.00 (0.72,1.29) | 0.97 (0.64,1.12) | 1.05 (0.78,1.38) | 0.79 (0.22,1.20) |
| PCR/NGS-CEBPA :: (+ve) | 13/21 | 451 | 314 | (-ve) | 183/272 | 378 | 299 | 1.01 (0.85,1.34) | 0.98 (0.74,1.12) | 1.02 (0.85,1.35) | 0.99 (0.46,1.68) |
| PCR/NGS-IDH2 :: (+ve) | 27/46 | 624 | 438 | (-ve) | 176/254 | 331 | 268 | 0.84 (0.57,0.97) | 0.93 (0.64,1.03) | 0.72 (0.37,0.86) | 0.73 (0.31,1.00) |
| PCR/NGS-IDH1 :: (+ve) | 11/24 | 994 | 378 | (-ve) | 191/275 | 347 | 259 | 0.73 (0.45,0.87) | 0.77 (0.44,0.94) | 0.62 (0.25,0.77) | 0.47 (0.09,1.00) |

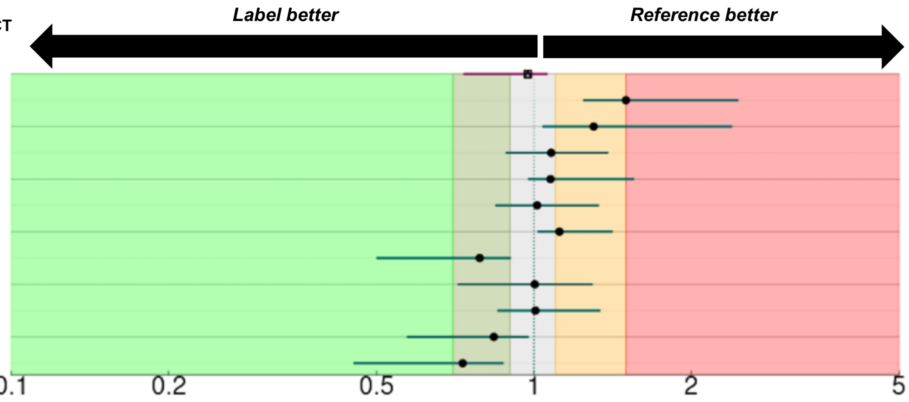


#### E. Flow cytometry (FC).

| Label | Counts | $t_{0.5}$: All | $t_{0.5}$: allo-HCT | Reference | Counts | $t_{0.5}$: All | $t_{0.5}$: allo-HCT | aHR: allo-HCT (95%) | aHR: ex-allo-HCT (95%) | aHR: All (95%) | aOR: CR/CRh (95%) |
|---|---|---|---|---|---|---|---|---|---|---|---|
| Age :: >75 | 80/96 | 322 | 299 | ≤75 | 132/220 | 431 | 286 | 1.06 (1.00,1.41) | 0.97 (0.82,1.00) | 1.30 (1.16,1.82) | 1.05 (1.00,1.97) |
| FC-CD16 :: (+ve) | 4/5 | 75 | 75 | (-ve) | 154/248 | 378 | 279 | 1.85 (1.35,8.77) | NA | 1.58 (1.06,3.68) | 1.64 (1.00,63.64) |
| FC-CD36 :: (+ve) | 28/37 | 171 | 136 | (-ve) | 74/124 | 397 | 289 | 1.32 (1.09,2.10) | 1.14 (1.00,1.59) | 1.35 (1.15,2.10) | 1.10 (1.00,2.35) |
| FC-CD71 :: (+ve) | 6/7 | 171 | 171 | (-ve) | 4/8 | 896 | 470 | 1.61 (1.26,5.91) | 1.20 (1.00,2.17) | 1.58 (1.22,3.22) | 1.22 (1.00,25.38) |
| FC-CD7 :: (+ve) | 62/84 | 207 | 199 | (-ve) | 128/201 | 483 | 397 | 1.28 (1.12,1.89) | 1.13 (1.00,1.62) | 1.31 (1.17,1.79) | 1.18 (1.00,2.81) |
| FC-CD14 :: (+ve) | 12/16 | 249 | 234 | (-ve) | 183/277 | 386 | 289 | 0.93 (0.43,1.01) | 0.95 (0.51,1.00) | 0.99 (0.65,1.12) | 0.95 (0.21,1.00) |
| FC-CD56 :: (+ve) | 54/77 | 253 | 234 | (-ve) | 131/202 | 390 | 311 | 1.14 (1.04,1.65) | 1.04 (1.00,1.31) | 1.13 (1.03,1.48) | 1.11 (1.00,2.92) |
| FC-CD11B :: (+ve) | 70/84 | 268 | 264 | (-ve) | 121/203 | 483 | 292 | 1.08 (1.00,1.50) | 1.01 (0.99,1.12) | 1.20 (1.10,1.55) | 1.05 (1.00,3.33) |
| FC-CD64 :: (+ve) | 69/94 | 315 | 264 | (-ve) | 119/189 | 431 | 317 | 1.07 (1.01,1.50) | 1.04 (1.00,1.33) | 1.09 (1.01,1.38) | 0.92 (0.33,1.00) |
| FC-CD5 :: (+ve) | 6/7 | 317 | 317 | (-ve) | 37/66 | 438 | 406 | 1.21 (1.06,2.52) | 1.04 (1.00,1.36) | 1.33 (1.13,2.43) | 1.09 (1.00,4.81) |
| FC-CD15 :: (+ve) | 23/32 | 319 | 299 | (-ve) | 116/181 | 391 | 311 | 0.92 (0.40,1.00) | 0.94 (0.69,1.00) | 0.93 (0.68,1.01) | 0.94 (0.32,1.00) |
| FC-CD10 :: (+ve) | 8/12 | 322 | 329 | (-ve) | 123/205 | 391 | 286 | 0.75 (0.22,0.98) | 0.87 (0.43,1.00) | 0.92 (0.51,1.05) | 0.89 (0.04,1.00) |
| FC-CD123 :: (+ve) | 83/133 | 349 | 256 | (-ve) | 17/33 | 419 | 207 | 0.99 (0.68,1.01) | 1.02 (1.00,1.11) | 0.92 (0.70,0.97) | 0.93 (0.11,1.00) |
| FC-CD45 :: (+ve) | 135/193 | 357 | 292 | (-ve) | 37/69 | 519 | 311 | 1.04 (1.00,1.41) | 0.99 (0.91,1.03) | 1.11 (1.04,1.48) | 0.96 (0.52,1.00) |
| FC-CD13 :: (+ve) | 180/265 | 375 | 264 | (-ve) | 20/34 | 582 | 732 | 1.05 (1.00,1.43) | 1.01 (0.98,1.23) | 1.08 (1.00,1.51) | 1.05 (1.00,2.19) |
| FC-CD33 :: (+ve) | 174/261 | 375 | 268 | (-ve) | 26/38 | 579 | 406 | 1.01 (0.96,1.22) | 1.02 (0.99,1.17) | 0.98 (0.82,1.04) | 1.04 (1.00,2.51) |
| FC-CD34 :: (+ve) | 164/243 | 375 | 259 | (-ve) | 41/63 | 438 | 342 | 1.19 (1.08,2.25) | 1.08 (1.00,1.44) | 1.14 (1.05,1.62) | 1.09 (1.00,3.44) |
| FC-CD117 :: (+ve) | 192/286 | 377 | 289 | (-ve) | 12/19 | 869 | 869 | 1.08 (1.00,1.90) | 1.02 (0.98,1.28) | 1.03 (0.94,1.31) | 1.01 (1.00,2.92) |
| FC-HLADR :: (+ve) | 173/253 | 377 | 268 | (-ve) | 29/48 | 492 | 331 | 1.01 (0.97,1.22) | 1.00 (0.90,1.06) | 1.05 (0.98,1.26) | 1.03 (1.00,2.63) |
| FC-CD38 :: (+ve) | 179/266 | 378 | 289 | (-ve) | 15/25 | 391 | 317 | 0.99 (0.80,1.03) | 1.00 (0.98,1.12) | 0.99 (0.83,1.05) | 1.04 (1.00,6.38) |
| FC-CD19 :: (+ve) | 6/8 | 431 | 460 | (-ve) | 185/280 | 375 | 279 | 0.83 (0.40,1.00) | 0.89 (0.50,1.00) | 0.96 (0.61,1.10) | 1.03 (1.00,8.22) |
| FC-MPO :: (+ve) | 108/160 | 433 | 319 | (-ve) | 70/104 | 337 | 256 | 0.92 (0.65,1.00) | 0.96 (0.75,1.00) | 0.94 (0.77,1.00) | 0.94 (0.40,1.00) |
| FC-CD11C :: (+ve) | 3/5 | 438 | 113 | (-ve) | 4/11 | NA | 896 | 1.30 (1.09,2.65) | NA | 1.22 (1.00,2.79) | NA |
| FC-CD22 :: (+ve) | 6/6 | 520.5 | 769 | (-ve) | 149/226 | 375 | 268 | 0.93 (0.60,1.02) | NA | 1.03 (0.92,1.41) | 0.88 (0.02,1.00) |
| FC-TDT :: (+ve) | 14/25 | 874 | 256 | (-ve) | 125/186 | 349 | 286 | 1.00 (0.93,1.29) | 1.01 (0.96,1.18) | 0.82 (0.52,0.95) | 0.85 (0.20,1.00) |

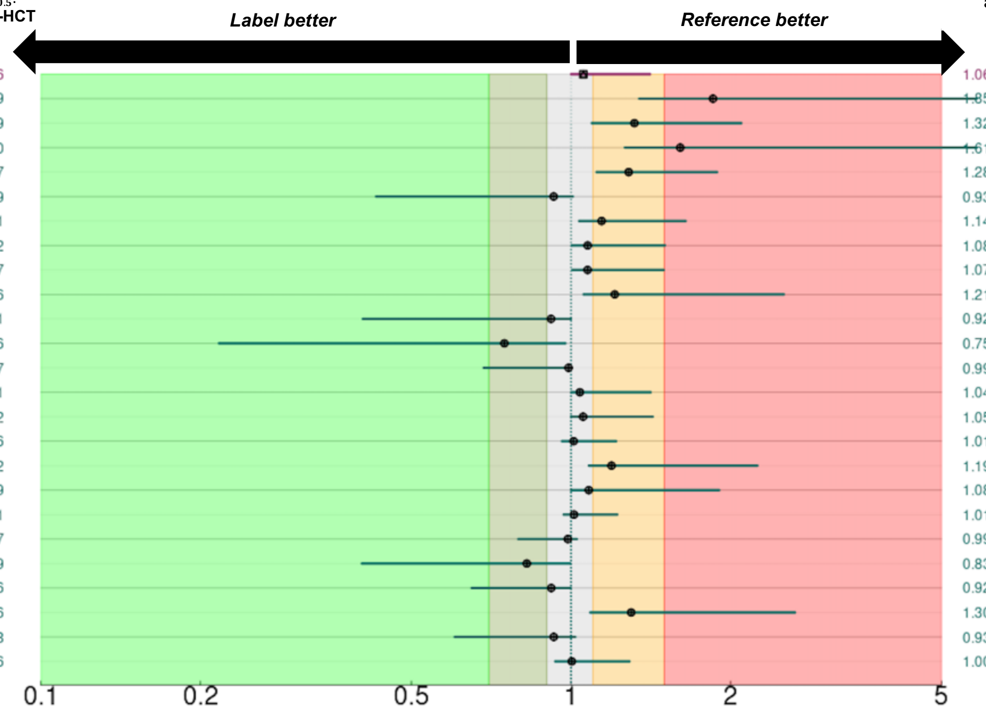

**Supplemental Table 4. Penalized regressions based adjusted hazard ratios (aHRs) of prognostic factors for event free survival (EFS).**

### A. Next generation sequencing (NGS).

| Label | Counts | $t_{0.5}$: All | $t_{0.5}$: allo-HCT | Reference | Counts | $t_{0.5}$: All | $t_{0.5}$: allo-HCT | aHR: allo-HCT (95%) | aHR: ex-allo-HCT (95%) | aHR: All (95%) |
|---|---|---|---|---|---|---|---|---|---|---|
| Age :: >75 | 75/91 | 248 | 248 | ≤75 | 147/212 | 182 | 158 | 0.99 (0.86,1.00) | 0.93 (0.73,1.00) | 1.09 (1.00,1.56) |
| NGS-KRAS :: (+ve) | 10/12 | 28 | 28 | (-ve) | 198/270 | 219 | 194 | 1.21 (1.00,2.25) | 1.33 (1.00,2.78) | 1.36 (1.00,2.79) |
| NGS-PTPN11 :: (+ve) | 17/19 | 30 | 30 | (-ve) | 191/263 | 231 | 209 | 1.21 (1.00,2.07) | 1.19 (1.00,1.69) | 1.29 (1.00,2.43) |
| NGS-WT1 :: (+ve) | 6/7 | 31 | 31 | (-ve) | 202/275 | 215 | 192 | 1.14 (1.00,2.09) | 1.02 (0.93,1.61) | 1.25 (1.00,2.72) |
| NGS-KMT2A :: (+ve) | 4/6 | 36 | 36 | (-ve) | 186/255 | 202 | 182 | 1.16 (1.00,2.43) | NA | 1.23 (1.00,2.88) |
| NGS-SH2B3 :: (+ve) | 5/6 | 90.5 | 42 | (-ve) | 83/132 | 186 | 142 | 1.04 (1.00,1.95) | NA | 1.26 (1.00,2.19) |
| NGS-GATA2 :: (+ve) | 10/12 | 109.5 | 109.5 | (-ve) | 195/265 | 215 | 192 | 1.09 (1.00,1.70) | 1.01 (0.96,1.38) | 1.12 (1.00,1.93) |
| NGS-U2AF1 :: (+ve) | 17/24 | 117 | 117 | (-ve) | 188/253 | 215 | 194 | 1.02 (1.00,1.41) | 1.01 (0.97,1.26) | 1.07 (1.00,1.47) |
| NGS-NRAS :: (+ve) | 20/26 | 117.5 | 99 | (-ve) | 189/256 | 215 | 194 | 1.04 (1.00,1.52) | 0.99 (0.72,1.04) | 1.03 (1.00,1.44) |
| NGS-NF1 :: (+ve) | 7/9 | 128 | 391 | (-ve) | 81/129 | 183 | 142 | 1.00 (0.73,1.04) | 1.05 (0.98,1.60) | 1.15 (1.00,1.89) |
| NGS-CSF3R :: (+ve) | 3/5 | 163 | 163 | (-ve) | 204/274 | 206 | 189 | 1.05 (1.00,1.54) | NA | 0.98 (0.48,1.00) |
| NGS-SMC1A :: (+ve) | 6/6 | 164 | 164 | (-ve) | 125/150 | 241 | 219 | 1.13 (1.00,1.53) | 1.08 (1.00,1.50) | 1.22 (1.05,1.79) |
| NGS-KIT :: (+ve) | 4/5 | 166 | 166 | (-ve) | 204/277 | 215 | 192 | 0.91 (0.44,1.00) | NA | 1.01 (0.96,1.69) |
| NGS-STAG2 :: (+ve) | 14/20 | 193.5 | 136 | (-ve) | 193/261 | 209 | 189 | 0.98 (0.69,1.00) | 1.00 (0.79,1.04) | 0.98 (0.69,1.00) |
| NGS-ASXL1 :: (+ve) | 43/60 | 219 | 189 | (-ve) | 165/222 | 202 | 192 | 0.97 (0.73,1.00) | 0.99 (0.76,1.01) | 0.93 (0.72,1.00) |
| NGS-EZH2 :: (+ve) | 14/15 | 241 | 241 | (-ve) | 194/267 | 206 | 182 | 1.02 (1.00,1.44) | 1.02 (0.98,1.26) | 1.07 (1.00,1.50) |
| NGS-TET2 :: (+ve) | 47/58 | 248 | 248 | (-ve) | 162/225 | 206 | 186 | 0.96 (0.71,1.00) | 0.95 (0.78,1.00) | 1.01 (1.00,1.23) |
| NGS-RAD21 :: (+ve) | 5/8 | 256 | 256 | (-ve) | 200/269 | 206 | 189 | 0.93 (0.49,1.00) | 0.94 (0.56,1.00) | 0.89 (0.47,1.00) |
| NGS-BCOR :: (+ve) | 11/20 | 278 | 397 | (-ve) | 197/262 | 196 | 182 | 0.86 (0.43,1.00) | 0.86 (0.57,1.00) | 0.83 (0.53,1.00) |
| NGS-DNMT3A :: (+ve) | 41/62 | 315 | 275 | (-ve) | 167/219 | 166 | 158 | 0.90 (0.67,1.00) | 0.91 (0.58,1.00) | 0.88 (0.65,1.00) |
| NGS-BCORL1 :: (+ve) | 9/10 | 318 | 318 | (-ve) | 196/266 | 206 | 186 | 1.00 (0.79,1.07) | 0.99 (0.76,1.04) | 1.08 (1.00,1.45) |
| NGS-PHF6 :: (+ve) | 9/12 | 352 | 256 | (-ve) | 197/266 | 202 | 186 | 1.03 (1.00,2.23) | 1.08 (1.00,1.78) | 0.99 (0.67,1.00) |
| NGS-SRSF2 :: (+ve) | 29/44 | 402 | 346 | (-ve) | 177/234 | 183 | 164 | 0.92 (0.65,1.00) | 0.92 (0.69,1.00) | 0.86 (0.61,1.00) |
| NGS-SETBP1 :: (+ve) | 4/8 | 448 | 312 | (-ve) | 204/274 | 206 | 189 | 0.96 (0.57,1.00) | 0.99 (0.57,1.10) | 0.87 (0.53,1.00) |
| NGS-SF3B1 :: (+ve) | 14/19 | 469 | 384 | (-ve) | 190/256 | 194 | 182 | 0.95 (0.70,1.00) | 0.94 (0.76,1.00) | 0.89 (0.58,1.00) |
| NGS-DDX41 :: (+ve) | 2/6 | NA | NA | (-ve) | 85/131 | 163 | 142 | 0.95 (0.57,1.00) | NA | 0.77 (0.37,1.00) |
| NGS-ZRSR2 :: (+ve) | 2/6 | NA | NA | (-ve) | 203/271 | 206 | 189 | 0.94 (0.64,1.00) | NA | 0.76 (0.41,1.00) |

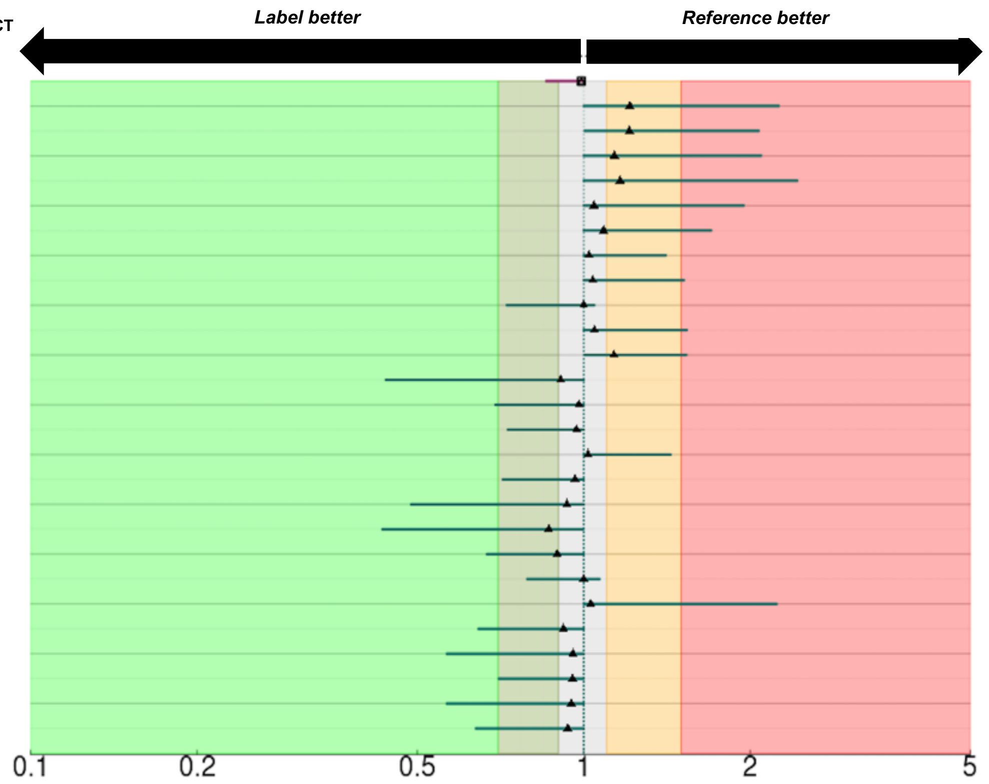

### Supplemental Table 4, cont'd

#### B. Cytogenetics (CYT).

| Label | Counts | $t_{0.5}$: All | $t_{0.5}$: allo-HCT | Reference | Counts | $t_{0.5}$: All | $t_{0.5}$: allo-HCT | aHR: allo-HCT (95%) | aHR: ex-allo-HCT (95%) | aHR: All (95%) |
|---|---|---|---|---|---|---|---|---|---|---|
| Age :: >75 | 75/91 | 248 | 248 | ≤75 | 147/212 | 182 | 158 | 1.04 (0.94,1.55) | 0.88 (0.66,0.98) | 1.33 (1.13,1.90) |
| CYT-Inv3 :: (+ve) | 5/5 | 27 | 27 | (-ve) | 207/286 | 225 | 196 | 5.91 (2.96,16.87) | NA | 4.85 (2.56,14.48) |
| CYT-Minus5 :: (+ve) | 9/9 | 28 | 28 | (-ve) | 203/282 | 225 | 194 | 1.37 (0.97,2.48) | 1.39 (1.05,3.42) | 1.49 (1.15,2.95) |
| CYT-Del7q :: (+ve) | 10/12 | 42 | 42 | (-ve) | 202/279 | 225 | 209 | 1.68 (1.16,3.04) | 1.41 (1.06,2.58) | 1.45 (1.00,2.74) |
| CYT-T911 :: (+ve) | 7/7 | 54 | 54 | (-ve) | 205/284 | 231 | 196 | 2.53 (1.66,7.46) | NA | 3.11 (2.10,7.66) |
| CYT-Minus7 :: (+ve) | 24/27 | 93 | 93 | (-ve) | 188/264 | 239 | 209 | 1.11 (0.90,1.70) | 1.13 (1.00,2.20) | 1.07 (0.74,1.34) |
| CYT-Complex :: (+ve) | 74/90 | 102 | 102 | (-ve) | 138/201 | 292 | 251 | 2.24 (1.28,5.05) | 1.21 (1.07,5.05) | 1.79 (1.25,5.36) |
| CYT-Minus17 :: (+ve) | 21/23 | 103 | 103 | (-ve) | 191/268 | 241 | 196 | 1.43 (1.17,2.90) | 1.15 (1.04,1.74) | 1.54 (1.24,3.38) |
| CYT-Monosomal :: (+ve) | 47/55 | 103 | 102 | (-ve) | 165/236 | 248 | 215 | 0.70 (0.31,0.95) | 1.01 (0.51,1.10) | 0.80 (0.37,1.01) |
| CYT risk -Poor :: (+ve) | 79/98 | 103 | 102 | (-ve) | 143/205 | 256 | 239 | 1.15 (1.03,1.89) | 1.09 (1.00,1.61) | 1.09 (0.94,1.48) |
| CYT risk -Indeterminant :: (+ve) | 54/73 | 194 | 194 | (-ve) | 168/230 | 215 | 186 | 1.59 (1.12,4.14) | 1.08 (0.99,3.07) | 1.38 (1.12,5.50) |
| CYT risk -Good :: (+ve) | 3/5 | 264 | 264 | (-ve) | 219/298 | 202 | 182 | 0.64 (0.14,1.00) | 0.73 (0.17,0.98) | 0.72 (0.24,1.08) |
| CYT-Del5q :: (+ve) | 23/32 | 272 | 225 | (-ve) | 189/259 | 202 | 189 | 0.67 (0.39,0.84) | 0.88 (0.42,1.00) | 0.66 (0.36,0.84) |
| CYT risk -Intermediate :: (+ve) | 88/128 | 275 | 239 | (-ve) | 134/175 | 166 | 158 | 1.29 (1.01,4.59) | 0.99 (0.92,2.89) | 1.16 (1.01,3.83) |
| CYT-Normal :: (+ve) | 70/108 | 397 | 275 | (-ve) | 142/183 | 147 | 147 | 0.83 (0.58,0.95) | 0.88 (0.43,0.98) | 0.85 (0.67,0.96) |

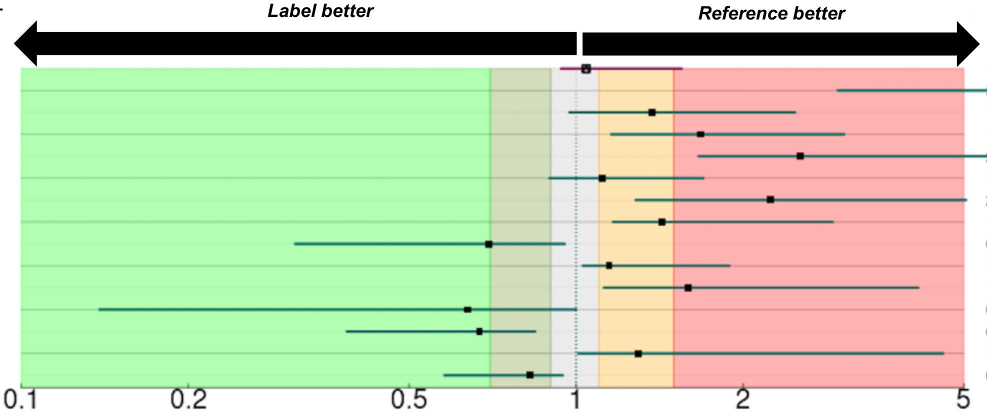


#### C. Fluorescence in situ hybridization (FISH).

| Label | Counts | $t_{0.5}$: All | $t_{0.5}$: allo-HCT | Reference | Counts | $t_{0.5}$: All | $t_{0.5}$: allo-HCT | aHR: allo-HCT (95%) | aHR: ex-allo-HCT (95%) | aHR: All (95%) |
|---|---|---|---|---|---|---|---|---|---|---|
| Age :: >75 | 75/91 | 248 | 248 | ≤75 | 147/212 | 182 | 158 | 0.98 (0.81,1.07) | 0.90 (0.71,0.99) | 1.13 (1.03,1.65) |
| FISH-MECOM [Rearrangement] :: (+ve) | 6/6 | 29 | 29 | (-ve) | 50/89 | 315 | 186 | 1.66 (1.20,2.98) | NA | 2.07 (1.56,4.95) |
| FISH-CHR 7 ABN [Loss] :: (+ve) | 50/57 | 38 | 38 | (-ve) | 140/207 | 278 | 241 | 1.32 (1.15,2.50) | 1.21 (1.06,1.67) | 1.29 (1.10,1.73) |
| FISH-RARA [Loss] :: (+ve) | 6/7 | 38 | 38 | (-ve) | 45/63 | 292 | 292 | 1.17 (0.96,2.02) | 1.21 (1.00,1.99) | 1.23 (1.00,2.18) |
| FISH-DEK [Loss] :: (+ve) | 3/6 | 44 | 44 | (-ve) | 57/94 | 202 | 152 | 1.03 (0.78,1.56) | NA | 0.98 (0.61,1.36) |
| FISH-CHR 7 ABN [Gain] :: (+ve) | 7/9 | 45 | 45 | (-ve) | 183/255 | 215 | 192 | 0.95 (0.55,1.12) | 0.92 (0.48,1.04) | 1.13 (0.96,2.36) |
| FISH-MLL [Rearrangement] :: (+ve) | 16/17 | 54 | 54 | (-ve) | 167/236 | 256 | 215 | 1.66 (1.20,4.12) | 1.43 (1.06,4.81) | 1.96 (1.33,4.71) |
| FISH-MECOM [Gain] :: (+ve) | 7/10 | 123.5 | 45 | (-ve) | 49/85 | 278 | 163 | 1.03 (0.85,1.43) | 1.33 (1.15,2.10) | 0.95 (0.23,1.14) |
| FISH-CHR 8 ABN [Loss] :: (+ve) | 11/13 | 158 | 158 | (-ve) | 126/159 | 251 | 239 | 1.35 (1.13,2.79) | 1.11 (1.00,2.38) | 1.30 (1.02,2.52) |
| FISH-DEK [Normal] :: (+ve) | 50/85 | 163 | 151 | (-ve) | 10/15 | 186 | 186 | 1.08 (1.03,1.55) | 1.05 (1.00,1.31) | 1.03 (0.98,1.27) |
| FISH-DEK [Gain] :: (+ve) | 6/8 | 186 | 186 | (-ve) | 54/92 | 272 | 151 | 0.94 (0.60,1.16) | NA | 1.06 (0.82,1.54) |
| FISH-CHR 5 ABN [Loss] :: (+ve) | 47/57 | 186 | 158 | (-ve) | 131/192 | 241 | 215 | 1.03 (0.94,1.14) | 1.03 (0.96,1.14) | 1.04 (0.92,1.17) |
| FISH-MLL [Gain] :: (+ve) | 29/36 | 196 | 152 | (-ve) | 154/217 | 241 | 209 | 0.93 (0.36,1.01) | 1.00 (0.51,1.06) | 0.91 (0.40,1.01) |
| FISH-CHR 8 ABN [Gain] :: (+ve) | 39/43 | 209 | 209 | (-ve) | 98/129 | 251 | 215 | 1.08 (1.00,1.43) | 1.03 (0.97,1.30) | 1.12 (1.01,1.60) |
| FISH-CHR 5 ABN [Normal] :: (+ve) | 125/186 | 256 | 215 | (-ve) | 53/63 | 158 | 152 | 0.96 (0.86,1.00) | 0.99 (0.92,1.06) | 0.93 (0.71,0.99) |
| FISH-CHR 8 ABN [Normal] :: (+ve) | 89/118 | 261 | 239 | (-ve) | 48/54 | 183 | 194 | 0.95 (0.85,1.00) | 0.96 (0.86,1.00) | 0.95 (0.86,1.01) |
| FISH-MLL [Normal] :: (+ve) | 135/197 | 272 | 239 | (-ve) | 48/56 | 102 | 102 | 0.86 (0.68,0.94) | 0.90 (0.68,0.98) | 0.85 (0.62,0.94) |
| FISH-CHR 5 ABN [Gain] :: (+ve) | 8/10 | 274 | 274 | (-ve) | 170/239 | 215 | 194 | 0.95 (0.69,1.13) | 0.97 (0.65,1.10) | 0.88 (0.55,1.07) |
| FISH-CHR 7 ABN [Normal] :: (+ve) | 133/198 | 278 | 241 | (-ve) | 57/66 | 43 | 43 | 0.88 (0.72,0.95) | 0.91 (0.78,0.98) | 0.86 (0.69,0.93) |
| FISH-RARA [Normal] :: (+ve) | 44/62 | 315 | 292 | (-ve) | 7/8 | 36 | 36 | 0.93 (0.78,1.00) | 0.90 (0.76,0.99) | 0.97 (0.81,1.03) |
| FISH-MECOM [Normal] :: (+ve) | 40/76 | 377 | 272 | (-ve) | 16/19 | 44 | 44 | 0.87 (0.18,0.94) | 0.90 (0.16,0.99) | 0.84 (0.15,0.93) |

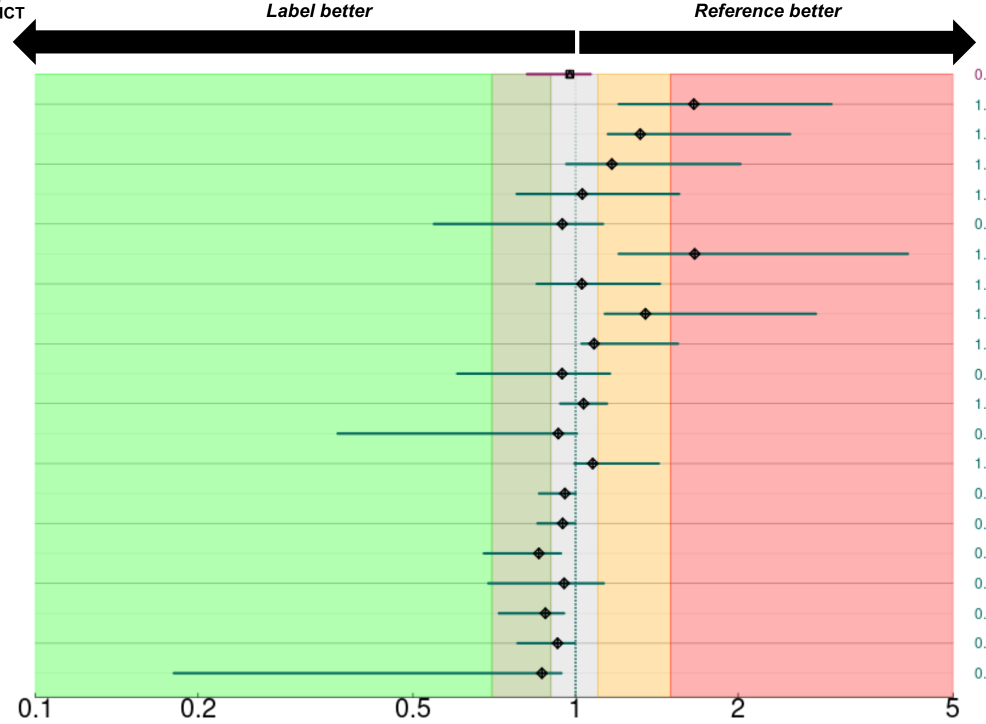

### Supplemental Table 4, cont'd

#### D. AML specific composite mutations.

| Label | Counts | $t_{0.5}$: All | $t_{0.5}$: allo-HCT | Reference | Counts | $t_{0.5}$: All | $t_{0.5}$: allo-HCT | aHR: allo-HCT (95%) | aHR: ex-allo-HCT (95%) | aHR: All (95%) |
|---|---|---|---|---|---|---|---|---|---|---|
| Age :: >75 | 75/91 | 248 | 248 | ≤75 | 147/212 | 182 | 158 | 0.89 (0.63,1.00) | 0.82 (0.57,0.98) | 1.10 (1.02,1.38) |
| PCR/NGS-JAK2 :: (+ve) | 9/11 | 49 | 49 | (-ve) | 199/272 | 215 | 194 | 1.42 (1.09,2.76) | 1.33 (1.01,2.39) | 1.24 (1.00,2.04) |
| FISH/NGS-TP53 :: (+ve) | 55/67 | 103 | 93 | (-ve) | 155/217 | 256 | 241 | 1.30 (1.14,1.77) | 1.27 (1.07,1.75) | 1.16 (1.02,1.57) |
| FISH/PCR/NGS-RUNX :: (+ve) | 78/104 | 196 | 186 | (-ve) | 136/185 | 215 | 192 | 1.04 (0.94,1.23) | 1.07 (0.99,1.30) | 0.99 (0.78,1.05) |
| FISH/PCR-CBFB :: (+ve) | 17/20 | 202 | 225 | (-ve) | 155/217 | 251 | 194 | 0.96 (0.64,1.08) | 1.03 (0.89,1.29) | 1.09 (1.00,1.41) |
| PCR/NGS-FLT3 :: (+ve) | 44/62 | 206 | 181 | (-ve) | 169/226 | 202 | 189 | 1.03 (0.94,1.42) | 1.04 (0.95,1.36) | 1.04 (0.97,1.36) |
| PCR/NGS-FLT3ITD :: (+ve) | 24/33 | 206 | 181 | (-ve) | 189/255 | 202 | 189 | 1.09 (0.94,1.74) | 1.13 (0.97,2.02) | 1.08 (0.99,1.75) |
| FISH/PCR/NGS-PML :: (+ve) | 13/16 | 225 | 225 | (-ve) | 171/234 | 231 | 196 | 0.89 (0.54,1.02) | 0.93 (0.58,1.04) | 0.99 (0.69,1.09) |
| PCR/NGS-CEBPA :: (+ve) | 12/20 | 314 | 314 | (-ve) | 194/261 | 202 | 189 | 0.90 (0.58,1.04) | 0.90 (0.61,1.01) | 0.90 (0.56,1.00) |
| PCR/NGS-NPM1 :: (+ve) | 37/59 | 315 | 314 | (-ve) | 172/225 | 152 | 147 | 0.67 (0.38,0.83) | 0.74 (0.41,0.94) | 0.79 (0.42,0.91) |
| PCR/NGS-IDH2 :: (+ve) | 27/44 | 582 | 398 | (-ve) | 185/243 | 183 | 166 | 0.77 (0.49,0.92) | 0.90 (0.64,1.01) | 0.74 (0.36,0.87) |
| PCR/NGS-IDH1 :: (+ve) | 13/24 | 922 | 314 | (-ve) | 199/263 | 182 | 164 | 0.70 (0.40,0.86) | 0.78 (0.49,1.00) | 0.69 (0.33,0.84) |

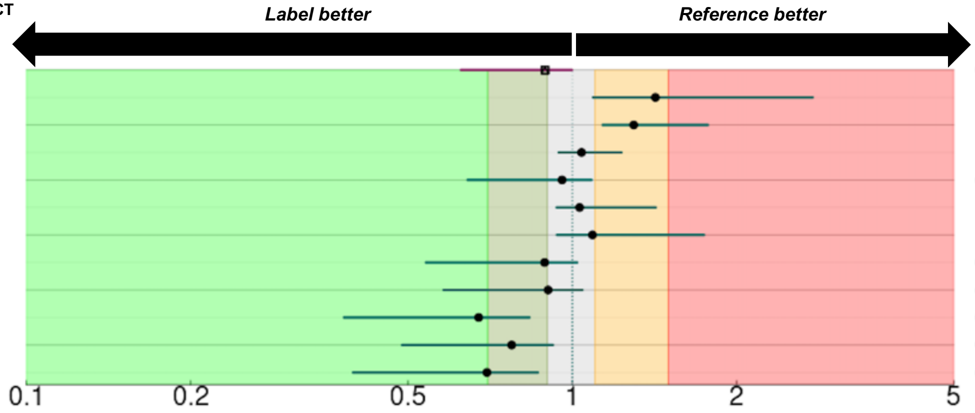


#### E. Flow cytometry (

| Label | Counts | $t_{0.5}$: All | $t_{0.5}$: allo-HCT | Reference | Counts | $t_{0.5}$: All | $t_{0.5}$: allo-HCT | aHR: allo-HCT (95%) | aHR: ex-allo-HCT (95%) | aHR: All (95%) |
|---|---|---|---|---|---|---|---|---|---|---|
| Age :: >75 | 75/91 | 248 | 248 | ≤75 | 147/212 | 182 | 158 | 0.97 (0.79,1.00) | 0.94 (0.73,1.00) | 1.09 (1.03,1.40) |
| FC-CD16 :: (+ve) | 5/5 | 28 | 28 | (-ve) | 163/236 | 215 | 192 | 2.09 (1.66,5.75) | NA | 1.73 (1.43,3.89) |
| FC-CD36 :: (+ve) | 30/36 | 44.5 | 44.5 | (-ve) | 81/119 | 256 | 176 | 1.23 (1.09,1.70) | 1.10 (1.00,1.49) | 1.23 (1.09,1.79) |
| FC-CD71 :: (+ve) | 6/7 | 45 | 45 | (-ve) | 5/8 | 431.5 | 342 | 1.44 (1.24,3.20) | 1.14 (1.01,1.71) | 1.28 (1.09,2.26) |
| FC-CD56 :: (+ve) | 58/75 | 103 | 99 | (-ve) | 135/192 | 202 | 189 | 1.18 (1.09,1.57) | 1.05 (1.00,1.36) | 1.13 (1.03,1.60) |
| FC-CD11B :: (+ve) | 70/81 | 136 | 136 | (-ve) | 131/194 | 261 | 196 | 1.18 (1.08,1.59) | 1.04 (1.00,1.28) | 1.22 (1.11,1.78) |
| FC-CD7 :: (+ve) | 60/81 | 142 | 142 | (-ve) | 138/192 | 231 | 196 | 1.10 (1.01,1.41) | 1.07 (1.00,1.37) | 1.08 (1.00,1.29) |
| FC-CD64 :: (+ve) | 72/92 | 164 | 152 | (-ve) | 125/179 | 231 | 189 | 1.08 (1.01,1.34) | 1.04 (1.00,1.29) | 1.08 (1.01,1.38) |
| FC-CD5 :: (+ve) | 6/7 | 176 | 176 | (-ve) | 43/64 | 147 | 117 | 1.25 (1.05,2.12) | 1.04 (1.00,1.39) | 1.25 (1.06,2.56) |
| FC-CD34 :: (+ve) | 175/233 | 182 | 158 | (-ve) | 41/61 | 315 | 292 | 1.33 (1.19,1.93) | 1.11 (1.00,1.55) | 1.19 (1.09,1.89) |
| FC-CD33 :: (+ve) | 183/250 | 183 | 164 | (-ve) | 27/37 | 342 | 256 | 1.07 (1.00,1.64) | 1.02 (0.99,1.15) | 1.03 (1.00,1.19) |
| FC-CD14 :: (+ve) | 12/16 | 183.5 | 152 | (-ve) | 193/265 | 194 | 182 | 0.86 (0.43,1.00) | 0.93 (0.53,1.00) | 0.91 (0.52,1.00) |
| FC-CD123 :: (+ve) | 90/128 | 186 | 166 | (-ve) | 20/31 | 62 | 62 | 0.98 (0.78,1.00) | 1.00 (0.98,1.07) | 0.95 (0.65,1.00) |
| FC-CD13 :: (+ve) | 189/253 | 186 | 166 | (-ve) | 21/34 | 582 | 342 | 1.11 (1.01,1.56) | 1.02 (0.99,1.19) | 1.12 (1.02,1.64) |
| FC-TDT :: (+ve) | 16/24 | 186 | 174.5 | (-ve) | 129/178 | 189 | 176 | 0.94 (0.63,1.00) | 1.00 (0.86,1.06) | 0.89 (0.59,1.00) |
| FC-CD38 :: (+ve) | 189/256 | 189 | 181 | (-ve) | 14/23 | 391 | 261 | 1.07 (1.00,1.43) | 1.03 (1.00,1.36) | 1.04 (1.00,1.39) |
| FC-HLADR :: (+ve) | 180/242 | 192 | 176 | (-ve) | 32/47 | 256 | 256 | 0.97 (0.73,1.00) | 1.01 (0.98,1.19) | 1.00 (0.96,1.15) |
| FC-CD117 :: (+ve) | 202/274 | 206 | 186 | (-ve) | 13/19 | 71 | 71 | 1.07 (1.00,1.44) | 1.04 (1.00,1.30) | 1.03 (1.00,1.30) |
| FC-CD22 :: (+ve) | 6/6 | 214.5 | 214.5 | (-ve) | 155/216 | 192 | 181 | 1.29 (1.05,2.69) | NA | 1.22 (1.03,2.24) |
| FC-CD45 :: (+ve) | 143/188 | 215 | 186 | (-ve) | 38/64 | 215 | 251 | 1.03 (1.00,1.27) | 0.99 (0.90,1.02) | 1.05 (1.00,1.31) |
| FC-CD15 :: (+ve) | 24/31 | 225 | 225 | (-ve) | 122/173 | 182 | 176 | 0.86 (0.54,0.99) | 0.92 (0.66,1.00) | 0.93 (0.64,1.00) |
| FC-MPO :: (+ve) | 112/154 | 248 | 215 | (-ve) | 72/99 | 152 | 176 | 0.90 (0.69,0.97) | 0.96 (0.78,1.00) | 0.93 (0.78,1.00) |
| FC-CD10 :: (+ve) | 8/12 | 293.5 | 272 | (-ve) | 130/194 | 209 | 186 | 0.68 (0.37,0.88) | 0.86 (0.40,1.00) | 0.86 (0.46,1.00) |
| FC-CD11C :: (+ve) | 3/5 | 398 | 71 | (-ve) | 6/11 | 521 | 521 | 1.12 (1.00,3.22) | NA | 0.96 (0.50,1.02) |
| FC-CD19 :: (+ve) | 5/7 | 402 | 402 | (-ve) | 195/269 | 194 | 181 | 0.79 (0.43,0.98) | 0.88 (0.49,1.00) | 0.90 (0.45,1.00) |

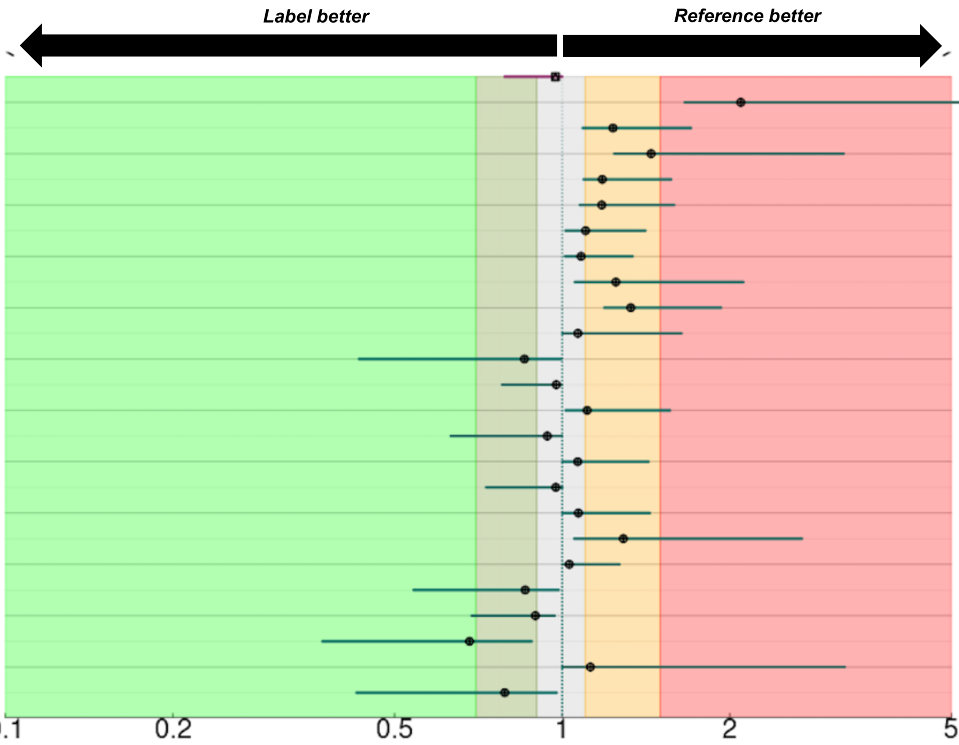

**Supplemental Table 5. Covariate-level classifications across different combinations.**

**A. Relationship among Rule-I, II, III, and IV.**

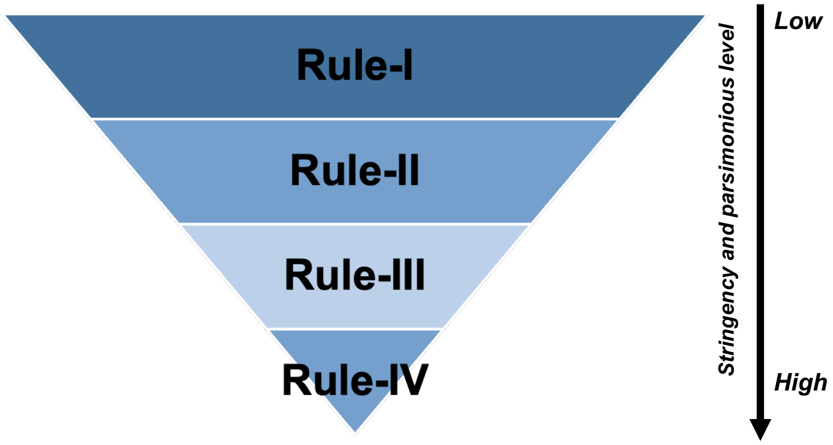


**B. Covariate-level risk classification based on Rule-I.**

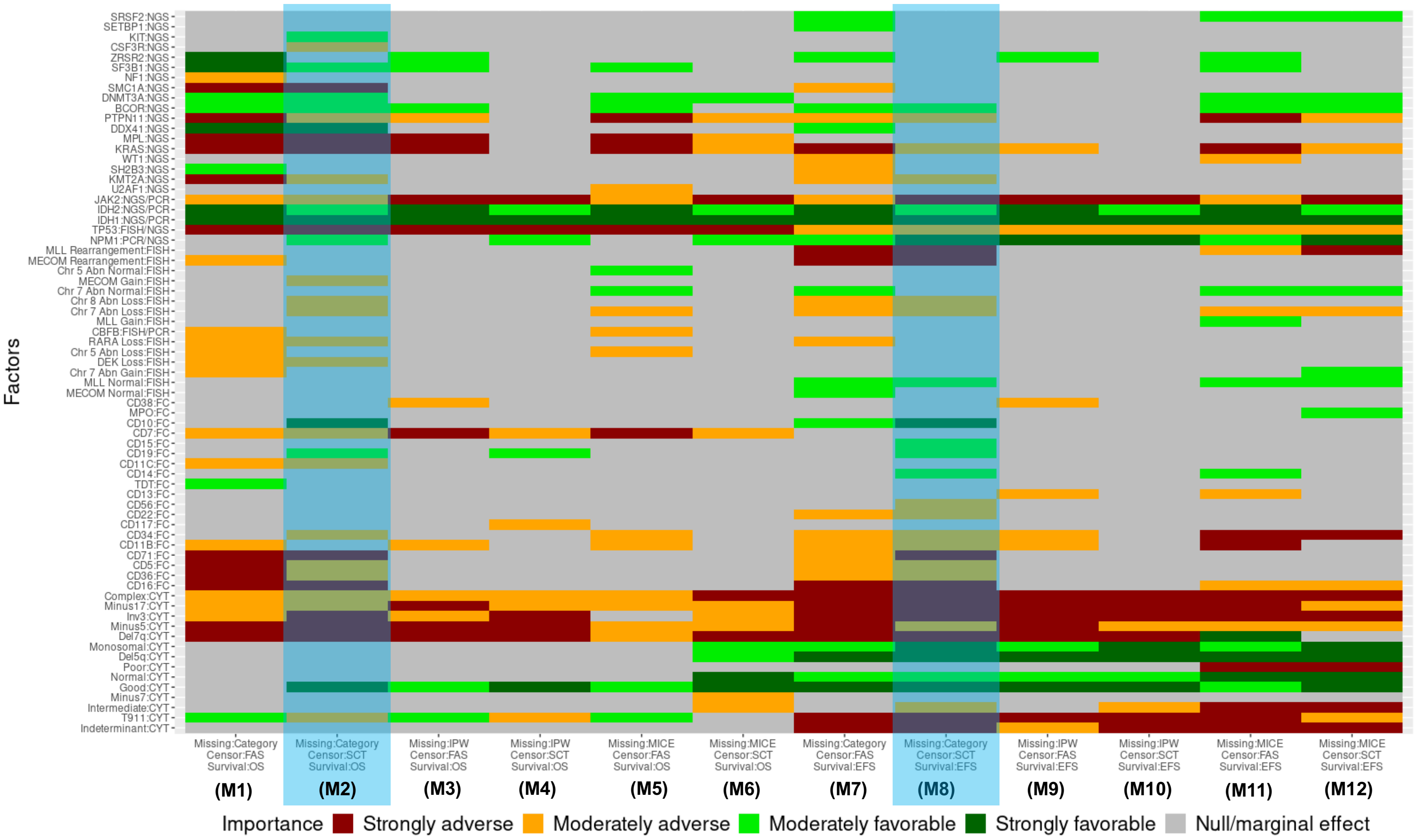

**Supplemental Table 5, cont'd**

### C. Covariate-level risk classification based on Rule-II.

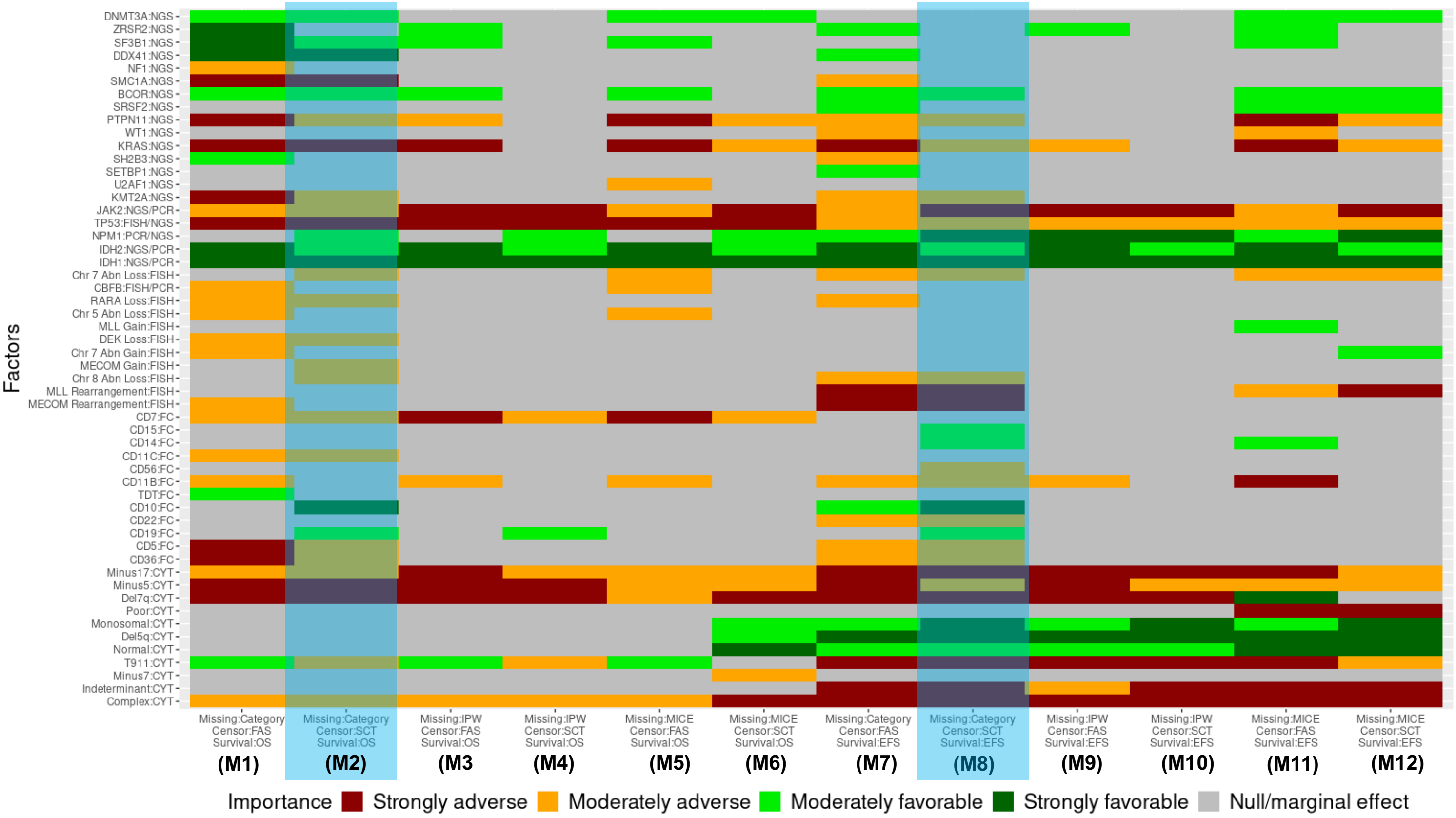


### D. Covariate-level risk classification based on Rule-III.

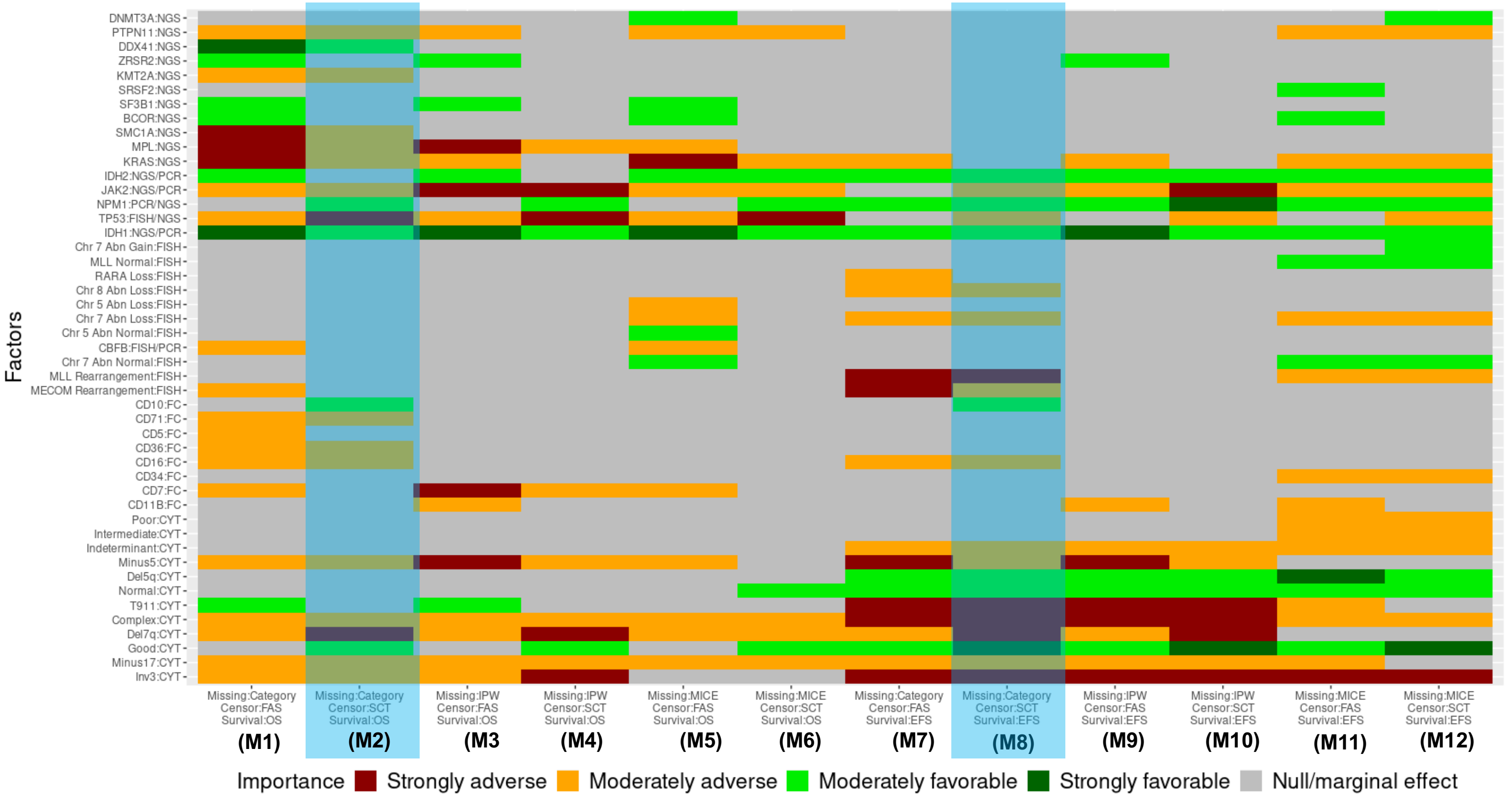

**Supplemental Table 5, cont'd**

**E. Covariate-level risk classification based on Rule-IV.**

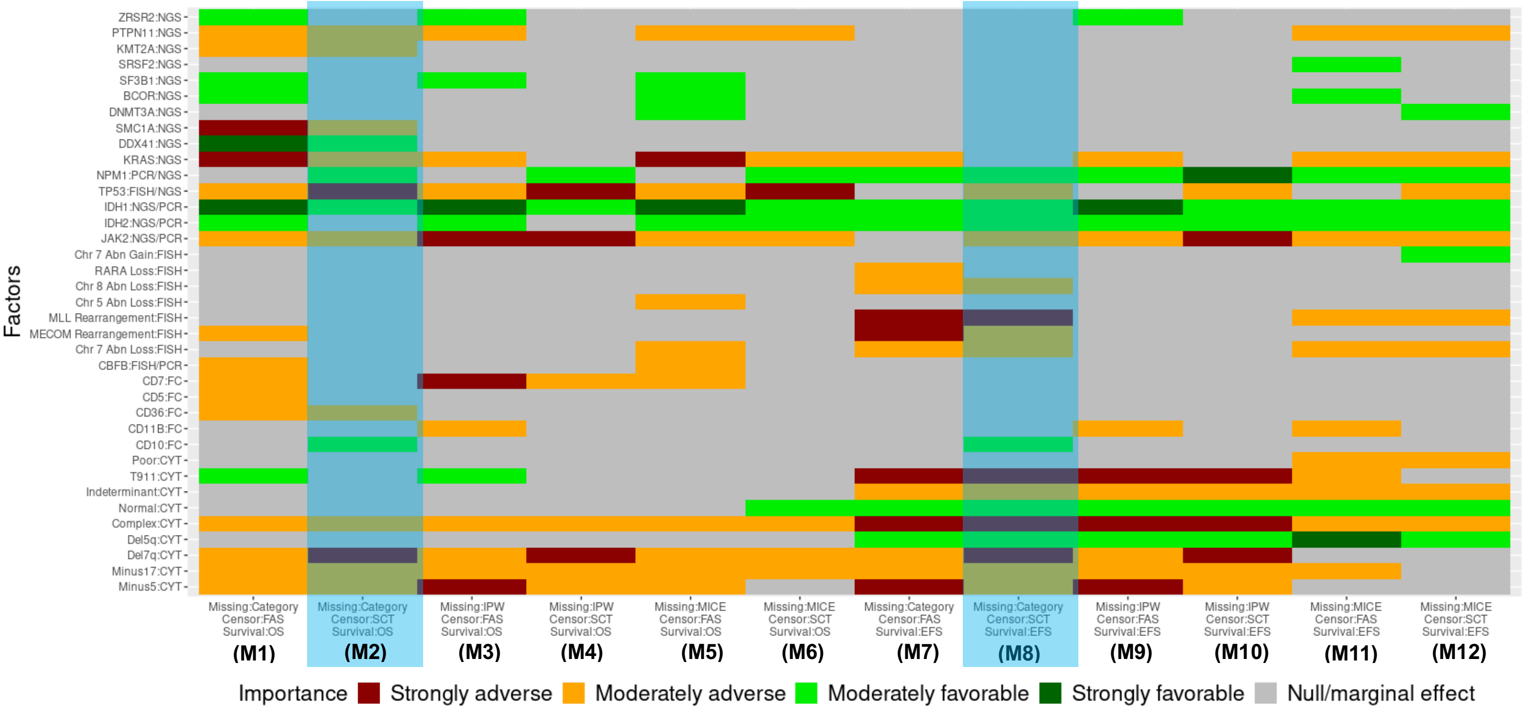

**Supplemental Table 6. Predictive validation of risk models for overall survival (OS).**

**A. OS, Rule-I.**

| | | Kaplan-Meier | | | mCOXr[a] | | |
|---|---|---|---|---|---|---|---|
| **Type of risk** | **Covariate-level classification for proposed** | **M-iAUC ($25^{th}$,$75^{th}$) over *t*** | **M-cAUC ($25^{th}$,$75^{th}$) over *t*** | **iBrier ($25^{th}$,$75^{th}$) over *t*** | **M-iAUC ($25^{th}$,$75^{th}$) over *t*** | **M-cAUC ($25^{th}$,$75^{th}$) over *t*** | **iBrier ($25^{th}$,$75^{th}$) over *t*** |
| **ELN risk** | | 0.56 [0.53,0.58] | 0.57 [<0.50,0.67] | 0.17 [0.12,0.19] | 0.55 [0.54,0.57] | 0.50 [<0.50,0.58] | 0.16 [0.13,0.20] |
| **Genetic plus phenotypic based risk signatures ($RM_{GP,I}$)** | (a) Retaining original list assuming fixed | 0.59 [0.56,0.64] (<0.001) | 0.64 [0.60,0.69] (<0.001) | 0.15 [0.12,0.20] | 0.58 [0.57,0.65] (<0.001) | 0.65 [0.62,0.70] (<0.001) | 0.15 [0.13,0.20] |
| | (b) Ensemble over $k$-CV | 0.58 [0.56,0.62] (<0.001) | 0.62 [0.60,0.68] (<0.001) | 0.15 [0.12,0.20] | 0.59 [0.56,0.62] (<0.001) | 0.62 [0.58,0.66] (<0.001) | 0.16 [0.13,0.20] |
| | (c) Ensemble over $k$-CV plus missing methods | 0.60 [0.55,0.65] (<0.001) | 0.62 [0.59,0.72] (<0.001) | 0.14 [0.10,0.18] | 0.59 [0.56,0.68] (<0.001) | 0.62 [0.54,0.70] (<0.001) | 0.14 [0.10,0.18] |
| | (d) Independent classification with in each $k$-CV | 0.55 [0.52,0.59] (0.129) | 0.53 [<0.50,0.65] (0.592) | 0.15 [0.13,0.21] | 0.55 [0.52,0.59] (<0.001) | 0.54 [<0.50,0.62] (<0.001) | 0.15 [0.13,0.21] |
| **Genetic based risk signatures ($RM_{G,I}$)** | (a) Retaining original list assuming fixed | 0.61 [0.58,0.62] (<0.001) | 0.67 [0.65,0.75] (<0.001) | 0.14 [0.12,0.18] | 0.63 [0.59,0.65] (<0.001) | 0.70 [0.63,0.76] (<0.001) | 0.14 [0.12,0.18] |
| | (b) Ensemble over $k$-CV | 0.58 [0.56,0.62] (<0.001) | 0.61 [0.58,0.67] (<0.001) | 0.15 [0.12,0.19] | 0.62 [0.55,0.63] (<0.001) | 0.66 [0.63,0.70] (<0.001) | 0.15 [0.13,0.18] |
| | (c) Ensemble over $k$-CV plus missing methods | 0.61 [0.58,0.64] (<0.001) | 0.68 [0.58,0.71] (<0.001) | 0.15 [0.11,0.18] | 0.63 [0.57,0.66] (<0.001) | 0.64 [0.57,0.69] (<0.001) | 0.15 [0.11,0.18] |
| | (d) Independent classification with in each $k$-CV | 0.57 [0.54,0.59] (0.350) | 0.60 [0.53,0.63] (<0.001) | 0.16 [0.12,0.20] | 0.56 [0.56,0.59] (<0.001) | 0.58 [0.52,0.66] (<0.001) | 0.15 [0.13,0.20] |

[a]Penalized CoxPH model adjusted by age, gender, race, and either ELN or proposed risk stratification variable (i.e., $RM_{GP,I}$, $RM_{G,I}$).

**Remarks:**

| | |
|---|---|
| The higher iAUC or cAUC value (i.e., close to 1) means better numerical performance | The lower iBrier value (i.e., close to 0) means better numerical performance |

Reported in parenthesis are the median of P-values (over 15-fold CV) testing the null hypothesis of inferiority of distributions of survival metrics of the proposed relative to ELN22.

**Supplemental Table 6, cont'd**

**B. OS, Rule-II.**

| | | Kaplan-Meier | | | mCOXr[a] | | |
|---|---|---|---|---|---|---|---|
| **Type of risk** | **Covariate-level classification for proposed** | **M-iAUC (25th,75th) over *t*** | **M-cAUC (25th,75th) over *t*** | **iBrier (25th,75th) over *t*** | **M-iAUC (25th,75th) over *t*** | **M-cAUC (25th,75th) over *t*** | **iBrier (25th,75th) over *t*** |
| **ELN risk** | | 0.56 [0.53,0.58] | 0.57 [0.48,0.67] | 0.17 [0.12,0.19] | 0.55 [0.54,0.57] | 0.50 [0.38,0.58] | 0.16 [0.13,0.20] |
| **Genetic plus phenotypic based risk signatures ($RM_{GP,II}$)** | (a) Retaining original list assuming fixed | 0.63 [0.57,0.66] (<0.001) | 0.70 [0.64,0.75] (<0.001) | 0.14 [0.11,0.18] | 0.64 [0.57,0.69] (<0.001) | 0.71 [0.67,0.74] (<0.001) | 0.14 [0.12,0.18] |
| | (b) Ensemble over $k$-CV | 0.60 [0.55,0.64] (<0.001) | 0.66 [0.61,0.72] (<0.001) | 0.15 [0.11,0.19] | 0.62 [0.56,0.66] (<0.001) | 0.67 [0.57,0.71] (<0.001) | 0.15 [0.12,0.19] |
| | (c) Ensemble over $k$-CV plus missing methods | 0.59 [0.55,0.64] (<0.001) | 0.62 [0.59,0.70] (<0.001) | 0.15 [0.11,0.20] | 0.59 [0.55,0.67] (<0.001) | 0.61 [0.53,0.69] (<0.001) | 0.16 [0.11,0.20] |
| | (d) Independent classification with in each $k$-CV | 0.57 [0.55,0.60] (0.633) | 0.61 [0.52,0.66] (0.001) | 0.14 [0.13,0.20] | 0.58 [0.56,0.61] (<0.001) | 0.60 [<0.50,0.68] (<0.001) | 0.14 [0.13,0.20] |
| **Genetic based risk signatures ($RM_{G,II}$)** | (a) Retaining original list assuming fixed | 0.61 [0.58,0.64] (<0.001) | 0.69 [0.65,0.73] (<0.001) | 0.14 [0.12,0.18] | [b]0.64 [0.59,0.66] (<0.001) | [b]0.70 [0.65,0.77] (<0.001) | [b]0.14 [0.12,0.19] |
| | (b) Ensemble over $k$-CV | 0.59 [0.56,0.63] (0.004) | 0.63 [0.58,0.68] (<0.001) | 0.15 [0.12,0.20] | 0.61 [0.57,0.64] (<0.001) | 0.64 [0.58,0.69] (<0.001) | 0.15 [0.12,0.19] |
| | (c) Ensemble over $k$-CV plus missing methods | 0.60 [0.57,0.64] (<0.001) | 0.61 [0.57,0.71] (<0.001) | 0.15 [0.11,0.20] | 0.62 [0.57,0.65] (<0.001) | 0.64 [0.56,0.71] (<0.001) | 0.16 [0.11,0.19] |
| | (d) Independent classification with in each $k$-CV | 0.57 [0.55,0.60] (0.395) | 0.61 [0.53,0.64] (<0.001) | 0.15 [0.12,0.20] | 0.57 [0.56,0.59] (<0.001) | 0.63 [0.53,0.67] (<0.001) | 0.15 [0.13,0.20] |

[a]Penalized CoxPH model adjusted by age, gender, race, and either ELN or proposed risk stratification variable (i.e., $RM_{GP,II}$, $RM_{G,II}$). [b]Reported in the Figure 4F.

Remarks:

| | |
|---|---|
| The higher iAUC or cAUC value (i.e., close to 1) means better numerical performance | The lower iBrier value (i.e., close to 0) means better numerical performance |

Reported in parenthesis are the median of P-values (over 15-fold CV) testing the null hypothesis of inferiority of distributions of survival metrics of the proposed relative to ELN22.

**Supplemental Table 6, cont'd**

**C. OS, Rule-III.**

| | | Kaplan-Meier | | | mCOXr[a] | | |
|---|---|---|---|---|---|---|---|
| **Type of risk** | **Covariate-level classification for proposed** | **M-iAUC ($25^{th}$,$75^{th}$) over *t*** | **M-cAUC ($25^{th}$,$75^{th}$) over *t*** | **iBrier ($25^{th}$,$75^{th}$) over *t*** | **M-iAUC ($25^{th}$,$75^{th}$) over *t*** | **M-cAUC ($25^{th}$,$75^{th}$) over *t*** | **iBrier ($25^{th}$,$75^{th}$) over *t*** |
| **ELN risk** | | 0.56 [0.53,0.58] | 0.57 [<0.50,0.67] | 0.17 [0.12,0.19] | 0.55 [0.54,0.57] | 0.50 [<0.50,0.58] | 0.16 [0.13,0.20] |
| **Genetic plus phenotypic based risk signatures ($RM_{GP,III}$)** | (a) Retaining original list assuming fixed | 0.61 [0.53,0.66] (<0.001) | 0.67 [0.56,0.73] (<0.001) | 0.16 [0.12,0.17] | 0.62 [0.56,0.64] (<0.001) | 0.66 [0.58,0.73] (<0.001) | 0.16 [0.12,0.17] |
| | (b) Ensemble over *k*-CV | 0.60 [0.56,0.65] (<0.001) | 0.66 [0.57,0.72] (<0.001) | 0.16 [0.12,0.17] | 0.61 [0.57,0.66] (<0.001) | 0.67 [0.60,0.71] (<0.001) | 0.16 [0.12,0.17] |
| | (c) Ensemble over *k*-CV plus missing methods | 0.59 [0.57,0.64] (<0.001) | 0.64 [0.57,0.72] (<0.001) | 0.16 [0.12,0.17] | 0.61 [0.57,0.65] (<0.001) | 0.65 [0.59,0.68] (<0.001) | 0.16 [0.12,0.18] |
| | (d) Independent classification with in each *k*-CV | 0.55 [0.52,0.60] (1.00) | 0.55 [<0.50,0.65] (0.012) | 0.16 [0.13,0.20] | 0.54 [0.53,0.58] (<0.001) | 0.55 [<0.50,0.62] (<0.001) | 0.16 [0.13,0.20] |
| **Genetic based risk signatures ($RM_{G,III}$)** | (a) Retaining original list assuming fixed | 0.60 [0.56,0.64] (<0.001) | 0.64 [0.60,0.71] (<0.001) | 0.16 [0.12,0.17] | 0.61 [0.56,0.65] (<0.001) | 0.62 [0.60,0.68] (<0.001) | 0.16 [0.12,0.18] |
| | (b) Ensemble over *k*-CV | 0.59 [0.57,0.63] (<0.001) | 0.64 [0.56,0.70] (<0.001) | 0.16 [0.12,0.17] | 0.61 [0.57,0.63] (<0.001) | 0.62 [0.59,0.69] (<0.001) | 0.16 [0.12,0.18] |
| | (c) Ensemble over *k*-CV plus missing methods | 0.59 [0.57,0.61] (<0.001) | 0.64 [0.60,0.70] (<0.001) | 0.16 [0.12,0.18] | 0.61 [0.57,0.63] (<0.001) | 0.62 [0.59,0.69] (<0.001) | 0.16 [0.12,0.18] |
| | (d) Independent classification with in each *k*-CV | 0.56 [0.53,0.61] (0.960) | 0.59 [0.51,0.63] (0.001) | 0.16 [0.13,0.19] | 0.56 [0.53,0.61] (<0.001) | 0.57 [0.52,0.64] (<0.001) | 0.16 [0.13,0.19] |

[a]Penalized CoxPH model adjusted by age, gender, race, and either ELN or proposed risk stratification variable (i.e., $RM_{GP,III}$, $RM_{G,III}$).

Remarks:

| The higher iAUC or cAUC value (i.e., close to 1) means better numerical performance | The lower iBrier value (i.e., close to 0) means better numerical performance |
|---|---|

Reported in parenthesis are the median of P-values (over 15-fold CV) testing the null hypothesis of inferiority of distributions of survival metrics of the proposed relative to ELN22.

**Supplemental Table 6, cont'd**

**D. OS, Rule-IV.**

| | | Kaplan-Meier | | | mCOXr[a] | | |
|---|---|---|---|---|---|---|---|
| **Type of risk** | **Covariate-level classification for proposed** | **M-iAUC ($25^{th}$,$75^{th}$) over $t$** | **M-cAUC ($25^{th}$,$75^{th}$) over $t$** | **iBrier ($25^{th}$,$75^{th}$) over $t$** | **M-iAUC ($25^{th}$,$75^{th}$) over $t$** | **M-cAUC ($25^{th}$,$75^{th}$) over $t$** | **iBrier ($25^{th}$,$75^{th}$) over $t$** |
| **ELN risk** | | 0.56 [0.53,0.58] | 0.57 [0.48,0.67] | 0.17 [0.12,0.19] | 0.55 [0.54,0.57] | 0.50 [0.38,0.58] | 0.16 [0.13,0.20] |
| **Genetic plus phenotypic based risk signatures ($RM_{GP,IV}$)** | (a) Retaining original list assuming fixed | 0.60 [0.54,0.65] (<0.001) | 0.64 [0.57,0.69] (<0.001) | 0.16 [0.12,0.18] | 0.62 [0.54,0.68] (<0.001) | 0.62 [0.57,0.70] (<0.001) | 0.16 [0.12,0.19] |
| | (b) Ensemble over $k$-CV | 0.59 [0.57,0.63] (<0.001) | 0.63 [0.58,0.70] (<0.001) | 0.16 [0.12,0.18] | 0.60 [0.56,0.64] (<0.001) | 0.64 [0.59,0.67] (<0.001) | 0.16 [0.12,0.19] |
| | (c) Ensemble over $k$-CV plus missing methods | 0.60 [0.55,0.62] (<0.001) | 0.63 [0.61,0.70] (<0.001) | 0.16 [0.12,0.19] | 0.60 [0.55,0.63] (<0.001) | 0.64 [0.58,0.66] (<0.001) | 0.16 [0.12,0.19] |
| | (d) Independent classification with in each $k$-CV | 0.55 [0.52,0.60] (0.997) | 0.56 [<0.50,0.63] (0.001) | 0.16 [0.14,0.20] | 0.54 [0.53,0.57] (0.983) | 0.54 [<0.50,0.62] (<0.001) | 0.16 [0.13,0.20] |
| **Genetic based risk signatures ($RM_{G,IV}$)** | (a) Retaining original list assuming fixed | 0.61 [0.56,0.65] (<0.001) | 0.66 [0.61,0.70] (<0.001) | 0.16 [0.12,0.18] | 0.60 [0.56,0.66] (<0.001) | 0.65 [0.59,0.69] (<0.001) | 0.16 [0.12,0.18] |
| | (b) Ensemble over $k$-CV | 0.59 [0.57,0.63] (<0.001) | 0.63 [0.58,0.70] (<0.001) | 0.16 [0.12,0.18] | 0.60 [0.56,0.64] (<0.001) | 0.64 [0.59,0.67] (<0.001) | 0.16 [0.12,0.19] |
| | (c) Ensemble over $k$-CV plus missing methods | 0.60 [0.55,0.62] (<0.001) | 0.63 [0.61,0.70] (<0.001) | 0.16 [0.12,0.19] | 0.60 [0.55,0.63] (<0.001) | 0.64 [0.58,0.66] (<0.001) | 0.16 [0.12,0.19] |
| | (d) Independent classification with in each $k$-CV | 0.56 [0.53,0.61] (0.964) | 0.59 [0.51,0.63] (0.001) | 0.16 [0.14,0.19] | 0.55 [0.53,0.61] (<0.001) | 0.58 [0.52,0.64] (<0.001) | 0.16 [0.13,0.19] |

[a]Penalized CoxPH model adjusted by age, gender, race, and either ELN or proposed risk stratification variable (i.e., $RM_{GP,IV}$, $RM_{G,IV}$).

Remarks:

| | |
|---|---|
| The higher iAUC or cAUC value (i.e., close to 1) means better numerical performance | The lower iBrier value (i.e., close to 0) means better numerical performance |

Reported in parenthesis are the median of P-values (over 15-fold CV) testing the null hypothesis of inferiority of distributions of survival metrics of the proposed relative to ELN22.

**Supplemental Table 7. Predictive validation of risk models for event free survival (EFS).**

### A. EFS, Rule-I.

| | | Kaplan-Meier | | | mCOXr[a] | | |
|---|---|---|---|---|---|---|---|
| **Type of risk** | **Covariate-level classification for proposed** | **M-iAUC ($25^{th}$,$75^{th}$) over $t$** | **M-cAUC ($25^{th}$,$75^{th}$) over $t$** | **iBrier ($25^{th}$,$75^{th}$) over $t$** | **M-iAUC ($25^{th}$,$75^{th}$) over $t$** | **M-cAUC ($25^{th}$,$75^{th}$) over $t$** | **iBrier ($25^{th}$,$75^{th}$) over $t$** |
| **ELN risk** | | 0.53 [0.51,0.55] | 0.51 [<0.50,0.57] | 0.18 [0.14,0.21] | 0.54 [0.51.58] | 0.56 [0.50,0.62] | 0.18 [0.14,0.22] |
| **Genetic plus phenotypic based risk signatures ($RM_{GP,I\text{-}EFS}$)** | (a) Retaining original list assuming fixed | 0.51 [0.50,0.52] (1.00) | 0.57 [0.54,0.62] (<0.001) | 0.20 [0.14,0.23] | 0.52 [0.52,0.55] (0.511) | 0.60 [<0.50,0.63] (0.450) | 0.19 [0.14,0.23] |
| | (b) Ensemble over $k$-CV | 0.54 [0.53,0.56] (0.755) | 0.63 [0.58,0.69] (<0.001) | 0.15 [0.14,0.22] | 0.56 [0.55,0.58] (<0.001) | 0.64 [0.59,0.70] (<0.001) | 0.17 [0.14,0.23] |
| | (c) Ensemble over $k$-CV plus missing methods | 0.51 [0.50,0.52] (1.00) | 0.60 [0.56,0.63] (<0.001) | 0.19 [0.14,0.23] | 0.53 [0.51,0.55] (1.00) | 0.55 [0.52,0.66] (0.030) | 0.19 [0.14,0.24] |
| | (d) Independent classification with in each $k$-CV | 0.53 [0.51,0.54] (0.044) | 0.56 [0.52,0.64] (0.004) | 0.18 [0.14,0.25] | 0.54 [0.52,0.56] (0.729) | 0.55 [0.48,0.69] (0.001) | 0.18 [0.14,0.24] |
| **Genetic based risk signatures ($RM_{G,I\text{-}EFS}$)** | (a) Retaining original list assuming fixed | 0.55 [0.54,0.57] (<0.001) | 0.65 [0.60,0.70] (<0.001) | 0.18 [0.14,0.21] | 0.58 [0.56,0.63] (<0.001) | 0.63 [0.59,0.72] (<0.001) | 0.17 [0.15,0.21] |
| | (b) Ensemble over $k$-CV | 0.56 [0.54,0.57] (0.006) | 0.66 [0.60,0.70] (<0.001) | 0.18 [0.14,0.21] | 0.59 [0.56,0.61] (<0.001) | 0.65 [0.61,0.73] (<0.001) | 0.17 [0.14,0.21] |
| | (c) Ensemble over $k$-CV plus missing methods | 0.56 [0.54,0.57] (0.006) | 0.66 [0.60,0.70] (<0.001) | 0.18 [0.14,0.21] | 0.59 [0.56,0.61] (<0.001) | 0.65 [0.61,0.73] (<0.001) | 0.17 [0.14,0.21] |
| | (d) Independent classification with in each $k$-CV | 0.55 [0.52,0.56] (0.015) | 0.59 [0.51,0.63] (0.001) | 0.18 [0.14,0.23] | 0.56 [0.53,0.61] (0.008) | 0.59 [0.52,0.61] (<0.001) | 0.18 [0.14,0.24] |

[a]Penalized CoxPH model adjusted by age, gender, race, and either ELN or proposed risk stratification variable (i.e., $RM_{GP,I\text{-}EFS}$, $RM_{G,I\text{-}EFS}$).

Remarks:

| | |
|---|---|
| The higher iAUC or cAUC value (i.e., close to 1) means better numerical performance | The lower iBrier value (i.e., close to 0) means better numerical performance |

Reported in parenthesis are the median of P-values (over 15-fold CV) testing the null hypothesis of inferiority of distributions of survival metrics of the proposed relative to ELN22.

**Supplemental Table 7, cont'd**

**B. EFS, Rule-I.**

| | | Kaplan-Meier | | | mCOXr[a] | | |
|---|---|---|---|---|---|---|---|
| **Type of risk** | **Covariate-level classification for proposed** | **M-iAUC ($25^{th}$,$75^{th}$) over *t*** | **M-cAUC ($25^{th}$,$75^{th}$) over *t*** | **iBrier ($25^{th}$,$75^{th}$) over *t*** | **M-iAUC ($25^{th}$,$75^{th}$) over *t*** | **M-cAUC ($25^{th}$,$75^{th}$) over *t*** | **iBrier ($25^{th}$,$75^{th}$) over *t*** |
| **ELN risk** | | 0.53 [0.51,0.55] | 0.51 [<0.50,0.57] | 0.18 [0.14,0.21] | 0.54 [0.51.58] | 0.56 [0.50,0.62] | 0.18 [0.14,0.22] |
| **Genetic plus phenotypic based risk signatures (RM$_{GP,II-EFS}$)** | (a) Retaining original list assuming fixed | 0.56 [0.53,0.58] (<0.001) | 0.65 [0.58,0.72] (<0.001) | 0.16 [0.14,0.22] | 0.55 [0.53,0.59] (0.001) | 0.61 [0.55,0.69] (<0.001) | 0.18 [0.14,0.22] |
| | (b) Ensemble over *k*-CV | 0.57 [0.56,0.64] (<0.001) | 0.67 [0.59,0.75] (<0.001) | 0.16 [0.14,0.21] | 0.58 [0.55,0.63] (<0.001) | 0.60 [0.54,0.71] (<0.001) | 0.16 [0.14,0.21] |
| | (c) Ensemble over *k*-CV plus missing methods | 0.57 [0.56,0.64] (<0.001) | 0.67 [0.59,0.75] (<0.001) | 0.16 [0.14,0.21] | 0.58 [0.55,0.63] (<0.001) | 0.60 [0.54,0.71] (<0.001) | 0.16 [0.14,0.21] |
| | (d) Independent classification with in each *k*-CV | 0.54 [0.53,0.59] (<0.001) | 0.63 [0.54,0.66] (<0.001) | 0.19 [0.15,0.22] | 0.54 [0.53,0.59] (<0.001) | 0.59 [0.53,0.69] (<0.001) | 0.18 [0.16,0.22] |
| **Genetic based risk signatures (RM$_{G,II-EFS}$)** | (a) Retaining original list assuming fixed | 0.56 [0.54,0.61] (<0.001) | 0.63 [0.60,0.72] (<0.001) | 0.18 [0.14,0.22] | 0.60 [0.55,0.64] (<0.001) | 0.65 [0.58,0.72] (<0.001) | 0.16 [0.15,0.22] |
| | (b) Ensemble over *k*-CV | 0.58 [0.56,0.63] (<0.001) | 0.66 [0.60,0.74] (<0.001) | 0.17 [0.14,0.21] | 0.58 [0.56,0.64] (<0.001) | 0.62 [0.56,0.75] (<0.001) | 0.16 [0.15,0.21] |
| | (c) Ensemble over *k*-CV plus missing methods | 0.58 [0.56,0.63] (<0.001) | 0.66 [0.60,0.74] (<0.001) | 0.17 [0.14,0.21] | 0.58 [0.56,0.64] (<0.001) | 0.62 [0.56,0.75] (<0.001) | 0.16 [0.15,0.21] |
| | (d) Independent classification with in each *k*-CV | 0.55 [0.54,0.57] (<0.001) | 0.61 [0.54,0.67] (<0.001) | 0.19 [0.15,0.22] | 0.57 [0.53,0.58] (<0.001) | 0.58 [0.56,0.67] (<0.001) | 0.19 [0.15,0.22] |

[a]Penalized CoxPH model adjusted by age, gender, race, and either ELN or proposed risk stratification variable (i.e., RM$_{GP,II-EFS}$, RM$_{G,II-EFS}$). [b]Reported in the Figure 5E.

Remarks:

| The higher iAUC or cAUC value (i.e., close to 1) means better numerical performance | The lower iBrier value (i.e., close to 0) means better numerical performance |
|---|---|

Reported in parenthesis are the median of P-values (over 15-fold CV) testing the null hypothesis of inferiority of distributions of survival metrics of the proposed relative to ELN22.

**Supplemental Table 7, cont'd**

**C. EFS, Rule-III.**

| | | Kaplan-Meier | | | mCOXr[a] | | |
|---|---|---|---|---|---|---|---|
| **Type of risk** | **Covariate-level classification for proposed** | **M-iAUC ($25^{th}$,$75^{th}$) over *t*** | **M-cAUC ($25^{th}$,$75^{th}$) over *t*** | **iBrier ($25^{th}$,$75^{th}$) over *t*** | **M-iAUC ($25^{th}$,$75^{th}$) over *t*** | **M-cAUC ($25^{th}$,$75^{th}$) over *t*** | **iBrier ($25^{th}$,$75^{th}$) over *t*** |
| **ELN risk** | | 0.53 [0.51,0.55] | 0.51 [<0.50,0.57] | 0.18 [0.14,0.21] | 0.54 [0.51.58] | 0.56 [0.50,0.62] | 0.18 [0.14,0.22] |
| **Genetic plus phenotypic based risk signatures ($RM_{GP,III-EFS}$)** | (a) Retaining original list assuming fixed | 0.58 [0.53,0.62] (<0.001) | 0.63 [0.60,0.68] (<0.001) | 0.18 [0.15,0.22] | 0.58 [0.53,0.61] (<0.001) | 0.63 [0.55,0.72] (<0.001) | 0.18 [0.15,0.23] |
| | (b) Ensemble over $k$-CV | 0.57 [0.55,0.60] (<0.001) | 0.68 [0.61,0.76] (<0.001) | 0.17 [0.14,0.19] | 0.60 [0.56,0.64] (<0.001) | 0.67 [0.58,0.76] (<0.001) | 0.17 [0.14,0.20] |
| | (c) Ensemble over $k$-CV plus missing methods | 0.58 [0.53,0.62] (<0.001) | 0.63 [0.60,0.68] (<0.001) | 0.18 [0.15,0.22] | 0.58 [0.53,0.61] (<0.001) | 0.63 [0.55,0.72] (<0.001) | 0.18 [0.15,0.23] |
| | (d) Independent classification with in each $k$-CV | 0.55 [0.53,0.58] (<0.001) | 0.57 [0.55,0.65] (<0.001) | 0.19 [0.14,0.23] | 0.56 [0.54,0.58] (0.003) | 0.56 [0.53,0.61] (0.008) | 0.18 [0.15,0.24] |
| **Genetic based risk signatures ($RM_{G,III-EFS}$)** | (a) Retaining original list assuming fixed | 0.58 [0.53,0.62] (<0.001) | 0.63 [0.60,0.68] (<0.001) | 0.18 [0.15,0.22] | 0.58 [0.52,0.61] (<0.001) | 0.61 [0.54,0.72] (<0.001) | 0.18 [0.15,0.23] |
| | (b) Ensemble over $k$-CV | 0.57 [0.55,0.60] (<0.001) | 0.68 [0.61,0.74] (<0.001) | 0.17 [0.14,0.19] | 0.59 [0.55,0.64] (<0.001) | 0.66 [0.58,0.78] (<0.001) | 0.17 [0.14,0.20] |
| | (c) Ensemble over $k$-CV plus missing methods | 0.58 [0.53,0.62] (<0.001) | 0.63 [0.60,0.68] (<0.001) | 0.18 [0.15,0.22] | 0.58 [0.52,0.61] (<0.001) | 0.61 [0.54,0.72] (<0.001) | 0.18 [0.15,0.23] |
| | (d) Independent classification with in each $k$-CV | 0.55 [0.53,0.57] (0.002) | 0.57 [0.55,0.65] (<0.001) | 0.19 [0.14,0.23] | 0.55 [0.54,0.58] (<0.001) | 0.56 [0.53,0.62] (0.008) | 0.18 [0.15,0.24] |

[a]Penalized CoxPH model adjusted by age, gender, race, and either ELN or proposed risk stratification variable (i.e., $RM_{GP,III-EFS}$, $RM_{G,III-EFS}$).

Remarks:

| | |
|---|---|
| The higher iAUC or cAUC value (i.e., close to 1) means better numerical performance | The lower iBrier value (i.e., close to 0) means better numerical performance |

Reported in parenthesis are the median of P-values (over 15-fold CV) testing the null hypothesis of inferiority of distributions of survival metrics of the proposed relative to ELN22.

**Supplemental Table 7, cont'd**

**D. EFS, Rule-IV.**

| Type of risk | Covariate-level classification for proposed | Kaplan-Meier M-iAUC (25th,75th) over *t* | Kaplan-Meier M-cAUC (25th,75th) over *t* | Kaplan-Meier iBrier (25th,75th) over *t* | mCOXr[a] M-iAUC (25th,75th) over *t* | mCOXr[a] M-cAUC (25th,75th) over *t* | mCOXr[a] iBrier (25th,75th) over *t* |
|---|---|---|---|---|---|---|---|
| **ELN risk** | | 0.53 [0.51,0.55] | 0.51 [<0.50,0.57] | 0.18 [0.14,0.21] | 0.54 [0.51.58] | 0.56 [0.50,0.62] | 0.18 [0.14,0.22] |
| **Genetic plus phenotypic based risk signatures ($RM_{GP,IV\text{-}EFS}$)** | (a) Retaining original list assuming fixed | 0.58 [0.54,0.61] (<0.001) | 0.63 [0.60,0.67] (<0.001) | 0.18 [0.15,0.23] | 0.58 [0.53,0.60] (<0.001) | 0.64 [0.52,0.73] (<0.001) | 0.18 [0.15,0.23] |
| | (b) Ensemble over *k*-CV | 0.57 [0.55,0.60] (<0.001) | 0.68 [0.61,0.74] (<0.001) | 0.16 [0.14,0.20] | 0.59 [0.55,0.63] (<0.001) | 0.66 [0.59,0.75] (<0.001) | 0.16 [0.14,0.20] |
| | (c) Ensemble over *k*-CV plus missing methods | 0.58 [0.54,0.61] (<0.001) | 0.63 [0.60,0.68] (<0.001) | 0.18 [0.15,0.23] | 0.58 [0.53,0.60] (<0.001) | 0.64 [0.52,0.73] (<0.001) | 0.18 [0.15,0.23] |
| | (d) Independent classification with in each *k*-CV | 0.55 [0.53,0.58] (0.001) | 0.59 [0.56,0.65] (<0.001) | 0.18 [0.15,0.22] | 0.58 [0.53,0.58] (0.003) | 0.56 [0.53,0.67] (<0.001) | 0.18 [0.15,0.23] |
| **Genetic based risk signatures ($RM_{G,IV\text{-}EFS}$)** | (a) Retaining original list assuming fixed | 0.58 [0.54,0.61] (<0.001) | 0.63 [0.60,0.68] (<0.001) | 0.18 [0.15,0.23] | 0.58 [0.53,0.60] (<0.001) | 0.64 [0.52,0.73] (<0.001) | 0.18 [0.15,0.23] |
| | (b) Ensemble over *k*-CV | 0.57 [0.55,0.60] (<0.001) | 0.68 [0.61,0.74] (<0.001) | 0.16 [0.14,0.20] | 0.59 [0.55,0.63] (<0.001) | 0.66 [0.59,0.75] (<0.001) | 0.16 [0.14,0.20] |
| | (c) Ensemble over *k*-CV plus missing methods | 0.58 [0.54,0.61] (<0.001) | 0.63 [0.60,0.68] (<0.001) | 0.18 [0.15,0.23] | 0.58 [0.53,0.60] (<0.001) | 0.64 [0.52,0.73] (<0.001) | 0.18 [0.15,0.23] |
| | (d) Independent classification with in each *k*-CV | 0.55 [0.53,0.58] (0.001) | 0.59 [0.56,0.65] (<0.001) | 0.18 [0.15,0.22] | 0.58 [0.53,0.58] (0.001) | 0.56 [0.53,0.67] (<0.001) | 0.18 [0.15,0.23] |

[a]Penalized CoxPH model adjusted by age, gender, race, and either ELN or proposed risk stratification variable (i.e., $RM_{GP,IV\text{-}EFS}$, $RM_{G,IV\text{-}EFS}$).

Remarks:

| | |
|---|---|
| The higher iAUC or cAUC value (i.e., close to 1) means better numerical performance | The lower iBrier value (i.e., close to 0) means better numerical performance |

Reported in parenthesis are the median of P-values (over 15-fold CV) testing the null hypothesis of inferiority of distributions of survival metrics of the proposed relative to ELN22.

### SUPPLEMENTAL FIGURE LEGENDS

**Supplemental Figure 1. Covariate-level classifications by counterfactual risk profiles for genetic and phenotypic features.** A) “strongly” or “moderately” Adverse features based on overall survival (OS); B) “strongly” or “moderately” Favorable features based on OS; C) “strongly” or “moderately” Adverse features based on event free survival (EFS); D) “strongly” or “moderately” Favorable features based on EFS. Bootstrap based 95% confidence intervals and *P* values testing superiority were provided. *Green* and *dark-green* color correspond to the control group (i.e., (-ve) or wild-type) population; *orange* and *dark-orange* correspond to the genetically or phenotypically abnormal population (i.e., (+ve) or mutated). Curves with dark-green and dark-orange color correspond to the median of bootstrap estimates of 2,000 runs. Note that the curves with light and dark green color are very close to one another and thus visually not discernible.

**Supplemental Figure 2. Overall survival (OS) based on the genetic feature specific risk model using the covariate-level classification Rule-I ($RM_{G,I}$).** A) Definitions of features associated with the $RM_{G,I}$ AML risk groups. The category column includes color legends for reference; B) Application of the $RM_{G,I}$ to the CU ven/aza treated cohort after excluding allo-HCT patients for OS (left panel) and best response (BR) (right panel); C) Application of the $RM_{G,I}$ to the ven/aza treated CU cohort based on the full analytical set (FAS) for OS (left panel) and BR (right panel); D) $RM_{G,I}$ based on OS treating allo-HCT recipients as censored (left panel) and only within the cohort comprised of allo-HCT recipients (right panel). Throughout Supplemental Figures, BCOR, DNMT3A, DDX41, SF3B1, KIT and similar designations refer to NGS mutations. IDH1, IDH2, NPM1 are composite mutations defined by different testing technologies as described in Methods and Supplemental Table 1E. “p53m” throughout is a combination of p53 abnormalities by NGS bundled with FISH losses or gains of 17p. Chr 7 abn:loss, Chr 8 abn:loss, DEK:loss, MECOM:gain, RARA:loss and similar designations refer to FISH features. Terms including “ x risk cytogenetics”, “minusx” and “Delx” refer to karyotypic features.

**Supplemental Figure 3. Overall survival (OS) based on the genetic features specific risk model using the covariate-level classification Rule-III ($RM_{G,III}$).** A) Definitions of features associated with the $RM_{G,III}$ AML risk groups. The category column includes color legends for reference; B) Application of the $RM_{G,III}$ to the CU ven/aza treated cohort after excluding allo-HCT patients for OS (left panel) and best response (BR) (right panel); C) Application of the $RM_{G,III}$ to the ven/aza treated CU cohort based on the full analytical set (FAS) for OS (left panel) and BR (right panel); D) $RM_{G,III}$ based on OS treating allo-HCT recipients as censored (left panel) and only within the cohort comprised of allo-HCT recipients (right panel).

**Supplemental Figure 4. Overall survival (OS) based on the genetic features specific risk model using the covariate-level classification Rule-IV ($RM_{G,IV}$).** A) Definitions of features associated with the $RM_{G,IV}$ AML risk groups. The category column includes color legends for reference; B) Application of the $RM_{G,IV}$ to the CU ven/aza treated cohort after excluding allo-HCT patients for OS (left panel) and best response (BR) (right panel); and C) Application of the $RM_{G,IV}$ to the ven/aza treated CU cohort based on the full analytical set (FAS) for OS (left panel) and BR (right panel); D) $RM_{G,IV}$ based on OS treating allo-HCT recipients as censored (left panel) and only within the cohort comprised of allo-HCT recipients (right panel).

**Supplemental Figure 5. ELN22 risk categorization model applied to overall survival (OS) and best response (BR).** A) OS and BR with allo-HCT recipients excluded; B) OS and BR based on the full analytical set (FAS); C) Pairwise comparison between the $RM_{G,II}$ and ELN risk models for the Adverse, Intermediate, and Favorable risk groups in the cohort excluding allo-HCT patients; D) Comparison of inter-risk group conformity between the $RM_{G,II}$ and ELN22 based on the FAS; E) Comparison of inter-risk group conformity between the $RM_{G,II}$ and ELN22 based on the FAS; F) Predictability of OS based on the $RM_{G,II}$ and ELN22 over follow-up time up to four years. Note BCOR, DNMT3A, DDX41, SF3B1, KRAS, KMT2A, PTPN11, SMC1A, JAK2 are mutations defined by NGS. IDH1, IDH2, NPM1 and p53m are defined by composite molecular tests as defined in Methods and Supplemental Table 1E for "composite mutation features". Chr 7 abn:loss, Chr 8 abn:loss, DEK:loss, MECOM:gain, RARA:loss are FISH features. The remainder including Del7q, T(9;11), Minus5, Minus17, Complex cytogenetics are karyotype features.

**Supplemental Figure 6. Overall survival (OS) based on the genetic-plus-phenotype features specific risk model using the covariate-level classification Rule-I ($RM_{GP,I}$).** A) Definitions of features associated with the $RM_{GP,I}$ AML risk groups. The category column includes color legends for reference; B) Application of the $RM_{GP,I}$ to the ven/aza treated cohort after excluding allo-HCT patients for OS (left panel) and best response (BR) (right panel); C) Application of the $RM_{GP,I}$ to the ven/aza treated cohort based on the full analytical set (FAS) for OS (left panel) and BR (right panel); D) $RM_{GP,I}$ based on OS treating allo-HCT recipients as censored (left panel) and only within the cohort comprised of allo-HCT recipients (right panel).

**Supplemental Figure 7. Overall survival (OS) based on the genetic-plus-phenotypic features specific risk model using the covariate-level classification Rule-II ($RM_{GP,II}$).** A) Definitions of features associated with the $RM_{GP,II}$ AML risk groups. The category column includes color legends for reference; B) Application of the $RM_{GP,II}$ model to the ven/aza treated cohort after excluding allo-HCT patients for OS (left panel) and best response (BR) (right panel); C) Application of the $RM_{GP,II}$ model to the ven/aza treated cohort based on the full analytical set (FAS) for OS (left panel) BR (right panel); D) $RM_{GP,II}$ based on OS treating allo-HCT recipients as censored (left panel) and only within the cohort comprised of allo-HCT recipients (right panel).

**Supplemental Figure 8. Overall survival (OS) based on the genetic-plus-phenotypic features specific risk model using the covariate-level classification Rule-III ($RM_{GP,III}$).** A) Definitions of features associated with the $RM_{GP,III}$ AML risk groups. The category column includes color legends for reference; B) Application of the $RM_{GP,III}$ model to the ven/aza treated cohort after excluding allo-HCT patients for OS (left panel) and best response (right panel); C) Application of the $RM_{GP,III}$ model to the ven/aza treated cohort based on the full analytical set (FAS) for OS (left panel) and best response (right panel); D) $RM_{GP,III}$ based on OS treating allo-HCT recipients as censored (left panel) and only within the cohort comprised of allo-HCT recipients (right panel).

**Supplemental Figure 9. Overall survival (OS) based on the genetic-plus-phenotypic features specific risk model using the covariate-level classification Rule-IV ($RM_{GP,IV}$).** A) Definitions of features associated with the $RM_{GP,IV}$ AML risk groups. The category column includes color legends for reference; B) Application of the $RM_{GP,IV}$ to the ven/aza treated cohort after excluding allo-HCT patients for OS (left panel) and best response (BR) (right panel); C) Application of the $RM_{GP,IV}$ model to the ven/aza treated cohort based on the full analytical set (FAS) for OS (left panel) and BR (right panel); D) $RM_{GP,IV}$ based on OS treating allo-HCT recipients as censored (left panel) and only within the cohort comprised of allo-HCT recipients (right panel).

**Supplemental Figure 10. Event free survival (EFS) based on the genetic features specific risk model using the covariate-level classification Rule-I ($RM_{G,I\text{-}EFS}$).** A) Definitions of features associated with the $RM_{G,I\text{-}EFS}$ AML risk groups. The category column includes color legends for reference; B) Application of the $RM_{G,I\text{-}EFS}$ to the full analytical set (FAS) (left panel) and the set excluding (right panel) allo-HCT recipients; C) $RM_{G,I\text{-}EFS}$ based on OS treating allo-HCT recipients as censored (left panel) and only within the cohort comprised of allo-HCT recipients (right panel).

**Supplemental Figure 11. Event free survival (EFS) based on the genetic features specific risk model using the covariate-level classification Rule-III ($RM_{G,III\text{-}EFS}$).** A) Definitions of features associated with the $RM_{G,III\text{-}EFS}$ AML risk groups. The category column includes color legends for reference; B) Application of the $RM_{G,III\text{-}EFS}$ to the full analytical set (FAS) (left panel) and the set excluding (right panel) allo-HCT recipients; C) $RM_{G,III\text{-}EFS}$ based on OS treating allo-HCT recipients as censored (left panel) and only within the cohort comprised of allo-HCT recipients (right panel).

**Supplemental Figure 12. Event free survival (EFS) based on the genetic features specific risk model using the covariate-level classification Rule-IV ($RM_{G,IV\text{-}EFS}$).** A) Definitions of features associated with the $RM_{G,IV\text{-}EFS}$ AML risk groups. The category column includes color legends for reference; B) Application of the $RM_{G,IV\text{-}EFS}$ to the full analytical set (FAS) (left panel) and the set excluding (right panel) allo-HCT recipients; C) $RM_{G,IV\text{-}EFS}$ based on OS treating allo-HCT recipients as censored (left panel) and only within the cohort comprised of allo-HCT recipients (right panel).

**Supplemental Figure 13. ELN22 and comparison with respect to $RM_{G,II\text{-}EFS}$ for event free survival (EFS).** A) EFS with allo-HCT recipients excluded; B) EFS based on the full analytical set (FAS); C) Pairwise comparison between the $RM_{G,II\text{-}EFS}$ and ELN risk models for the Adverse, Intermediate, and Favorable risk groups excluding allo-HCT patients; D) Comparison of inter-risk group conformity between the $RM_{G,II\text{-}EFS}$ and ELN22 risk model based on the FAS; E)

Predictability of OS based on the $RM_{G,II-EFS}$ and ELN22 over follow-up time up to four years. Note BCOR, KRAS, KMT2A, PTPN11, JAK2, were mutations defined by NGS. IDH1, IDH2, NPM1 and p53m are defined by composite molecular tests as defined in Methods and Supplemental Table 1E for “composite mutation features”, Chr 7 abn:loss, Chr 8 abn:loss, RARA:loss, MECOM:rearrangement, KMT2A:rearrangement are FISH features. The remainder including Del 5q Del7q, T(9;11), Minus5, Minus17, Complex cytogenetics, Indeterminate cytogenetics and Normal cytogenetics are karyotype features.

**Supplemental Figure 14. Event free survival (EFS) based on the genetic-plus-phenotypic features specific based risk model using the covariate-level classification Rule-I ($RM_{GP,I-EFS}$).** A) Definitions of features associated with the $RM_{GP,I-EFS}$ AML risk groups. The category column includes color legends for reference; B) Application of the $RM_{GP,I-EFS}$ to the full analytical set (FAS) (left panel) and the set excluding (right panel) allo-HCT recipients; C) $RM_{GP,I-EFS}$ based on OS treating allo-HCT recipients as censored (left panel) and only within the cohort comprised of allo-HCT recipients (right panel).

**Supplemental Figure 15. Event free survival (EFS) based on the genetic-plus-phenotypic features specific risk model using the covariate-level classification Rule-II ($RM_{GP,II-EFS}$).** A) Definitions of features associated with the $RM_{GP,II-EFS}$ AML risk groups. The category column includes color legends for reference; B) Application of the $RM_{GP,II-EFS}$ to the full analytical set (FAS) (left panel) and the set excluding (right panel) allo-HCT recipients; C) $RM_{GP,II-EFS}$ based on OS treating allo-HCT recipients as censored (left panel) and only within the cohort comprised of allo-HCT recipients (right panel).

**Supplemental Figure 16. Event free survival (EFS) based on the genetic-plus-phenotypic features specific risk model using the covariate-level classification Rule-III ($RM_{GP,III-EFS}$).** A)

Definitions of features associated with the $RM_{GP,III\text{-}EFS}$ AML risk groups. The category column includes color legends for reference; B) Application of the $RM_{GP,III\text{-}EFS}$ to the full analytical set (FAS) (left panel) and the set excluding (right panel) allo-HCT recipients; C) $RM_{GP,III\text{-}EFS}$ based on OS treating allo-HCT recipients as censored (left panel) and only within the cohort comprised of allo-HCT recipients (right panel).

**Supplemental Figure 17. Event free survival (EFS) based on the genetic-plus-phenotypic features specific risk model using the covariate-level classification Rule-IV ($RM_{GP,IV\text{-}EFS}$).** A) Definitions of features associated with the $RM_{GP,IV\text{-}EFS}$ AML risk groups. The category column includes color legends for reference; B) Application of the $RM_{GP,IV\text{-}EFS}$ to the full analytical set (FAS) (left panel) and the set excluding (right panel) allo-HCT recipients; C) $RM_{GP,IV\text{-}EFS}$ based on OS treating allo-HCT recipients as censored (left panel) and only within the cohort comprised of allo-HCT recipients (right panel).

**Supplemental Figure 18. Subject-level risk stratifications of 316 AML patients by different risk models (*as highlighted in Supplemental Table 4).*** Covariate-level classifications were performed by different rules and assumptions - A) Missing data adjusted as a separate category and allo-HCT recipients were not censored; B) Complete cases with inverse probability weights and allo-HCT recipients were not censored; C) Imputation of missing elements by MICE and allo-HCT recipients were not censored; D) Missing data adjusted as a separate category and allo-HCT recipients were censored; E) Complete cases with inverse probability weights and allo-HCT recipients were censored; F) Imputation of missing elements by MICE and allo-HCT recipients were censored.

**Supplemental Figure 19. Evaluation of the ELN22 for overall survival (OS) using the RWC dataset.** Reported are the results for OS including allo-HCT patients (left panel) and excluding allo-HCT patients (right panel).

**Supplemental Figure 20. Contingency tables for risk models.** Reported are the (top) confusion matrices with the risk groups based on the RWC-$RM_{G\text{-}II}$ and ELN22 compared to their recorded outcomes (0 = censored to last known alive date (LKAD) or 1 = deceased) and (bottom) a contingency table with dot plots highlighting the proportion of deceased patients for different risk groups across the RWC-$RM_{G\text{-}II}$ and ELN22.

**Supplemental Figure 21. Contingency tables for risk models at 450-day.** Reported are the (top) confusion matrices with the risk groups based on the RWC-$RM_{G\text{-}II}$ and ELN22 compared to their observed outcomes (0 = alive at 450-day or 1 = deceased before 450-day post-treatment) and (bottom) a contingency table with dot plots highlighting the proportion of deceased patients for different risk groups across the RWC-$RM_{G\text{-}II}$ and ELN22.

**Supplemental Figure 22. Evaluation of the proposed risk models (RMs) for overall survival (OS) using the RWC dataset.** Results are based on OS including allo-HCT patients (left panel) and excluding allo-HCT patients (right panel) with respect to the simplified versions of RMs. A) Application of the RWC-$RM_{G,I}$ to the full analytical set (FAS) (left panel) and the set excluding (right panel) allo-HCT recipients; B) Application of the RWC-$RM_{G,III}$ to the full analytical set (FAS) (left panel) and the set excluding (right panel) allo-HCT recipients; C) Application of the RWC-$RM_{G,IV}$ to the full analytical set (FAS) (left panel) and the set excluding (right panel) allo-HCT recipients; D) Application of the RWC-$RM_{GP,I}$ to the full analytical set (FAS) (left panel) and the set excluding (right panel) allo-HCT recipients; E) Application of the RWC-$RM_{GP,II}$ to the full analytical set (FAS) (left panel) and the set excluding (right panel) allo-HCT recipients; F) Application of the RWC-$RM_{GP,III}$ to the full analytical set (FAS) (left panel) and the set excluding (right panel) allo-HCT recipients; G) Application of the RWC-$RM_{GP,IV}$ to the full analytical set (FAS) (left panel) and the set excluding (right panel) allo-HCT recipients

**Supplemental Figure 23. Conformity between risk groups assigned by the overall survival (OS) specific risk models and ELN22 based on the RWC.** Results are based on A) the full analytical set (FAS) with *Imputation-by-mode* and B) an imputed sample with respect to MICE. Patients with missing ELN22 were shown as “white” vertical lines.

**Supplemental Figure 24. Evaluation of predictive performance based on the RWC.** Results are based on the (top) FAS with missing data being imputed by *imputation-by-mode* approach and (bottom) average over 10 imputed analytical sets. Results reported in the main draft are highlighted in red box.

**Supplemental Figure 25. Sensitivity analysis for the evaluation of predictive performance based on the RWC.** Penalized CoxPH model is trained on the CU dataset adjusting for age, gender, and risk classification variable using either ELN22 or RWC-RM. A) Counts of patients assigned to each risk group when the reduced RWC feature definitions (i.e., RWC-RMs) were applied to the CU cohort; B) Time-dependent AUC values are based on the FAS where missing data elements being imputed by *imputation-by-mode* approach; C) Time-dependent AUC values are based on the FAS where missing data elements being imputed by MICE approach.

**Supplemental Figure 1A. Counterfactual risk profiles for genetic and phenotypic markers classified as "strongly" or "moderate" adverse for overall survival.** Note that the curves with light and dark green color are very close to one another and thus visually not discernible.

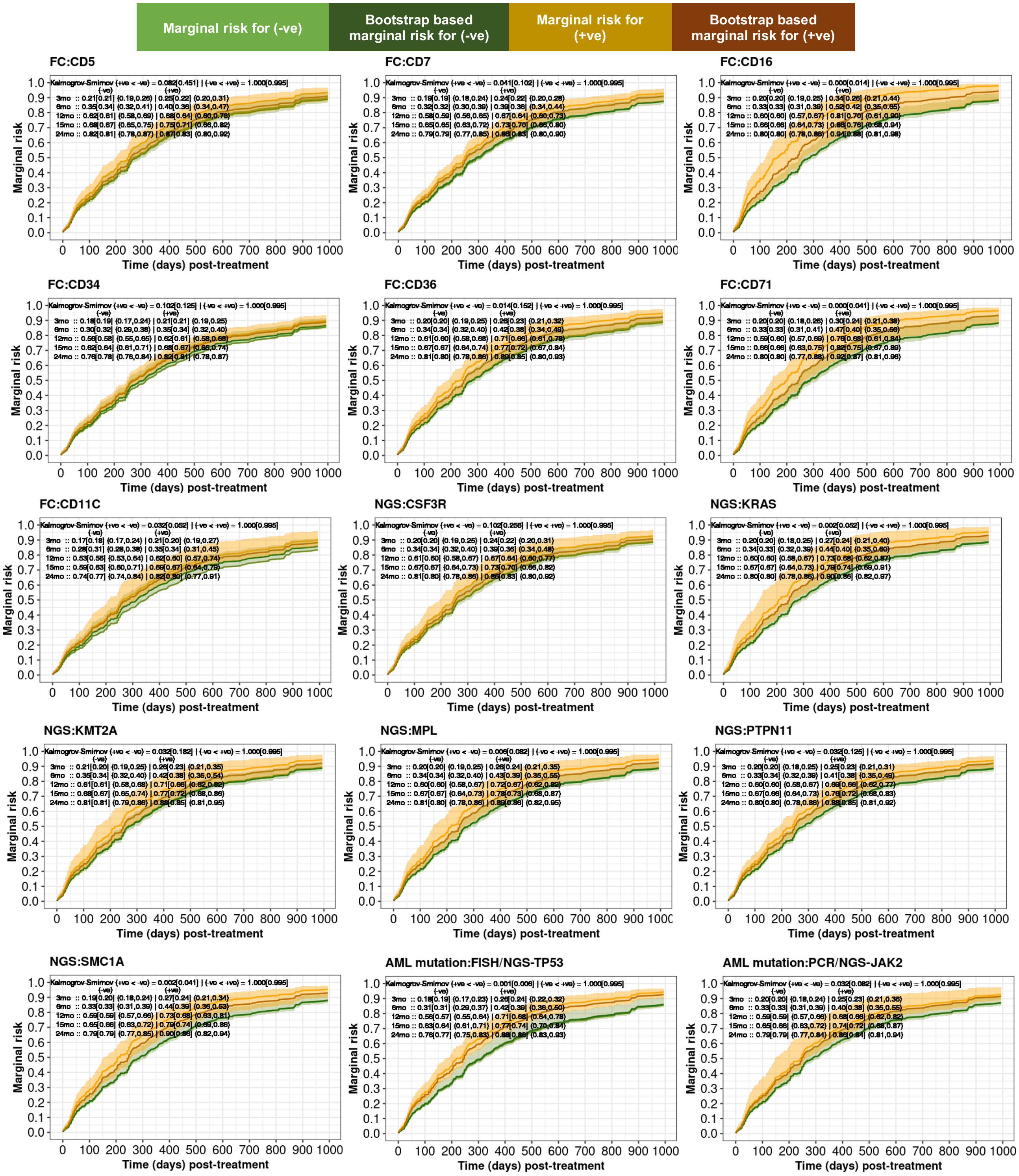

### Supplemental Figure 1A, cont'd

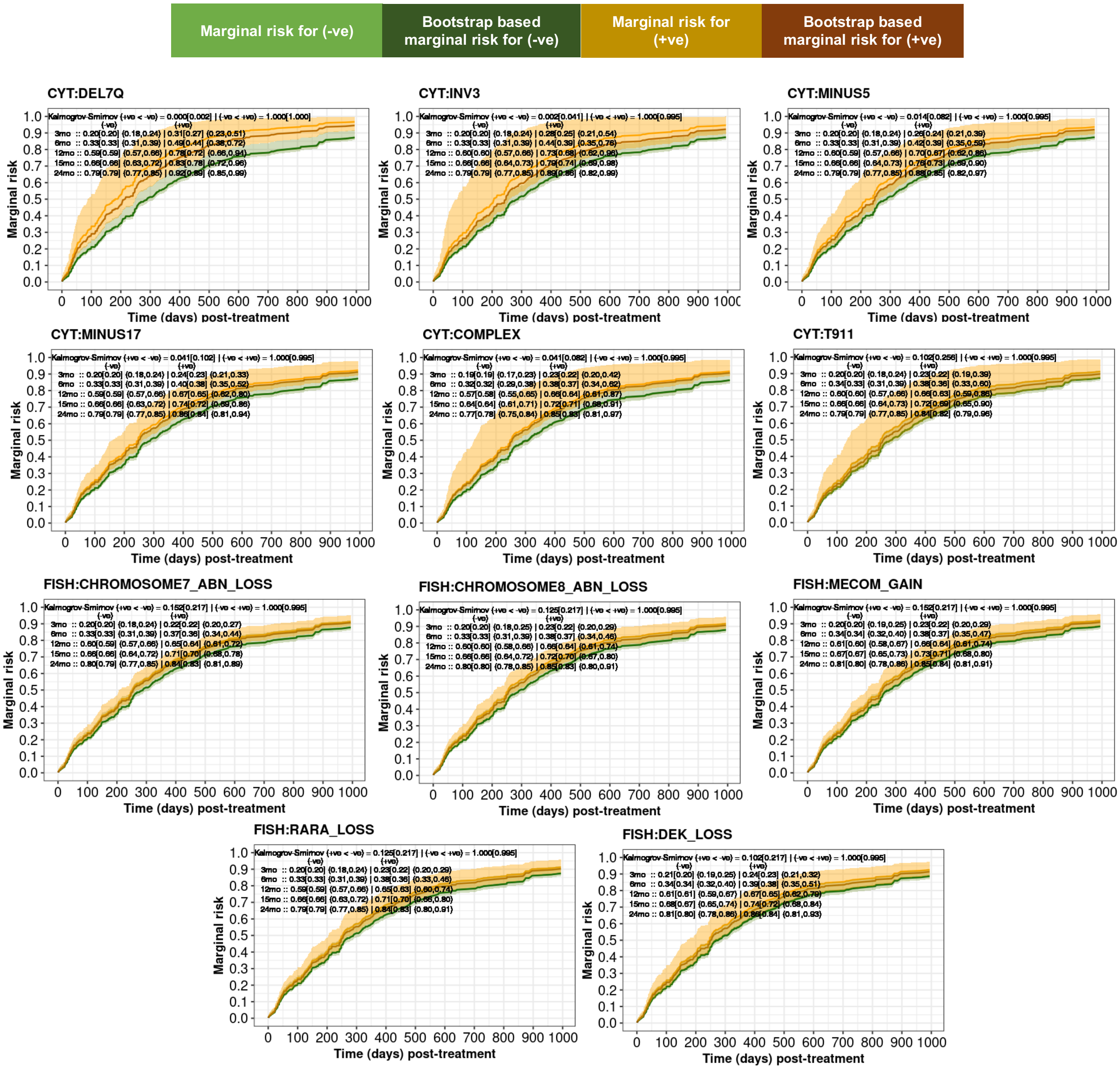

**Supplemental Figure 1B. Counterfactual risk profiles for genetic and phenotypic markers classified as “strongly” or “moderate” favorable for overall survival.** Note that the curves with light and dark green color are very close to one another and thus visually not discernible.

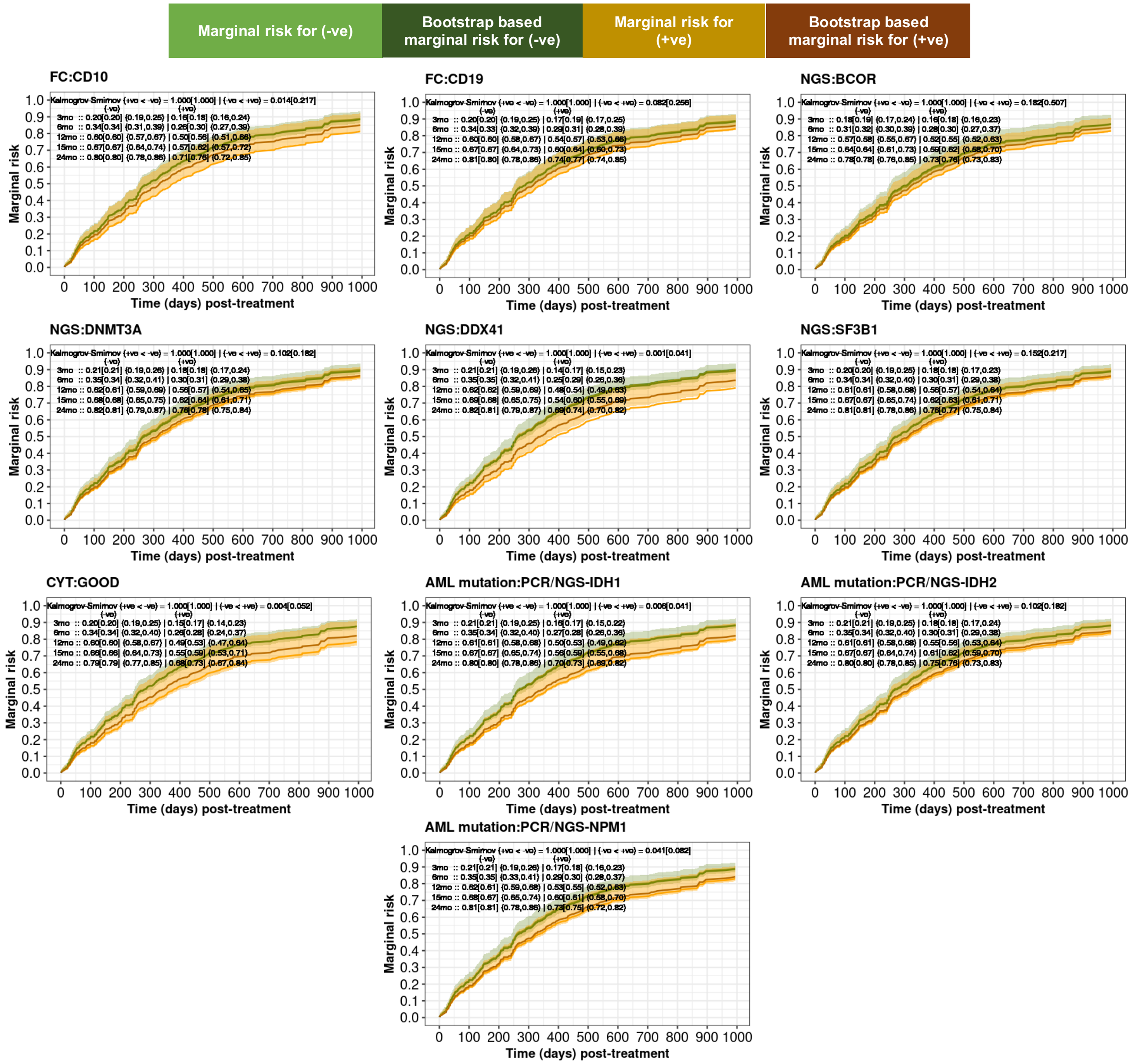

**Supplemental Figure 1C. Counterfactual risk profiles for genetic and phenotypic markers classified as "strongly" or "moderate" adverse for event free survival.** Note that the curves with light and dark green color are very close to one another and thus visually not discernible.

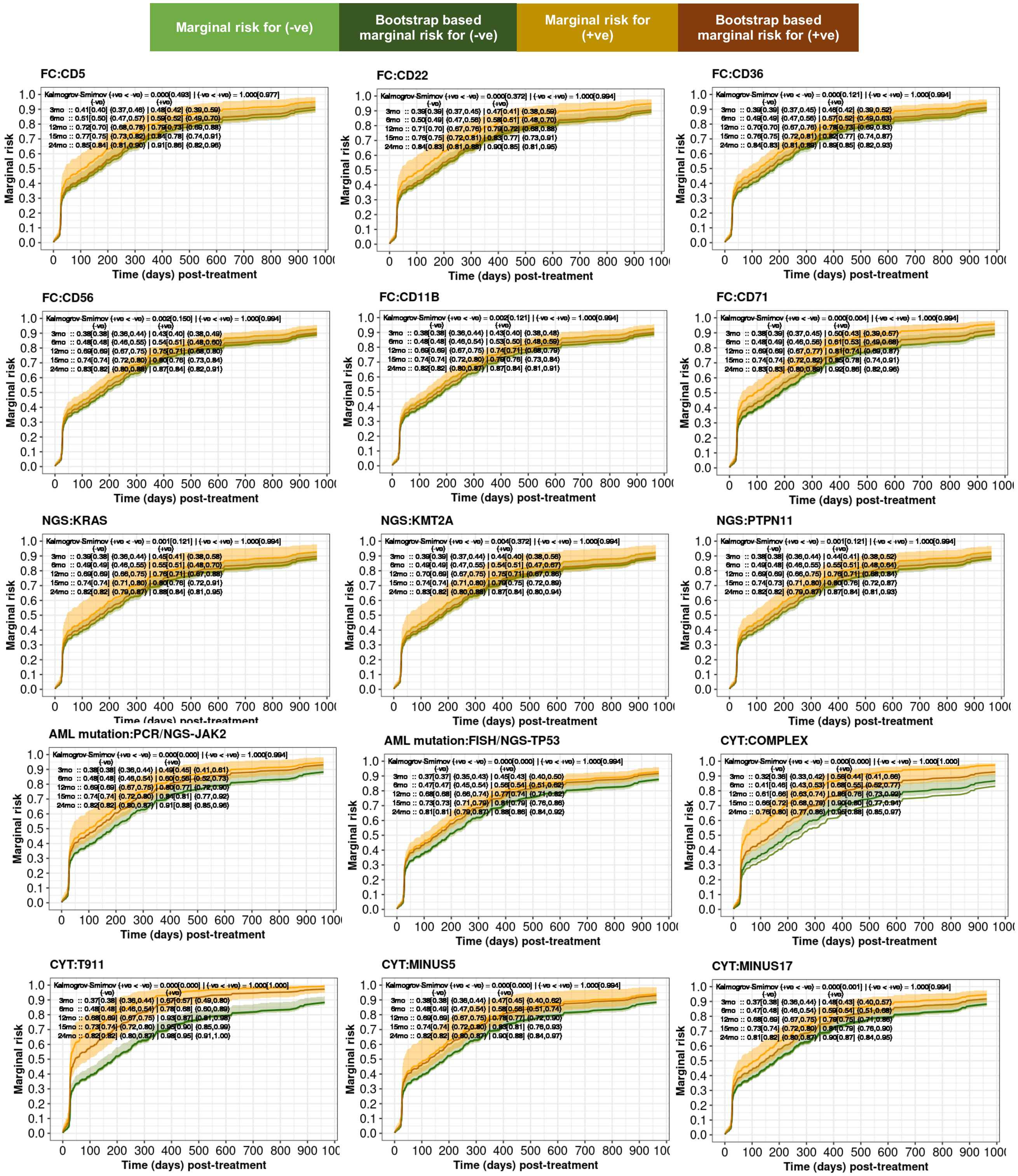

### Supplemental Figure 1C, cont'd

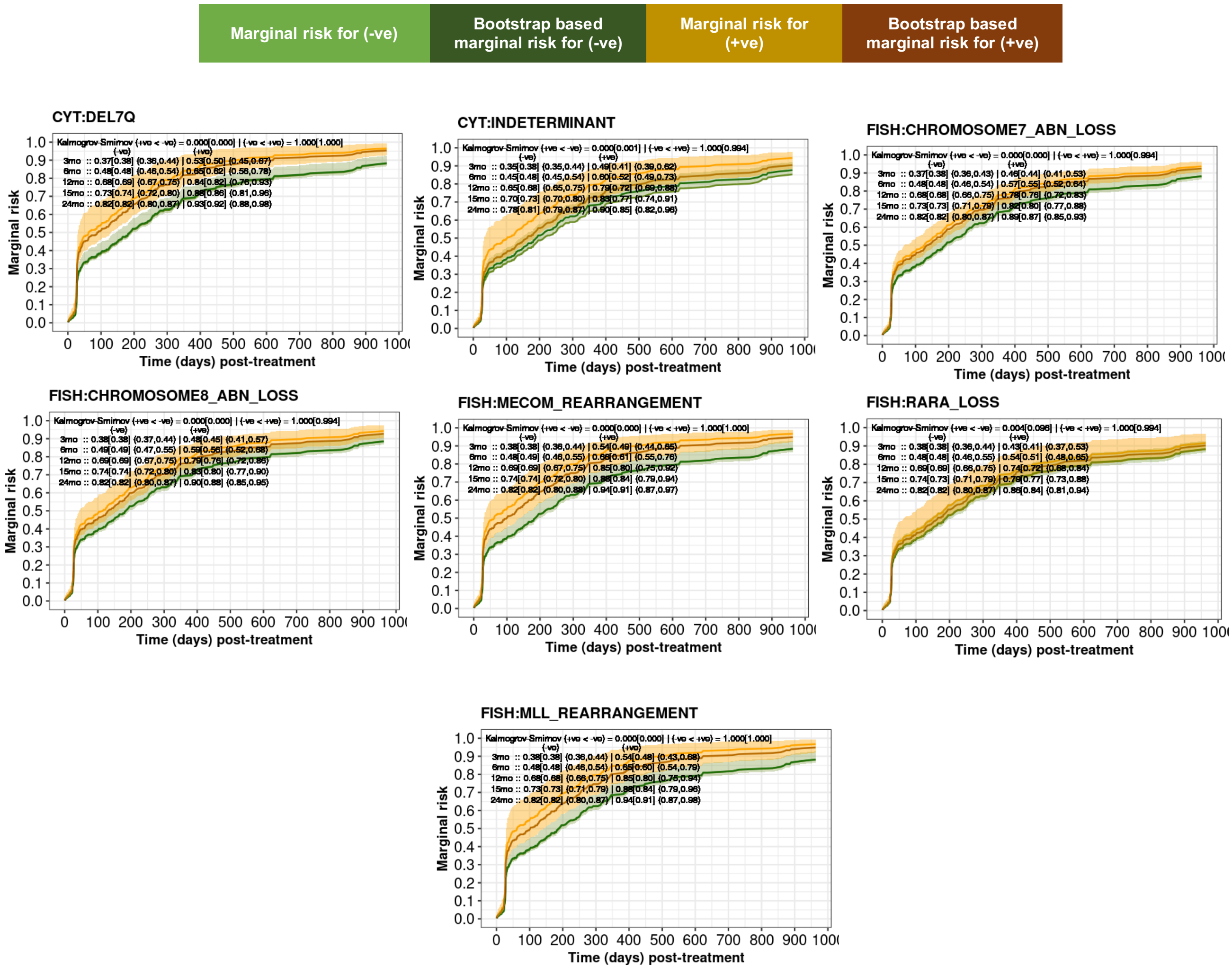

**Supplemental Figure 1D. Counterfactual risk profiles for genetic and phenotypic markers classified as "strongly" or "moderate" favorable for event free survival.** Note that the curves with light and dark green color are very close to one another and thus visually not discernible.

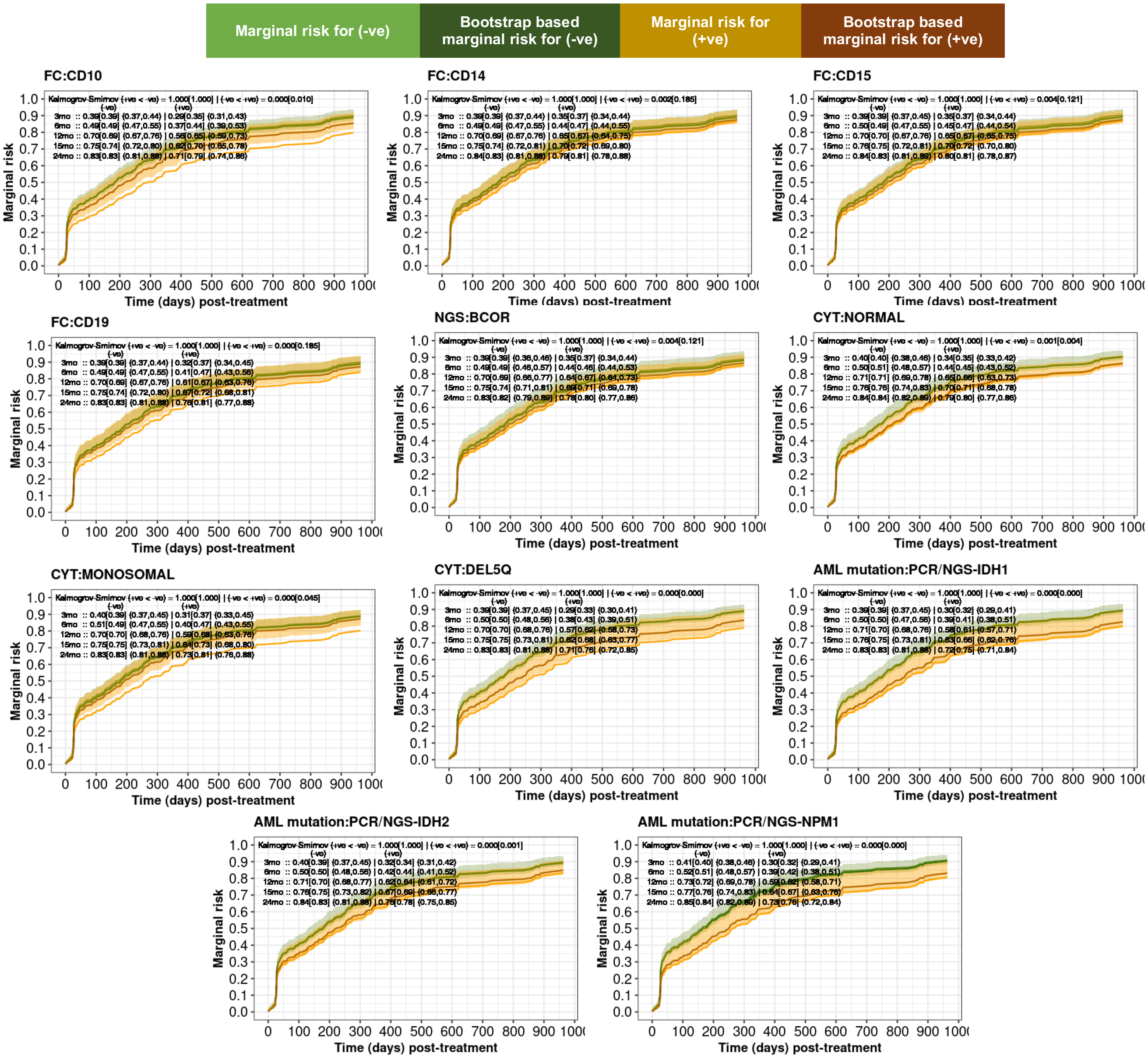

**Supplemental Figure 2. Overall survival (OS) based on the genetic features specific risk model using Rule-I ($RM_{G,I}$).**

**A. Feature identification strategy and $RM_{G,I}$ specific features.**

| Category | Genetic features | Frequency† | Frequency¶ |
|---|---|---|---|
| **Favorable** | Any of ***BCOR, DNMT3A, DDX41, SF3B1, KIT, Good risk cytogenetics, IDH1, IDH2, NPM1*** (in the absence of any Adverse features) | **74/224 (33%)** | **103/316 (33%)** |
| **Intermediate** | No Favorable or Adverse features, or ≥1 Favorable features with ≥1 Adverse features | **67/224 (30%)** | **105/316 (33%)** |
| **Adverse** | Any of ***CSF3R, KRAS, KMT2A, MPL, PTPN11, SMC1A, JAK2, p53m, Del7q, Inv(3), t(9;11), Minus5, Minus17, Complex cytogenetics, Chr 7 abn:loss, Chr 8 abn:loss, DEK:loss, MECOM:gain, RARA:loss*** (in the absence of any Favorable features) | **83/224 (37%)** | **108/316 (34%)** |

† Counts (proportions) of patients after excluding allo-HCT
¶ Counts (proportions) of patients without excluding allo-HCT

**Supplemental Figure 2, cont'd**

**B. $RM_{G,I}$ on OS (left) and best response (right), excluding allo-HCT patients.**

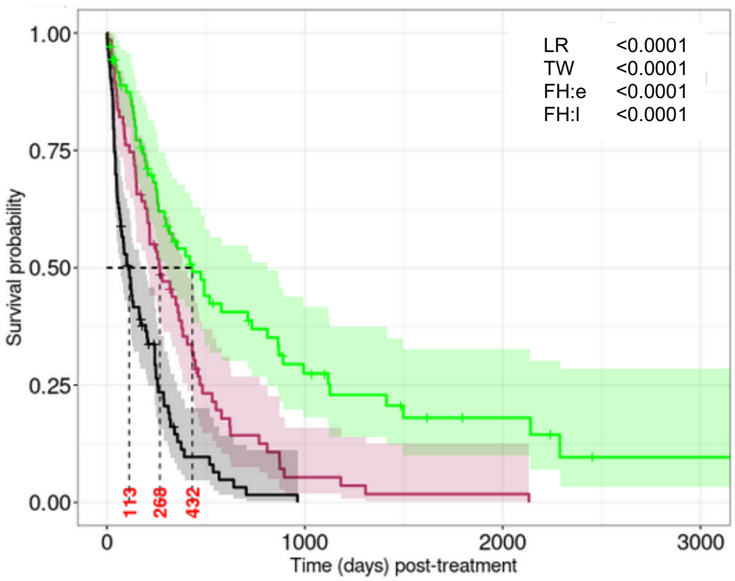


**Number at risk (number censored)**

| | 0 | 1000 | 2000 | 3000 |
|---|---|---|---|---|
| green | 74 (0) | 14 (14) | 5 (19) | 1 (21) |
| red | 67 (0) | 3 (5) | 1 (5) | 0 (5) |
| black | 83 (0) | 0 (7) | 0 (7) | 0 (7) |

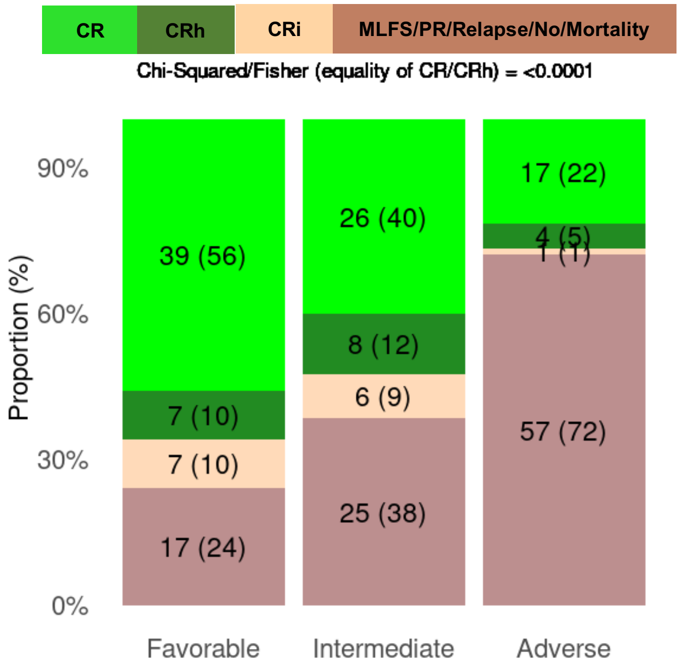


**C. $RM_{G,I}$ on OS (left) and best response (right), with respect to the FAS.**

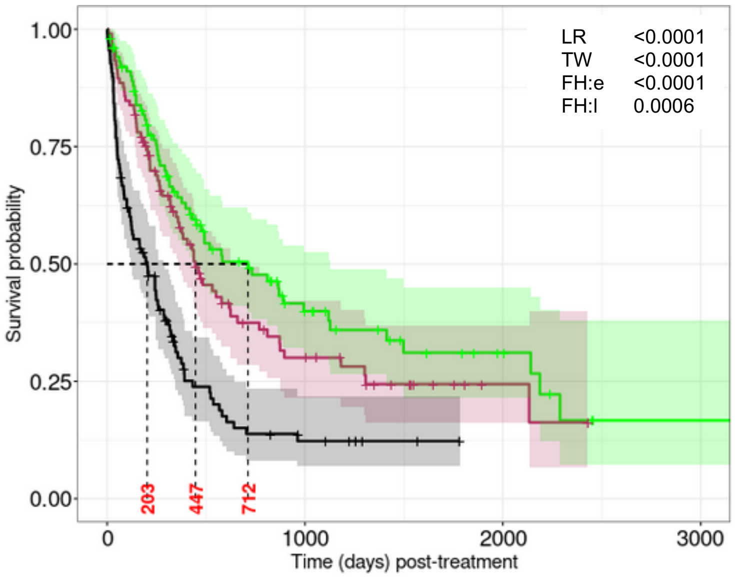


**Number at risk (number censored)**

| | 0 | 1000 | 2000 | 3000 |
|---|---|---|---|---|
| green | 103 (0) | 24 (27) | 8 (39) | 1 (43) |
| red | 105 (0) | 20 (22) | 3 (36) | 0 (38) |
| black | 108 (0) | 8 (14) | 0 (22) | 0 (22) |

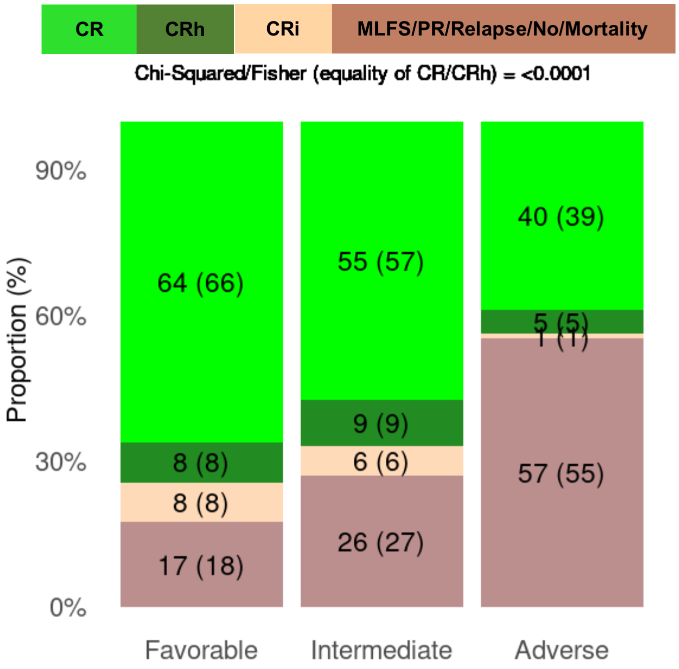

**Supplemental Figure 2, cont'd**

**D. $RM_{G,I}$ on OS censoring allo-HCT (left) and only within allo-HCT (right) recipients.**

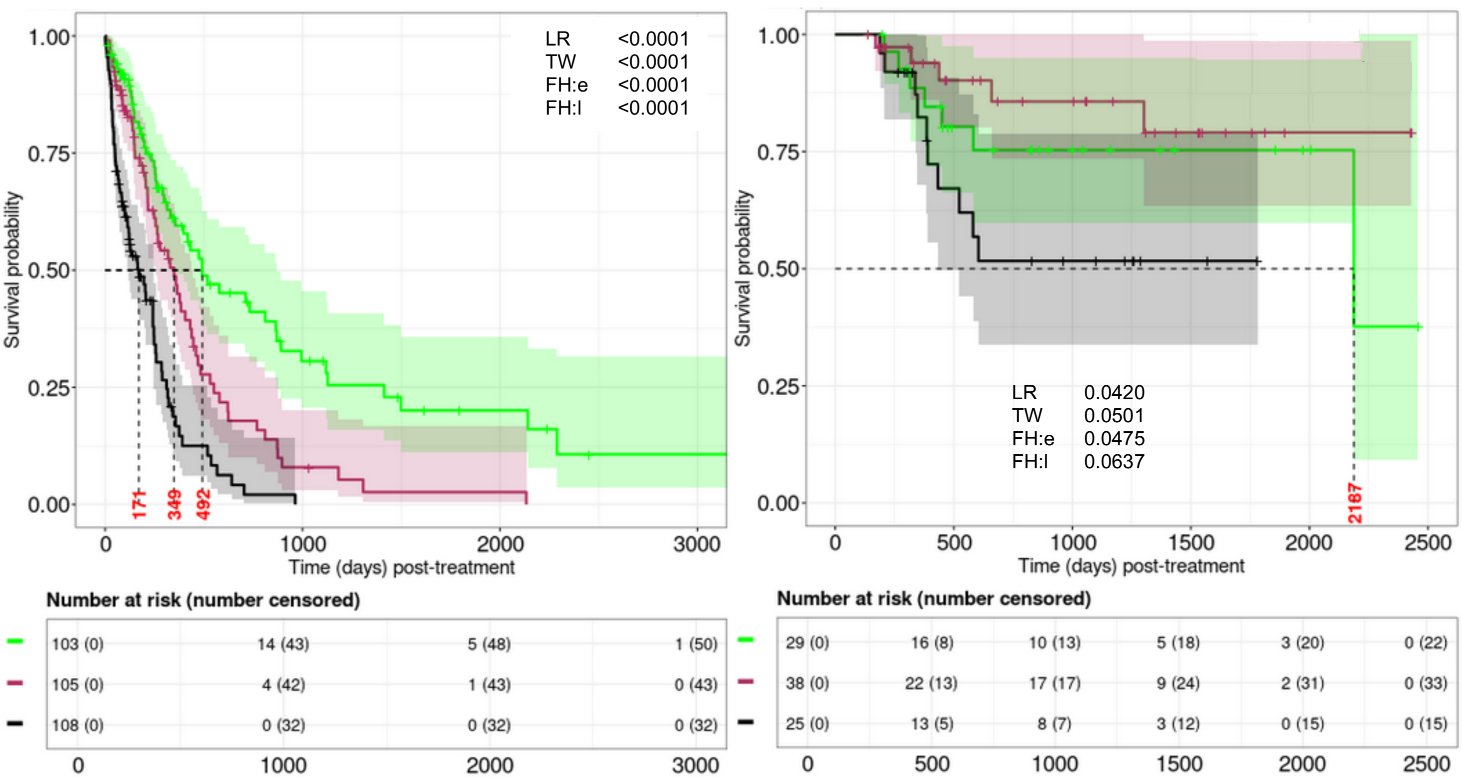

**Supplemental Figure 3. Overall survival (OS) based on the genetic features specific risk model using Rule-III ($RM_{G,III}$).**

**A. Feature identification strategy and $RM_{G,III}$ features.**

| Category | Genetic features | Frequency† | Frequency¶ |
|---|---|---|---|
| **Favorable** | Any of ***DDX41, Cytogenetics good, IDH1, NPM1*** (in the absence of any Adverse features) | **48/224 (21%)** | **69/316 (22%)** |
| **Intermediate** | No Favorable or Adverse features, or $\geq$1 Favorable features with $\geq$1 Adverse features | **78/224 (35%)** | **111/316 (35%)** |
| **Adverse** | Any of ***KRAS, KMT2A, MPL, PTPN11, SMC1A, JAK2, p53m, Inv(3), Del7q, Minus5, Minus17, Complex cytogenetics*** (in the absence of any Favorable features) | **98/224 (44%)** | **136/316 (43%)** |

† Counts (proportions) of patients after excluding allo-HCT
¶ Counts (proportions) of patients without excluding allo-HCT

**Supplemental Figure 3, cont'd**

**B. $RM_{G,III}$ on OS (left) and best response (right), excluding allo-HCT recipients.**

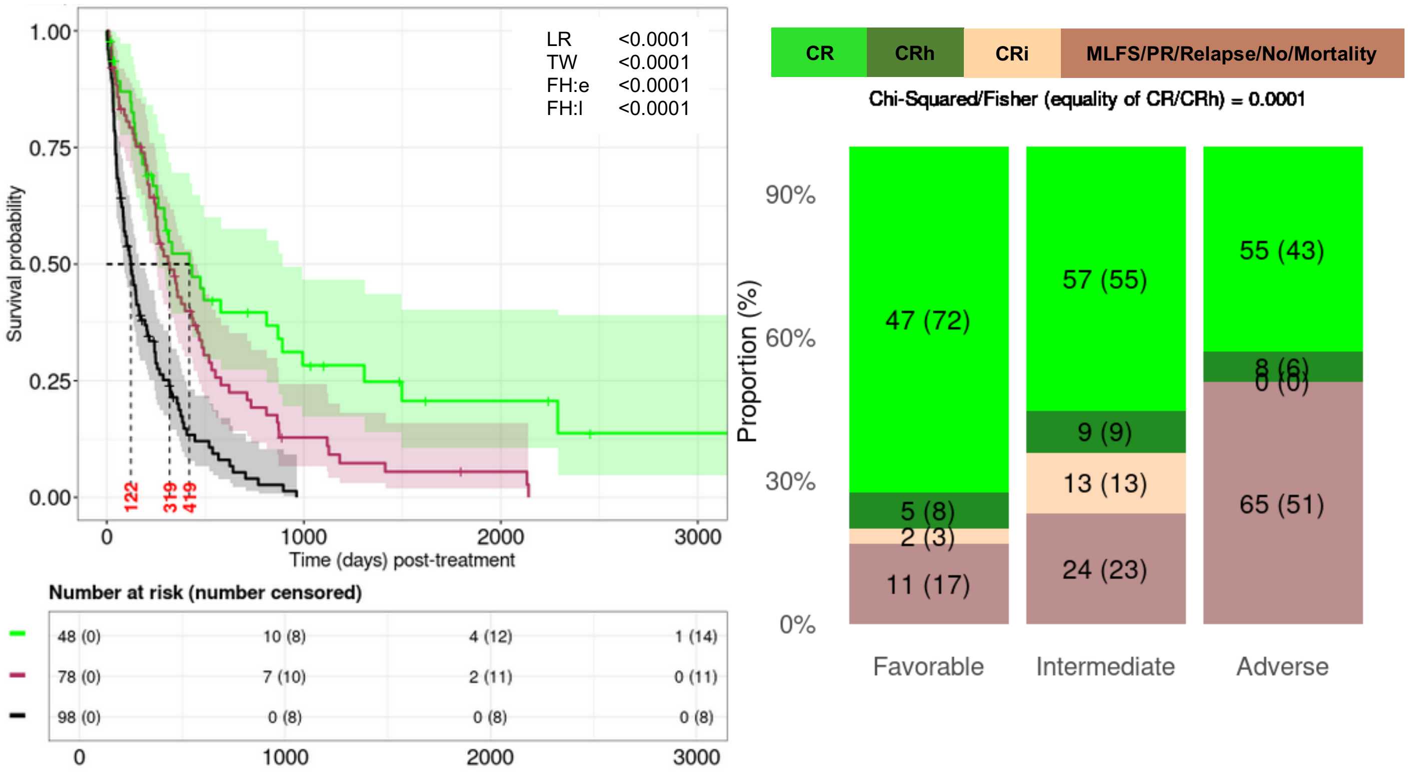


**C. $RM_{G,III}$ for OS (left) and best response (right), with respect to the FAS.**

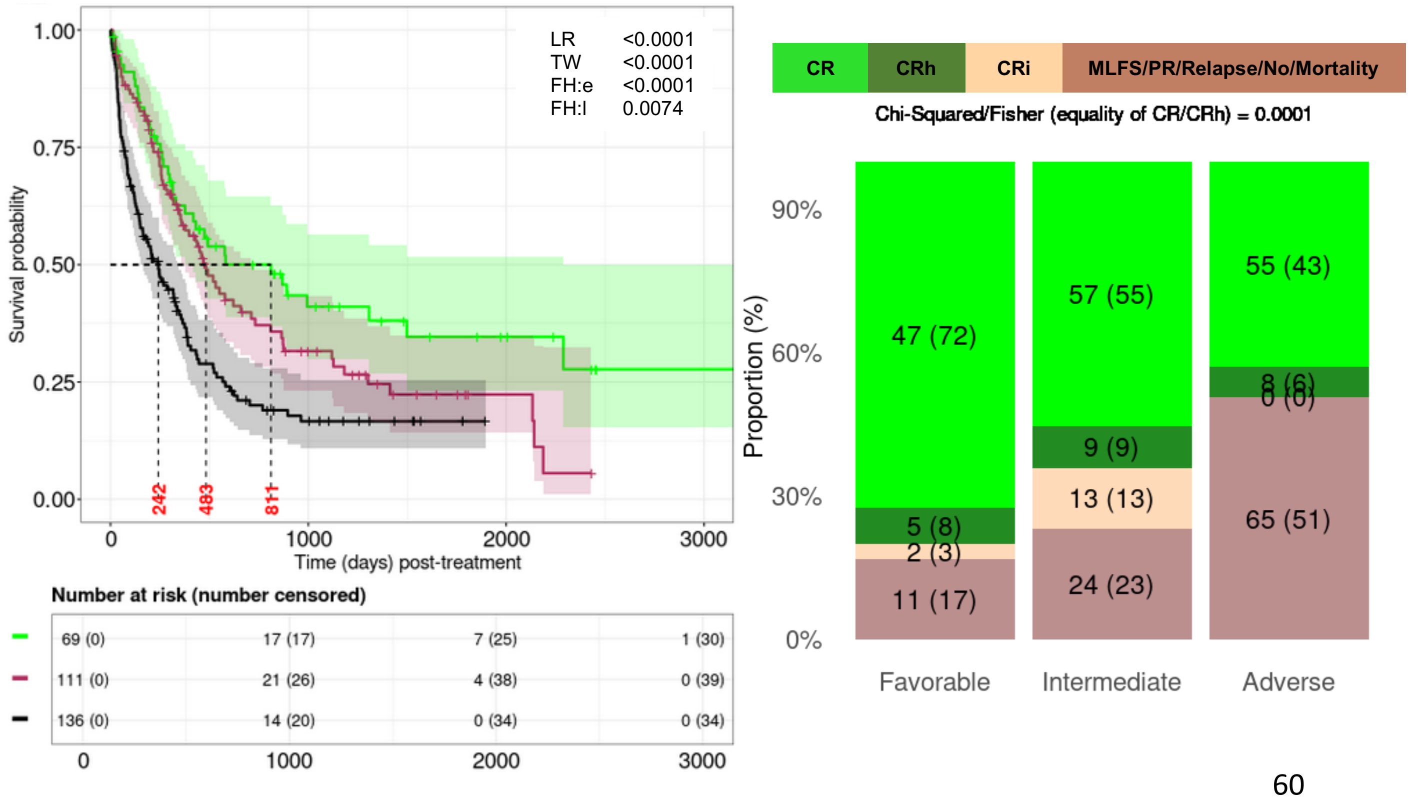

**Supplemental Figure 3, cont'd**

**D. $RM_{G,III}$ on OS censoring allo-HCT (left) and only within allo-HCT (right) recipients.**

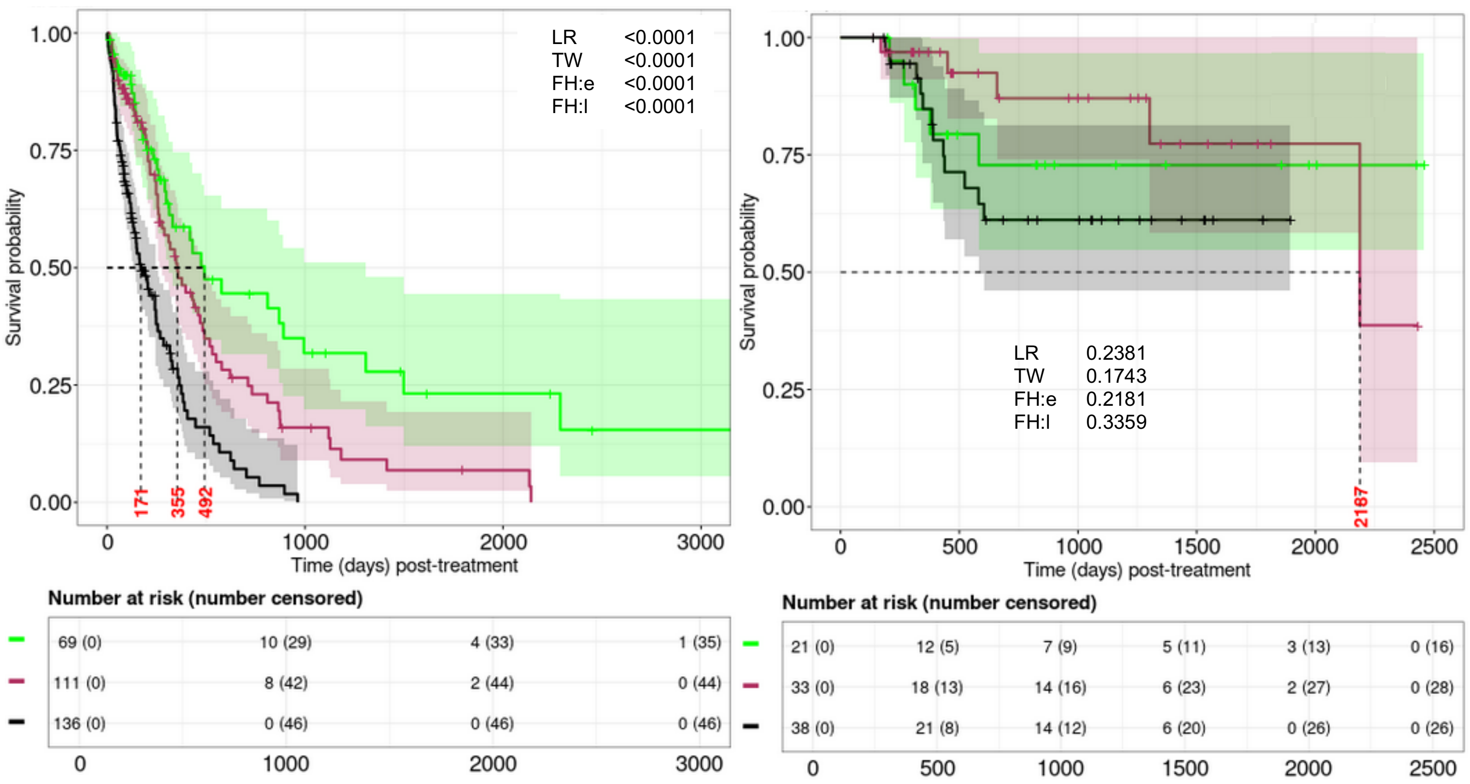

**Supplemental Figure 4. Overall survival (OS) based on the genetic features specific risk model using Rule-IV ($RM_{G,IV}$).**

**A. Feature identification strategy and $RM_{G,IV}$ features.**

| Category | Genetic features | Frequency[†] | Frequency[¶] |
|---|---|---|---|
| **Favorable** | Any of ***DDX41, IDH1, NPM1*** (in the absence of any Adverse features) | **46/224 (21%)** | **67/316 (21%)** |
| **Intermediate** | No Favorable or Adverse features, or $\geq$1 Favorable features with $\geq$1 Adverse features | **79/224 (35%)** | **114/316 (36%)** |
| **Adverse** | Any of ***KRAS, KMT2A, PTPN11, SMC1A, JAK2, p53m, Del7q, Minus5, Minus17, Complex cytogenetics*** (in the absence of any Favorable features) | **99/224 (44%)** | **135/316 (43%)** |

[†] Counts (proportions) of patients after excluding allo-HCT
[¶] Counts (proportions) of patients without excluding allo-HCT

**Supplemental Figure 4, cont'd**

**B. $RM_{G,IV}$ on OS (left) and best response (right), excluding allo-HCT recipients.**

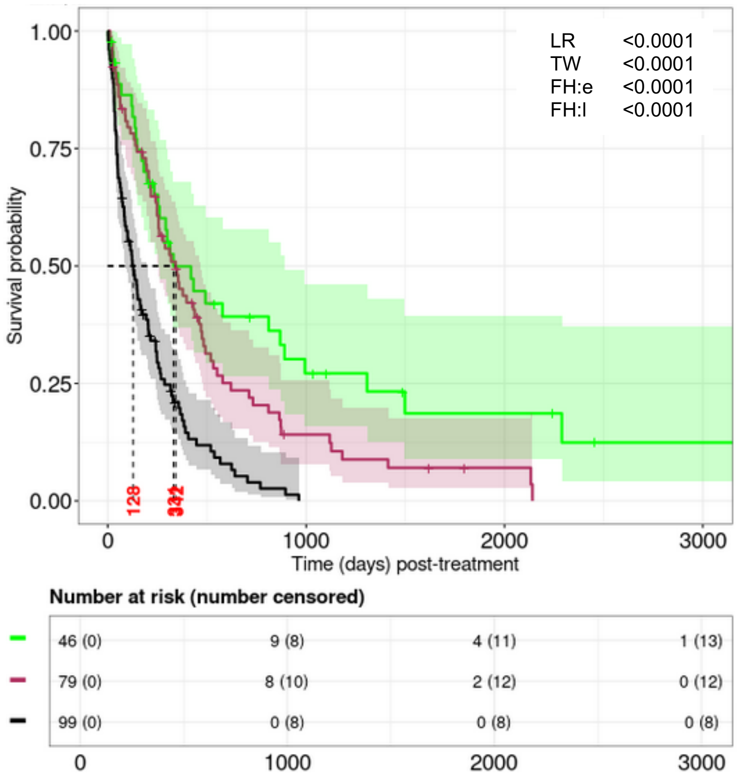


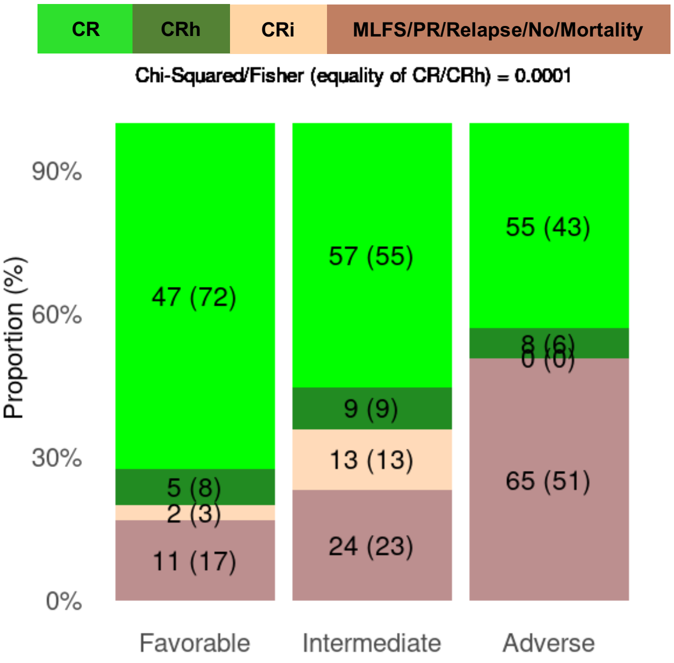


**C. $RM_{G,IV}$ for OS (left) and best response (right), with respect to the FAS.**

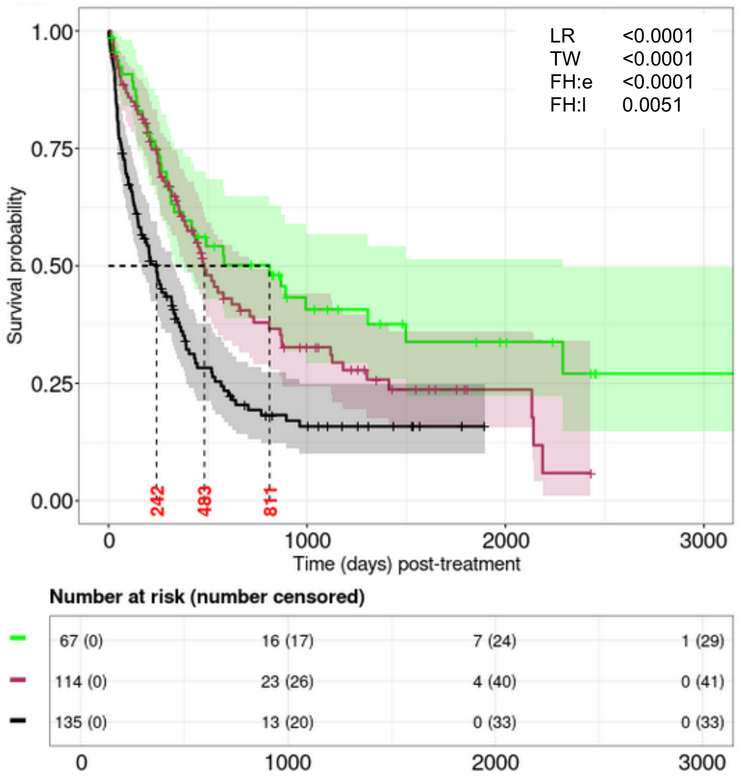


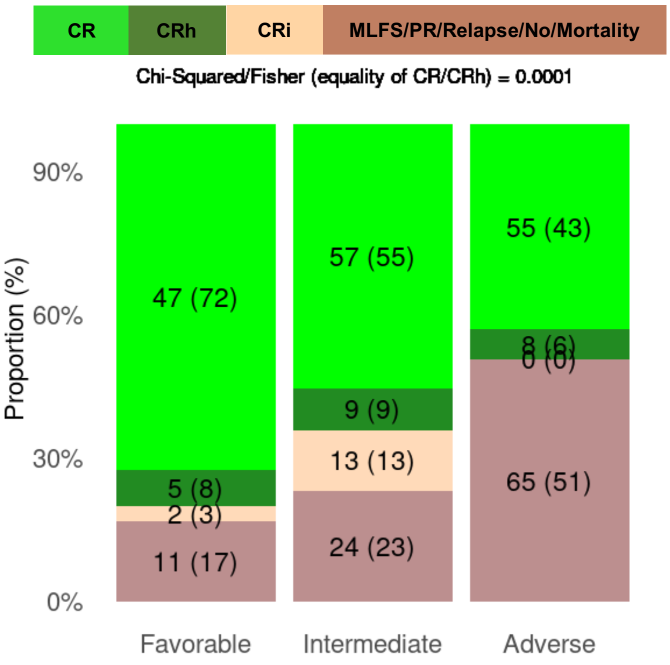

**Supplemental Figure 4, cont'd**

**D. $RM_{G,IV}$ on OS censoring allo-HCT (left) and only within allo-HCT (right) recipients.**

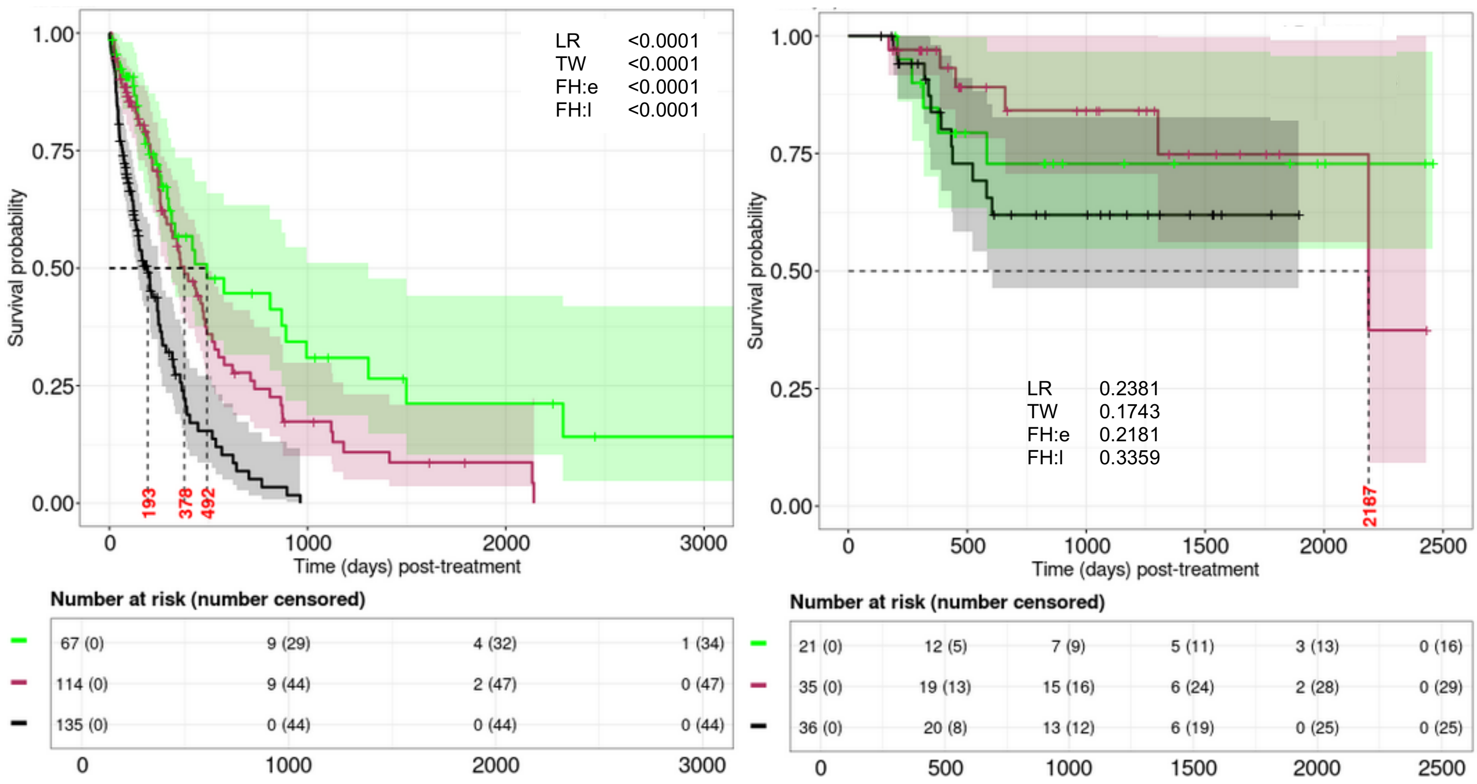

**Supplemental Figure 5. ELN22 and comparison with respect to $RM_{G,II}$ for overall survival (OS).**

**A. ELN22 on OS (left) and best response (right), excluding allo-HCT patients.**

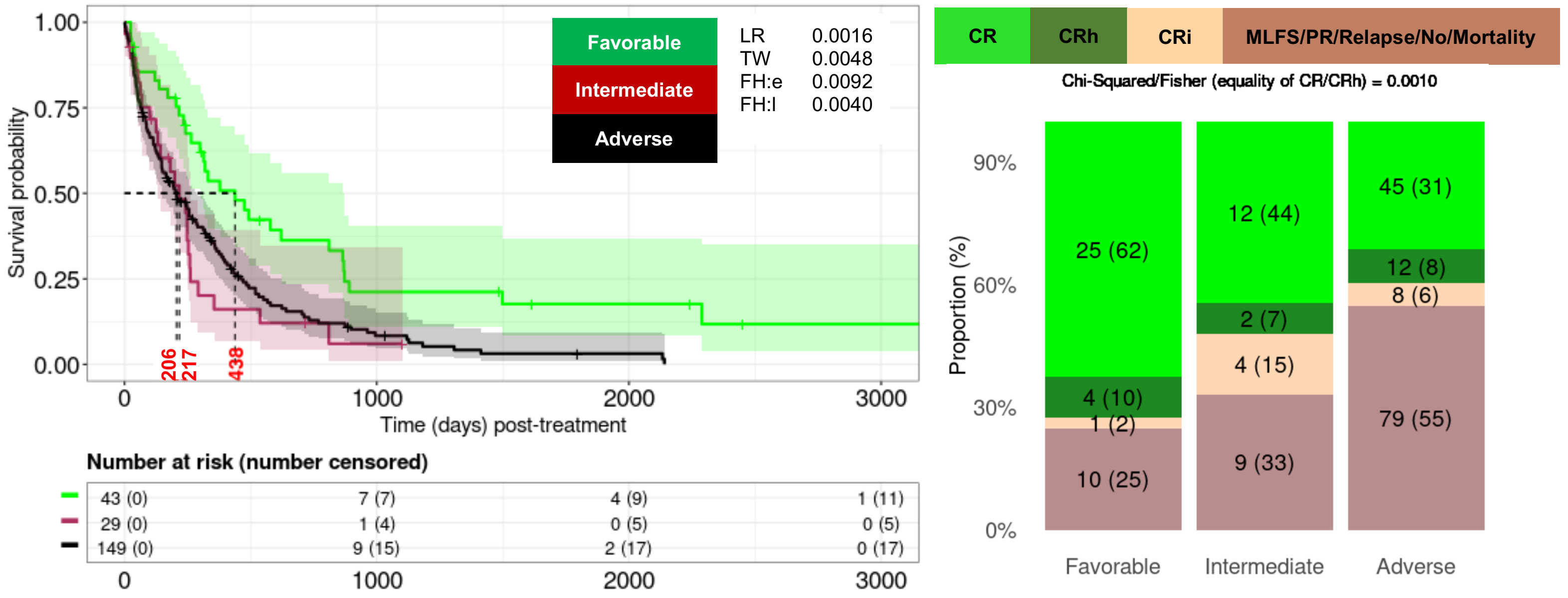


**B. ELN22 OS (left) and best response (right), with respect to the FAS.**

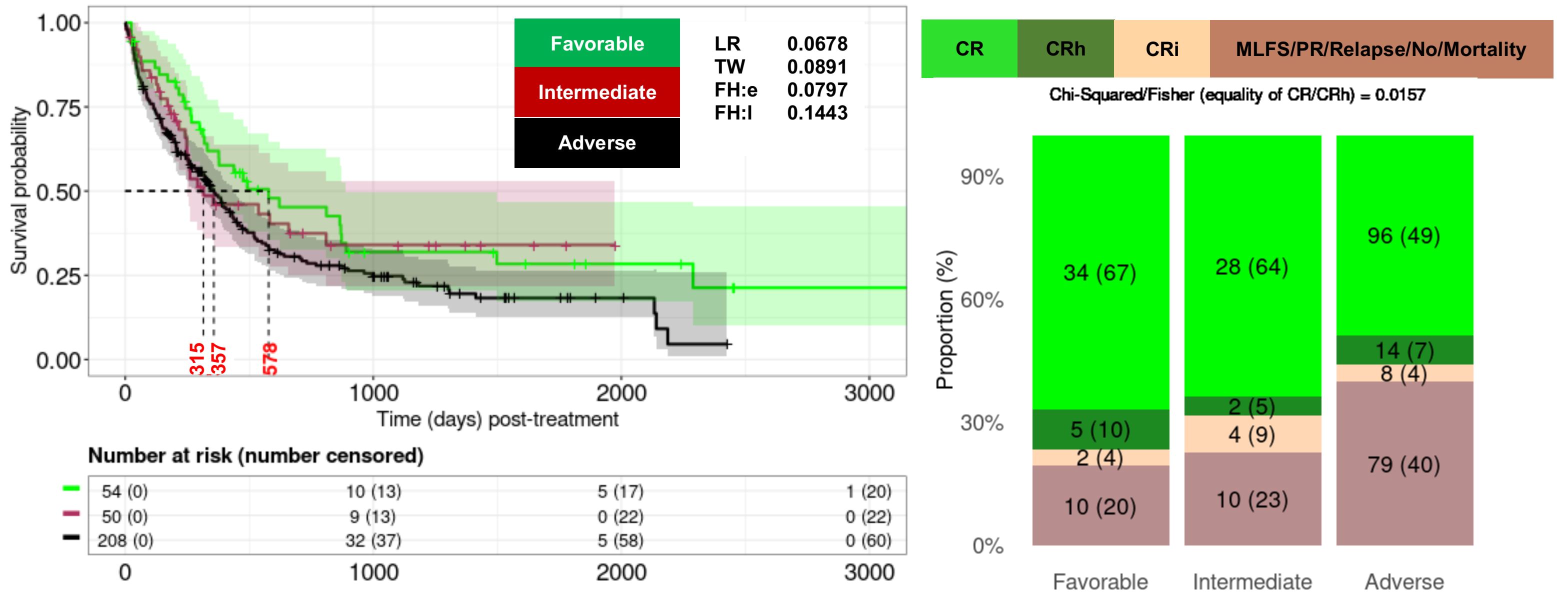

**Supplemental Figure 5, cont'd**

### C. Pairwise comparison between $RM_{G,II}$ and ELN22, excluding allo-HCT recipients.

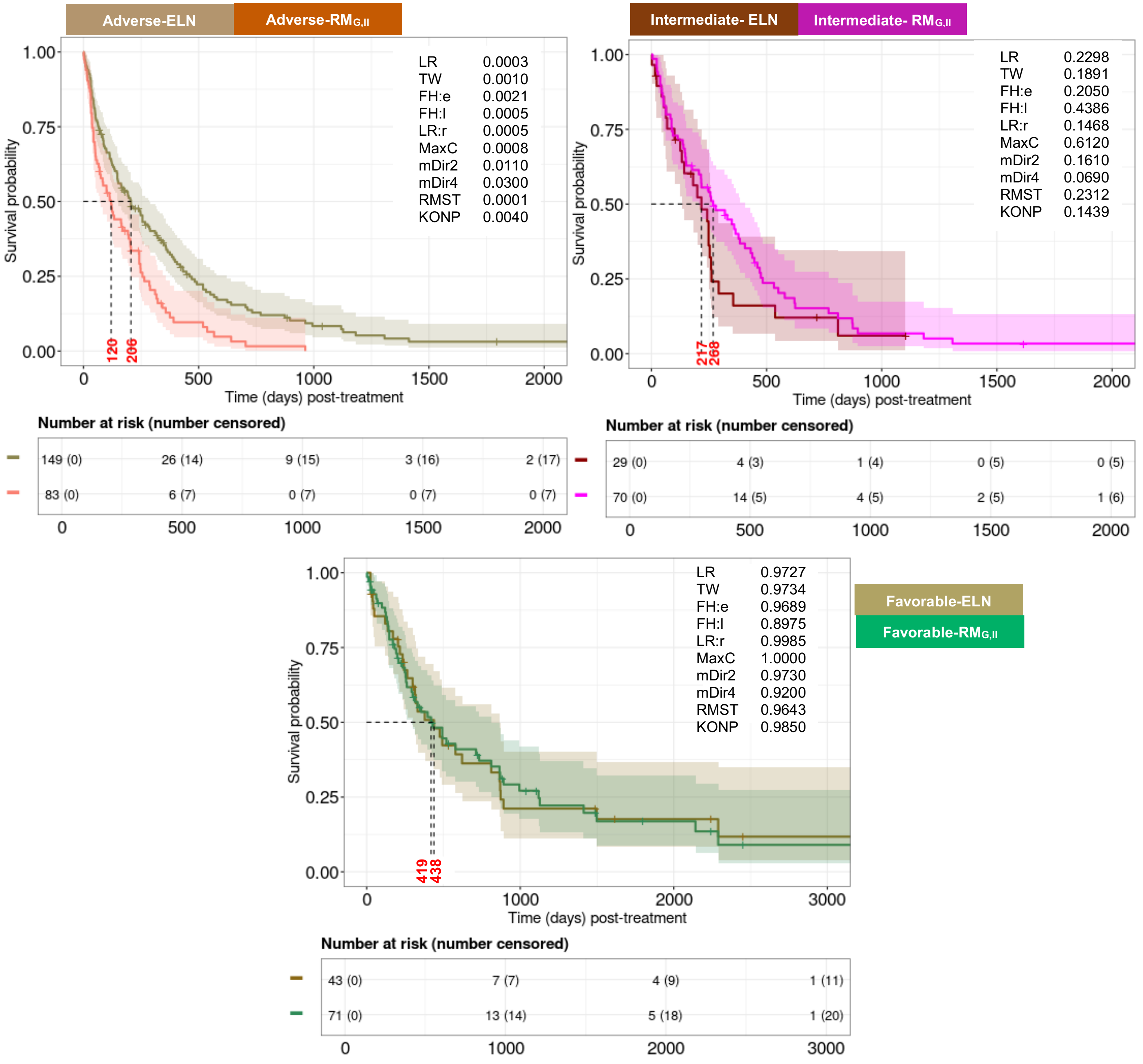

**Supplemental Figure 5, cont'd**

**D. Agreement between ELN22 and RM$_{G,II}$.**

| | Favorable | Intermediate | Adverse |
|---|---|---|---|
| [a]Fleiss kappa (*P* values) | **0.25 (<0.001)** | **-0.10 (0.090)** | **0.17 (0.002)** |

**Remarks:**
- The higher positive value (i.e., close to 1) means more agreement
- The lower negative value (i.e., close to -1) means less agreement
- Value close to 0 means agreement is no better than obtained by chance

[a]Analytical set is comprised of the patients for who ELN22 was observed in the CU dataset.

**E. Predictive evaluation of ELN22 and RM$_{G,II}$.**

| [a]Model | M-iAUC ($25^{th}$,$75^{th}$) over $t$ | M-cAUC ($25^{th}$,$75^{th}$) over $t$ | iBrier ($25^{th}$,$75^{th}$) over $t$ |
|---|---|---|---|
| **ELN22** | 0.55 [0.54,0.57] | 0.50 [<0.50,0.58] | 0.16 [0.13,0.20] |
| **RM$_{G,II-EFS}$** | 0.64 [0.59,0.66] (<0.001) | 0.70 [0.65,0.77] (<0.001) | 0.14 [0.12,0.19] |

- [a]Penalized CoxPH model adjusted by age, gender, race, and either ELN22 or RM$_{G,II}$.
- Reported in parenthesis are the median of P-values (over 15-fold CV) testing the null hypothesis of inferiority of distributions of survival metrics of the RM$_{G,II}$ relative to that of the ELN22.

**Supplemental Figure 6. Overall survival (OS) based on the genetic-plus-phenotype features specific risk model using Rule-I ($RM_{GP,I}$).**

**A. Feature identification strategy and $RM_{GP,I}$ specific features.**

| Category | Genetic plus phenotypic features | Frequency† | Frequency¶ |
|---|---|---|---|
| **Favorable** | Any of ***CD10, CD19, BCOR, DNMT3A, DDX41, SF3B1, Good risk cytogenetics, IDH1, IDH2, KIT, NPM1*** (in the absence of any Adverse features) | **29/224 (13%)** | **39/316 (12%)** |
| **Intermediate** | No Favorable or Adverse features, or ≥1 Favorable features with ≥1 Adverse features | **94/224 (42%)** | **140/316 (44%)** |
| **Adverse** | Any of ***CD5, CD7, CD16, CD34, CD36, CD71, CD11C, CSF3R, KRAS, KMT2A, MPL, PTPN11, SMC1A, JAK2, p53m, Del7q, Inv(3), t(9;11), Minus5, Minus17, Complex cytogenetics, Chr 7 abn:loss, Chr 8 abn:loss, DEK:loss, MECOM:gain, RARA:loss*** (in the absence of any Favorable features) | **101/224 (45%)** | **137/316 (43%)** |

† Counts (proportions) of patients after excluding allo-HCT
¶ Counts (proportions) of patients without excluding allo-HCT

**Supplemental Figure 6, cont'd**

**B. $RM_{GP,I}$ on OS (left) and best response (right), excluding allo-HCT patients.**

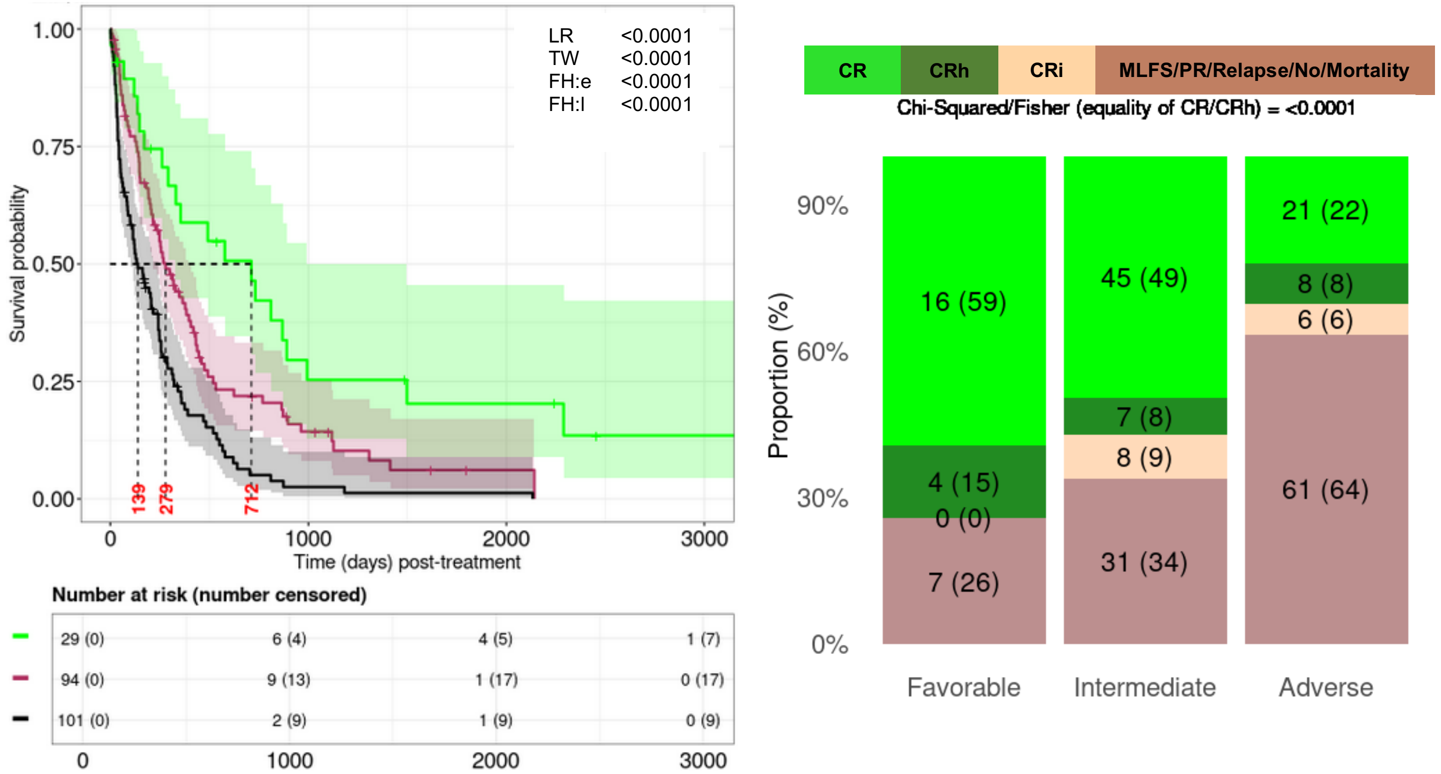


**C. $RM_{GP,I}$ on OS (left) and best response (right), with respect to the FAS.**

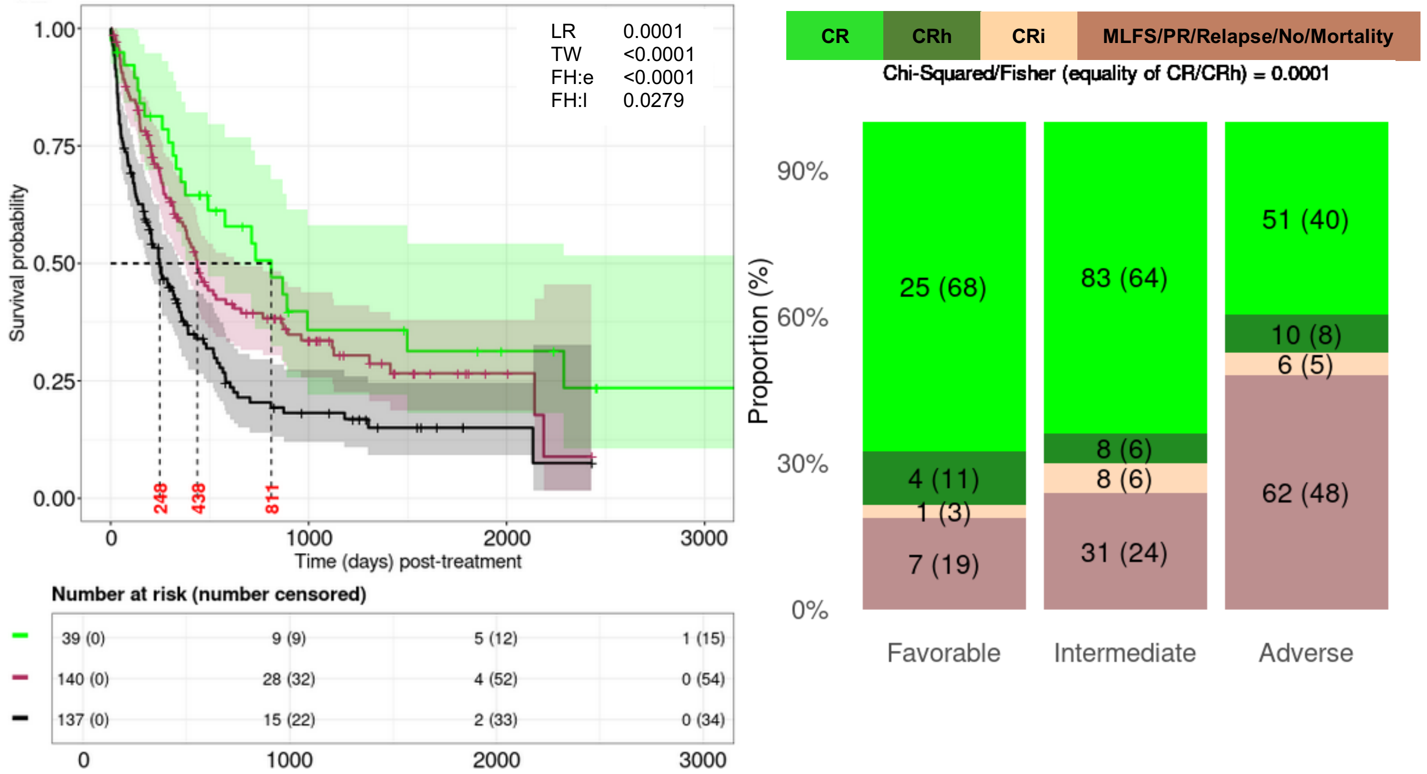

**Supplemental Figure 6, cont'd**

**D. $RM_{GP,I}$ on OS censoring allo-HCT (left) and only within allo-HCT (right) recipients.**

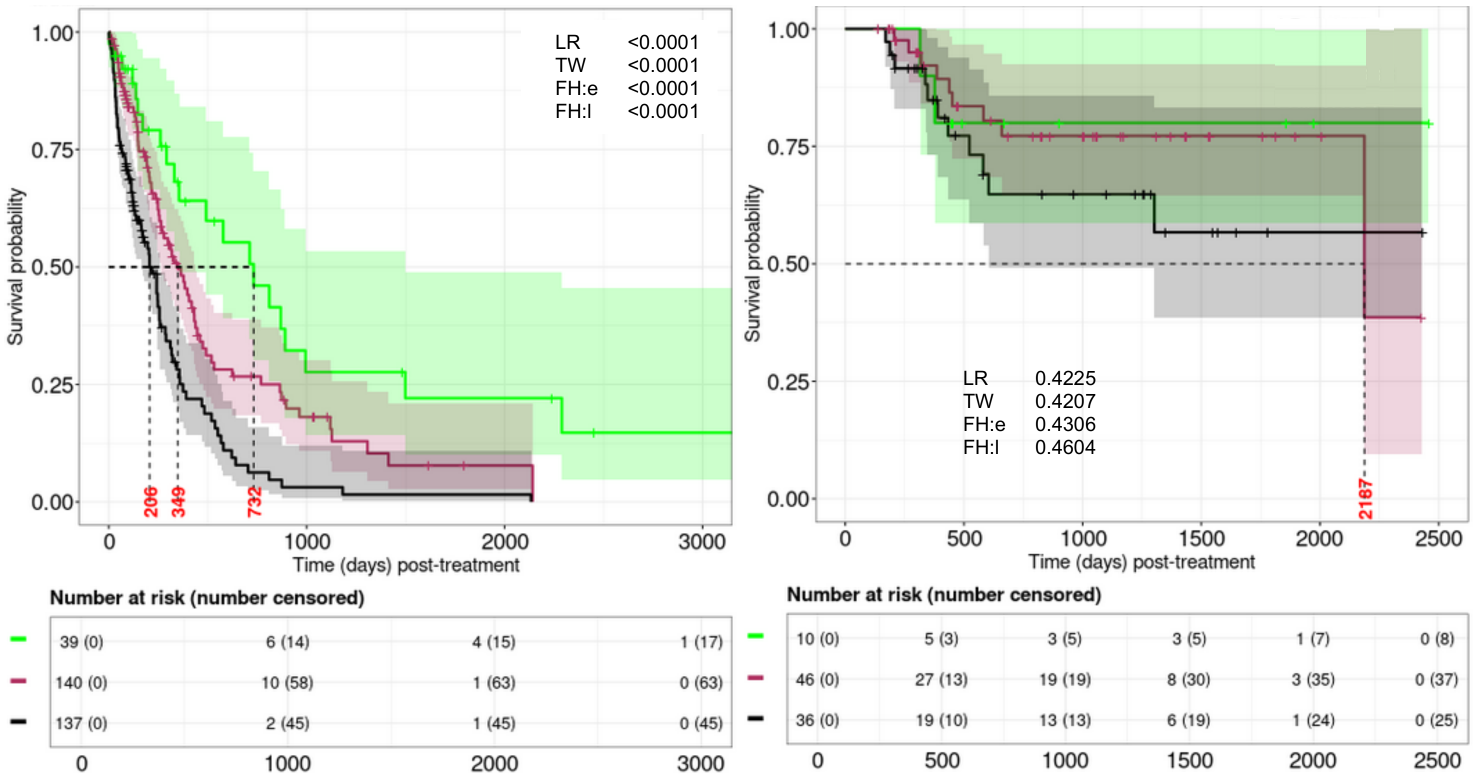

**Supplemental Figure 7. Overall survival (OS) based on the genetic-plus-phenotypic features specific risk model using Rule-II ($RM_{GP,II}$).**

**A. Feature identification strategy and $RM_{GP,II}$ features.**

| Category | Genetic plus phenotypic features | Frequency[†] | Frequency[¶] |
|---|---|---|---|
| **Favorable** | Any of ***CD10, CD19, BCOR, DNMT3A, DDX41, SF3B1, IDH1, IDH2, NPM1*** (in the absence of any Adverse features) | **57/224 (25%)** | **82/316 (26%)** |
| **Intermediate** | No Favorable or Adverse features, or ≥1 Favorable features with ≥1 Adverse features | **79/224 (35%)** | **116/316 (37%)** |
| **Adverse** | Any of ***CD5, CD7, CD36, CD11C, KRAS, KMT2A, PTPN11, SMC1A, JAK2, p53m, Del7q, Minus5, Minus17, t(9;11), Complex cytogenetics, Chr 7 abn:loss, Chr 8 abn:loss, DEK:loss, MECOM:gain, RARA:loss*** (in the absence of any Favorable features) | **88/224 (39%)** | **118/316 (37%)** |

[†] Counts (proportions) of patients after excluding allo-HCT
[¶] Counts (proportions) of patients without excluding allo-HCT

**Supplemental Figure 7, cont'd**

**B. $RM_{GP,II}$ on OS (left) and best response (right), excluding allo-HCT recipients.**

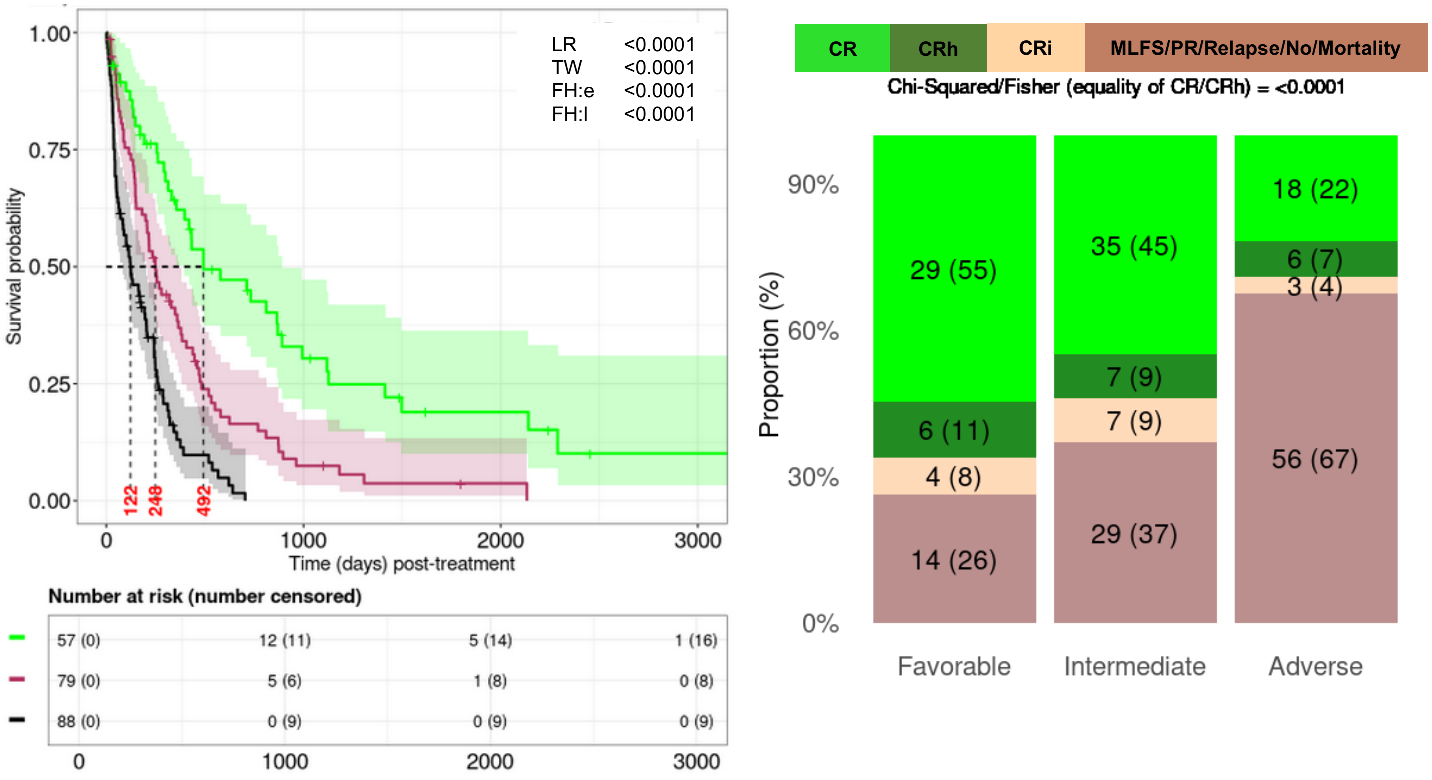


**C. $RM_{GP,II}$ on OS (left) and best response (right), with respect to the FAS.**

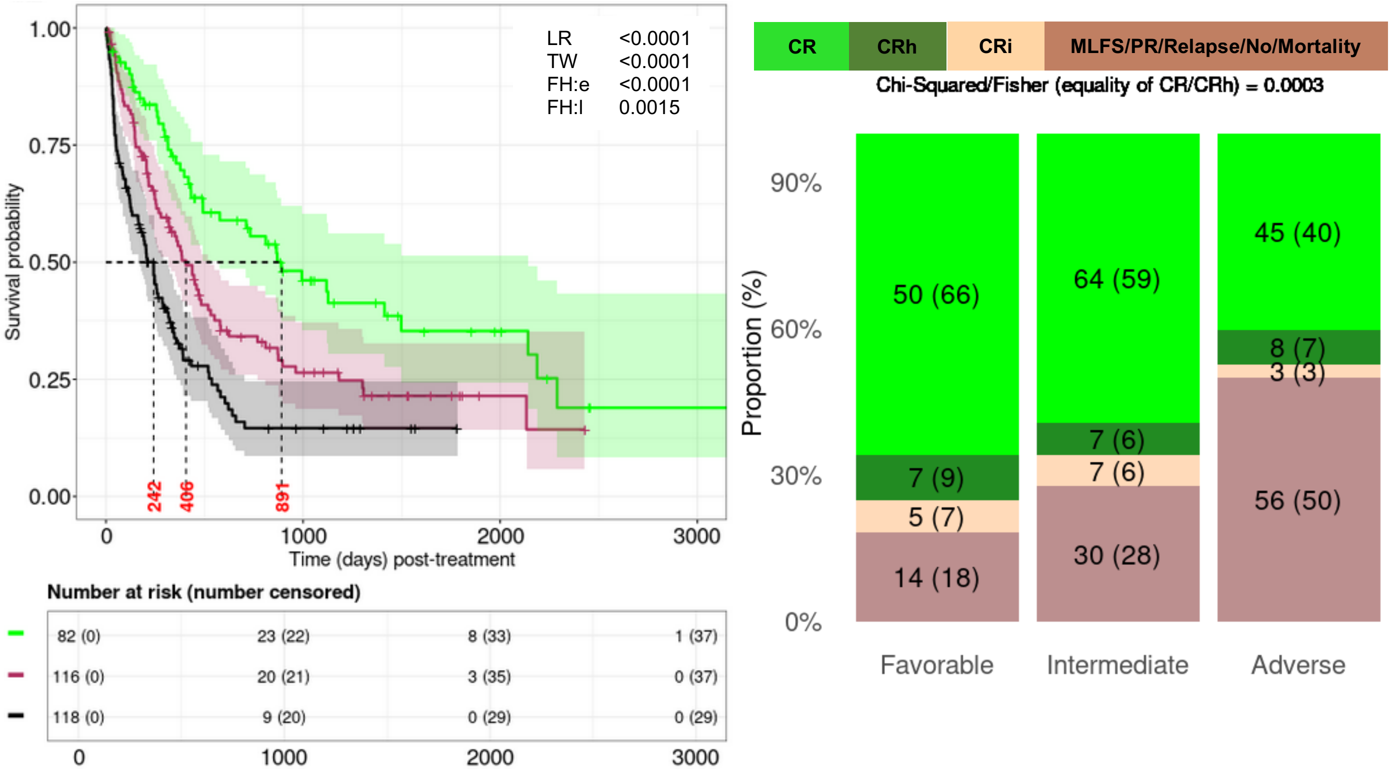

**Supplemental Figure 7, cont'd**

**D. $RM_{GP,II}$ on OS censoring allo-HCT (left) and only within allo-HCT (right) recipients.**

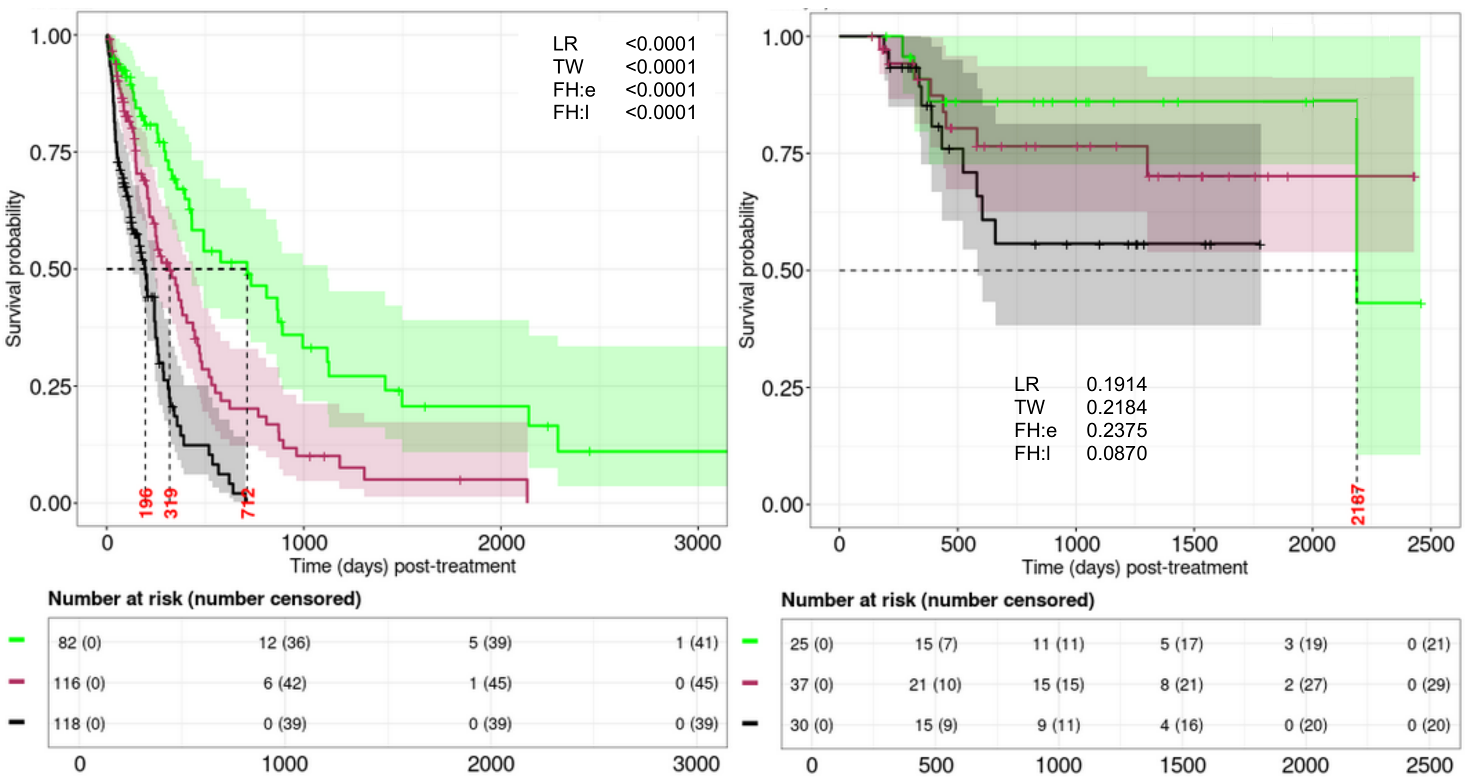

**Supplemental Figure 8. Overall survival (OS) based on the genetic-plus-phenotypic features specific risk model using Rule-III ($RM_{GP,III}$).**

**A. Feature identification strategy and $RM_{GP,III}$ features.**

| Category | Genetic plus phenotypic features | Frequency[†] | Frequency[¶] |
|---|---|---|---|
| **Favorable** | Any of ***CD10, DDX41, Good risk cytogenetics, IDH1, NPM1*** (in the absence of any Adverse features) | **45/224 (20%)** | **64/316 (20%)** |
| **Intermediate** | No Favorable or Adverse features, or ≥1 Favorable features with ≥1 Adverse features | **82/224 (37%)** | **114/316 (36%)** |
| **Adverse** | Any of ***CD16, CD36, CD71, KRAS, KMT2A, PTPN11, SMC1A, MPL, JAK2, p53m, Inv(3), Del7q, Minus5, Minus17, Complex cytogenetics*** (in the absence of any Favorable features) | **97/224 (43%)** | **138/316 (44%)** |

[†] Counts (proportions) of patients after excluding allo-HCT
[¶] Counts (proportions) of patients without excluding allo-HCT

**Supplemental Figure 8, cont'd**

**B. RM$_{GP,III}$ on OS (left) and best response (right), excluding allo-HCT patients.**

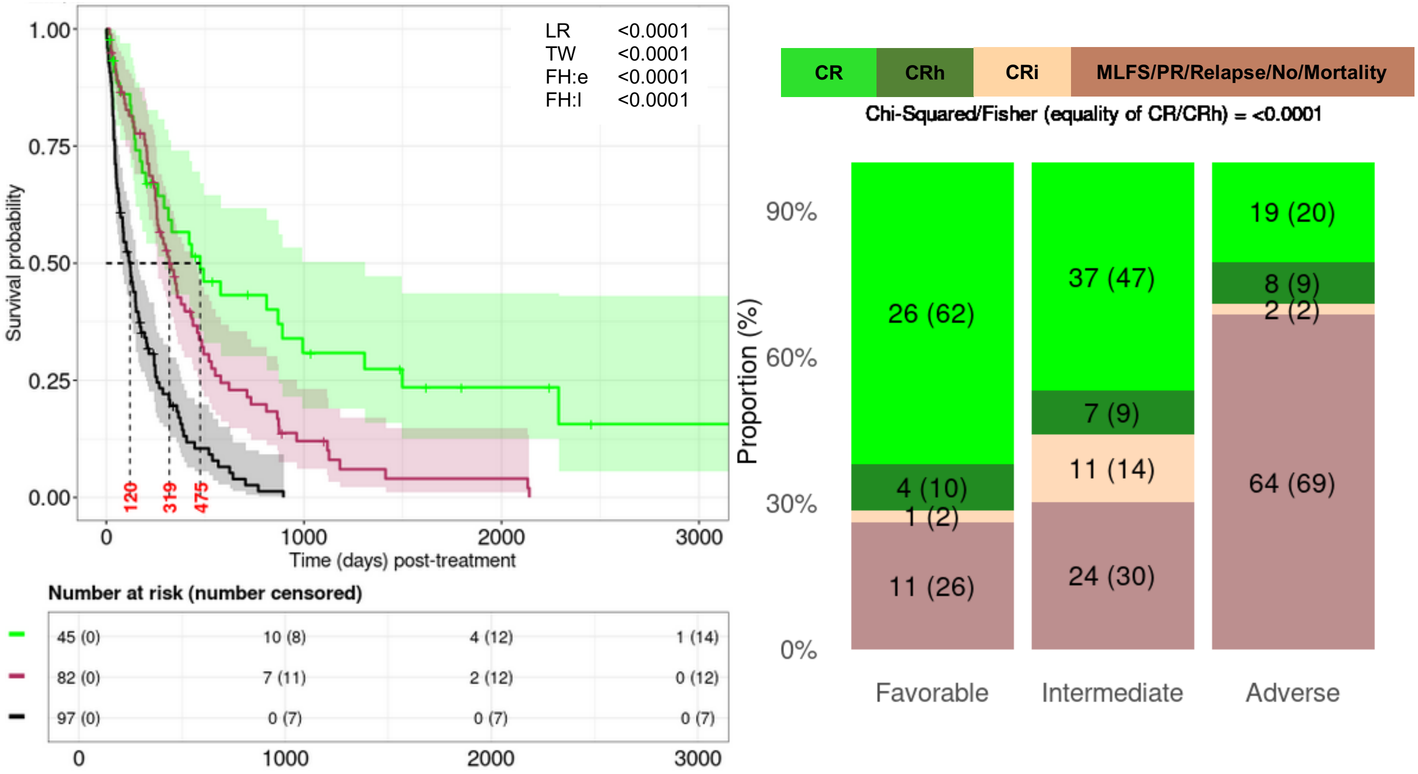


**C. RM$_{GP,III}$ on OS (left) and best response (right), with respect to the FAS.**

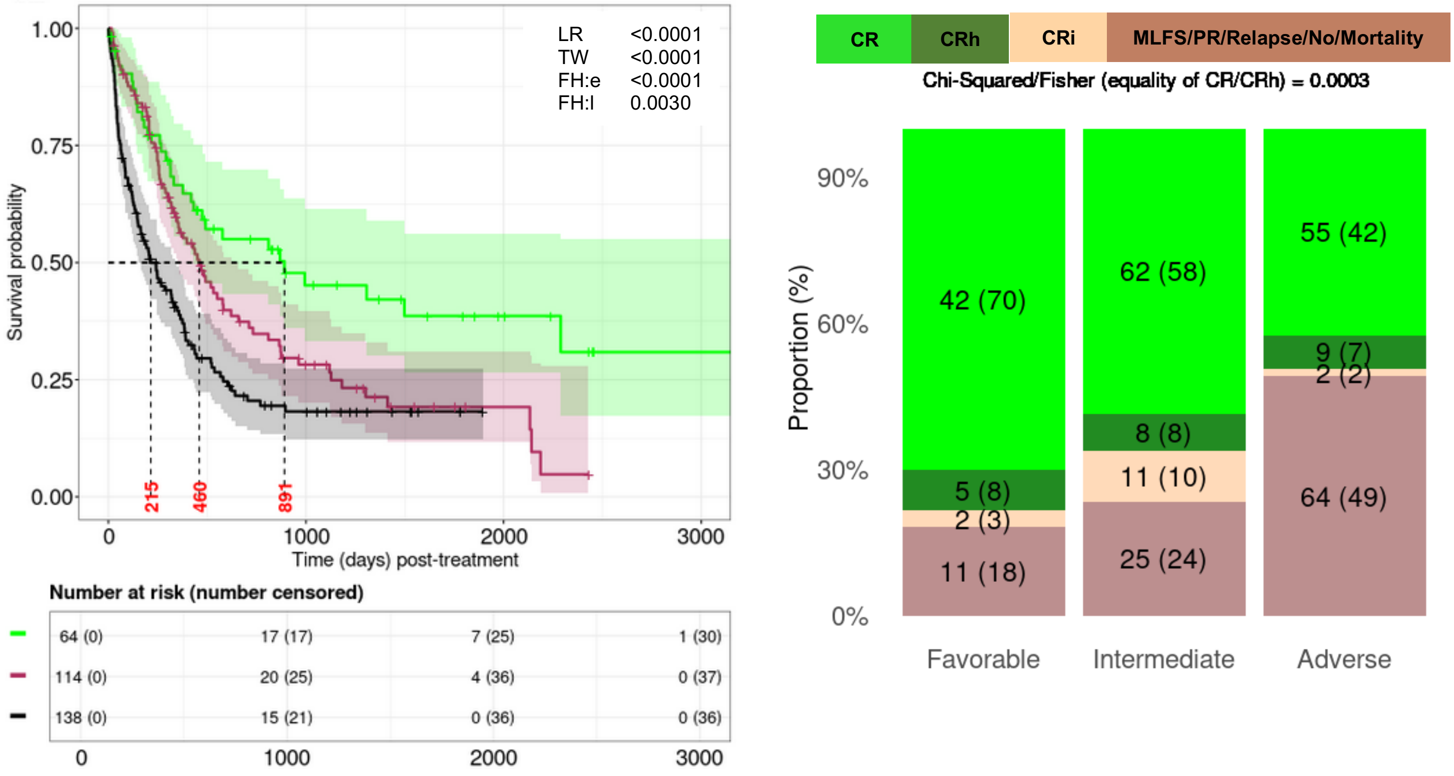

**Supplemental Figure 8, cont'd**

**D. $RM_{GP,III}$ on OS censoring allo-HCT (left) and only within allo-HCT (right) recipients.**

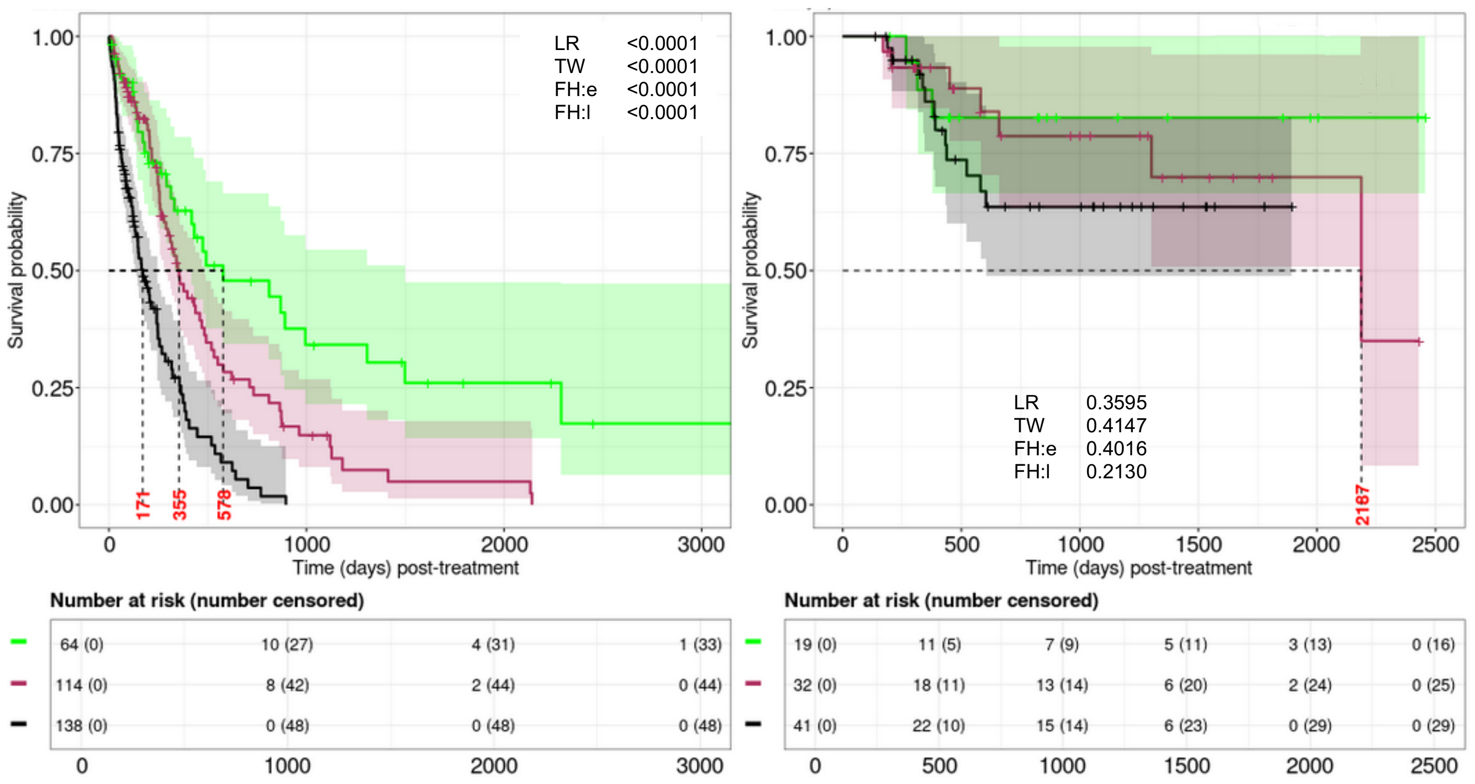

**Supplemental Figure 9. Overall survival (OS) based on the genetic-plus-phenotypic features specific risk model using Rule-IV ($RM_{GP,IV}$).**

**A. Feature identification strategy and $RM_{GP,IV}$ features.**

| Category | Genetic plus phenotypic features | Frequency† | Frequency¶ |
|---|---|---|---|
| **Favorable** | Any of ***CD10, DDX41, IDH1, NPM1*** (in the absence of any Adverse features) | **44/224 (20%)** | **63/316 (20%)** |
| **Intermediate** | No Favorable or Adverse features, or $\geq$1 Favorable features with $\geq$1 Adverse features | **85/224 (38%)** | **119/316 (38%)** |
| **Adverse** | Any of ***CD36, KRAS, KMT2A, PTPN11, SMC1A, JAK2, p53m, Del7q, Minus5, Minus17, Complex cytogenetics*** (in the absence of any Favorable features) | **95/224 (42%)** | **134/316 (42%)** |

† Counts (proportions) of patients after excluding allo-HCT
¶ Counts (proportions) of patients without excluding allo-HCT

**Supplemental Figure 9, cont'd**

**B. $RM_{GP,IV}$ on OS (left) and best response (right), excluding allo-HCT patients.**

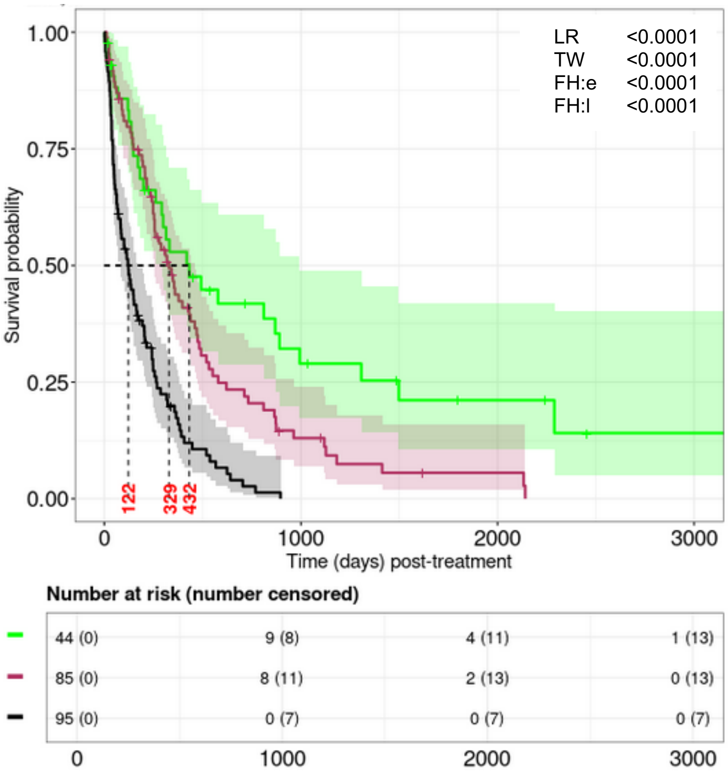


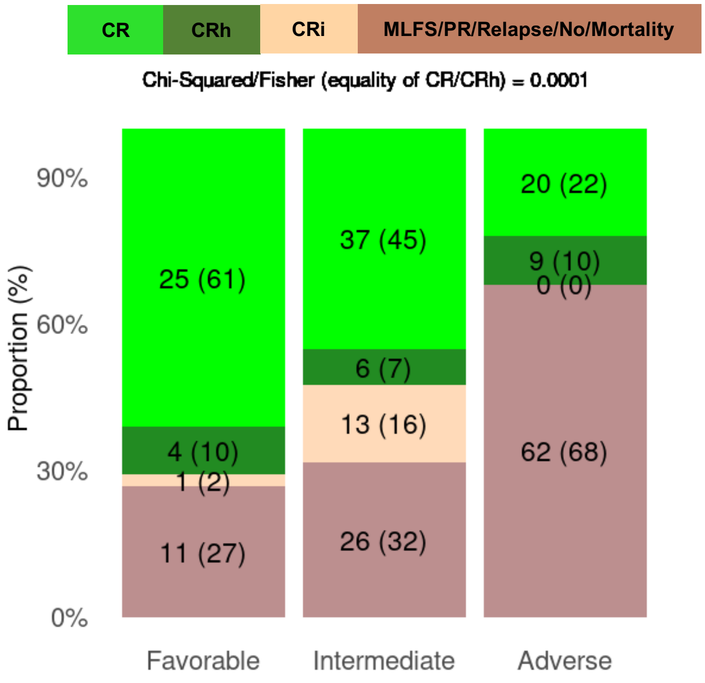


**C. $RM_{GP,IV}$ on OS (left) and best response (right), with respect to the FAS.**

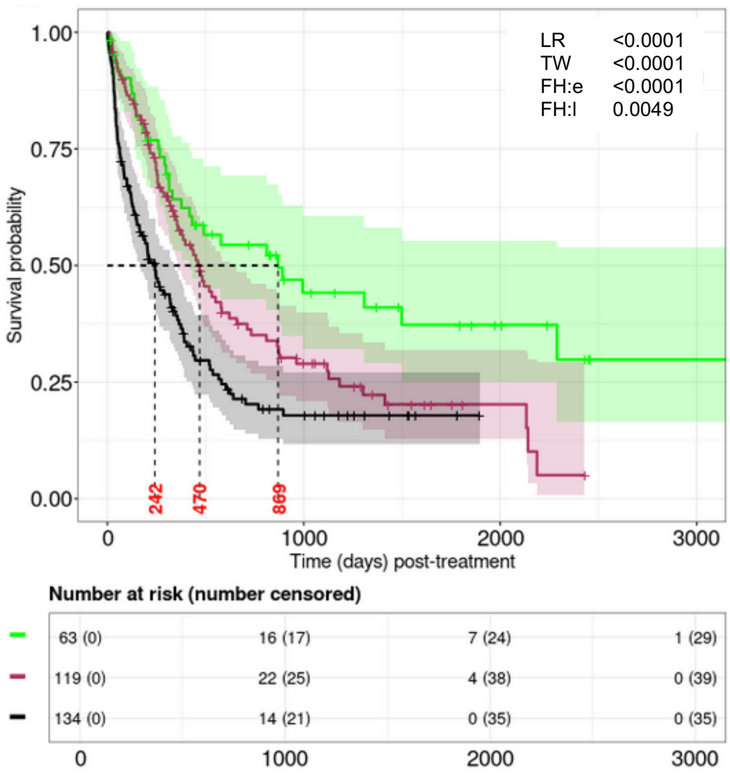


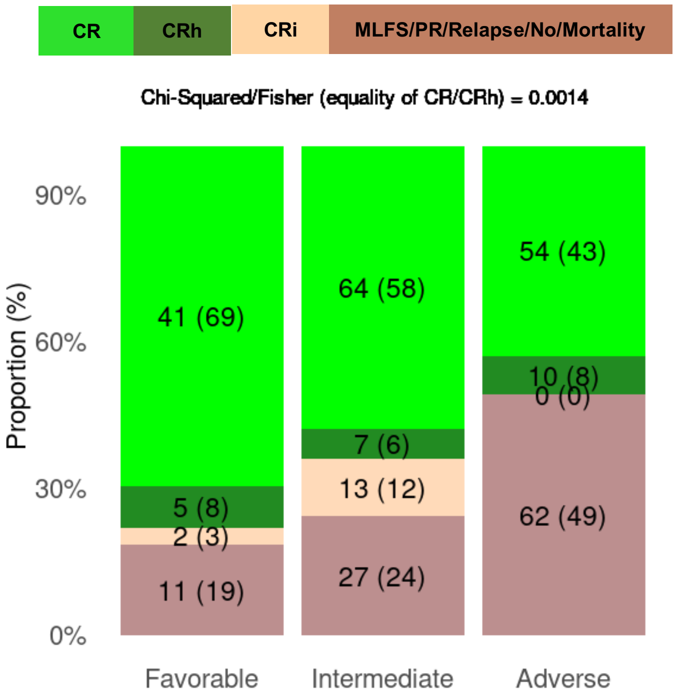

**Supplemental Figure 9, cont'd**

**D. $RM_{GP,IV}$ censoring allo-HCT (left) and only within allo-HCT (right) recipients.**

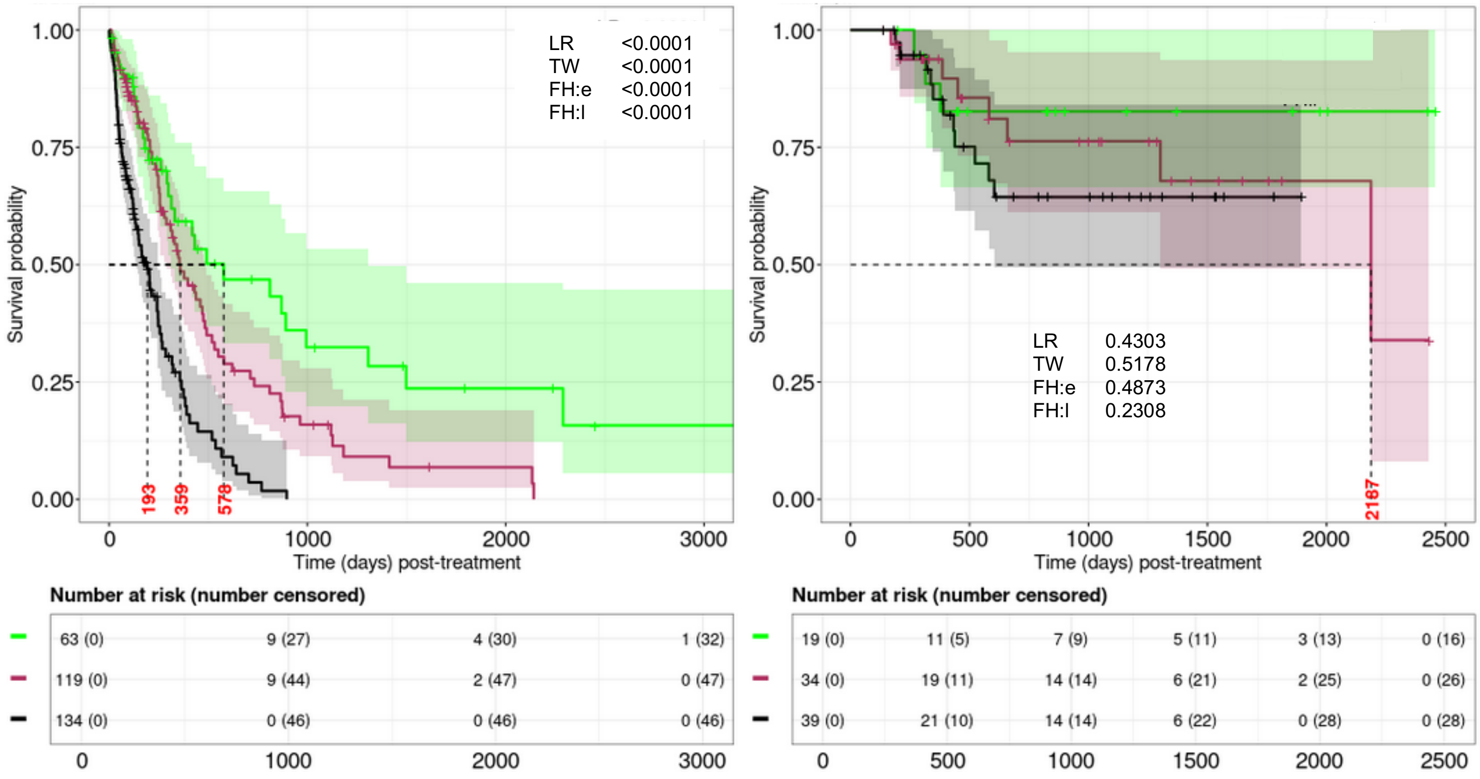

**Supplemental Figure 10. Event free survival (EFS) based on the genetic features specific risk model using Rule-I ($RM_{G,I-EFS}$).**

**A. Feature identification strategy and $RM_{G,I-EFS}$ features.**

| Category | Genetic features | Frequency[†] | Frequency[¶] |
|---|---|---|---|
| **Favorable** | Any of ***BCOR, Cytogenetics normal, Monosomal, Good cytogenetics, MECOM:normal, KMT2A:normal, Del5q, IDH1, IDH2, NPM1*** (in the absence of any Adverse features) | **7/211 (3%)** | **10/303 (3%)** |
| **Intermediate** | No Favorable or Adverse features, or ≥1 Favorable features with ≥1 Adverse features | **183/211 (87%)** | **261/303 (86%)** |
| **Adverse** | Any of ***KRAS, KMT2A, PTPN11, JAK2, p53m, Del7q, Inv(3), t(9;11), Minus5, Minus17, Complex cytogenetics, Indeterminant cytogenetics, Intermediate cytogenetics, Chr 7 abn:loss, Chr 8 abn:loss, RARA:loss, MECOM:rearrangement, KMT2A:rearrangement*** (in the absence of any Favorable features) | **21/211 (10%)** | **32/303 (11%)** |

[†] Counts (proportions) of patients after excluding allo-HCT
[¶] Counts (proportions) of patients without excluding allo-HCT

**Supplemental Figure 10, cont'd**

**B. $RM_{G,I\text{-}EFS}$ with respect to the FAS (left) and dataset excluding allo-HCT (right) patients.**

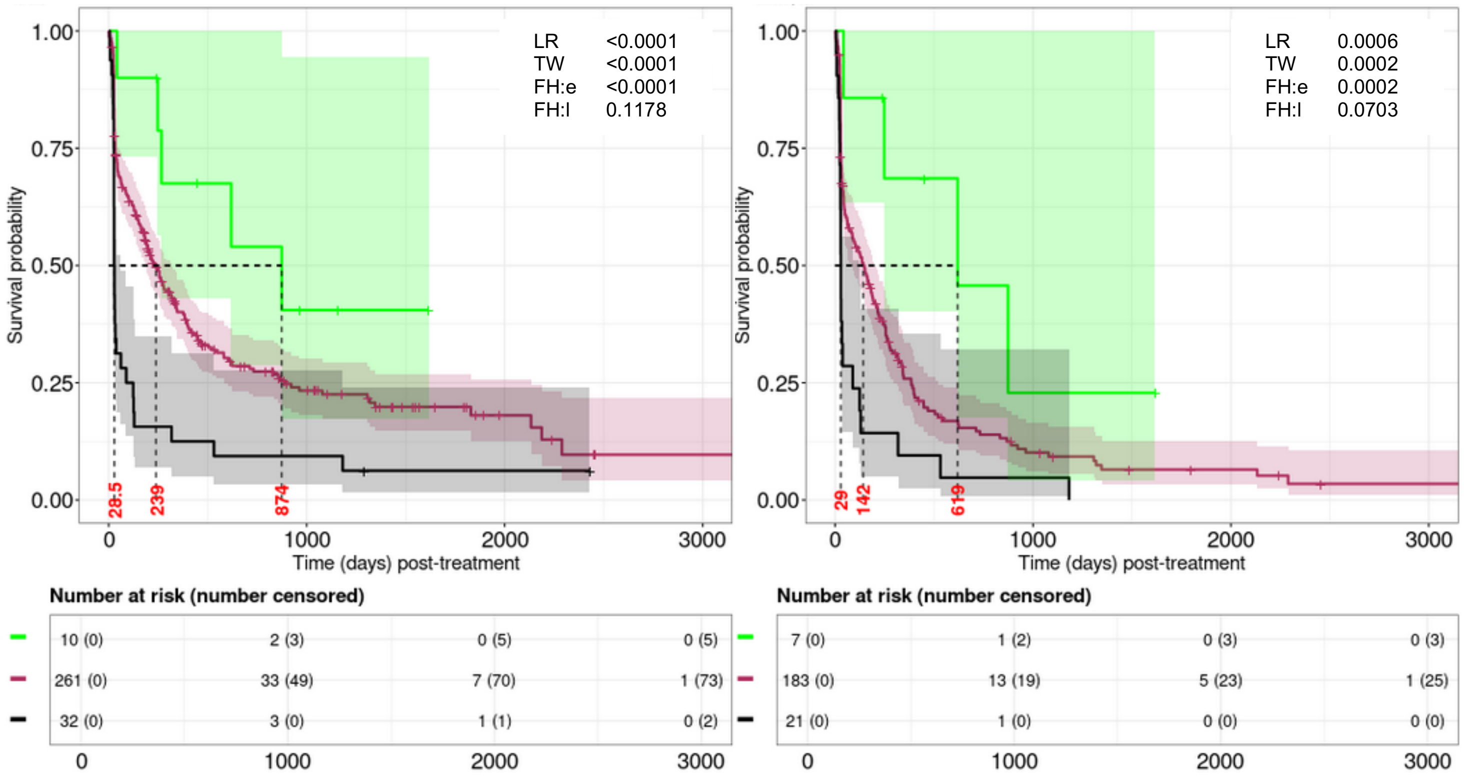


**C. $RM_{G,I\text{-}EFS}$ censoring allo-HCT (left) and only within allo-HCT (right) recipients.**

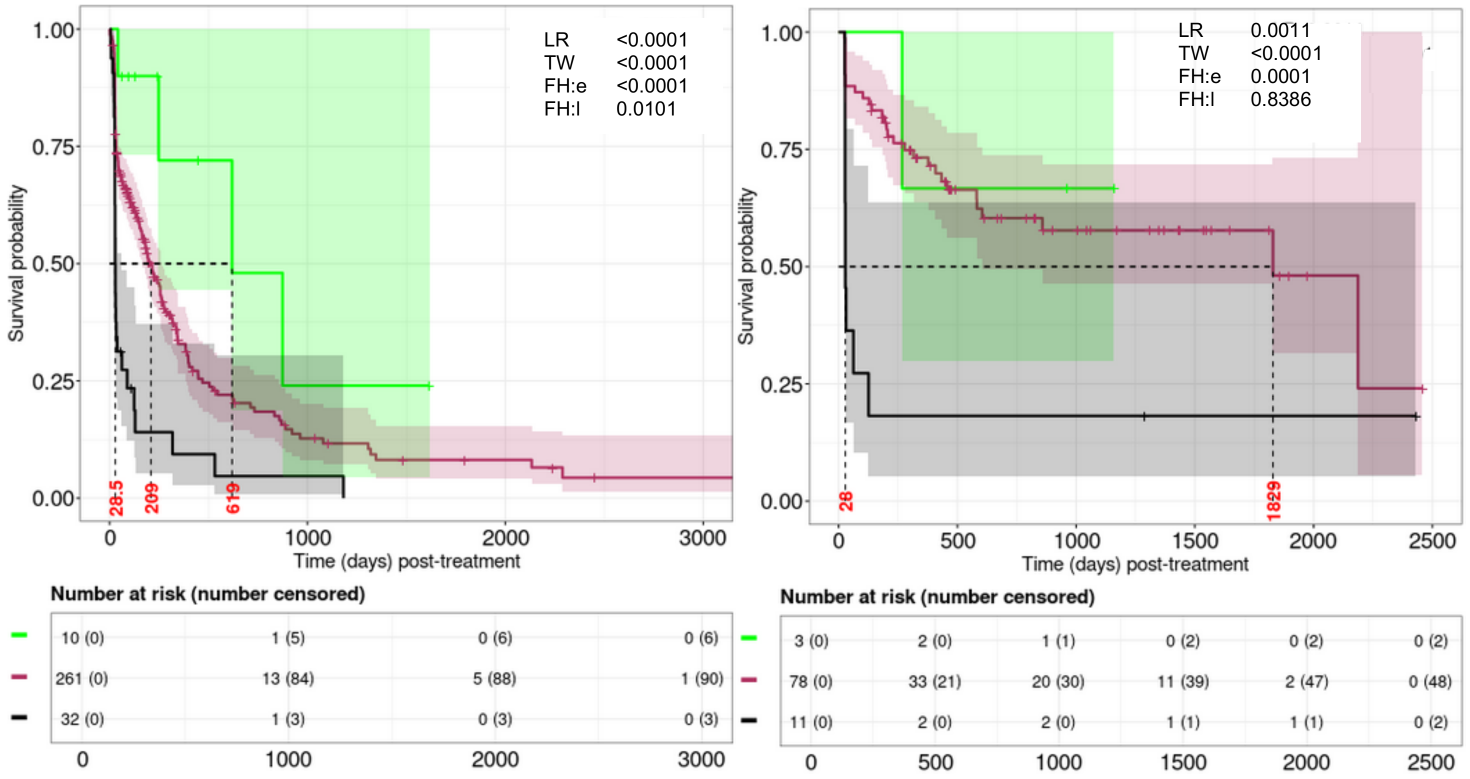

**Supplemental Figure 11. Event free survival (EFS) based on the genetic features specific risk model using Rule-III ($RM_{G,III\text{-}EFS}$).**

**A. Feature identification strategy and $RM_{G,III\text{-}EFS}$ features.**

| Category | Genetic features | Frequency[†] | Frequency[¶] |
|---|---|---|---|
| **Favorable** | Any of ***Normal cytogenetics, Good risk cytogenetics, Del5q, IDH1, IDH2, NPM1*** (in the absence of any Adverse features) | **76/211 (36%)** | **109/303 (36%)** |
| **Intermediate** | No Favorable or Adverse features, or ≥1 Favorable features with ≥1 Adverse features | **49/211 (23%)** | **75/303 (25%)** |
| **Adverse** | Any of ***JAK2, p53m, Inv3, Del7q, t(9;11), Minus5, Minus17, Complex cytogenetics, Indeterminant cytogenetics, Chr 7 abn:loss, Chr 8 abn:loss, MECOM:rearrangement, KMT2A:rearrangement*** (in the absence of any Favorable features) | **86/211 (41%)** | **119/303 (39%)** |

† Counts (proportions) of patients after excluding allo-HCT
¶ Counts (proportions) of patients without excluding allo-HCT

**Supplemental Figure 11, cont'd**

**B. $RM_{G,III\text{-}EFS}$ with respect to the FAS (left) and excluding allo-HCT (right) patients.**

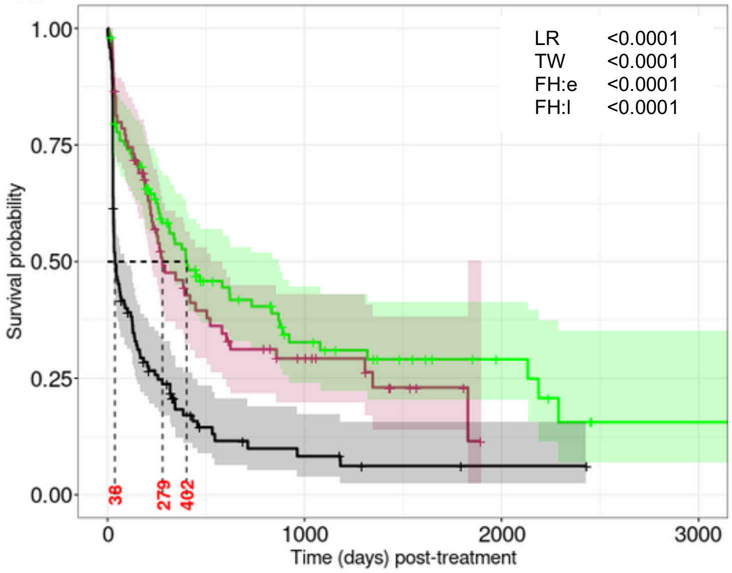


**Number at risk (number censored)**

| | 0 | 1000 | 2000 | 3000 |
|---|---|---|---|---|
| | 109 (0) | 20 (25) | 7 (36) | 1 (39) |
| | 75 (0) | 13 (14) | 0 (24) | 0 (24) |
| | 119 (0) | 5 (13) | 1 (16) | 0 (17) |

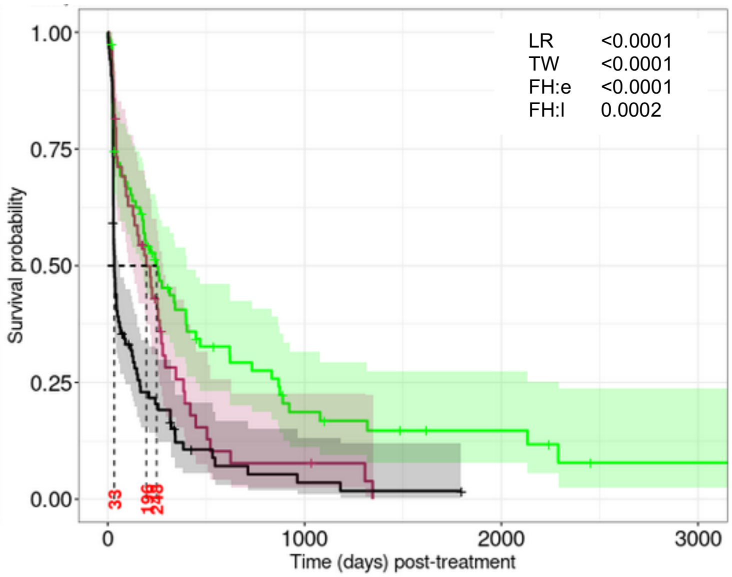


**Number at risk (number censored)**

| | 0 | 1000 | 2000 | 3000 |
|---|---|---|---|---|
| | 76 (0) | 10 (10) | 5 (13) | 1 (15) |
| | 49 (0) | 3 (5) | 0 (6) | 0 (6) |
| | 86 (0) | 2 (6) | 0 (7) | 0 (7) |

**C. $RM_{G,III\text{-}EFS}$ censoring allo-HCT (left) and only within allo-HCT (right) recipients.**

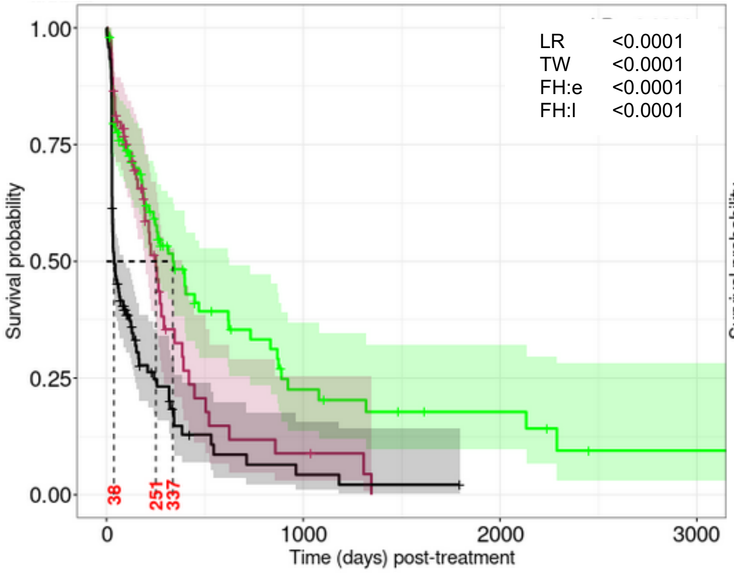


**Number at risk (number censored)**

| | 0 | 1000 | 2000 | 3000 |
|---|---|---|---|---|
| | 109 (0) | 10 (40) | 5 (43) | 1 (45) |
| | 75 (0) | 3 (28) | 0 (29) | 0 (29) |
| | 119 (0) | 2 (24) | 0 (25) | 0 (25) |

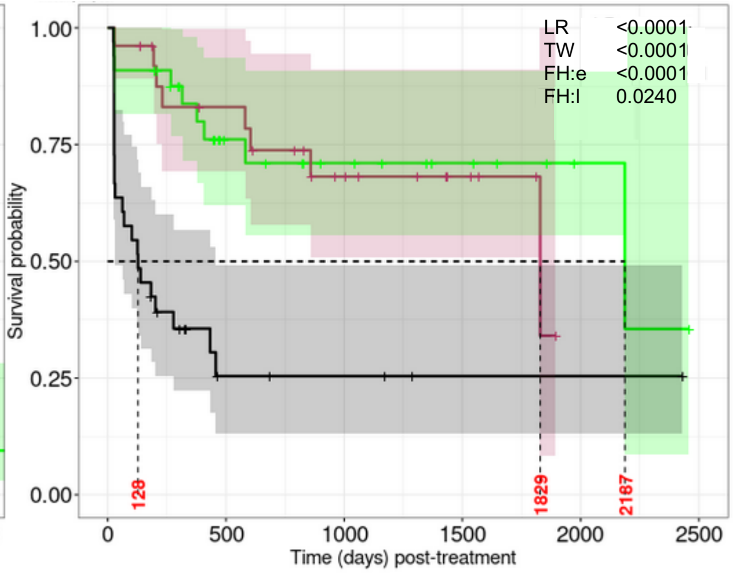


**Number at risk (number censored)**

| | 0 | 500 | 1000 | 1500 | 2000 | 2500 |
|---|---|---|---|---|---|---|
| | 33 (0) | 15 (11) | 10 (15) | 6 (19) | 2 (23) | 0 (24) |
| | 26 (0) | 18 (4) | 10 (9) | 5 (14) | 0 (18) | 0 (18) |
| | 33 (0) | 4 (6) | 3 (7) | 1 (9) | 1 (9) | 0 (10) |

**Supplemental Figure 12. Event free survival (EFS) based on the genetic features specific risk model using Rule-IV ($RM_{G,IV-EFS}$).**

**A. Feature identification strategy and $RM_{G,IV-EFS}$ features.**

| Category | Genetic features | Frequency† | Frequency¶ |
|---|---|---|---|
| **Favorable** | Any of ***Normal cytogenetics, Del5q, IDH1, IDH2, NPM1*** (in the absence of any Adverse features) | **73/211 (35%)** | **106/303 (35%)** |
| **Intermediate** | No Favorable or Adverse features, or ≥1 Favorable features with ≥1 Adverse features | **50/211 (24%)** | **77/303 (25%)** |
| **Adverse** | Any of ***JAK2, p53m, Del7q, t(9;11), Minus5, Minus17, Complex cytogenetics, Indeterminant cytogenetics, Chr 7 abn:loss, Chr 8 abn:loss, MECOM:rearrangement, KMT2A:rearrangement*** (in the absence of any Favorable features) | **88/211 (42%)** | **120/303 (40%)** |

† Counts (proportions) of patients after excluding allo-HCT
¶ Counts (proportions) of patients without excluding allo-HCT

**Supplemental Figure 12, cont'd**

**B. $RM_{G,IV\text{-}EFS}$ with respect to the FAS (left) and set excluding allo-HCT (right) patients.**

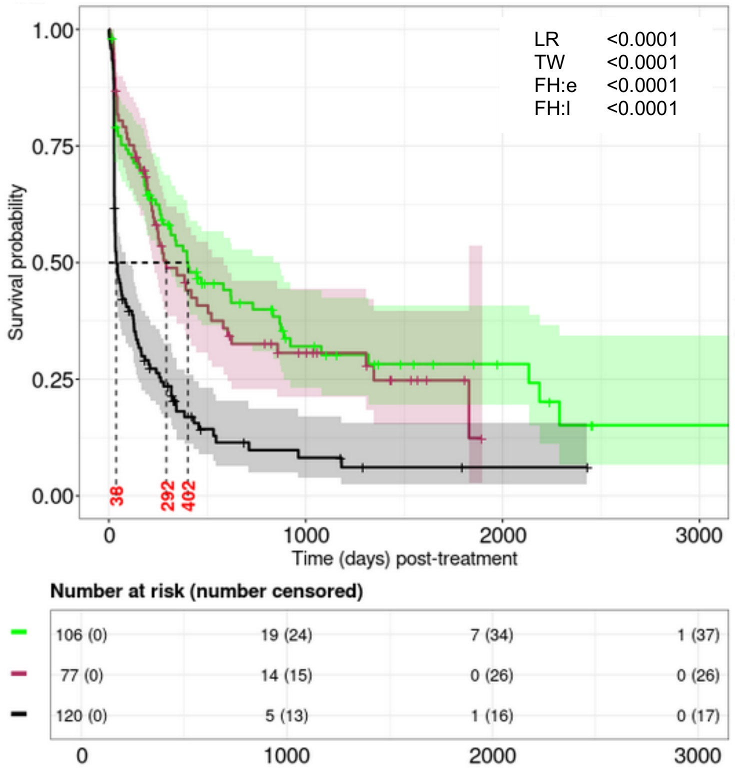


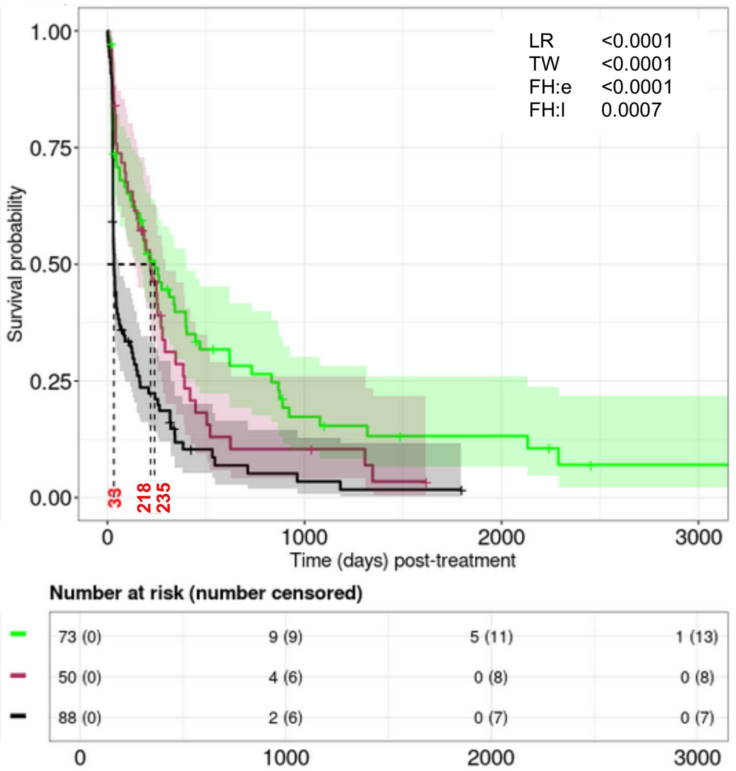


**C. $RM_{G,IV\text{-}EFS}$ censoring allo-HCT (left) and only within allo-HCT (right) recipients.**

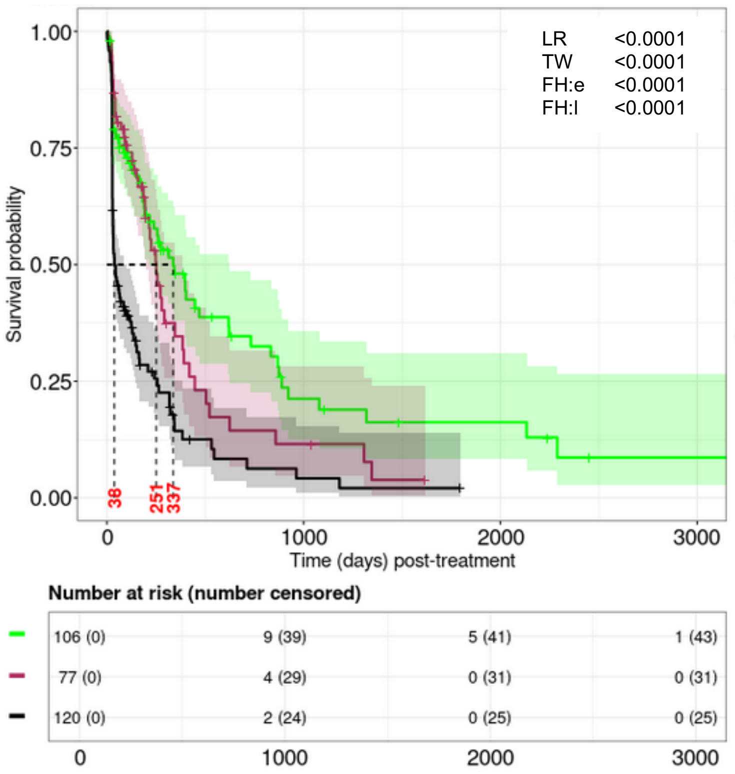


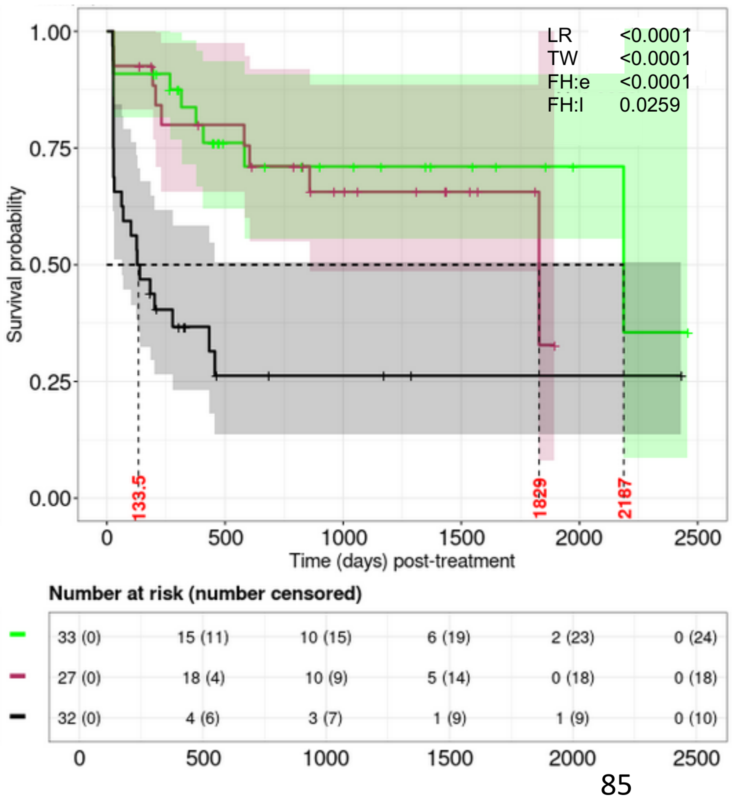

**Supplemental Figure 13. ELN22 and comparison with respect to $RM_{G,II\text{-}EFS}$ for event free survival (EFS).**

#### A. ELN22 on EFS excluding allo-HCT patients.

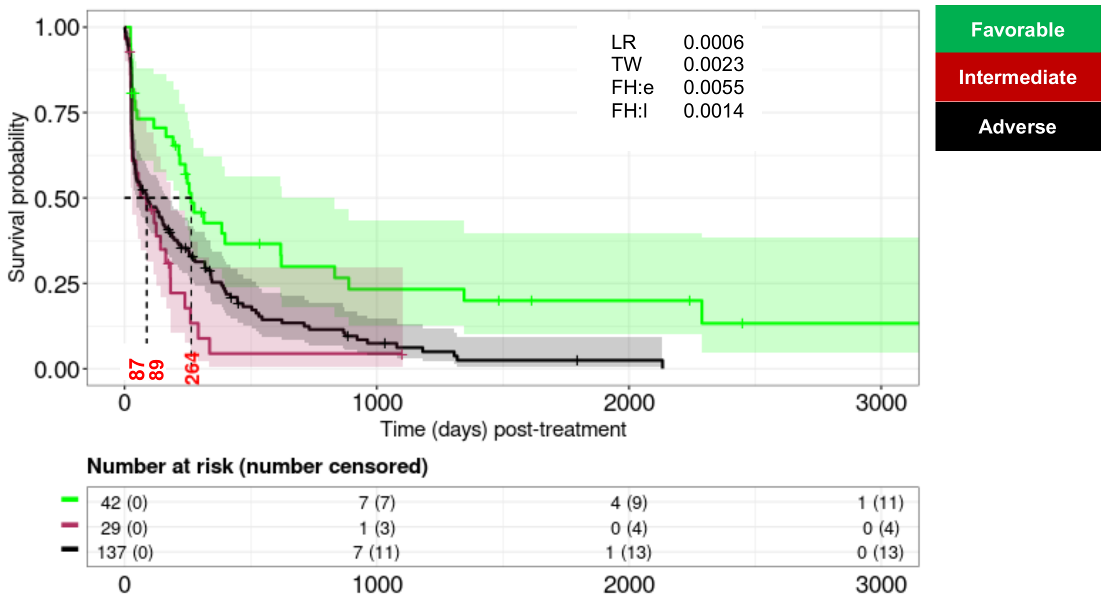


#### B. ELN22 on EFS with respect to the FAS.

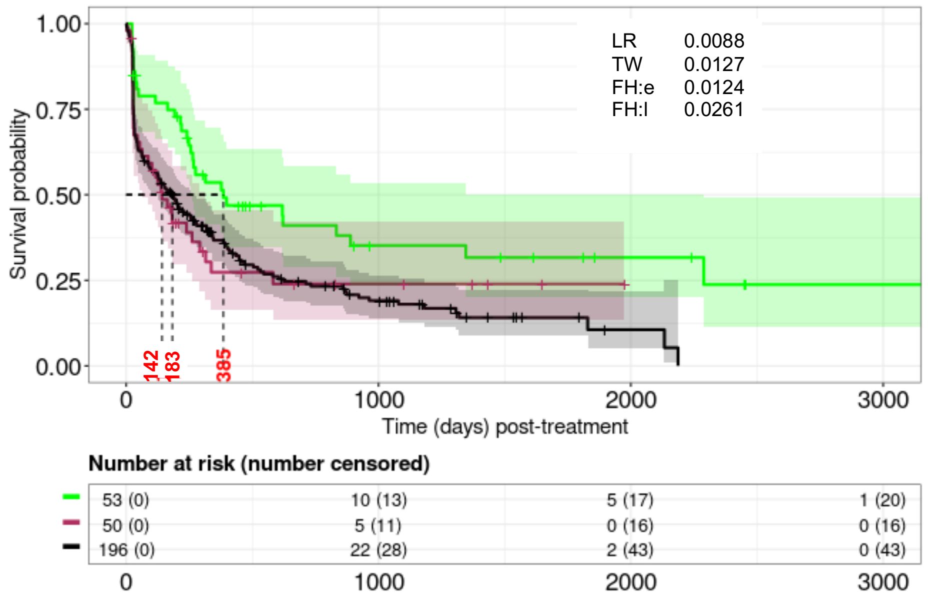

**Supplemental Figure 13, cont'd**

**C. Pairwise comparison between $RM_{G,II\text{-}EFS}$ and ELN22, excluding allo-HCT recipients.**

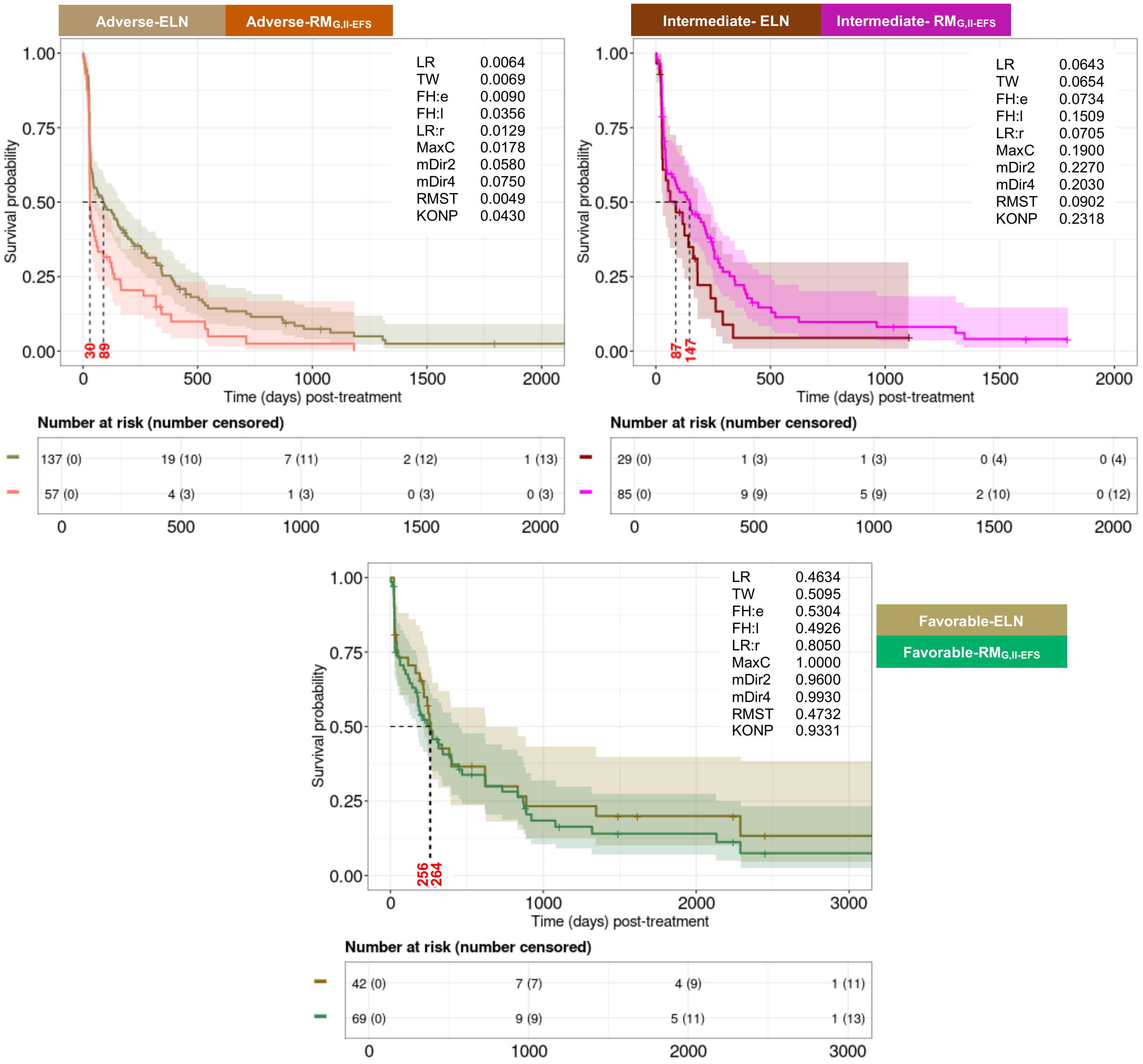


**Supplemental Figure 13, cont'd**

### D. Agreements between $RM_{G,II\text{-}EFS}$ and ELN22 risk models based on the FAS

| | Favorable | Intermediate | Adverse |
|---|---|---|---|
| [a]Fleiss kappa (*P* values) | **0.24 (<0.001)** | **-0.26 (0.090)** | **-0.02 (0.752)** |

**Remarks:**

**The higher positive value (i.e., close to 1) means more agreement**
**The lower negative value (i.e., close to -1) means less agreement**
**Value close to 0 means agreement is no better than obtained by chance**

[a]Analytical set to enumerate this conformity metric is comprised of the patients for who ELN22 was observed in the CU.

### E. Predictive evaluation of ELN22 and $RM_{G,II\text{-}EFS}$.

| [a]Model | M-iAUC ($25^{th}$,$75^{th}$) over $t$ | M-cAUC ($25^{th}$,$75^{th}$) over $t$ | iBrier ($25^{th}$,$75^{th}$) over $t$ |
|---|---|---|---|
| **ELN22** | 0.54 [0.51.58] | 0.56 [0.50,0.62] | 0.18 [0.14,0.22] |
| **$RM_{G,II\text{-}EFS}$** | 0.60 [0.55,0.64] (<0.001) | 0.65 [0.58,0.72] (<0.001) | 0.16 [0.15,0.22] |

- [a]Penalized CoxPH model adjusted by age, gender, race, and either ELN22 or $RM_{G,II\text{-}EFS}$.
- Reported in parenthesis are the median of P-values (over 15-fold CV) testing the null hypothesis of inferiority of distributions of survival metrics of the $RM_{G,II\text{-}EFS}$ relative to that of the ELN22.

**Supplemental Figure 14. Event free survival (EFS) based on the genetic-plus-phenotypic features specific risk model using Rule-I ($RM_{GP,I\text{-}EFS}$).**

**A. Feature identification strategy and $RM_{GP,I\text{-}EFS}$ features.**

| Category | Genetic plus phenotypic features | Frequency[†] | Frequency[¶] |
|---|---|---|---|
| **Favorable** | Any of ***CD10, CD14, CD15, CD19, BCOR, Normal cytogenetics, Good risk cytogenetics, Monosomal karyotype, Del5q, MECOM:normal, KMT2A:normal, IDH1, IDH2, NPM1*** (in the absence of any Adverse features) | **1/211 (0.5%)** | **1/303 (0.3%)** |
| **Intermediate** | No Favorable or Adverse features, or $\geq$1 Favorable features with $\geq$1 Adverse features | **191/211 (91%)** | **274/303 (90%)** |
| **Adverse** | Any of ***CD5, CD16, CD22, CD34, CD36, CD56, CD71, CD11B, KRAS, KMT2A, PTPN11, JAK2, p53m, Inv(3), t(9;11), Del7q, Minus5, Minus17, Complex cytogenetics, Indeterminant cytogenetics, Intermediate cytogenetics, Chr 7 abn:loss, Chr 8 abn:loss, RARA:loss, MECOM:rearrangement, KMT2A:rearrangement*** (in the absence of any Favorable features) | **19/211 (9%)** | **28/303 (9%)** |

[†] Counts (proportions) of patients after excluding allo-HCT
[¶] Counts (proportions) of patients without excluding allo-HCT

**Supplemental Figure 14, cont'd**

**B. $RM_{GP,I\text{-}EFS}$ with respect to the FAS (left) and set excluding allo-HCT (right) patients.**

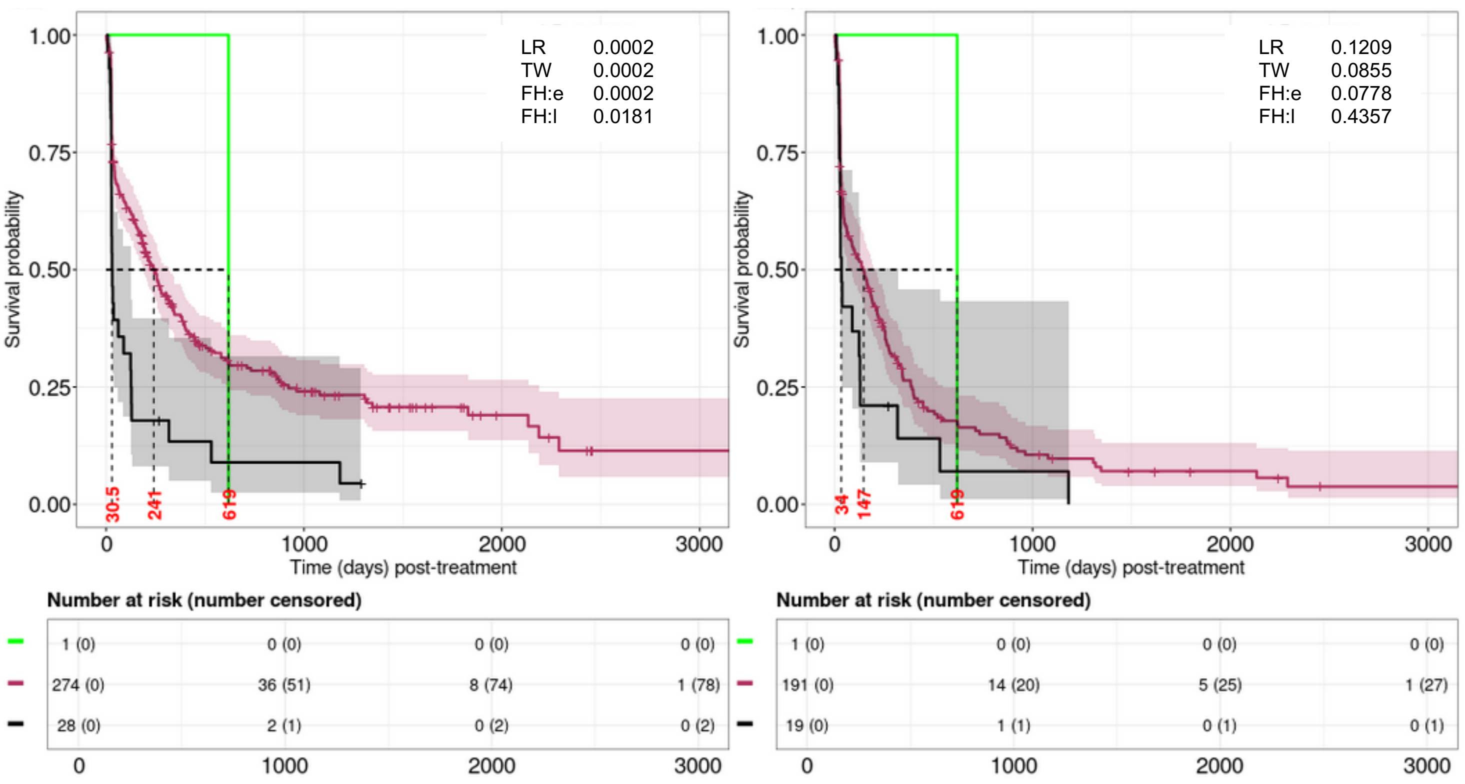


**C. $RM_{GP,I\text{-}EFS}$ censoring allo-HCT (left) and only within allo-HCT (right) recipients.**

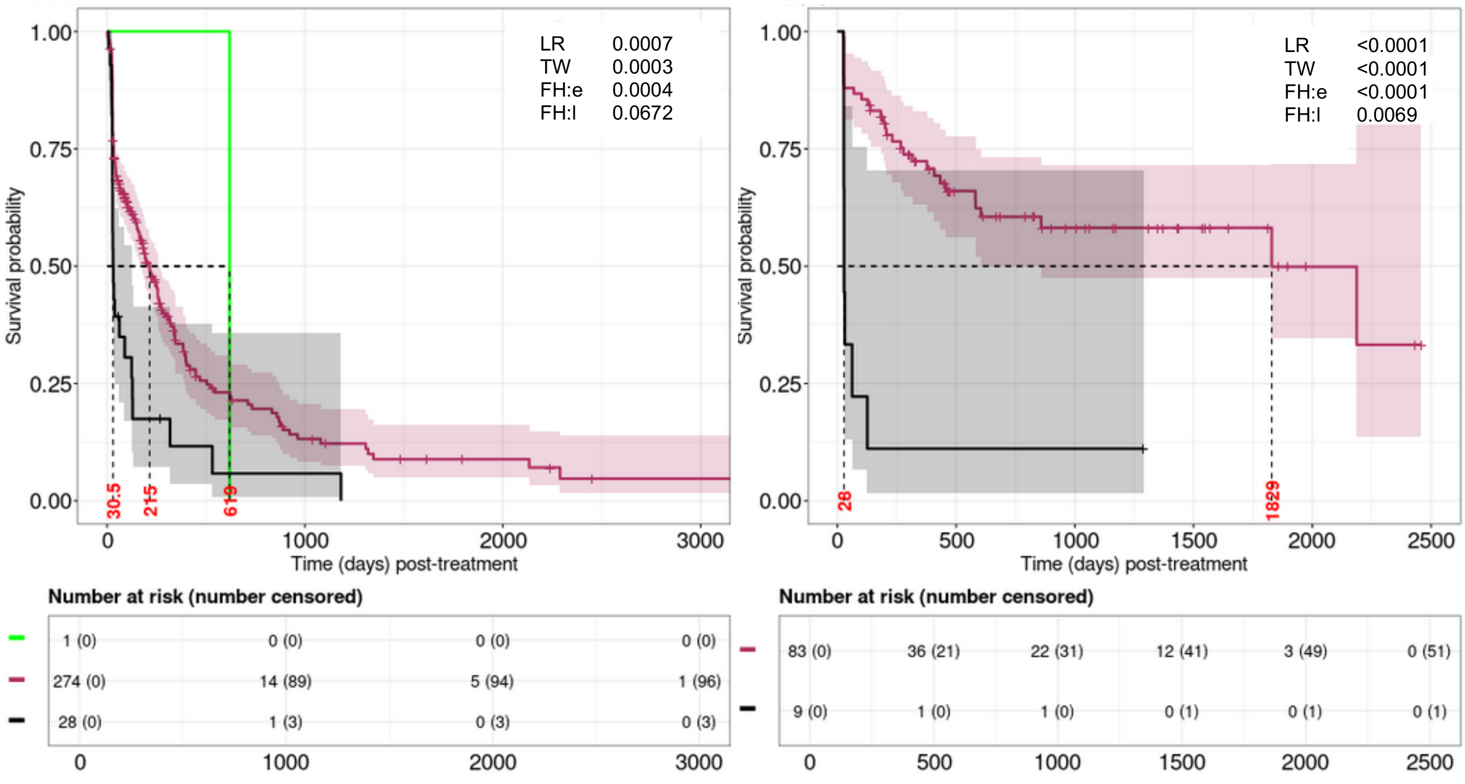

**Supplemental Figure 15. Event free survival (EFS) based on the genetic-plus-phenotypic features specific risk model using Rule-II ($RM_{GP,II-EFS}$).**

**A. Feature identification strategy and $RM_{GP,II-EFS}$ features.**

| Category | Genetic plus phenotypic features | Frequency[†] | Frequency[¶] |
|---|---|---|---|
| **Favorable** | Any of ***CD10, CD14, CD15, CD19, BCOR, Normal cytogenetics, Monosomal karyotype, Del5q, IDH1, IDH2, NPM1*** (in the absence of any Adverse features) | **30/211 (14%)** | **54/303 (18%)** |
| **Intermediate** | No Favorable or Adverse features, or $\geq$1 Favorable features with $\geq$1 Adverse features | **133/211 (63%)** | **186/303 (61%)** |
| **Adverse** | Any of ***CD5, CD22, CD36, CD56, CD11B, KRAS, KMT2A, PTPN11, JAK2, p53m, Del7q, t(9;11), Minus5, Minus17, Complex cytogenetics, Indeterminant cytogenetics, Chr 7 abn:loss, Chr 8 abn:loss, RARA:loss, MECOM:rearrangement, KMT2A:rearrangement*** (in the absence of any Favorable features) | **48/211 (23%)** | **63/303 (21%)** |

[†] Counts (proportions) of patients after excluding allo-HCT
[¶] Counts (proportions) of patients without excluding allo-HCT

**Supplemental Figure 15, cont'd**

**B. $RM_{GP,II\text{-}EFS}$ with respect to the FAS (left) and set excluding allo-HCT (right) patients.**

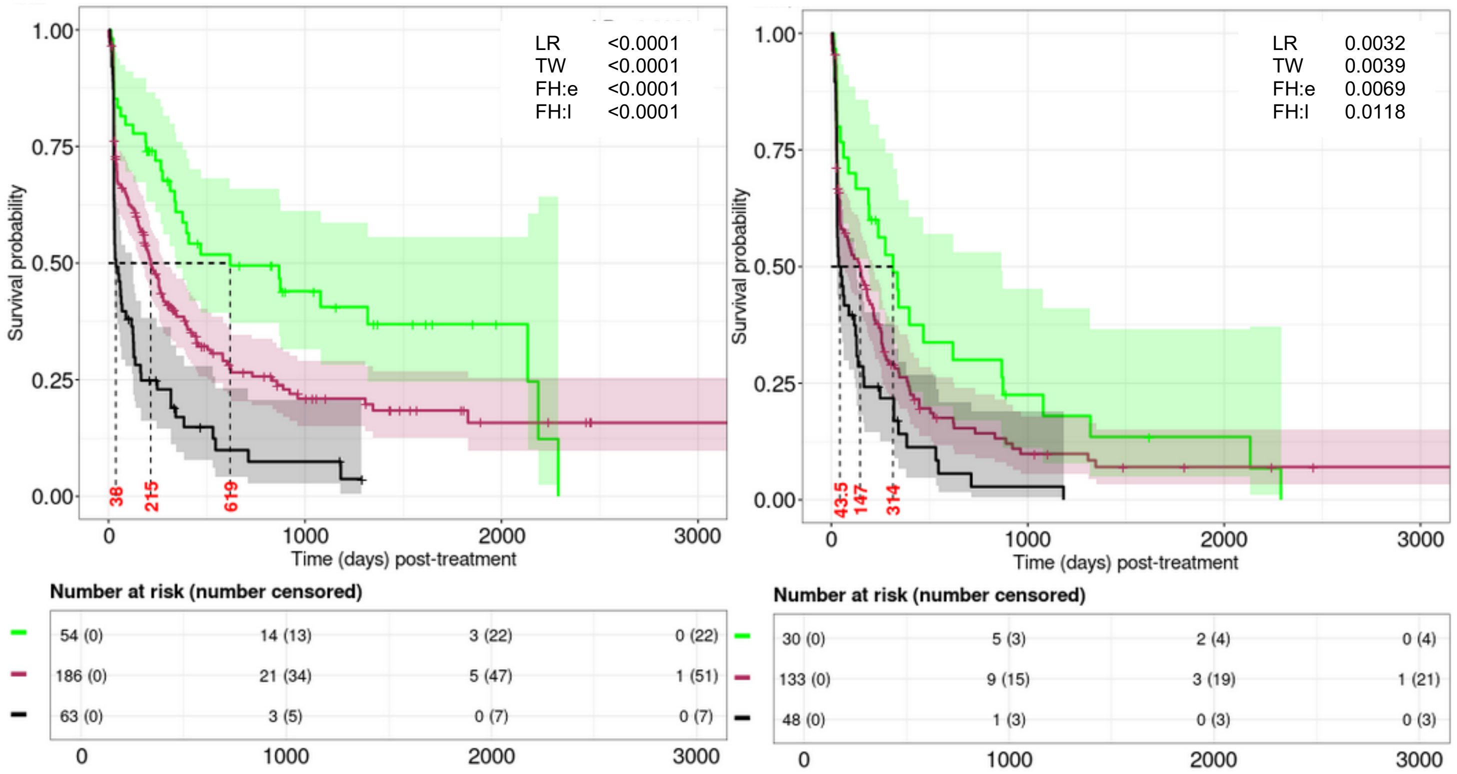


**C. $RM_{GP,II\text{-}EFS}$ censoring allo-HCT (left) and only within allo-HCT (right) recipients.**

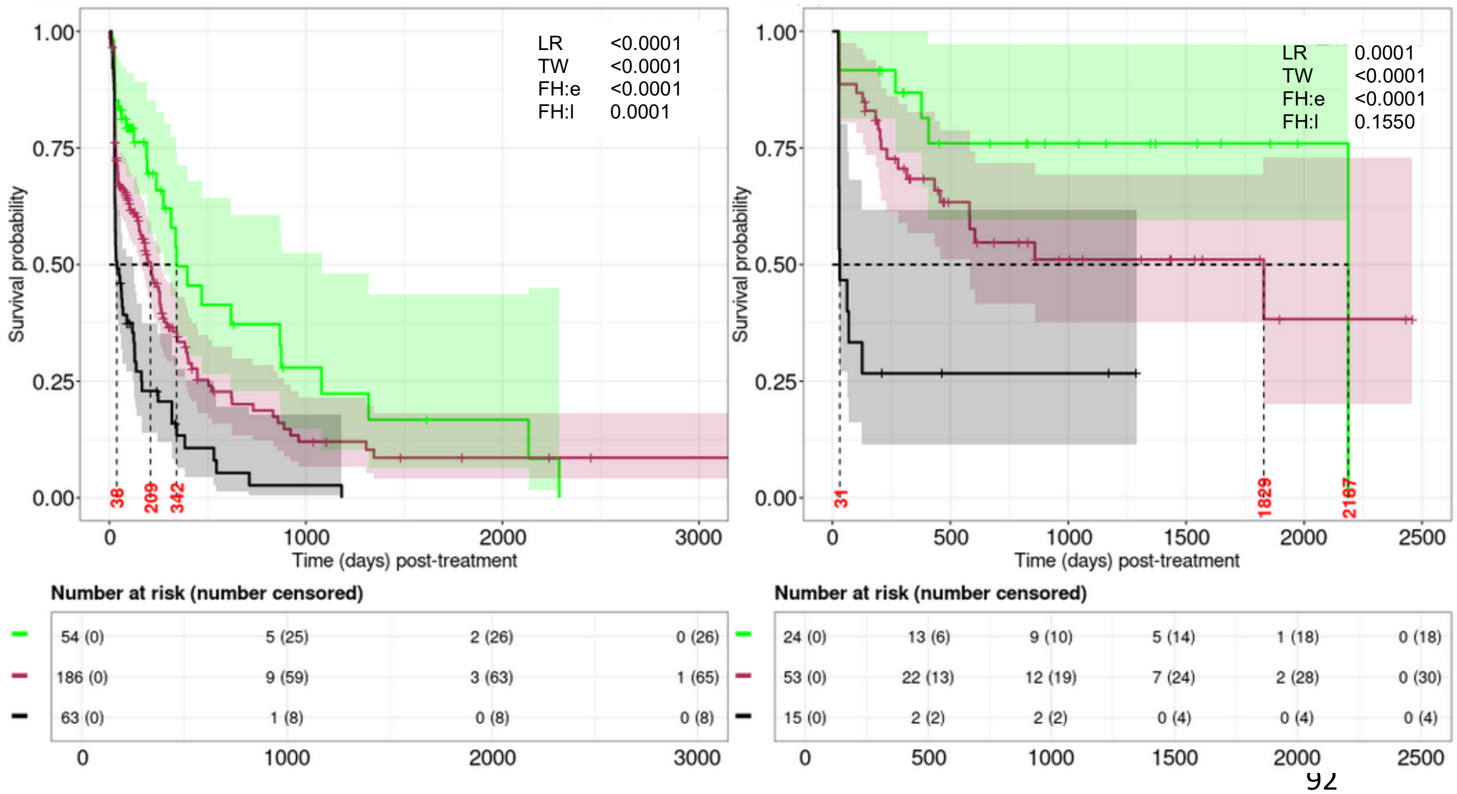

**Supplemental Figure 15, cont'd**

**F. Agreement between ELN22 and $RM_{G,II\text{-}EFS}$.**

| | Favorable | Intermediate | Adverse |
|---|---|---|---|
| **Fleiss kappa (*P* values)** | **0.24 (<0.001)** | **-0.26 (0.090)** | **-0.02 (0.752)** |

**Remarks:**

- The higher positive value (i.e., close to 1) means more agreement
- The lower negative value (i.e., close to -1) means less agreement
- Value close to 0 means agreement is no better than obtained by chance

**G. Predictive evaluation of ELN22 and $RM_{G,II\text{-}EFS}$.**

| [a]Model | M-iAUC ($25^{th}$,$75^{th}$) over *t* | M-cAUC ($25^{th}$,$75^{th}$) over *t* | iBrier ($25^{th}$,$75^{th}$) over *t* |
|---|---|---|---|
| **ELN22** | 0.54 [0.51.58] | 0.56 [0.50,0.62] | 0.18 [0.14,0.22] |
| **$RM_{G,II\text{-}EFS}$** | 0.60 [0.55,0.64] (<0.001) | 0.65 [0.58,0.72] (<0.001) | 0.16 [0.15,0.22] |

- [a]Penalized CoxPH model adjusted by age, gender, race, and either ELN22 or $RM_{G,II\text{-}EFS}$.
- Reported in parenthesis are the median of P-values (over 15-fold CV) testing the null hypothesis of inferiority of distributions of survival metrics of the $RM_{G,II\text{-}EFS}$ relative to that of the ELN22.

**Supplemental Figure 16. Event free survival (EFS) based on the genetic-plus-phenotypic features specific risk model using Rule-III ($RM_{GP,III\text{-}EFS}$).**

**A. $RM_{GP,III\text{-}EFS}$ features**

| Category | Genetic features | Frequency† | Frequency¶ |
|---|---|---|---|
| **Favorable** | Any of ***CD10, Normal cytogenetics, Good risk cytogenetics, Del5q, IDH1, IDH2, NPM1*** (in the absence of any Adverse features) | **73/211 (35%)** | **106/303 (35%)** |
| **Intermediate** | No Favorable or Adverse features, or ≥1 Favorable features with ≥1 Adverse features | **57/211 (27%)** | **83/303 (27%)** |
| **Adverse** | Any of ***CD16, CD71, JAK2, p53m, Inv3, Del7q, t(9;11), Minus5, Minus17, Complex cytogenetics, Indeterminant cytogenetics, Chr 7 abn:loss, Chr 8 abn:loss, MECOM:rearrangement, KMT2A:rearrangement*** (in the absence of any Favorable features) | **81/211 (38%)** | **114/303 (38%)** |

† Counts (proportions) of patients after excluding allo-HCT
¶ Counts (proportions) of patients without excluding allo-HCT

**Supplemental Figure 16, cont'd**

**B. $RM_{GP,III\text{-}EFS}$ with respect to the FAS (left) and excluding allo-HCT (right) patients.**

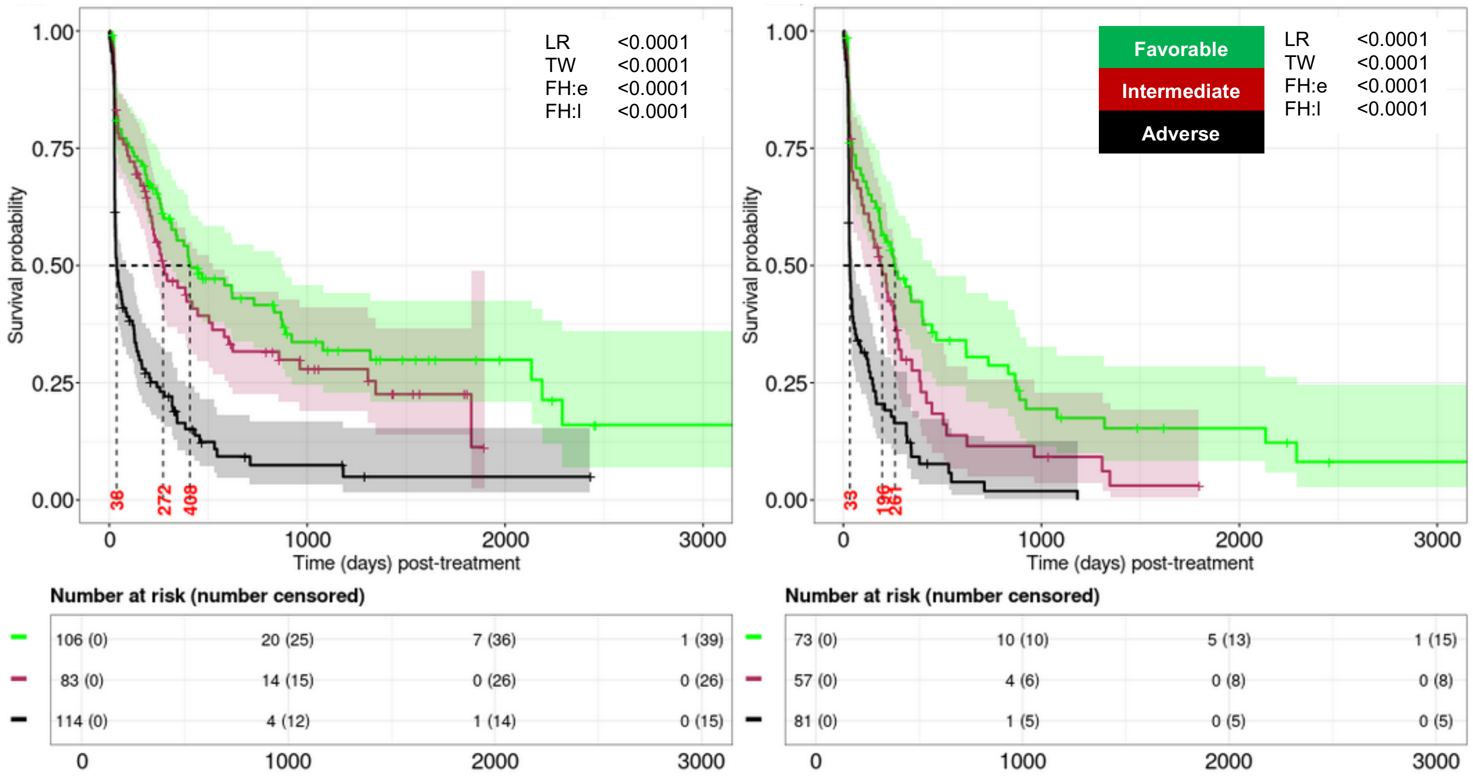


**D. $RM_{GP,III\text{-}EFS}$ censoring allo-HCT (left) and only within allo-HCT (right) recipients.**

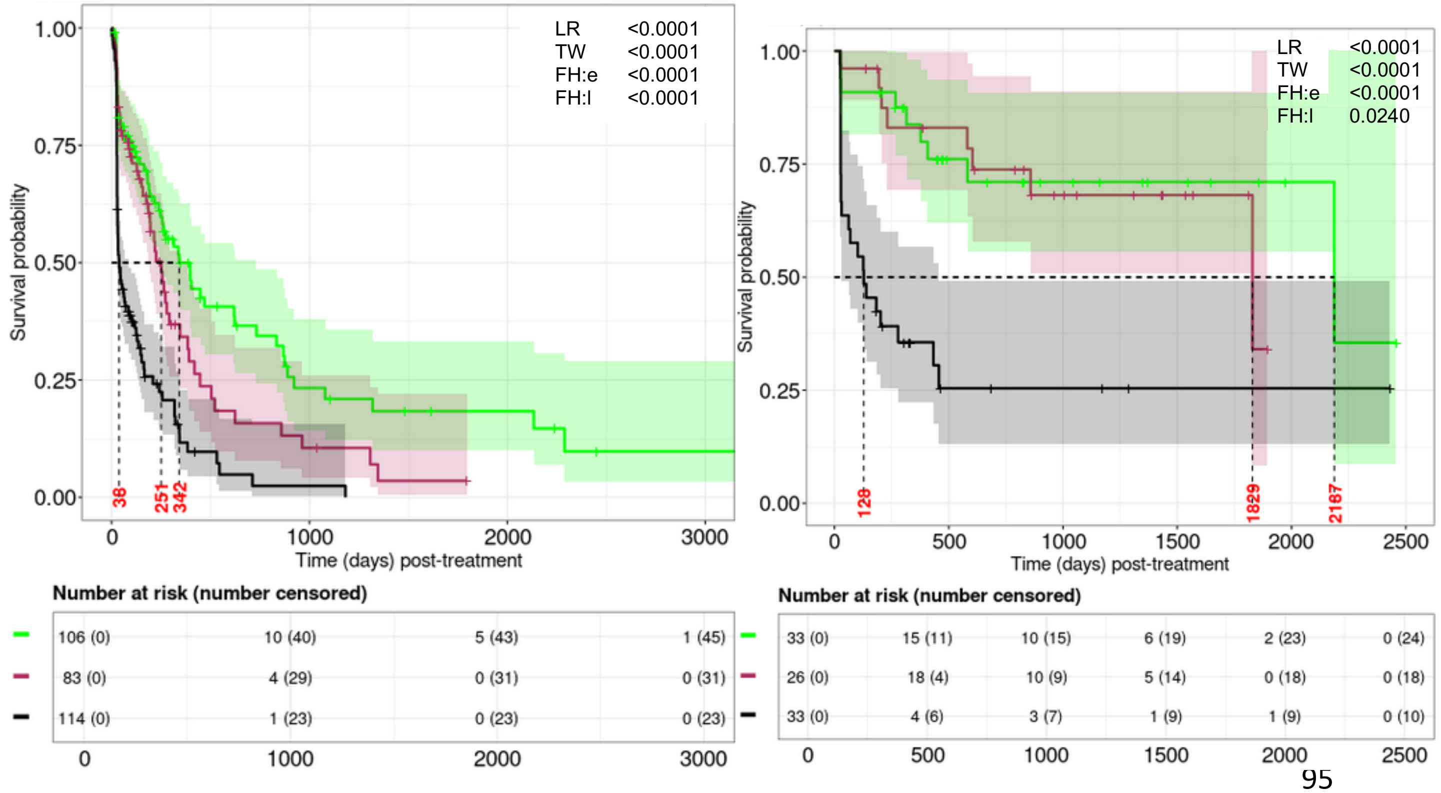

**Supplemental Figure 17. Event free survival (EFS) based on the genetic-plus-phenotypic features specific risk model using Rule-IV ($RM_{GP,IV\text{-}EFS}$).**

A. **Feature identification strategy and $RM_{GP,IV\text{-}EFS}$ features.**

| Category | Genetic features | Frequency† | Frequency¶ |
|---|---|---|---|
| **Favorable** (green) | Any of ***CD10, Normal cytogenetics , Del5q, IDH1, IDH2, NPM1*** (in the absence of any Adverse features) | **73/211 (35%)** | **106/303 (35%)** |
| **Intermediate** (red) | No Favorable or Adverse features, or ≥1 Favorable features with ≥1 Adverse features | **55/211 (26%)** | **82/303 (27%)** |
| **Adverse** (black) | Any of ***JAK2, p53m, Del7q, t(9;11), Minus5, Minus17, Complex cytogenetics, Indeterminant cytogenetics, Chr 7 abn:loss, Chr 8 abn:loss, MECOM:rearrangement, KMT2A:rearrangement*** (in the absence of any Favorable features) | **83/211 (39%)** | **115/303 (38%)** |

† Counts (proportions) of patients after excluding allo-HCT
¶ Counts (proportions) of patients without excluding allo-HCT

**Supplemental Figure 17, cont'd**

**B. RM$_{GP,IV\text{-}EFS}$ with respect to the FAS (left) and excluding allo-HCT (right) patients.**

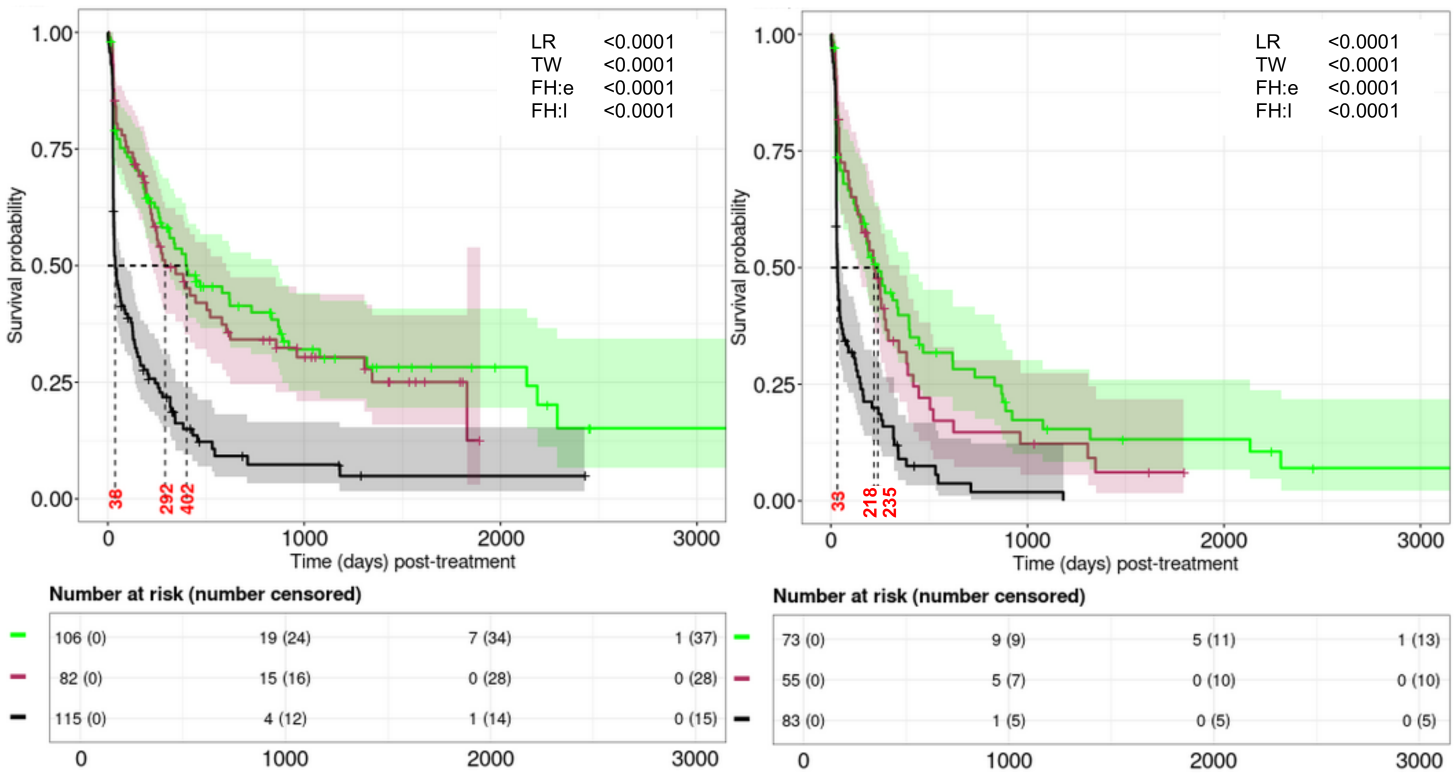


**C. RM$_{GP,IV\text{-}EFS}$ censoring allo-HCT (left) and only within allo-HCT (right) recipients.**

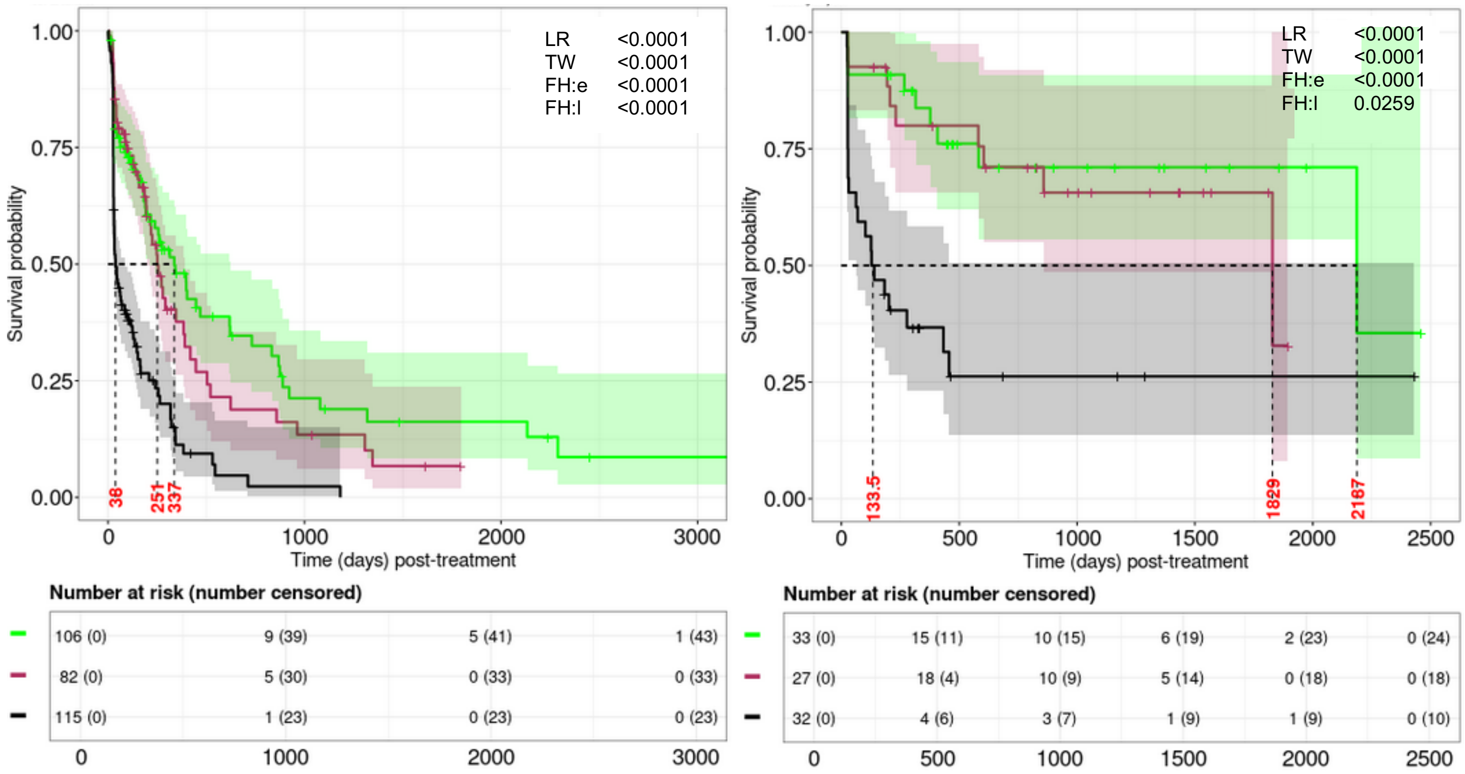

**Supplemental Figure 18. Subject-level risk stratifications of 316 AML patients by different risk models as described Supplemental Table 4.**

**A.** Covariate-level classification: Missing data adjusted as a separate category and allo-HCT recipients were not censored (Supplemental Table 4B-4E with (M1) and (M7)).

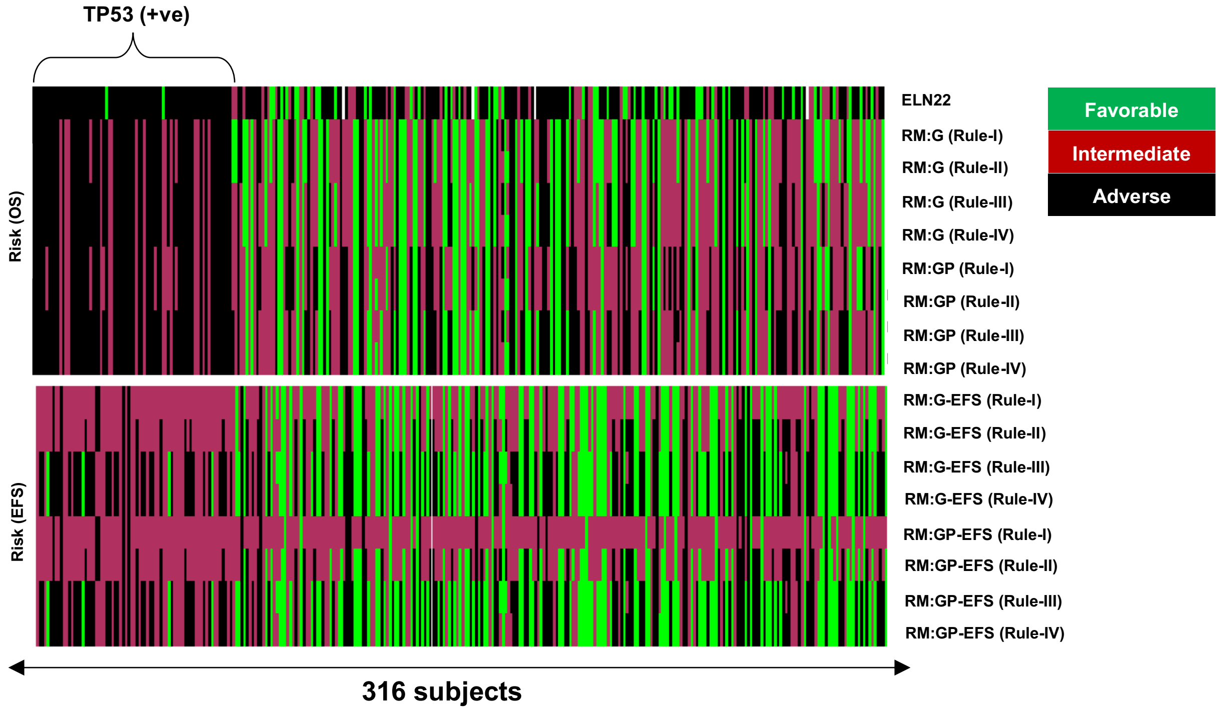


**B.** Covariate-level classification: Complete cases with weights and allo-HCT recipients were not censored (Supplemental Table 4B-4E with (M3) and (M9)).

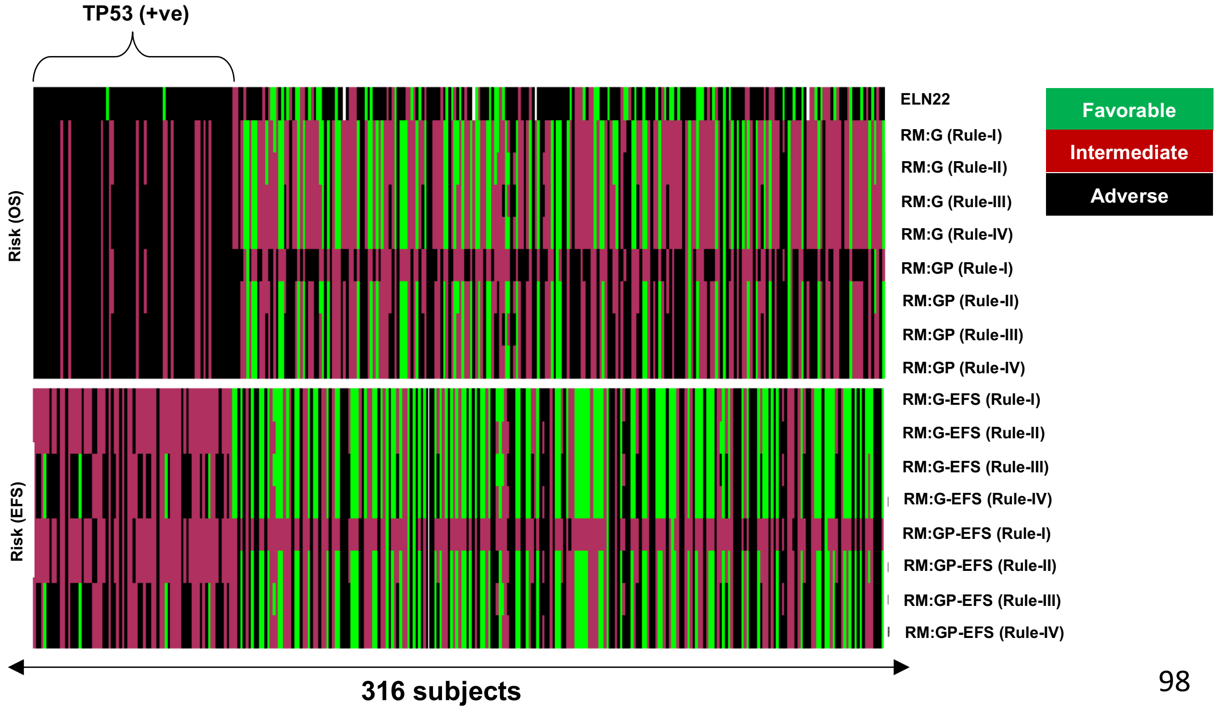

**Supplemental Figure 18, cont'd**

**C.** Covariate-level classification: Imputation of missing elements and allo-HCT recipients were not censored (Supplemental Table 4B-4E with (M5) and (M11)).

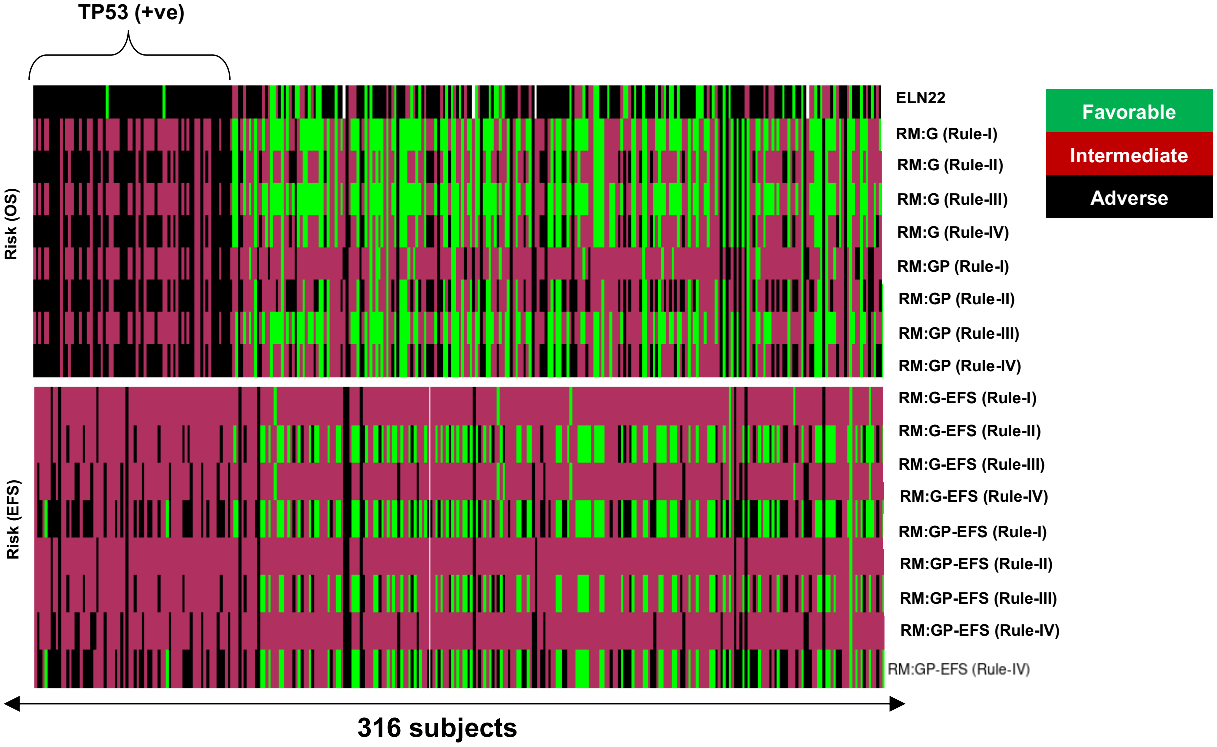


**D.** Missing data adjusted as a separate category and allo-HCT recipients were censored (Supplemental Table 4B-4E with (M2) and (M8)).

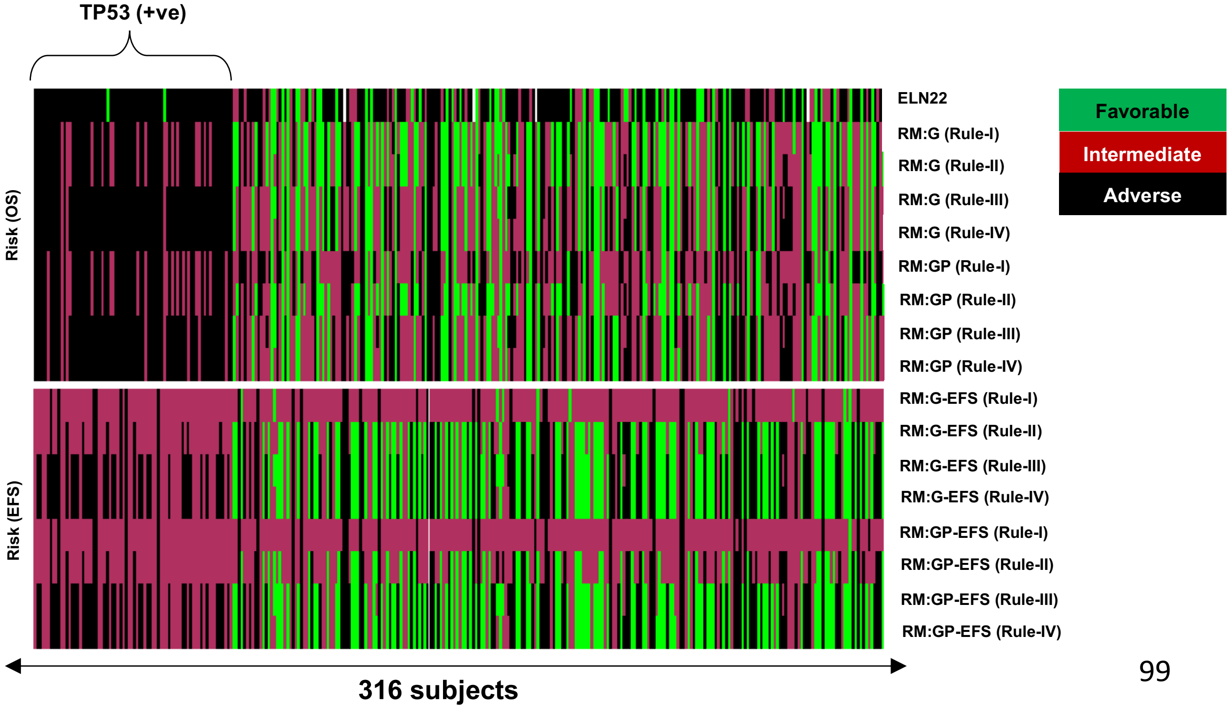

**Supplemental Figure 18, cont'd**

**E.** Covariate-level classification: Complete cases with weights and allo-HCT recipients were censored (Supplemental Table 4B-4E with (M4) and (M10)).

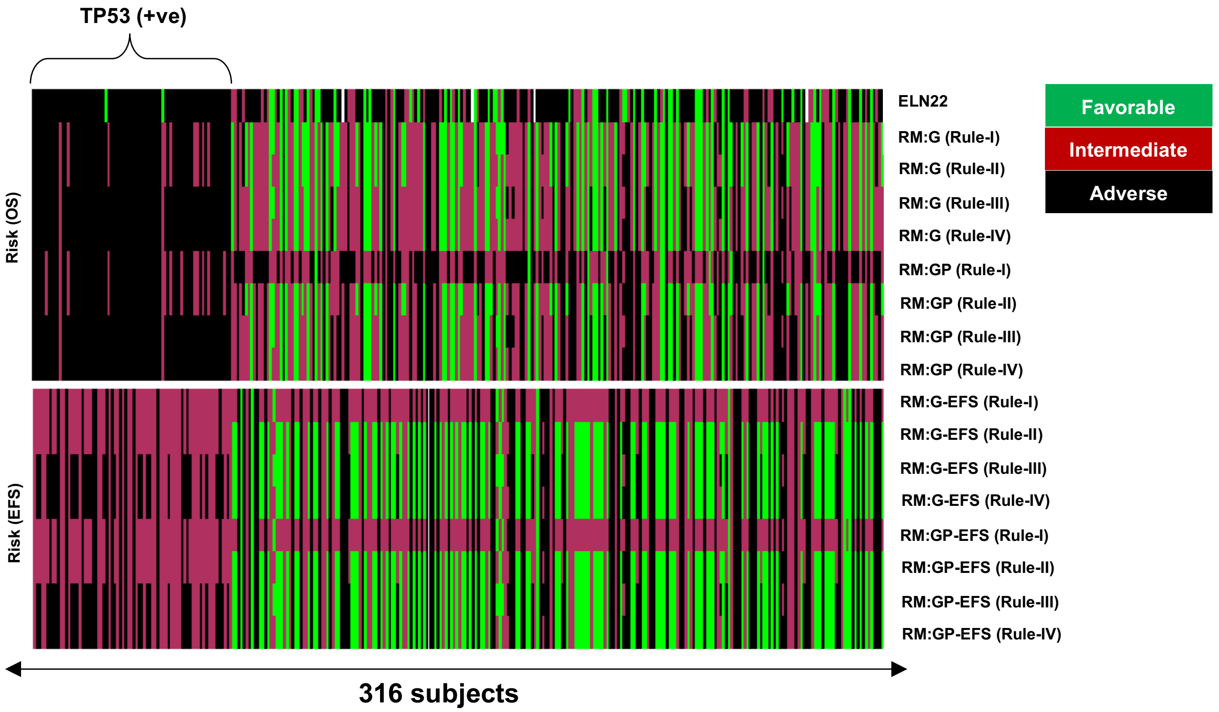


**F.** Covariate-level classification: Imputation of missing elements and allo-HCT recipients were censored (Supplemental Table 4B-4E with (M6) and (M12)).

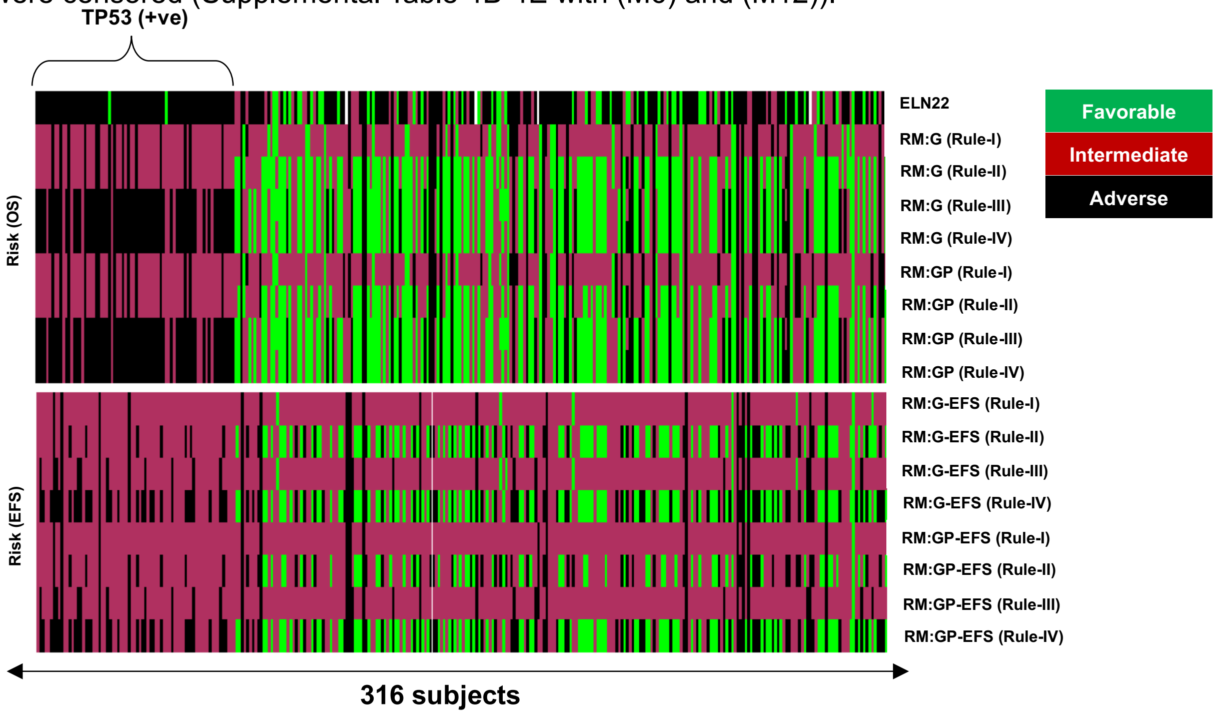

**Supplemental Figure 19. Evaluation of the ELN22 for overall survival (OS) using the RWC dataset.** Reported are the results for OS including allo-HCT patients (left panel) and excluding allo-HCT patients (right panel).

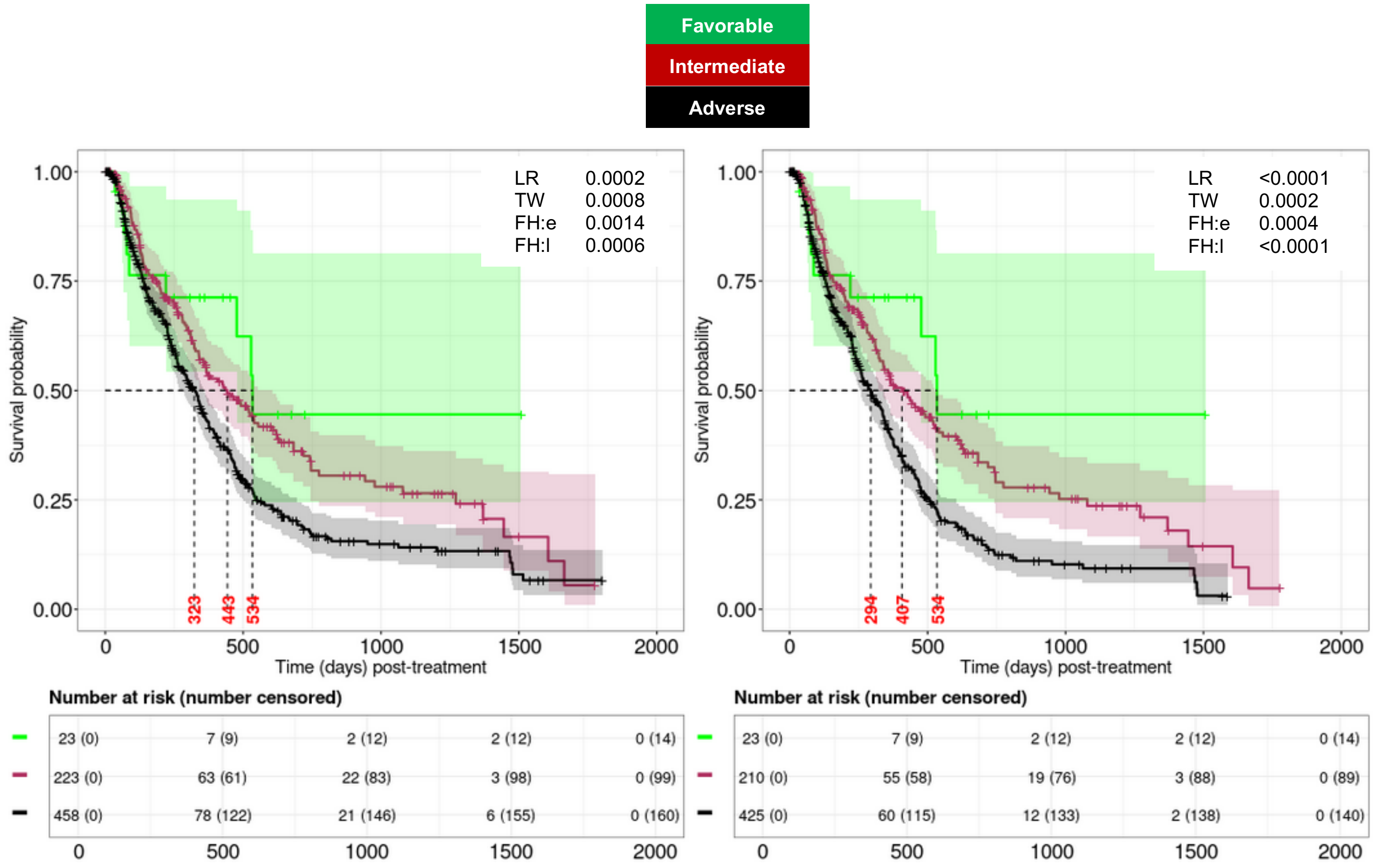

**Supplemental Figure 20. Contingency tables for risk models with respect to the real-world cohort (RWC) full analytical set (FAS).** Reported are the (top) confusion matrices with risk groups based on the RWC-RM$_{G\text{-}II}$ and ELN22 compared to their recorded outcomes (0 = censored to last known alive date (LKAD) or 1 = deceased) and (bottom) a contingency table with dot plots highlighting the proportion of deceased patients for different risk groups across the RWC-RM$_{G\text{-}II}$ and ELN22.

| RWC-RM$_{G\text{-}II}$ | Censored to LKAD | Deceased |
|---|---|---|
| Adverse ($n_{adv}$ = 285) | 76 (27%) | 209 (73%) |
| Intermediate ($n_{int}$ = 506) | 193 (38%) | 313 (62%) |
| Favorable ($n_{fav}$ = 180) | 94 (52%) | 86 (48%) |

| ELN22 | Censored to LKAD | Deceased |
|---|---|---|
| Adverse ($n_{adv}$ = 458) | 160 (35%) | 298 (65%) |
| Intermediate ($n_{int}$ = 223) | 99 (44%) | 124 (56%) |
| Favorable ($n_{fav}$ = 23) | 14 (61%) | 9 (39%) |
| Missing ($n_{na}$ = 267) | 90 (34%) | 177 (66%) |

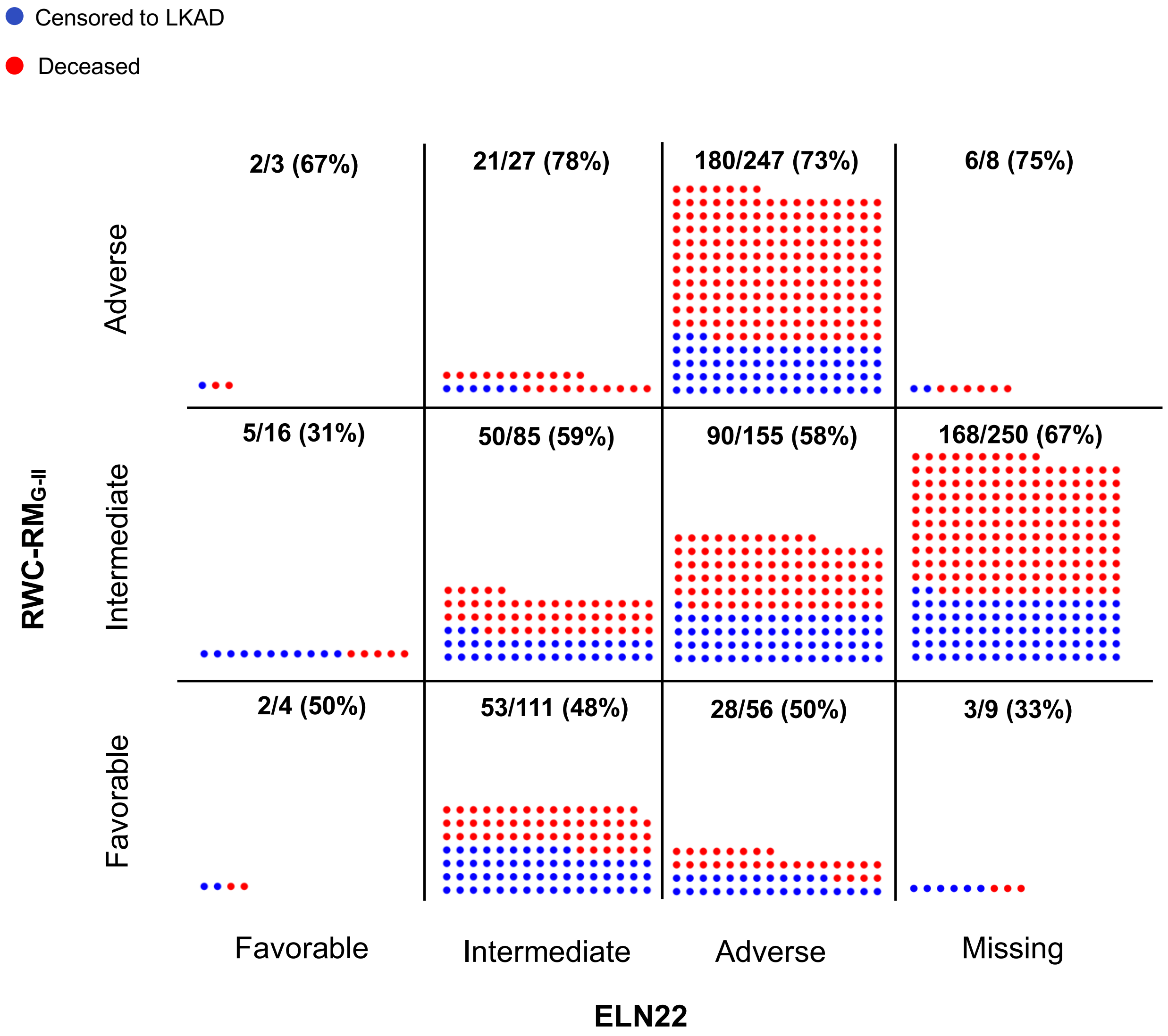


***Note:*** *Patients taking values 0 are censored at LKAD and the true outcomes for these censored patients are indeed unknown and their corresponding follow-up times vary. Therefore, the results need to be interpreted with caution.*

**Supplemental Figure 21. Contingency tables for risk models at 450-day.** Reported are the (top) confusion matrices with risk groups based on the RWC-RM$_{G\text{-}II}$ and ELN22 compared to their observed outcomes (0 = alive at 450-day or 1 = deceased before 450-day post-treatment) and (bottom) a contingency table with dot plots highlighting the proportion of deceased patients before 450-day for different risk groups across the RWC-RM$_{G\text{-}II}$ and ELN22.

| *RWC-RM$_{G\text{-}II}$ | Alive at 450d | Died before 450d |
|---|---|---|
| Adverse ($n_{adv}$ = 216) | 37 (17%) | 179 (83%) |
| Intermediate ($n_{int}$ = 381) | 148 (39%) | 233 (61%) |
| Favorable ($n_{fav}$ = 132) | 69 (52%) | 63 (48%) |

*242 patients were excluded as they were censored to LKA[illegible]

| *ELN22 | Alive at 450d | Died before 450d |
|---|---|---|
| Adverse ($n_{adv}$ = 343) | 103 (30%) | 240 (70%) |
| Intermediate ($n_{int}$ = 167) | 72 (43%) | 95 (57%) |
| Favorable ($n_{fav}$ = 15) | 9 (60%) | 6 (40%) |
| Missing ($n_{na}$ = 204) | 70 (34%) | 134 (66%) |

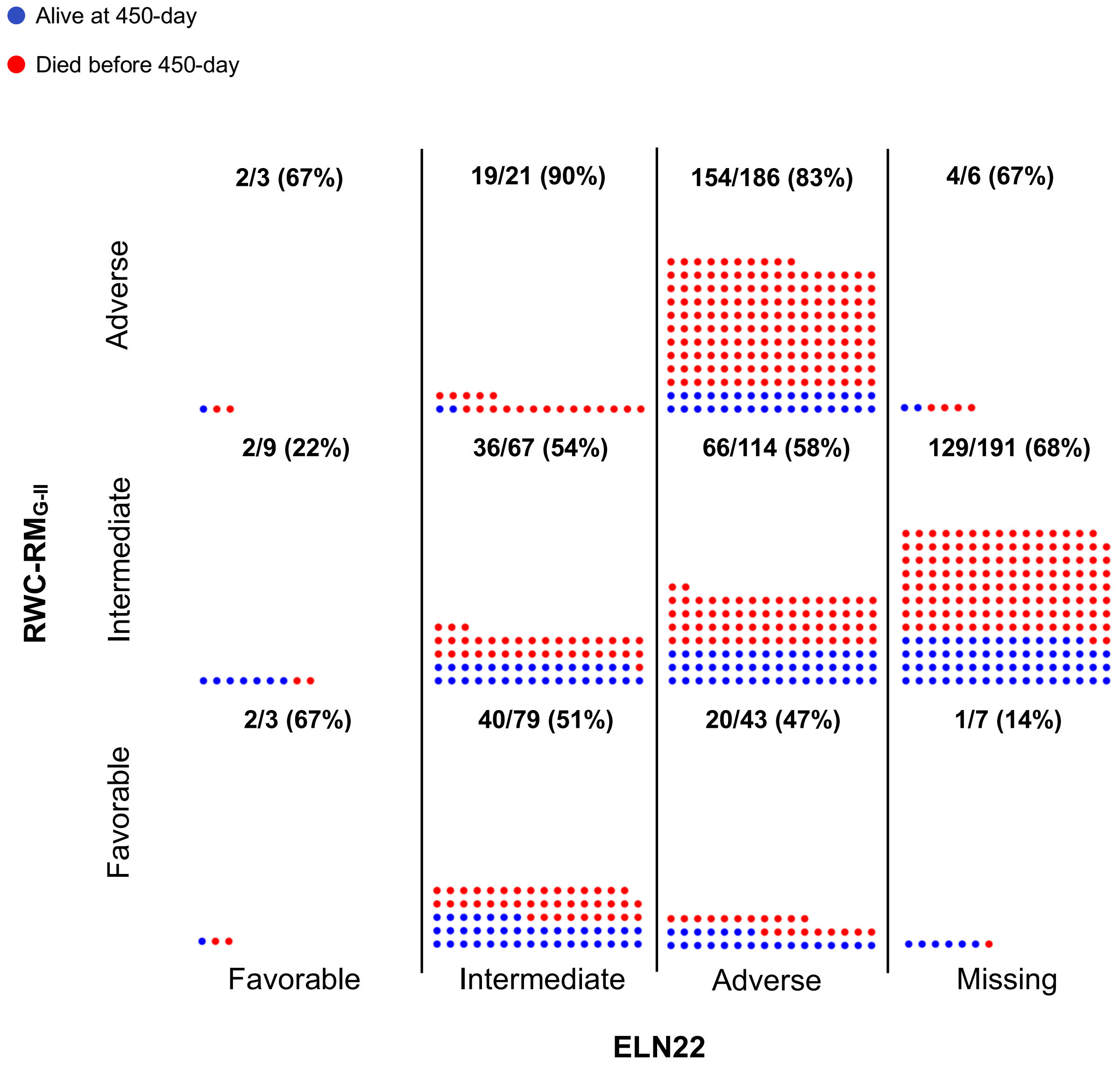


***Note:*** *While the above descriptive statistics facilitate direct outcome comparison at 450-day post-treatment, this approach reduces sample size by excluding 242 censored patients prior to 450-day. Therefore, the results need to be interpreted with caution.*

**Supplemental Figure 22. Evaluation of the proposed risk models (RMs) for overall survival (OS) using the RWC dataset.** Results are based on OS including allo-HCT patients (left panel) and excluding allo-HCT patients (right panel) with respect to A) RWC-$RM_{G,I}$; B) RWC-$RM_{G,III}$; C) RWC-$RM_{G,IV}$; D) RWC-$RM_{GP,I}$; E) RWC-$RM_{GP,II}$; F) RWC-$RM_{GP,III}$; and G) RWC-$RM_{GP,IV}$.

#### A. RWC-$RM_{G,I}$

| Category | $RM_{G,I}$ classification | [a]RWC-$RM_{G,I}$ classification |
|---|---|---|
| Favorable | Any of ***BCOR, DNMT3A, DDX41, SF3B1, KIT, Good risk cytogenetics, IDH1, IDH2, NPM1*** (in the absence of any Adverse features) | Any of ***DNMT3A, DDX41, KIT, Good risk cytogenetics, IDH1, IDH2, NPM1*** (in the absence of any Adverse features) |
| Intermediate | No Favorable or Adverse features, or $\geq$1 Favorable features with $\geq$1 Adverse features | No Favorable or Adverse features, or $\geq$1 Favorable features with $\geq$1 Adverse features |
| Adverse | Any of ***CSF3R, KRAS, KMT2A, MPL, PTPN11, SMC1A, JAK2, p53m, Del7q, Inv(3), t(9;11), Minus5, Minus17, Complex cytogenetics, Chr 7 abn:loss, Chr 8 abn:loss, DEK:loss, MECOM:gain, RARA:loss*** (in the absence of any Favorable features) | Any of ***CSF3R, KRAS, MPL, JAK2, p53m, Minus7, Inv(3), t(9;11), Minus5, Minus17, Complex cytogenetics, Chr 7 abn:loss, Chr 8 abn:loss, DEK:loss, MECOM:gain, RARA:loss*** (in the absence of any Favorable features) |

[a]Biomarkers such as *BCOR, SF3B1, SMC1A, KMT2A, PTPN11, Chr 8 abn:loss, DEK:loss, MECOM:gain,* and *RARA:loss* were unavailable in the RWC. *Chromosome 7 deletion* (i.e., *Minus 7*) was used as a proxy of *Chromosome 7q deletion* (i.e., *Del7Q*).

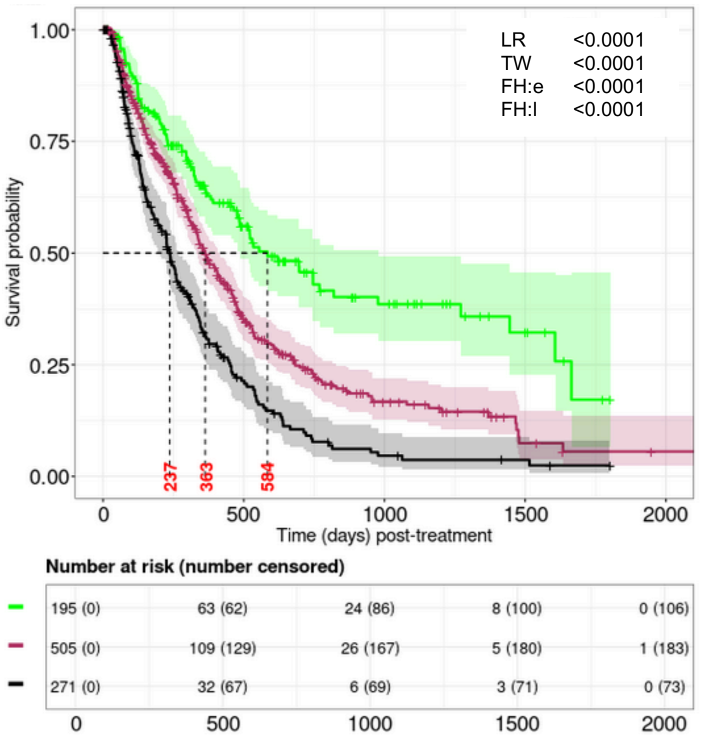


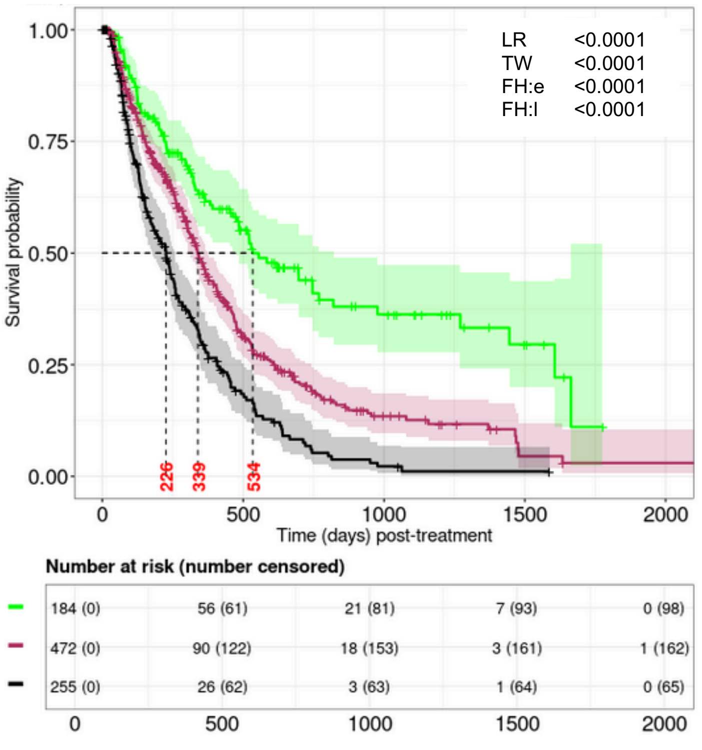

**Supplemental Figure 22, cont'd**

### B. RWC-RM$_{G,III}$

| Category | RM$_{G,III}$ classification | [a]RWC-RM$_{G,III}$ classification |
|---|---|---|
| Favorable | Any of ***DDX41, Cytogenetics good, IDH1, NPM1*** (in the absence of any Adverse features) | Any of ***DDX41, Cytogenetics good, IDH1, NPM1*** (in the absence of any Adverse features) |
| Intermediate | No Favorable or Adverse features, or >1 Favorable features with >1 Adverse features | No Favorable or Adverse features, or >1 Favorable features with >1 Adverse features |
| Adverse | Any of ***KRAS, KMT2A, MPL, PTPN11, SMC1A, JAK2, p53m, Inv(3), Del7q, Minus5, Minus17, Complex cytogenetics*** (in the absence of any Favorable features) | Any of ***KRAS, MPL, JAK2, p53m, Inv(3), Minus7, Minus5, Minus17, Complex cytogenetics*** (in the absence of any Favorable features) |

[a]Biomarkers such as *SMC1A, KMT2A,* and *PTPN11* were unavailable in the RWC. *Chromosome 7 deletion* (i.e., *Minus 7*) was used as a proxy of *Chromosome 7q deletion* (i.e., *Del7Q*).

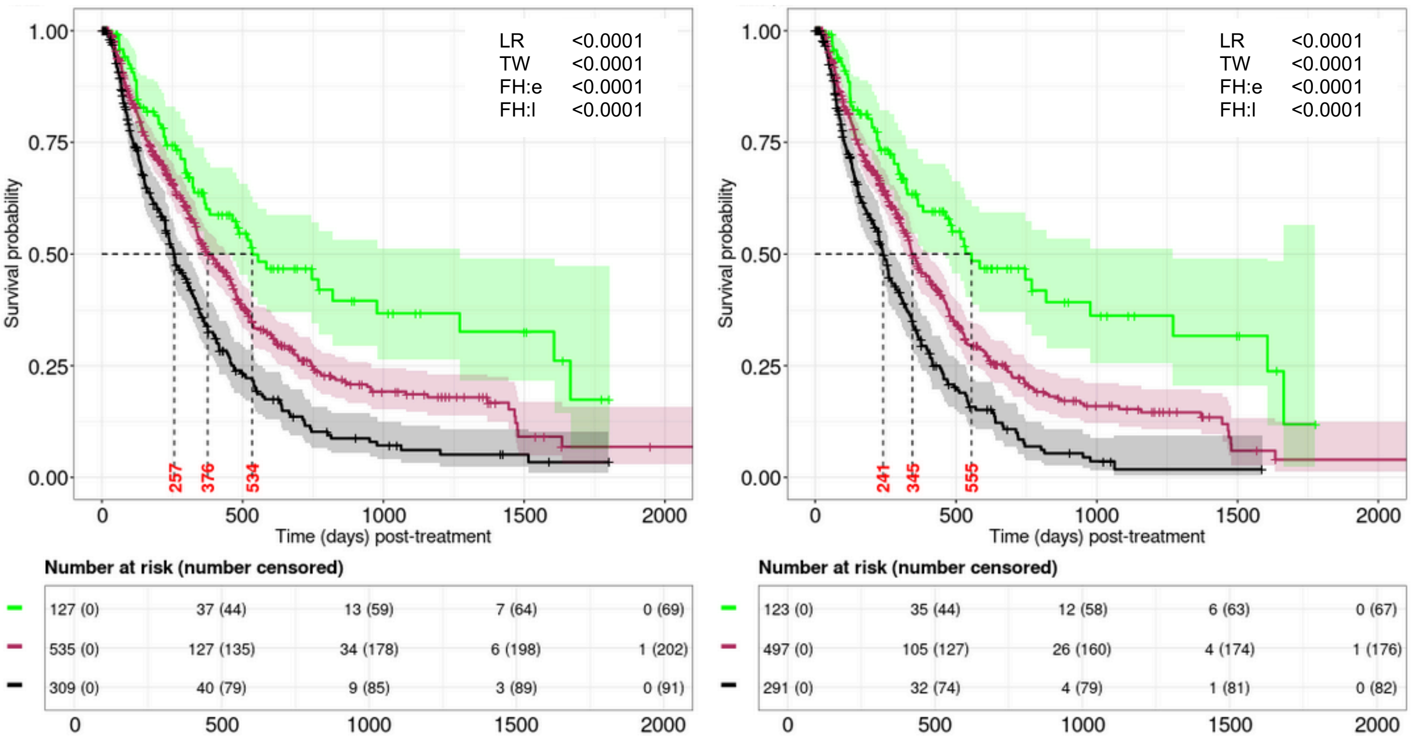

**Supplemental Figure 22, cont'd**

**C. RWC-RM$_{G,IV}$**

| Category | RM$_{G,IV}$ classification | [a]RWC-RM$_{G,IV}$ classification |
|---|---|---|
| **Favorable** | Any of ***DDX41, IDH1, NPM1*** (in the absence of any Adverse features) | Any of ***DDX41, IDH1, NPM1*** (in the absence of any Adverse features) |
| **Intermediate** | No Favorable or Adverse features, or $\geq$1 Favorable features with $\geq$1 Adverse features | No Favorable or Adverse features, or $\geq$1 Favorable features with $\geq$1 Adverse features |
| **Adverse** | Any of ***KRAS, KMT2A, PTPN11, SMC1A, JAK2, p53m, Del7q, Minus5, Minus17, Complex cytogenetics*** (in the absence of any Favorable features) | Any of ***KRAS, JAK2, p53m, Minus7, Minus5, Minus17, Complex cytogenetics*** (in the absence of any Favorable features) |

[a]Biomarkers such as *SMC1A, KMT2A,* and *PTPN11* were unavailable in the RWC. *Chromosome 7 deletion* (i.e., *Minus 7*) was used as a proxy of *Chromosome 7q deletion* (i.e., *Del7Q*).

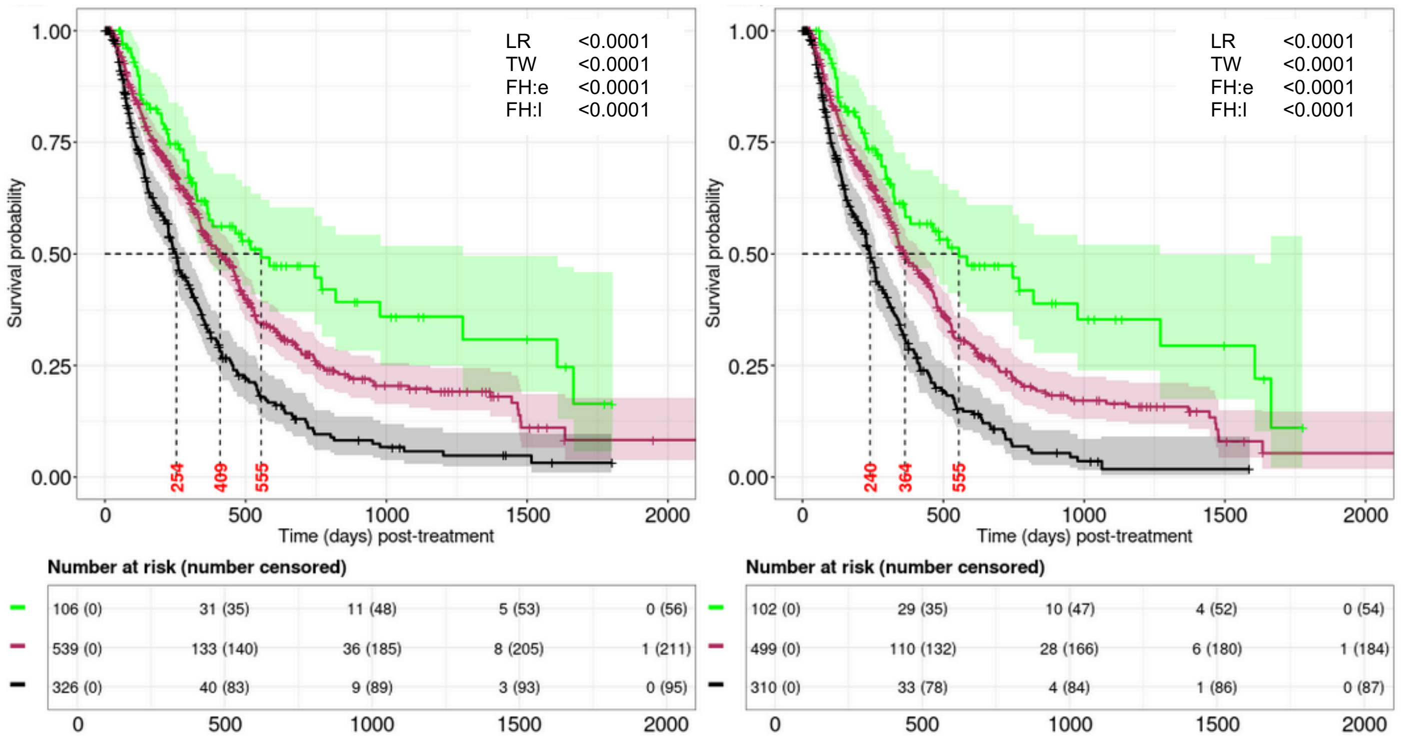

**Supplemental Figure 22, cont'd**

**D. RWC-RM$_{GP,I}$**

| Category | RM$_{GP,I}$ classification | [a]RWC-RM$_{GP,I}$ classification |
|---|---|---|
| **Favorable** | Any of ***CD10, CD19, BCOR, DNMT3A, DDX41, SF3B1, Good risk cytogenetics, IDH1, IDH2, KIT, NPM1*** (in the absence of any Adverse features) | Any of ***CD10, CD19, DNMT3A, DDX41, Good risk cytogenetics, IDH1, IDH2, KIT, NPM1*** (in the absence of any Adverse features) |
| **Intermediate** | No Favorable or Adverse features, or ≥1 Favorable features with ≥1 Adverse features | No Favorable or Adverse features, or ≥1 Favorable features with ≥1 Adverse features |
| **Adverse** | Any of ***CD5, CD7, CD16, CD34, CD36, CD71, CD11C, CSF3R, KRAS, KMT2A, MPL, PTPN11, SMC1A, JAK2, p53m, Del7q, Inv(3), t(9;11), Minus5, Minus17, Complex cytogenetics, Chr 7 abn:loss, Chr 8 abn:loss, DEK:loss, MECOM:gain, RARA:loss*** (in the absence of any Favorable features) | Any of ***CD34, CD36, CSF3R, KRAS, KMT2A, MPL, PTPN11, SMC1A, JAK2, p53m, Minus7, Inv(3), t(9;11), Minus5, Minus17, Complex cytogenetics, Chr 7 abn:loss, Chr 8 abn:loss, DEK:loss, MECOM:gain, RARA:loss*** (in the absence of any Favorable features) |

[a]Biomarkers such as *CD5, CD7, CD16, CD71, CD11C, BCOR, SF3B1, SMC1A, KMT2A, PTPN11, Chr 8 abn:loss, DEK:loss, MECOM:gain,* and *RARA:loss* were unavailable in the RWC. *Chromosome 7 deletion* (i.e., *Minus 7*) was used as a proxy of *Chromosome 7q deletion* (i.e., *Del7Q*).

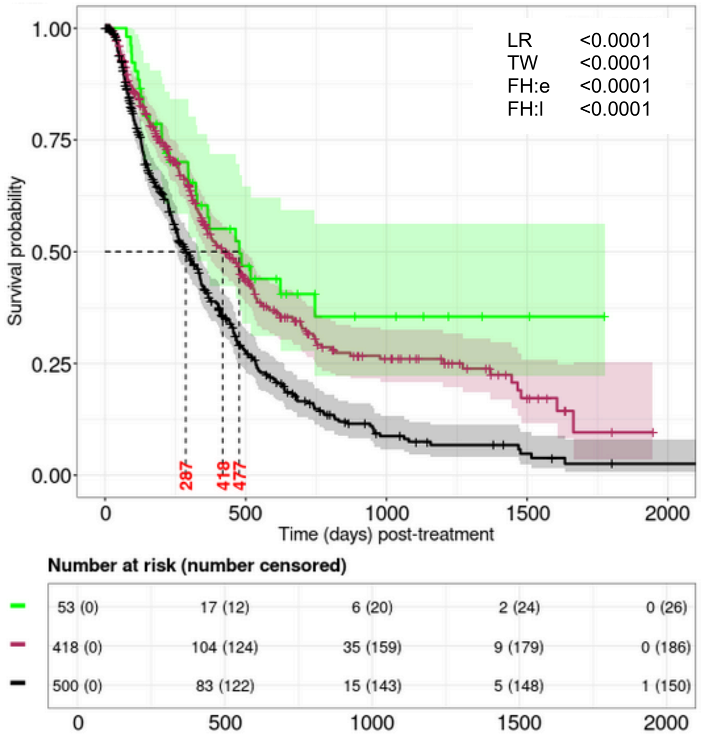


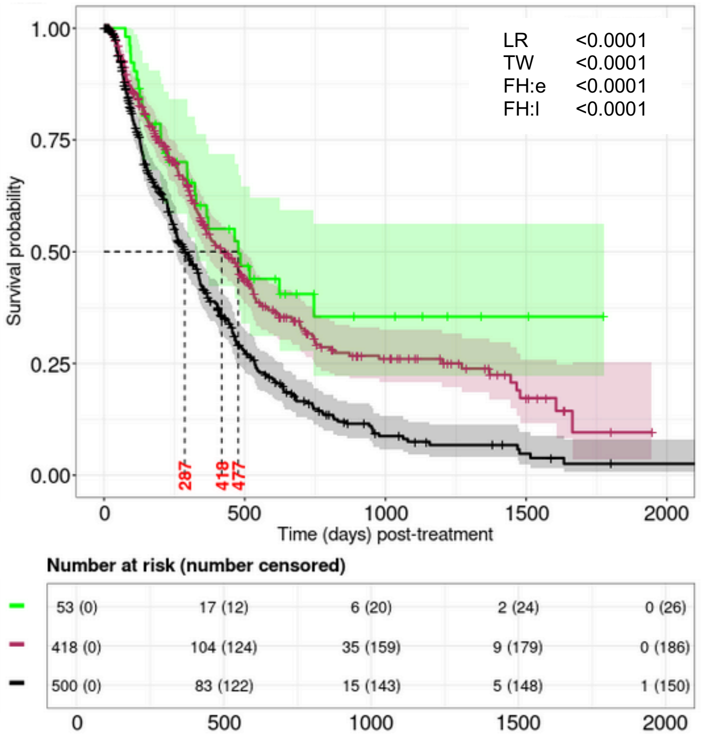

**Supplemental Figure 22, cont'd**

### E. RWC-$RM_{GP,II}$

| Category | $RM_{GP,II}$ classification | [a]RWC-$RM_{GP,II}$ classification |
|---|---|---|
| **Favorable** | Any of ***CD10, CD19, BCOR, DNMT3A, DDX41, SF3B1, IDH1, IDH2, NPM1*** (in the absence of any Adverse features) | Any of ***CD10, CD19, BCOR DNMT3A, DDX41, IDH1, IDH2, NPM1*** (in the absence of any Adverse features) |
| **Intermediate** | No Favorable or Adverse features, or >1 Favorable features with >1 Adverse features | No Favorable or Adverse features, or >1 Favorable features with >1 Adverse features |
| **Adverse** | Any of ***CD5, CD7, CD36, CD11C, KRAS, KMT2A, PTPN11, SMC1A, JAK2, p53m, Del7q, Minus5, Minus17, t(9;11), Complex cytogenetics, Chr 7 abn:loss, Chr 8 abn:loss, DEK:loss, MECOM:gain, RARA:loss*** (in the absence of any Favorable features) | Any of ***CD36, KRAS, JAK2, p53m, Minus7, Minus5, Minus17, t(9;11), Complex cytogenetics, Chr 7 abn:loss*** (in the absence of any Favorable features) |

[a]Biomarkers such as *CD5, CD7, CD11C, BCOR, SF3B1, SMC1A, KMT2A, PTPN11, Chr 8 abn:loss, DEK:loss, MECOM:gain,* and *RARA:loss* were unavailable in the RWC. *Chromosome 7 deletion* (i.e., *Minus 7*) was used as a proxy of *Chromosome 7q deletion* (i.e., *Del7Q*).

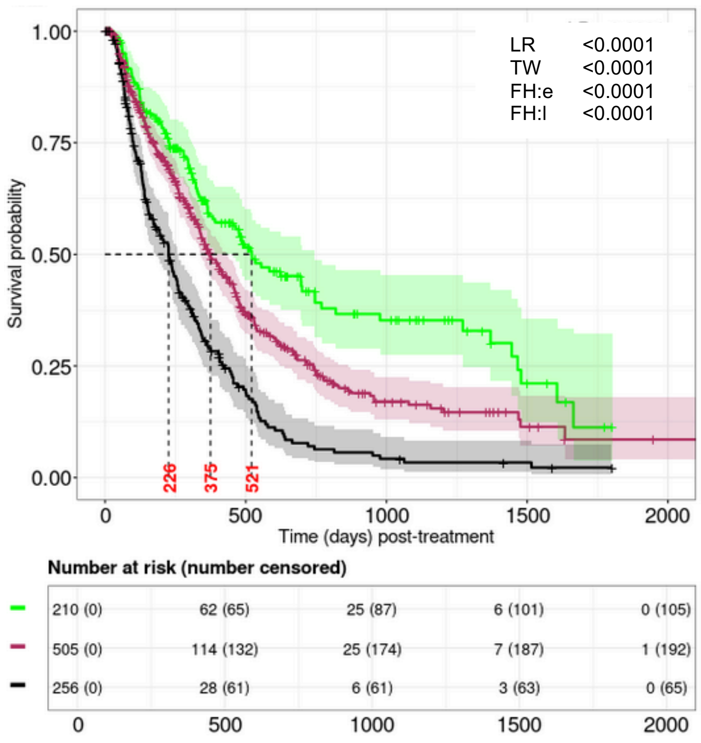


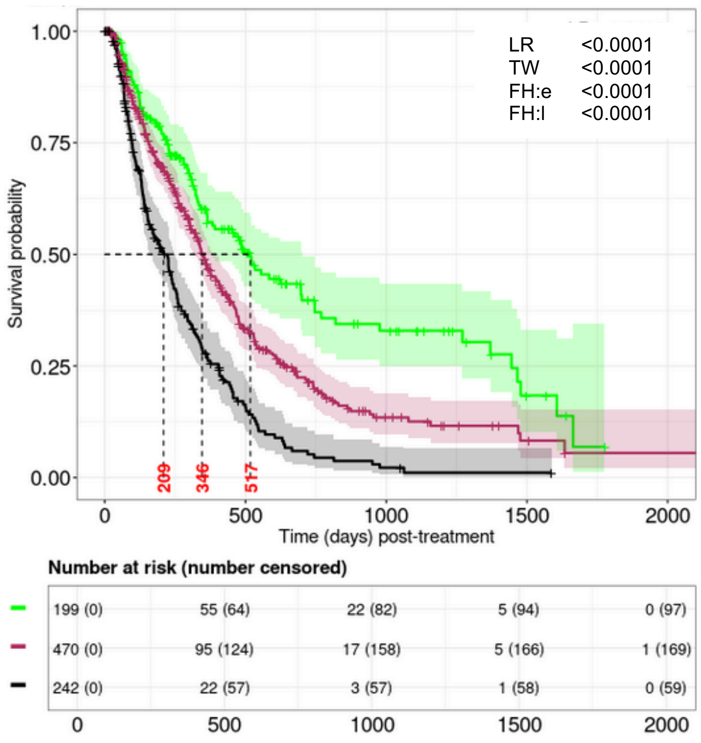

**Supplemental Figure 22, cont'd**

**F. RWC-RM$_{GP,III}$**

| Category | RM$_{GP,II}$ classification | [a]RWC-RM$_{GP,II}$ classification |
|---|---|---|
| **Favorable** | Any of ***CD10, DDX41, Good risk cytogenetics, IDH1, NPM1*** (in the absence of any Adverse features) | Any of ***CD10, DDX41, Good risk cytogenetics, IDH1, NPM1*** (in the absence of any Adverse features) |
| **Intermediate** | No Favorable or Adverse features, or ≥1 Favorable features with ≥1 Adverse features | No Favorable or Adverse features, or ≥1 Favorable features with ≥1 Adverse features |
| **Adverse** | Any of ***CD16, CD36, CD71, KRAS, KMT2A, PTPN11, SMC1A, MPL, JAK2, p53m, Inv(3), Del7q, Minus5, Minus17, Complex cytogenetics*** (in the absence of any Favorable features) | Any of ***CD36, KRAS, MPL, JAK2, p53m, Inv(3), Minus7, Minus5, Minus17, Complex cytogenetics*** (in the absence of any Favorable features) |

[a]Biomarkers such as *CD16, CD71, SMC1A, KMT2A,* and *PTPN11* were unavailable in the RWC. *Chromosome 7 deletion* (i.e., *Minus 7*) was used as a proxy of *Chromosome 7q deletion* (i.e., *Del7Q*).

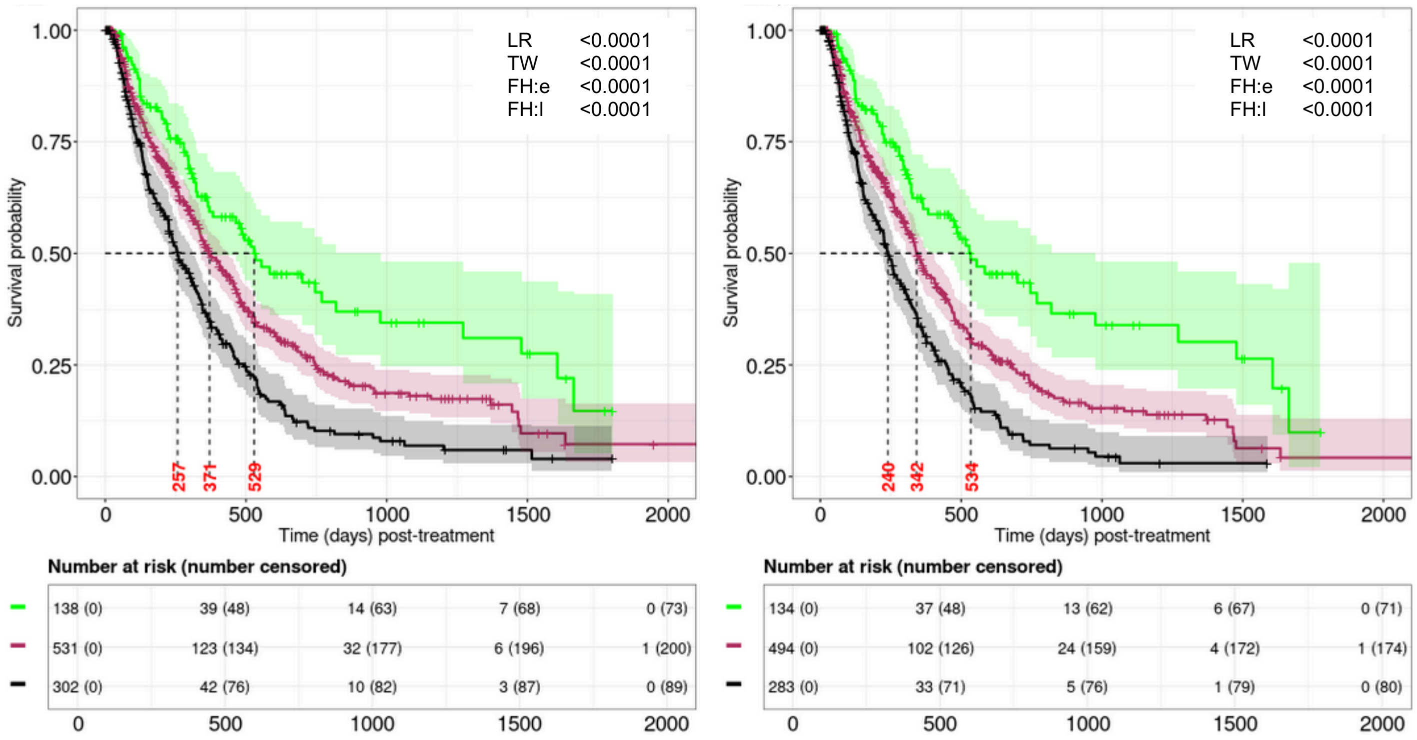

**Supplemental Figure 22, cont'd**

### G. RWC-RM$_{GP,IV}$

| Category | RM$_{GP,IV}$ classification | [a]RWC-RM$_{GP,IV}$ classification |
|---|---|---|
| **Favorable** | Any of ***CD10, DDX41, IDH1, NPM1*** (in the absence of any Adverse features) | Any of ***CD10, DDX41, IDH1, NPM1*** (in the absence of any Adverse features) |
| **Intermediate** | No Favorable or Adverse features, or >1 Favorable features with >1 Adverse features | No Favorable or Adverse features, or >1 Favorable features with >1 Adverse features |
| **Adverse** | Any of ***CD36, KRAS, KMT2A, PTPN11, SMC1A, JAK2, p53m, Del7q, Minus5, Minus17, Complex cytogenetics*** (in the absence of any Favorable features) | Any of ***CD36, KRAS, JAK2, p53m, Minus7, Minus5, Minus17, Complex cytogenetics*** (in the absence of any Favorable features) |

[a]Biomarkers such as *SMC1A, KMT2A,* and *PTPN11* were unavailable in the RWC. *Chromosome 7 deletion* (i.e., *Minus 7*) was used as a proxy of *Chromosome 7q deletion* (i.e., *Del7Q*).

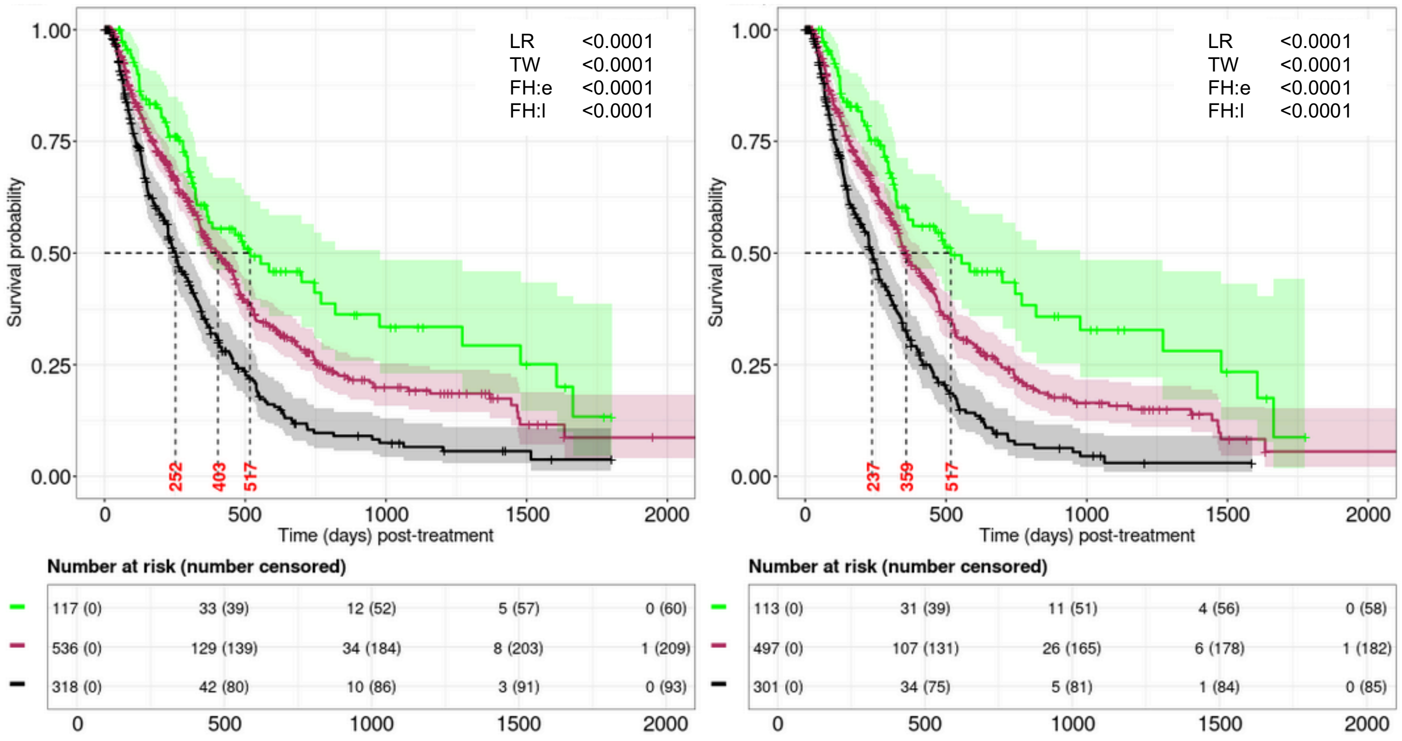

**Supplemental Figure 23. Agreements between the overall survival (OS) specific risk models and ELN22 based on the RWC.** Results are based on A) the full analytical set (FAS) with *Imputation-by-mode* and B) an imputed sample with respect to MICE. Patients with missing ELN22 were shown as “white” vertical lines.

**A. Agreements between the ELN22 and RWC-RMs based on the FAS.**

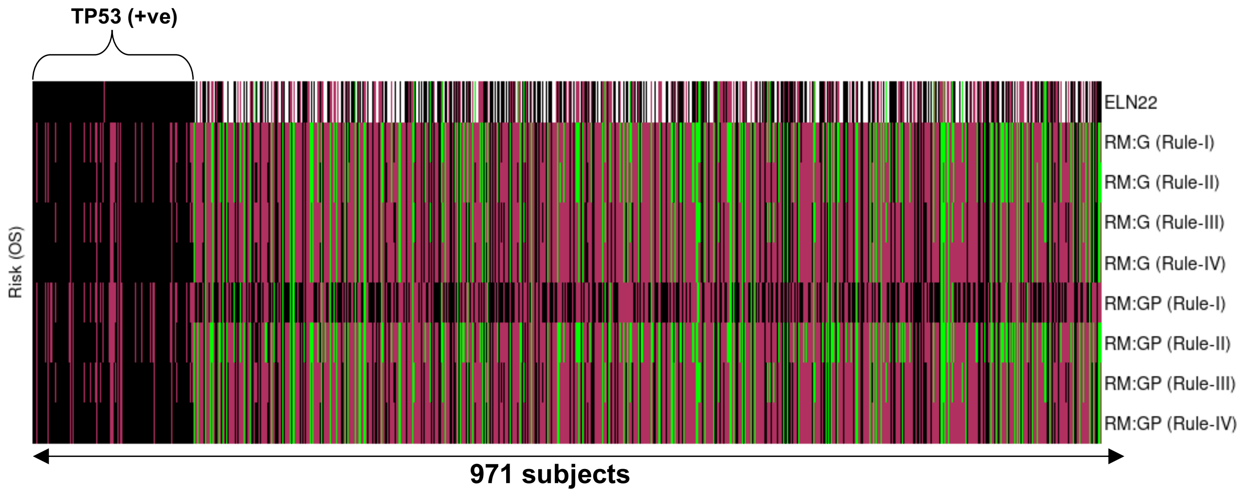


| *[a]Fleiss kappa (P values)* | *Favorable* | *Intermediate* | *Adverse* |
|---:|---|---|---|
| ***$RWC\text{-}RM_{G,I}$*** | 0.05 (0.181) | 0.03 (0.448) | 0.30 (<0.001) |
| ***$RWC\text{-}RM_{G,III}$*** | 0.14 (<0.001) | 0.20 (<0.001) | 0.37 (<0.001) |
| ***$RWC\text{-}RM_{G,IV}$*** | -0.08 (0.047) | 0.13 (<0.001) | 0.39 (<0.001) |
| ***$RWC\text{-}RM_{GP,I}$*** | 0.06 (0.135) | 0.00 (1.000) | 0.20 (<0.001) |
| ***$RWC\text{-}RM_{GP,II}$*** | -0.08 (0.048) | -0.04 (0.320) | 0.23 (<0.001) |
| ***$RWC\text{-}RM_{GP,III}$*** | 0.18 (<0.001) | 0.08 (0.041) | 0.30 (<0.001) |
| ***$RWC\text{-}RM_{GP,IV}$*** | -0.08 (0.028) | 0.07 (0.072) | 0.33 (<0.001) |

**Remarks:**

**The higher positive value (i.e., close to 1) means more agreement**
**The lower negative value (i.e., close to -1) means less agreement**
**Value close to 0 means agreement is no better than obtained by chance**

[a]Analytical set is comprised of the patients for who ELN22 was observed in the RWC.

**Supplemental Figure 23, cont'd**

**B. Agreements between the ELN22 and RWC-RMs based on an imputed set.**

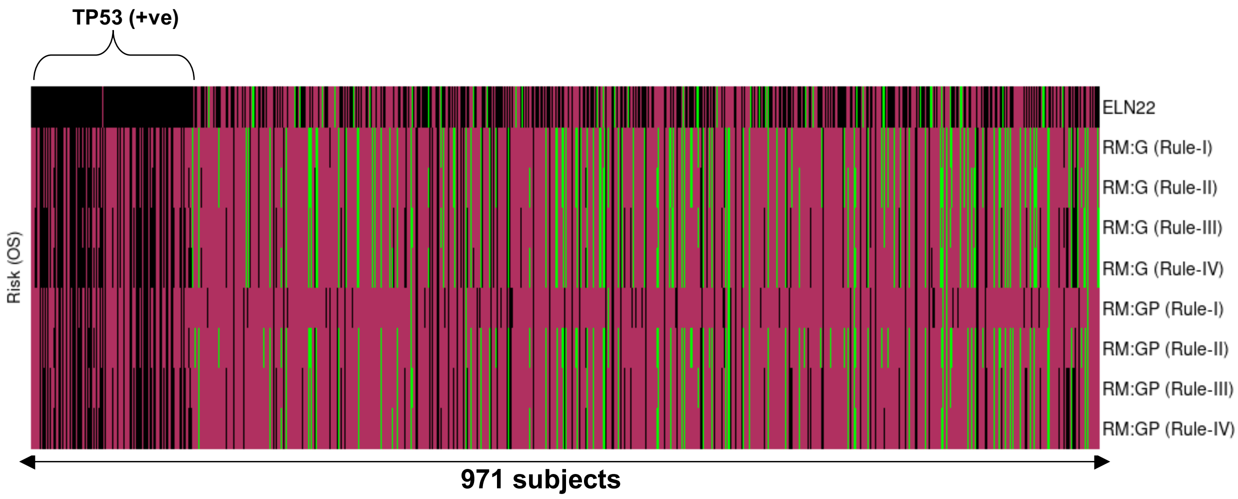


| *Fleiss kappa (P values)* | *Favorable* | *Intermediate* | *Adverse* |
|---|---|---|---|
| ***RWC-RM$_{G,I}$*** | -0.02 (0.563) | -0.02 (0.641) | 0.04 (0.201) |
| ***RWC-RM$_{G,II}$*** | -0.03 (0.339) | -0.00 (0.899) | 0.07 (0.038) |
| ***RWC-RM$_{G,III}$*** | 0.03 (0.428) | 0.04 (0.232) | 0.13 (<0.001) |
| ***RWC-RM$_{G,IV}$*** | -0.02 (0.499) | 0.04 (0.213) | 0.14 (<0.001) |
| ***RWC-RM$_{GP,I}$*** | -0.03 (0.320) | -0.08 (0.018) | 0.02 (0.651) |
| ***RWC-RM$_{GP,II}$*** | -0.02 (0.455) | -0.06 (0.051) | -0.01 (0.797) |
| ***RWC-RM$_{GP,III}$*** | 0.02 (0.499) | -0.04 (0.255) | 0.06 (0.076) |
| ***RWC-RM$_{GP,IV}$*** | -0.02 (0.586) | -0.03 (0.397) | 0.07 (0.025) |

**Remarks:**

**The higher positive value (i.e., close to 1) means more agreement**
**The lower negative value (i.e., close to -1) means less agreement**
**Value close to 0 means agreement is no better than obtained by chance**

**Supplemental Figure 24. Evaluation of predictive performance based on the RWC.**

Penalized CoxPH model is trained on the CU dataset adjusting for age, gender, and risk levels (e.g., Adverse, Intermediate, Favorable) using either ELN22 or the parental RM. Time-dependent AUC values are based on the (top) FAS with missing data being imputed by *imputation-by-mode* approach and (bottom) average over 10 imputed analytical sets. Results for RWC-$RM_{G,II}$ are also reported in the main draft and highlighted in red box.

| | cAUC ($2.5^{th}$,$97.5^{th}$) | ($25^{th}$, $75^{th}$) | $cAUC_{15}$ | P-value |
|---|---|---|---|---|
| ELN22 | 0.52 (0.50, 0.59) | (0.51, 0.53) | 0.52 | |
| RM:G Rule-I | 0.62 (0.59, 0.75) | (0.60, 0.66) | 0.64 | <0.0001 |
| RM:G Rule-II | 0.62 (0.60, 0.75) | (0.61, 0.66) | 0.64 | <0.0001 |
| RM:G Rule-III | 0.61 (0.56, 0.73) | (0.59, 0.64) | 0.62 | <0.0001 |
| RM:G Rule-IV | 0.61 (0.57, 0.72) | (0.59, 0.65) | 0.63 | <0.0001 |
| RM:GP Rule-I | 0.58 (0.56, 0.70) | (0.57, 0.61) | 0.58 | <0.0001 |
| RM:GP Rule-II | 0.62 (0.60, 0.74) | (0.61, 0.66) | 0.64 | <0.0001 |
| RM:GP Rule-III | 0.60 (0.57, 0.69) | (0.59, 0.64) | 0.61 | <0.0001 |
| RM:GP Rule-IV | 0.61 (0.58, 0.70) | (0.59, 0.64) | 0.61 | <0.0001 |

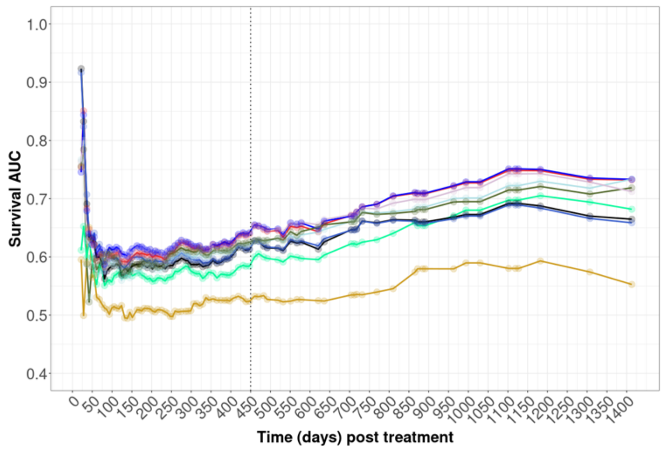


| | cAUC ($2.5^{th}$,$97.5^{th}$) | ($25^{th}$, $75^{th}$) | $cAUC_{15}$ | P-value |
|---|---|---|---|---|
| ELN22 | 0.51 (<0.50, 0.58) | (0.50, 0.52) | 0.52 | |
| RM:G Rule-I | 0.58 (0.55, 0.65) | (0.56, 0.60) | 0.58 | <0.0001 |
| RM:G Rule-II | 0.57 (0.55, 0.65) | (0.56, 0.60) | 0.58 | <0.0001 |
| RM:G Rule-III | 0.58 (0.55, 0.63) | (0.57, 0.59) | 0.59 | <0.0001 |
| RM:G Rule-IV | 0.57 (0.56, 0.62) | (0.56, 0.59) | 0.59 | <0.0001 |
| RM:GP Rule-I | 0.53 (0.51, 0.64) | (0.52, 0.55) | 0.54 | <0.0001 |
| RM:GP Rule-II | 0.57 (0.54, 0.66) | (0.55, 0.59) | 0.58 | <0.0001 |
| RM:GP Rule-III | 0.57 (0.55, 0.68) | (0.55, 0.59) | 0.58 | <0.0001 |
| RM:GP Rule-IV | 0.56 (0.54, 0.67) | (0.55, 0.59) | 0.58 | <0.0001 |

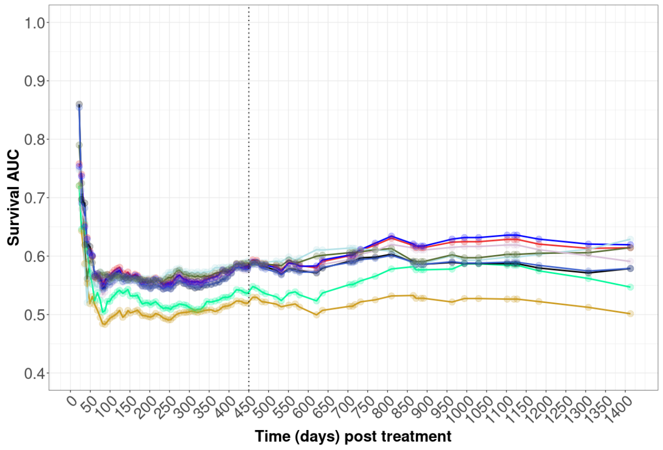

**Supplemental Figure 25. Sensitivity analysis for the evaluation of predictive performance based on the RWC.** Penalized CoxPH model is trained on the CU dataset adjusting for age, gender, and risk classification variable using either ELN22 or RWC-RM.

**A. Counts of patients assigned to each risk group when the reduced RWC feature definitions (i.e., RWC-RMs) were applied to the CU cohort.**

| | **RWC-RM$_{G,I}$** | | **RWC-RM$_{G,II}$** | | **RWC-RM$_{G,III}$** | | **RWC-RM$_{G,IV}$** | |
|---|---|---|---|---|---|---|---|---|
| **Risk group** | n[†] | n[¶] | n[†] | n[¶] | n[†] | n[¶] | n[†] | n[¶] |
| **Favorable** | 74/224 (33%) | 101/316 (32%) | 71/224 (32%) | 98/316 (31%) | 51/224 (23%) | 72/316 (23%) | 49/224 (22%) | 70/316 (22%) |
| **Intermediate** | 68/224 (30%) | 106/316 (34%) | 72/224 (32%) | 110/316 (35%) | 82/224 (37%) | 117/316 (37%) | 83/224 (37%) | 120/316 (38%) |
| **Adverse** | 82/224 (37%) | 109/316 (34%) | 81/224 (36%) | 108/316 (34%) | 91/224 (41%) | 127/316 (40%) | 92/224 (41%) | 126/316 (40%) |

| | **RWC-RM$_{GP,I}$** | | **RWC-RM$_{GP,II}$** | | **RWC-RM$_{GP,III}$** | | **RWC-RM$_{GP,IV}$** | |
|---|---|---|---|---|---|---|---|---|
| **Risk group** | n[†] | n[¶] | n[†] | n[¶] | n[†] | n[¶] | n[†] | n[¶] |
| **Favorable** | 35/224 (16%) | 45/316 (14%) | 71/224 (32%) | 95/316 (30%) | 49/224 (22%) | 67/316 (21%) | 47/224 (21%) | 65/316 (21%) |
| **Intermediate** | 82/224 (37%) | 120/316 (38%) | 74/224 (33%) | 115/316 (36%) | 86/224 (38%) | 121/316 (38%) | 87/224 (39%) | 124/316 (39%) |
| **Adverse** | 107/224 (48%) | 151/316 (48%) | 79/224 (35%) | 106/316 (34%) | 96/224 (40%) | 128/316 (41%) | 90/224 (40%) | 127/316 (40%) |

[†] Counts (proportions) of patients after excluding allo-HCT
[¶] Counts (proportions) of patients without excluding allo-HCT

**Supplemental Figure 25, cont'd**

**B. Time-dependent AUC values are based on the FAS where missing data elements being imputed by *imputation-by-mode* approach.**

| | cAUC (2.5th,97.5th) | (25th, 75th) | $cAUC_{15}$ | P-value |
|---|---|---|---|---|
| ELN22 | 0.52 (0.50, 0.59) | (0.51, 0.53) | 0.52 | |
| RM:G Rule-I | 0.62 (0.59, 0.75) | (0.60, 0.66) | 0.64 | <0.0001 |
| RM:G Rule-II | 0.62 (0.60, 0.75) | (0.61, 0.66) | 0.64 | <0.0001 |
| RM:G Rule-III | 0.61 (0.56, 0.73) | (0.59, 0.64) | 0.62 | <0.0001 |
| RM:G Rule-IV | 0.61 (0.57, 0.72) | (0.59, 0.65) | 0.63 | <0.0001 |
| RM:GP Rule-I | 0.58 (0.56, 0.70) | (0.57, 0.61) | 0.58 | <0.0001 |
| RM:GP Rule-II | 0.62 (0.60, 0.74) | (0.61, 0.66) | 0.64 | <0.0001 |
| RM:GP Rule-III | 0.60 (0.57, 0.69) | (0.59, 0.64) | 0.61 | <0.0001 |
| RM:GP Rule-IV | 0.62 (0.58, 0.70) | (0.60, 0.65) | 0.61 | <0.0001 |

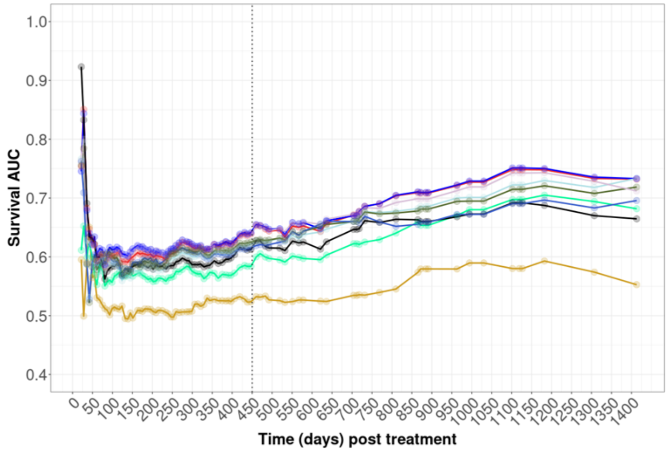


**C. Time-dependent AUC values are based on the FAS where missing data elements being imputed by MICE approach.**

| | cAUC (2.5th,97.5th) | (25th, 75th) | $cAUC_{15}$ | P-value |
|---|---|---|---|---|
| ELN22 | 0.52 (0.50, 0.60) | (0.51, 0.53) | 0.53 | |
| RM:G Rule-I | 0.58 (0.55, 0.65) | (0.56, 0.60) | 0.58 | <0.0001 |
| RM:G Rule-II | 0.57 (0.55, 0.65) | (0.56, 0.60) | 0.59 | <0.0001 |
| RM:G Rule-III | 0.58 (0.55, 0.63) | (0.57, 0.59) | 0.59 | <0.0001 |
| RM:G Rule-IV | 0.58 (0.56, 0.62) | (0.56, 0.60) | 0.59 | <0.0001 |
| RM:GP Rule-I | 0.53 (0.51, 0.64) | (0.52, 0.55) | 0.54 | <0.0001 |
| RM:GP Rule-II | 0.57 (0.55, 0.65) | (0.55, 0.59) | 0.58 | <0.0001 |
| RM:GP Rule-III | 0.57 (0.55, 0.66) | (0.56, 0.59) | 0.58 | <0.0001 |
| RM:GP Rule-IV | 0.57 (0.55, 0.64) | (0.56, 0.58) | 0.58 | <0.0001 |

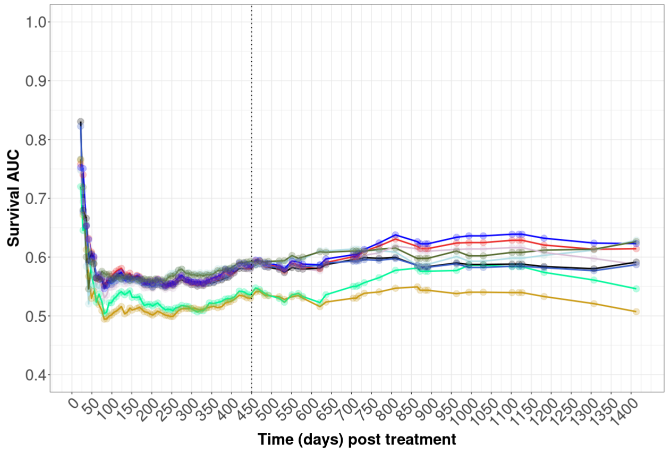
